# Clinical review before execution of executable guideline encodings: a prospective registered study of three ocular hypertension guidelines

**DOI:** 10.64898/2026.11.20.26368564

**Authors:** Andrea Ragosta, Francesco Bruni

## Abstract

**Objective:** To determine whether executable encodings of three ocular hypertension guidelines, produced under a prospectively frozen pipeline barring author-defined operationalisations, are judged faithful by independent clinical review before execution, and to characterise findings on retained content, exclusions and divisions.

**Methods:** Under a registered protocol, one ophthalmologist encoded European Glaucoma Society (EGS), American Academy of Ophthalmology (AAO) and National Institute for Health and Care Excellence (NICE) guidelines; a second encoded a sample. A gate barred author-defined operationalisations. Before execution, two other ophthalmologists reviewed branches, exclusions and division records against predefined materiality criteria.

**Results:** The gate admitted 0, 2 and 1 branches (EGS, AAO, NICE) from 748, 322 and 198 eligible units. Review of these branches raised no primary finding (AAO 0/2, NICE 0/1); at these denominators, uninformative. In the overlap, one NICE omission (laser trabeculoplasty) was judged material. The exclusion reviewer flagged material source-stated decision content in 601/2,109, 216/234 and 151/179 excluded passages. A masked post-hoc reader agreed with 31/50, 11/50 and 28/50 sampled flags; he judged 1/246 passages directly relevant to the initial treat-or-observe decision and 0/100, 0/68 and 8/78 needed to preserve a retained unit’s meaning. No synthetic case resolved to observation.

**Conclusion:** Without author-defined choices, few branches were instantiable and none resolved to observation. Fidelity was not determinable at these denominators; exclusion flags were reader-dependent, and some NICE exclusions may have carried meaning needed by retained units. Exclusion review shows what a scope restriction sets aside; its reproducibility must be established before it measures fidelity.

## 1. Introduction

Clinical practice guidelines are written for clinicians, while decision support needs logic that a computer can evaluate; computer-interpretable guideline (CIG) research has long concentrated on this translation [1,2]. It requires choices: recommendations must be segmented and atomised, and vague terms made determinate [3–5]. Clinicians and informaticians encode the same text differently [6,7], and formalisation exposes gaps in the guideline itself [8,9]. Complementary frameworks support markup, authoring, implementability appraisal and staged representation [3,4,10–12], and guideline producers now build computable artefacts themselves [13,14].

### Statement of Significance

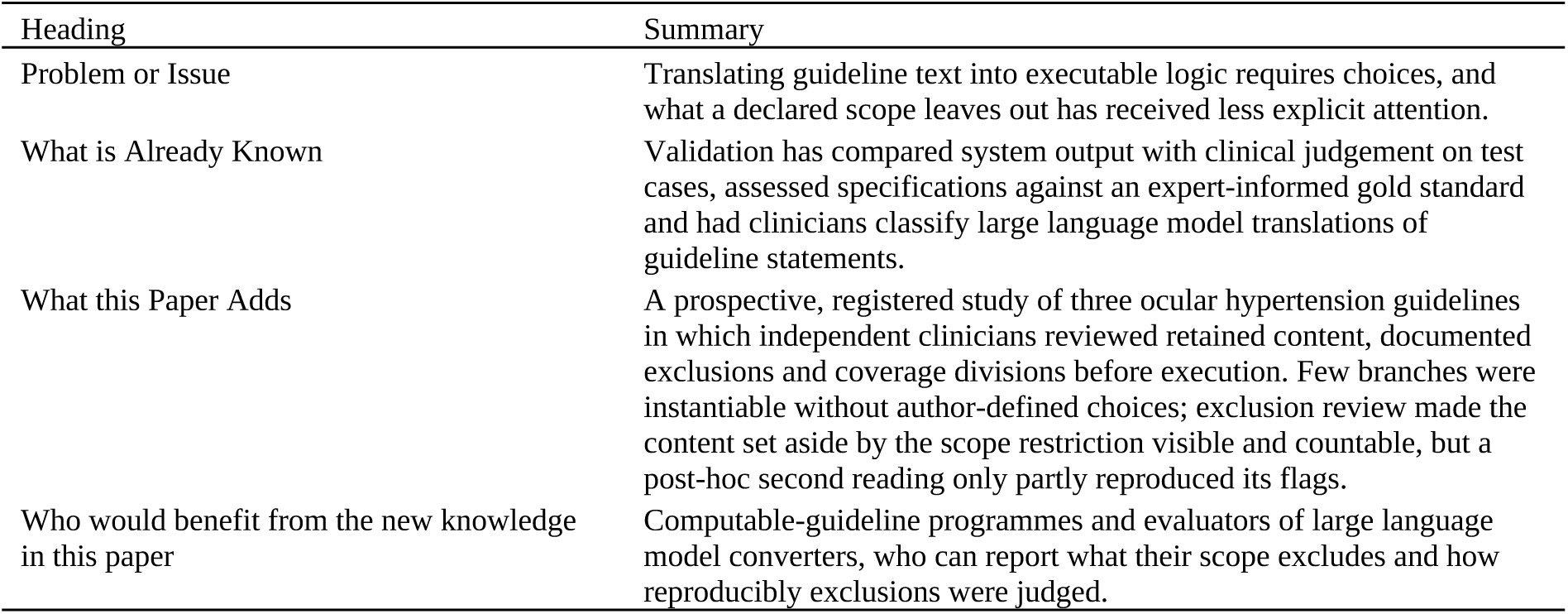

| Heading | Summary |
|---|---|
| Problem or Issue | Translating guideline text into executable logic requires choices, and what a declared scope leaves out has received less explicit attention. |
| What is Already Known | Validation has compared system output with clinical judgement on test cases, assessed specifications against an expert-informed gold standard and had clinicians classify large language model translations of guideline statements. |
| What this Paper Adds | A prospective, registered study of three ocular hypertension guidelines in which independent clinicians reviewed retained content, documented exclusions and coverage divisions before execution. Few branches were instantiable without author-defined choices; exclusion review made the content set aside by the scope restriction visible and countable, but a post-hoc second reading only partly reproduced its flags. |
| Who would benefit from the new knowledge in this paper | Computable-guideline programmes and evaluators of large language model converters, who can report what their scope excludes and how reproducibly exclusions were judged. |

## 2. Related Work

Validation has compared system output with clinical judgement on test cases [15], assessed specifications for completeness and correctness against an expert-informed gold standard [16], and checked consistency against a separately coded implementation, which does not establish fidelity [17]. Back-translation reviewed by clinical experts tests semantic fidelity directly [18]. Large language models (LLMs) make the question more pressing: Gupta and colleagues had clinicians classify each National Institute for Health and Care Excellence (NICE) statement’s translation into logic, including statements yielding no rule [19], and CPGPrompt validated generated guidance trees before evaluating LLM traversal [20]. What a declared scope leaves out of the encoding has received less explicit attention.

We therefore designed a prospective, registered study in which materiality criteria were fixed in advance, author-defined operationalisations were barred from execution, and independent clinicians reviewed what was carried, what was excluded and how the source was divided, before any case was executed.

As a test domain we chose the decision to treat or to monitor in ocular hypertension: binary, with explicit thresholds, yet reliant on judgement terms such as risk of visual impairment within the patient’s lifetime [21–23]. We analysed three current guidelines in parallel: the European Glaucoma Society (EGS) guidelines, the American Academy of Ophthalmology (AAO) Primary Open-Angle Glaucoma (POAG) Suspect Preferred Practice Pattern, and NICE NG81. **Objective:** to determine whether executable encodings of these guidelines, produced under a prospectively frozen pipeline barring author-defined operationalisations, are judged faithful to their sources by independent clinical review before execution, and to characterise the findings on retained content, exclusions and divisions.

## 3. Methods

### 3.1. Design and registration

The protocol was registered on OSF Registries (osf.io/bhmz2) on 1 October 2026, before v0.2 segmentation and encoding; it fixed the question, sources, encoding rules, acceptance criteria, outcomes and analysis. On 28 October 2026, after encoding and before any release to reviewers, governed amendments fixed what the omission-detection pass compares (each division’s source-stated decision content against that division’s records, not the executable form), revised the review instrument and added a claim limit to the primary outcome, without changing any denominator, class or bound; they were disclosed in a linked OSF entry that day. Two earlier registry updates (18 October) disclosed post-exposure changes to the reconciliation rules (Supplement S1.1). The review instrument and outcome-computation specification were locked before reviewer release. The registration is embargoed until peer review ends; the study is not a Registered Report. The prespecified study artefacts were hash-committed to an append-only freeze register (Figure 1); later freezes and deviations are dated in Supplement S1.

**Figure 1.**
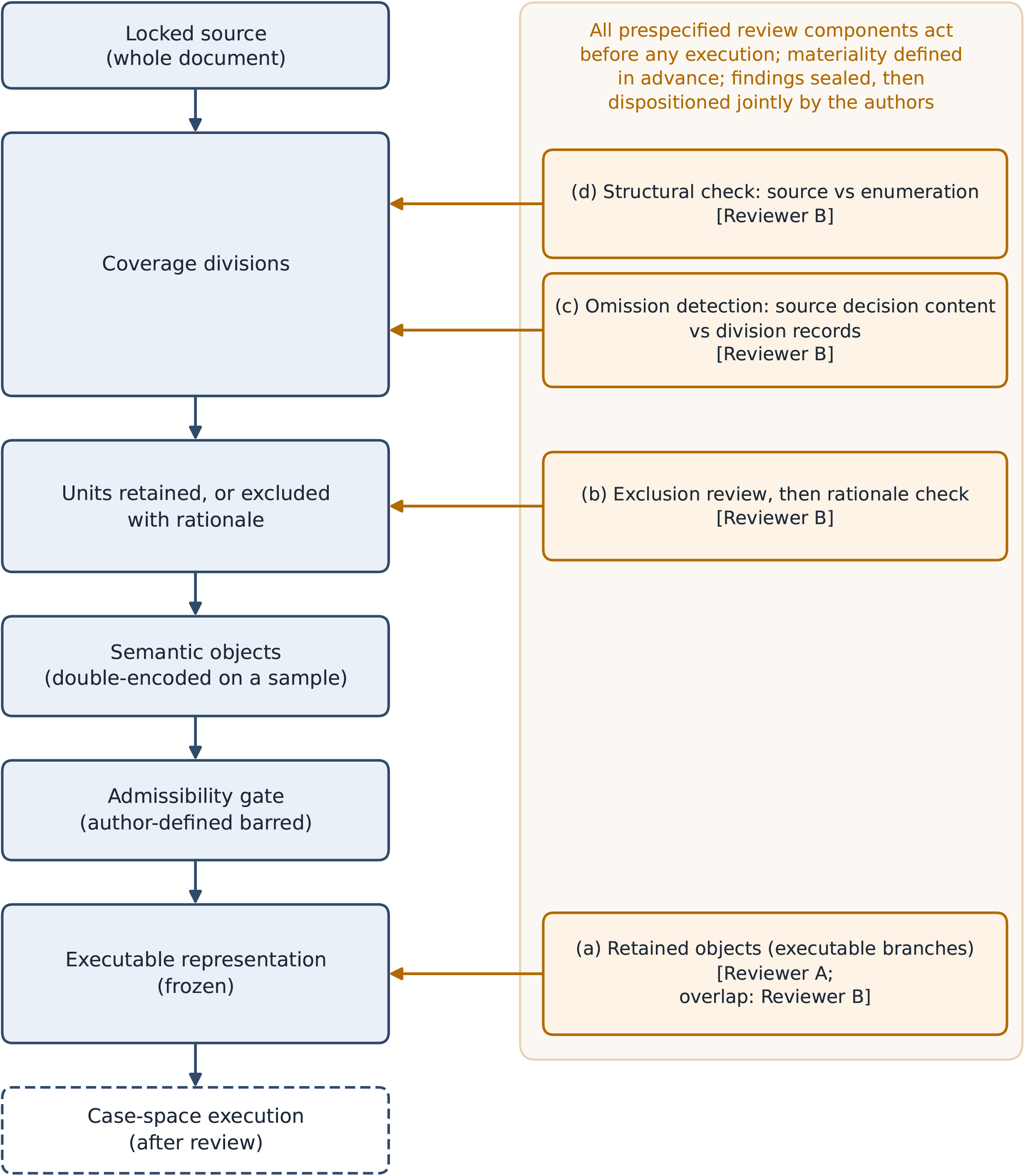
Study pipeline. From the locked source document to case-space execution: coverage divisions, units retained or excluded with a recorded rationale, semantic objects (double-encoded on a sample), the admissibility gate that bars author-defined operationalisations, and the frozen executable representation. Orange boxes mark where each review component acts. All prespecified review components act before any execution, against a materiality definition fixed in advance; findings were sealed and then dispositioned jointly by the authors. Drawn with plotting scripts written with AI assistance.

### 3.2. Sources and segmentation

The sources were the EGS guidelines, 6th edition (2025), the AAO POAG Suspect Preferred Practice Pattern (2025/2026) and NICE NG81 (updated 2022, reviewed 2025) [21–23], each hash-locked on 18 September 2026 and used in full. The study population was adults with ocular hypertension, without established glaucomatous damage and never treated. The decision domain was the initial choice between starting intraocular pressure (IOP)-lowering treatment and observation without treatment.

Each document (212, 49 and 37 pages) was enumerated into coverage divisions, structural sections covering the whole document (EGS 262, AAO 58, NICE 124), and text within divisions was segmented into units, one per separately assertable guidance-bearing proposition (Supplement S1.1). A unit was retained for semantic encoding or excluded with a recorded reason and rationale; exclusion was allowed only if the unit did not directly bear on the study population or decision domain and was not needed to preserve the meaning of any retained unit. Retained units formed a canonical record of 1,291 units, of which 1,268 were eligible for the executable stage (EGS 748, AAO 322, NICE 198).

### 3.3. Double semantic encoding

Encoder 1 (AR, the principal investigator (PI), who also authored the executable encoding, co-disposed findings and read the review-frame rule) semantically encoded all retained units; Encoder 2 (FB) independently encoded a duplicate sample of retained candidate-proposition (Class A) units, body-stratified and selected by a committed seed (EGS 83 of 581, AAO 74 of 302, NICE 60 of 156); the other retained units were dependency objects. Initial contributions were sealed before comparison. The encoders jointly reconciled a mechanically generated register of 2,434 discrepancies in 316 duplicated units and dependency objects (Supplement S1.8); 17 residual disagreements stayed unresolved under a non-use rule. There was no third adjudicator.

### 3.4. Admissibility gate and executable representations

Before execution, each executable form was classed by its dependence on the source as source-derived, source-constrained, author-defined or inadmissible (Table 1). Only source-derived forms, and source-constrained forms whose choice rule had been declared at study level before inspection, were eligible for execution; author-defined content was barred. Rule families were registered for each guideline before its units were inspected.

**Table 1.** Approximate conceptual correspondence between the study’s terms and established guideline-formalisation frameworks [3,10–12]. Admissibility classes: *source-derived*, the source fixes the form with no encoder choice; *source-constrained*, the encoder chooses within a set the source makes decidable; *author-defined*, supplied by the study; *inadmissible*, not representable under the registered rules. The correspondence is an analogy; the study does not implement, or claim conformance with, these frameworks or HL7 FHIR.

| Study term | What it is | CPG-on-FHIR knowledge level (analogy) | GEM / BRIDGE-Wiz | GLIA dimension |
|---|---|---|---|---|
| Locked source and coverage divisions | Whole guideline, hash-locked, enumerated into divisions | L1 narrative | Document markup | — |
| Units and semantic objects (canonical record) | Segmented source text and its encoded propositions, double-encoded on a sample | L2 semi-structured | Atomisation | — |
| Admissibility class (source-derived, source-constrained, author-defined, inadmissible) | How far the executable form is fixed by the source rather than by the encoder | L2 → L3 | De-abstraction, disambiguation | Decidability |
| Executable representation | Versioned branches with a closed output vocabulary (initiate, observe, unresolved) | L3 structured | Executable recommendation | Executability |
| Engine and case space | Deterministic evaluation over a synthetic grid | Analogous to L4 only; no clinical-system implementation | — | — |

Admissible units were authored as a versioned executable representation for each guideline. The output vocabulary mirrors the decision domain: resolved to initiate treatment, resolved to observe without treatment, or unresolved, with the blocking conditions named. Each representation passed validator and rendering-coverage checks, and Encoder 2 checked its transcription against the canonical record (design-side verification, not clinical review). Supplement S1.7 traces each admitted branch from source locator to executable condition, output and missing-data result.

### 3.5. Independent review before execution

Two ophthalmologists experienced in glaucoma, neither an author, reviewed the frozen versions after declaring their eligibility and relationships with the authors. Reviewer A had reviewed material from the historical v0.1 study; both were naïve to v0.2 (PI determination). Reviewer A reviewed the retained-object frame, which the registered rule defines as “the set of in-scope retained objects of all output-intended lineages … at the review-version freeze point”, generated “from the frozen review versions”. Before release, the PI read this as the objects encoded in the frozen executable representations (AAO two, NICE one, EGS none); the alternative reading, all retained objects of AAO and NICE (539), was not adopted. The narrower reading was chosen to shorten the review; an independent pre-release artificial intelligence (AI) audit (Kimi) found it compatible with the frozen records (Supplement S1.5). For each object Reviewer A answered 12 items, from source span to executable rendering (Table S3-6). Reviewers received a written manual and a non-scored exercise on a fictional guideline; there was no calibration or pilot on study material. Reviewer B judged every documented exclusion source-first at its location; after sealing those judgements, he checked each recorded exclusion rationale, ran an omission-detection pass over every division and checked each source structurally against its enumeration. In the initial review, each item received one judgement, except the overlap: retained objects (at most 10 per guideline, by committed seed; here all) also judged by Reviewer B. The exclusion instrument was streamlined (passages grouped by division, prefilled locators, one-click answers). Reviewer B reported using computing support, including scripts and AI assistance, to organise the passages, word his findings in a standard form and transcribe his decisions; each assessment was his own (Supplement S1.6).

What counts as material was defined before review (verbatim in Supplement S1.1): a change, as represented, in what the source states about a recommendation’s direction, strength or certainty, population or scope, or conditions for applying it. For each item the reviewer first recorded whether he raised a finding; category, stance and materiality answer (material, not material or cannot judge) were recorded only for raised findings. For exclusions, the task fixed before reviewer release asked the reviewer to judge “whether excluding it loses decision content”, and the registered definition states that, for a documented exclusion, “loss includes decision content of these kinds that the source states and the representation omits”; the 28 October amendments changed neither. In a dated clarifying addendum after the analysis (Supplement S1.6; sealed record unchanged), Reviewer B reported that his material answers on exclusions referred to the decision content the source states in the passage, not to whether the exclusion was proper for the study’s domain. In the initial phase, reviewers saw the sources and the representations under review (canonical semantic records and executable renderings), not the encoders’ initial or reconciliation records, agreement results or duplicated units; initial findings were sealed before the unblinded supplementary phase.

The two encoders then jointly dispositioned each sealed finding as change, no change, routed to the procedure responsible for the affected record, or accepted as unresolved. Exclusion findings were dispositioned with one standard reason that restates the recorded exclusion rationale; this was not an independent re-examination of each passage. A *no change* outcome records the disposers’ judgement; it does not refute the finding, which stays on record. Before the PI determined whether each version passed, an AI gatekeeper (Codex) checked the procedure.

After analysis, a further glaucoma ophthalmologist, not otherwise involved in the study, who collaborates with the corresponding author on unrelated research, read a stratified random sample of 246 documented exclusions (Table S3-12), masked to Reviewer B’s answers, the rationales and the dispositions, with the locked sources and views of the retained units (identifier, location and text). He answered the original exclusion task, given verbatim with the frozen definition but without Reviewer B’s addendum, and whether each passage bears directly on the initial treat-or-observe decision or is needed to preserve a retained unit’s meaning. This reading was post hoc and not registered.

### 3.6. Outcomes

The primary outcome, for each guideline, was the number of review units with at least one finding answered *material* (P-1) and with at least one asserted finding (P-2), for five components: (a) retained objects, (b) documented exclusions, (c) enumerated divisions, (d) the structural check and (e) version-level items. P-1’s upper bound adds indeterminate, unable-to-judge (J) and incomplete (R) units; P-2’s adds unresolved-stance, J and R units; query-only units are reported separately. Component (c) is reported with a structural indicator (S) for unenumerated observations. Secondary outcomes, in planned order, were finding categories, disposition profile, acceptance status, execution determinacy (the distribution of output states over the case space) and inter-encoder agreement. Exploratory outcomes included rationale findings and supplementary-phase findings; instantiation yield was reported as context.

### 3.7. Case space and execution

The seven case variables came only from the frozen representations; none was shared between guidelines. Numeric variables took values just below, at and just above each cut point. Every combination was generated (96 parent cases), with 672 variants in which one variable at a time was not established. A locked deterministic engine ran both representations on all cases (1,536 inputs); a replay was byte-identical.

### 3.8. Statistical analysis

Prespecified sampled inter-encoder agreement estimands carry exact design-based hypergeometric 95% intervals; Cohen’s κ and Gwet’s AC1 are reported together because κ is unstable when one category dominates [24,25]. For the post-hoc second reading, positive and negative percent agreement (on raising a finding, conditional on Reviewer B’s outcome) carry Clopper–Pearson exact 95% intervals; κ and AC1 are not reported because sampling was stratified by Reviewer B’s outcome. No hypothesis was tested. The prespecified engine, input validator, analysis plan and analysis scripts were locked before execution; an input-derivation script and input maps were pinned by hash rather than locked (Supplement S1). Codex, in a separate session, recomputed all 16 outputs byte for byte and independently re-implemented the primary outcome, with the same result.

## 4. Results

The gate admitted 0, 2 and 1 branches from 748, 322 and 198 eligible units (EGS, AAO, NICE), none author-defined (Figure 2). Their primary review was uninformative at these denominators. Findings concentrated in documented exclusions (601/2,109, 216/234 and 151/179) and were partly reproduced at post-hoc second reading. No synthetic case resolved to observation. Both reviewers sealed all initial-phase judgements, no material was eligible for the supplementary phase, and lower and upper bounds coincided, with no indeterminate, query-only, unable-to-judge (J) or incomplete (R) item.

**Figure 2.**
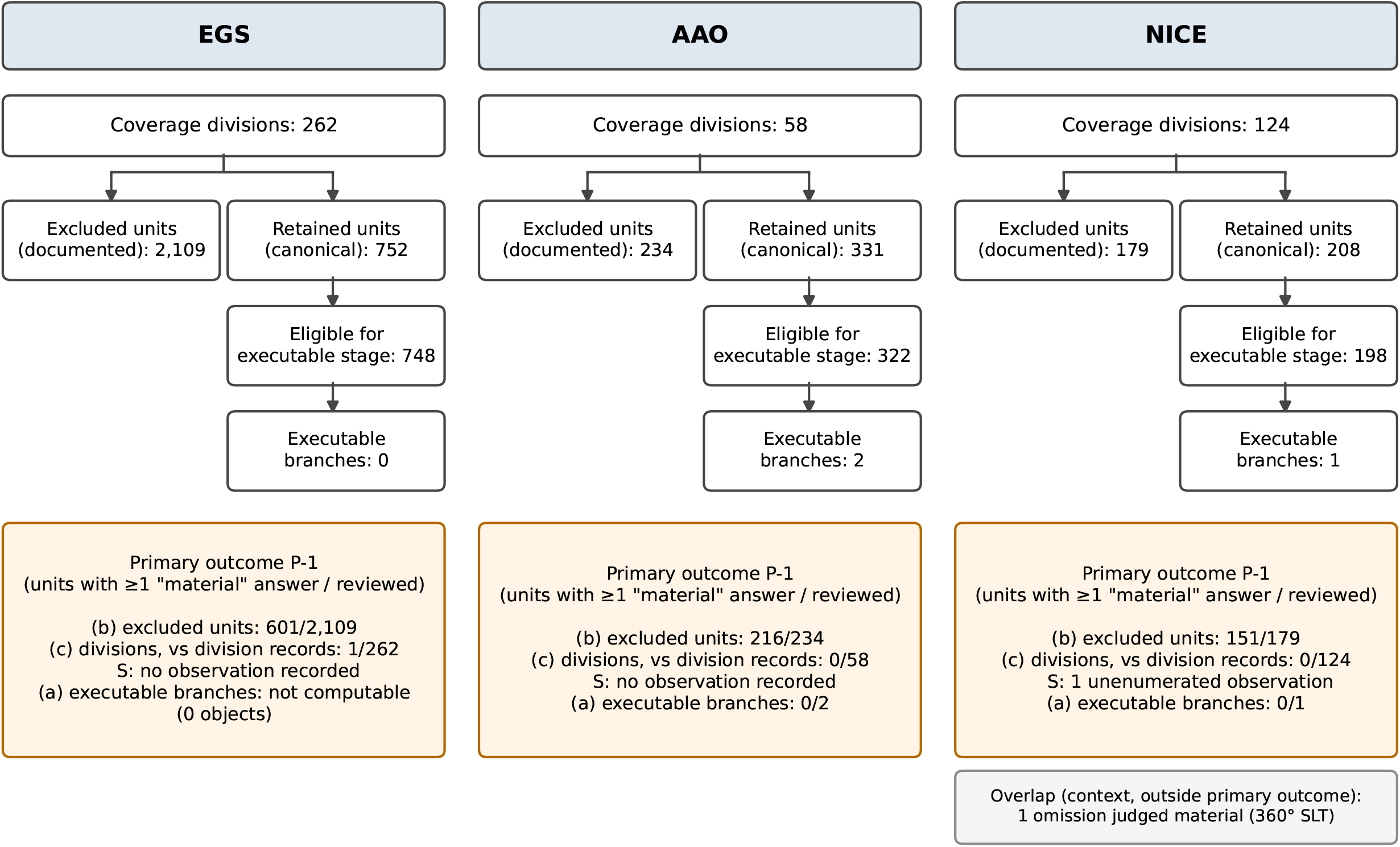
Flow for each guideline, shown in parallel without totals across guidelines. Excluded units are documented exclusions at extraction; retained units are those in the canonical record; the 23 retained units not eligible for the executable stage were declared non-admissible at the close of semantic encoding (register entry 151), with their unresolved states preserved. Orange boxes give, for each primary component, the number of review units with at least one answer of “material” over the number reviewed; these are reviewer judgements, not verified departures. For excluded units, “material” refers to decision content the source states in the excluded passage (Supplement S1.6). Component (c) compares source-stated decision content with each division’s records, not with the executable version. The selective laser trabeculoplasty (SLT) overlap finding, shown separately, is descriptive and outside the primary outcome. Drawn with plotting scripts written with AI assistance.

### 4.1. Primary outcome

Table 2 shows P-1 and P-2 for each component.

**Table 2.** Primary outcome by component and guideline: units with ≥1 finding answered material (P-1) and with ≥1 asserted finding (P-2). Lower and upper bounds coincided in every cell.

| Component | EGS | AAO | NICE |
|---|---|---|---|
| (a) Retained objects | NOT COMPUTABLE (denominator 0) | P-1 0/2; P-2 0/2 | P-1 0/1; P-2 0/1 |
| (b) Documented exclusions | P-1 601/2,109; P-2 601/2,109 | P-1 216/234; P-2 216/234 | P-1 151/179; P-2 151/179 |
| (c) Enumerated divisions (vs division records) | P-1 1/262; P-2 1/262 | P-1 0/58; P-2 0/58 | P-1 0/124; P-2 0/124 |
| Structural indicator S for (c) | No observation recorded | No observation recorded | Observation recorded (1 unenumerated observation) |
| (d) Structural check | P-1 0/1; P-2 0/1 | P-1 0/1; P-2 0/1 | P-1 0/1; P-2 1/1 |
| (e) Version-level items | NOT APPLICABLE | NOT APPLICABLE | NOT APPLICABLE |

In component (b), every raised finding was answered material in each guideline. Component (c) compares source-stated decision content with the enumerated division’s records, not with an executable version; it covers enumerated divisions only. No finding was J (unable to judge), R (incomplete judgement), indeterminate or query-only. P-2 is not presented as answering materiality.

- **(a) Retained objects** (adopted frame: the objects encoded in the executable representations): no finding on any of them (AAO 0/2, NICE 0/1); EGS not computable (denominator 0).
- **(b) Documented exclusions:** Reviewer B raised a finding stating that exclusion omitted decision-relevant source content for 601 of 2,109 EGS, 216 of 234 AAO and 151 of 179 NICE exclusions. Every exclusion finding was answered “material”; none was answered “not material” or “cannot judge”. Materiality was elicited only after a finding was raised, so exclusions without a finding received no materiality rating; within this component the observed pattern is finding versus no finding. They do not estimate inappropriate exclusion or in-domain clinical loss. P-2 was identical. Post-hoc tabulation by recorded material class showed no Class-C flags (EGS 0/40, AAO 0/9, NICE 0/7), lower Class-B than Class-A flag rates within each guideline, and Class-A flag rates of 31.9% (EGS), 99.5% (AAO) and 92.1% (NICE) (Table S3-11).
- **(c) Enumerated divisions:** one EGS division had a possible omission from its division records, answered material (1/262; judged present in the division’s records by the disposers; Table S3-7); none in AAO (0/58) or NICE (0/124). Table 2 gives the structural indicator S.
- **(d) Structural check:** one finding, in NICE (P-1 0/1; P-2 1/1), on a source page not covered by the enumeration, answered not material.
- **(e) Version-level items:** not applicable in every guideline.

All counts are units with reviewer-judged answers, not verified encoding departures, and a finding may concern the source rather than the encoding; absence of findings is not a fidelity determination.

### 4.2. Instantiation yield (context)

Per guideline (traces in Supplement S1.7):

- **EGS:** none of 748 units. The linked candidate recommendations were diagnostic or follow-up statements; the recorded rationales of 15 excluded passages naming ocular hypertension and treatment class them as modality or procedure selection, one leaving open whether it bears on the initial decision (Supplement S1.4).
- **AAO:** two branches from 322 units, both resolving to *initiate treatment*: patients with ocular hypertension at high risk of developing POAG, and an eye likely to progress to POAG.
- **NICE:** one branch from 198 units (recommendation 1.4.4): newly diagnosed ocular hypertension with IOP of 24 mmHg or more, not associated with pigment dispersion syndrome, at risk of visual impairment within the person’s lifetime, resolving to *initiate treatment*.

All three branches were source-constrained, with choice rules declared at study level; none was author-defined. The remaining units stayed non-instantiated; each guideline’s encoding phase was closed by PI decision with these units non-terminal (Supplement S1.4).

### 4.3. Secondary outcomes

#### Finding categories

All exclusion findings were *exclusion questioned*; the division finding was a *possible omission* and the structural-check finding *not classified*.

#### Disposition profile

Denominators were all initial-phase findings: EGS 602 (601 exclusion findings and 1 division finding), AAO 216 (all exclusion findings) and NICE 153 (151 exclusion findings, the structural-check finding and the overlap finding). The outcome was no change for EGS 602/602 (100.0%), AAO 216/216 (100.0%) and NICE 152/153 (99.3%); every questioned exclusion was kept, under that standard reason. The one NICE change (1/153, 0.7%) corrected the source partition flagged by the structural check. In the exploratory rationale check, Reviewer B raised no finding on any exclusion rationale (EGS 0/2,109, AAO 0/234, NICE 0/179); that absence does not validate the exclusions.

#### Acceptance

Both representations passed. Passing is procedural and says nothing about fidelity.

#### Overlap (context)

On the 36 overlap items (12 per object: AAO 24, NICE 12), the two reviewers’ answers differed on one NICE item. Reviewer B judged material that the NICE representation records the recommendation as initiating treatment but omits that the recommended treatment is 360° selective laser trabeculoplasty (SLT); Reviewer A raised no finding. The authors kept the representation, because the frozen output vocabulary records direction, not modality. This finding is descriptive and outside the primary outcome.

#### Execution determinacy

All 1,536 inputs executed, with no validation failure (Table 3). Of 96 parent cases, AAO resolved 72 (75.0%) to *initiate treatment* and NICE 8 (8.3%); the rest were unresolved.

**Table 3.** Execution determinacy over the synthetic case space: 96 parent cases and 672 variants with one variable not established, for each representation.

| Output state | AAO parents | AAO variants | NICE parents | NICE variants |
|---|---|---|---|---|
| Resolved: initiate treatment | 72/96 (75.0%) | 432/672 (64.3%) | 8/96 (8.3%) | 24/672 (3.6%) |
| Resolved: observe without treatment | 0/96 (0.0%) | 0/672 (0.0%) | 0/96 (0.0%) | 0/672 (0.0%) |
| Unresolved | 24/96 (25.0%) | 240/672 (35.7%) | 88/96 (91.7%) | 648/672 (96.4%) |

EGS: not applicable (no executable representation). No admitted branch outputs *observe without treatment*, so 0 in that row holds by construction. AAO resolves whenever either branch fires; AAO-CD-0036-C02 needs only the case-supplied “eye likely to progress to POAG”, which accounts for most resolved AAO cases. Validation failures 0; unexecuted inputs 0. The counts describe a designed grid and carry no prevalence. Determinacy is not fidelity or clinical correctness.

#### Inter-encoder agreement

Exact agreement on the segmented source text was 83/83 (100.0%) in EGS, 70/74 (94.6%) in AAO and 60/60 (100.0%) in NICE (Table 4). Agreement on whether explicit source evidence for decision content was identified was 65/83 (78.3%), 50/74 (67.6%) and 44/60 (73.3%). Where both encoders recorded a proposition, its wording matched exactly in 13 of 81 pairs in EGS, 29 of 70 in AAO and 11 of 51 in NICE; in the remaining pairs the evidence spans overlapped.

**Table 4.** Inter-encoder agreement on duplicated units. Sample n_h / frame N_h: EGS 83/581, AAO 74/302, NICE 60/156. The frame is the retained candidate-proposition (Class A) units; with the dependency (Class B) objects (EGS 171, AAO 29, NICE 52) they make up the 752, 331 and 208 retained units of the canonical record. Dependency-link equality counts each link recorded for a duplicated Class B object as equal only if both encoders recorded it identically; the denominator is those links. Raw agreement n/N (%) [95% design-based interval]; for two-category estimands, κ and AC1 with the ordered contingency counts (both positive / Encoder 1 only / Encoder 2 only / both negative).

| Estimand | EGS | AAO | NICE |
|---|---|---|---|
| Segmented source text, exact match | 83/83 (100.0%) [96.0, 100.0] | 70/74 (94.6%) [87.7, 98.0] | 60/60 (100.0%) [95.5, 100.0] |
| Explicit evidence identified for source-specified decision content | 65/83 (78.3%) [68.7, 86.1]; κ 0.58; AC1 0.57; 30/1/17/35 | 50/74 (67.6%) [57.3, 76.5]; κ 0.39; AC1 0.36; 28/2/22/22 | 44/60 (73.3%) [63.5, 81.4]; κ 0.32; AC1 0.58; 38/0/16/6 |
| Explicit evidence identified for decision-authority allocation | 82/83 (98.8%) [94.0, 99.8]; κ 0.00; AC1 0.99; 0/0/1/82 | 73/74 (98.6%) [93.7, 99.7]; κ 0.66; AC1 0.99; 1/0/1/72 | 59/60 (98.3%) [92.9, 99.4]; κ 0.85; AC1 0.98; 3/0/1/56 |
| Competing reading raised at the stage-1 interpretation review | 83/83 (100.0%) [96.0, 100.0]; κ not computable; AC1 1.00; 0/0/0/83 | 74/74 (100.0%) [96.0, 100.0]; κ not computable; AC1 1.00; 0/0/0/74 | 60/60 (100.0%) [95.5, 100.0]; κ not computable; AC1 1.00; 0/0/0/60 |
| Proposition wording, exact match where both recorded a value (conditional) | 13/81 (16.0%) | 29/70 (41.4%) | 11/51 (21.6%) |
| Dependency links of duplicated dependency objects, exact equality (descriptive) | 34/484 (7.0%) | 9/99 (9.1%) | 23/143 (16.1%) |
| Semantic-slot state agreement, range over 14 slots | 48/83 – 83/83 | 58/74 – 73/74 | 46/60 – 60/60 |

Dependency objects record source material that changes the meaning of a retained proposition, for example material that operates as a condition. No benchmark is applied. Per-slot contingency tables are in Supplement Table S3-4. The EGS κ of 0.00 beside an AC1 of 0.99 for decision-authority allocation reflects one discordant unit among 83, the known prevalence effect on κ. A discrepancy is a difference between the two encoders’ sealed initial records of a duplicated unit, detected by the locked comparison tool. Identifying whether a unit carries explicit evidence for decision content had the lowest raw agreement of the four unconditional estimands shown. Agreement between encoders is not fidelity to the source.

#### Exploratory outcomes

No alternative encoding was admitted for execution, and no case fell in the stratum governed by source non-identification.

### 4.4. Post-hoc second reading (not registered)

Of the exclusions on which Reviewer B raised a finding, the second reader also raised one on 31/50 in EGS, 11/50 in AAO and 28/50 in NICE; of those without a finding, he raised none on 43/50, 18/18 and 28/28 (Table S3-12). This is agreement on raising a finding; unlike Reviewer B, he answered not material on 9, 1 and 12 of his findings. He judged one sampled passage (NICE) directly relevant to the initial treat-or-observe decision, and 0/100, 0/68 and 8/78 needed to preserve the meaning of a retained unit; the eight include the directly relevant passage and one linked to the executed NICE branch (Table S3-12). These judgements were not adjudicated.

## 5. Discussion

Few treat-or-monitor branches were instantiated (EGS none, AAO two, NICE one), and their primary review was uninformative at these denominators. In the descriptive overlap, the second reviewer judged material that the NICE representation omits the source’s treatment modality. This is a partial omission, the most common discrepancy that Gupta and colleagues reported for LLM-converted NICE rules [19]. Most findings came from what had been excluded at extraction.

### 5.1. What the scope restriction leaves out

Exclusion happened at extraction, separately from the admissibility gate. As the reviewer’s addendum reports, the counts describe source-stated decision content in excluded passages; they do not establish that every exclusion was correct or quantify clinical loss. The nine examples in Table S3-9, selected by a fixed rule, range from adjacent treatment decisions to diagnostic, procedural and patient-information content. A flag records that the reviewer read source-stated decision content in the passage, not that this content bears on the initial treat-or-observe decision. In the post-hoc second reading, flags were only partly reproduced; the second reader judged almost no sampled passage directly relevant to the initial decision, but, in his judgement, eight NICE passages were needed to preserve retained meaning, an unadjudicated signal on the exclusion rule’s second limb. The counts are reader-dependent descriptions rather than a stable measure. The protocol permits parallel per-guideline reporting but prohibits cross-guideline comparison; the post-hoc tabulation describes finding rates by material class but does not explain them. A review limited to the produced rules does not show what a scope restriction sets aside; whole-document exclusion review makes it visible and countable and, unlike a coverage ledger alone, adds a clinical judgement on each excluded passage.

Unlike translation-level assessment [19] or completeness evaluation against a gold standard [16], our contribution is narrower and methodological: it separates extraction exclusions, division-record review, executable review and disposition, and reports each before execution. No improvement over existing completeness or fidelity methods was measured, and counts are not numerically comparable across these designs.

### 5.2. Execution and agreement

On the synthetic grid, neither representation ever resolved to observation, because each admitted branch states when to treat and none states when to observe. NICE 1.4.3, which advises against treatment for people not at risk of lifetime visual impairment, was not admitted, because the registered rules did not cover negative forms (Supplement S1.7). Judgement-bearing conditions such as “at high risk” entered as supplied case values: the engine evaluated them but did not ascertain them.

The encoders agreed on segmentation but less on extracted wording and dependency links; different wording is not by itself a disagreement about meaning. Reconciliation adopted Encoder 2’s value for most discrepancies (Supplement S1.8); because Encoder 1 alone encoded the non-duplicated units, under-identification of decision content there could have lowered yield, a hypothesis these data cannot test.

### 5.3. Limitations

- **Pipeline-specific yield.** The gate defines a restricted translatability, and many non-instantiated units remain in non-terminal states; the yield describes this pipeline, not a limit on computability. Phase closure was an unregistered PI implementation decision (Supplement S1.5), and one EGS passage was excluded with its bearing on the initial decision recorded as unsettled.
- **Single-author steps.** Source non-identification and scope-coverage determinations governing case generation were made by one author, with only an AI procedural check.
- **Disposition.** The encoders decided dispositions without a third adjudicator; review detected disagreements but did not resolve them.
- **Reviewers.** Outside the overlap and the post-hoc second-reading sample, each unit received one clinical judgement; exclusion and division counts do not estimate between-reviewer reproducibility. One reviewer completed the exclusion review with a streamlined instrument; the primary counts remain single-rater observations made under these conditions.
- **Post-analysis clarification and construct ambiguity.** Reviewer B reported his interpretation of exclusion materiality after analysis in a separate dated addendum, without item-specific adjudication; the reported findings remain the original sealed judgements. “Loss … as represented” remains under-specified for omitted units. The second reader’s answers also show uncertainty at the domain boundary (Table S3-12).
- **Review conduct.** The reviewer-contact log was compiled retrospectively; the PI was the only contact point, and four interpretive queries were not answered on the merits, under the contact rule.
- **Retained-object frame.** The denominator of component (a) rests on a PI reading of an ambiguous registered rule, made before release without a governed route; under the alternative reading the component would have covered 539 objects (Supplement S1.5).
- **AAO assessment universe.** The AAO scope-coverage assessment was limited to the two executed units.
- **No back-translation** or source-unprimed reading of the executable was performed.
- **Prior exposure.** Both encoders knew the earlier version of the project (v0.1), and Reviewer A had reviewed material from it; the initial reviewers were naïve to v0.2 material.

Supplement S2 states the further limitations the protocol requires; Supplement S1 lists every recorded deviation.

### 5.4. Implications

Computable-guideline programmes and evaluations of LLM converters should report what was excluded and why, how much decision content their scope sets aside, how reproducibly exclusions were judged and how review findings were disposed of, complementing recent guidance for developing computable guidelines [26]. Released records are these encoders’ and reviewers’ documented records, not adjudicated ground truth.

## 6. Conclusion

Across three ocular hypertension guidelines, the pipeline instantiated three treat-or-monitor branches without author-defined choices; their primary review was uninformative at its denominators, and the overlap review judged one omission material, a treatment modality that the direction-only output could not carry. Whole-document exclusion review made the content set aside by the scope restriction visible and countable, but a post-hoc second reading only partly reproduced its flags; it judged almost no sampled passage directly relevant to the initial decision, yet some NICE passages needed to preserve retained meaning. Exclusion review is a candidate component of fidelity assessment whose reproducibility must be established before it is used as a measure.

## Supporting information

Supplementary Data S4 - Freeze-register digest

## CRediT authorship contribution statement

Andrea Ragosta: Conceptualization, Methodology, Investigation (Encoder 1), Formal analysis, Data curation, Project administration, Writing – original draft, Writing – review & editing. Francesco Bruni: Investigation (Encoder 2), Validation (transcription verification), Writing – review & editing. Both authors jointly dispositioned the review findings and approved the final manuscript.

## Ethics statement

The study analysed published guideline documents, synthetic cases and judgements contributed by three ophthalmologist reviewers serving as study personnel. It used no patient data. All three reviewers consented to being acknowledged, and the two initial reviewers to publication of their verbatim findings and messages; none was compensated. Ethics committee review was not sought.

## Data availability

The registered protocol, frozen records, freeze register, analysis code, case space, execution records and the post-hoc second-reading files will be released on OSF (osf.io/bhmz2) when the embargo ends after peer review, and are available to editors and reviewers on request. Full guideline documents are not redistributed. A digest of the freeze register is provided as Supplementary Data S4.

## Declaration of generative AI and AI-assisted technologies in the manuscript preparation process

No AI tool is an author.

- Claude (Anthropic) structured the initial encoding and reconciliation packages.
- Codex (OpenAI) compiled the AAO workbook, proposed options for 34 encoding positions in reconciliation, acted as procedural gatekeeper before each governed act and recomputed the stage-10 outputs in a separate session.
- Kimi (Moonshot AI) audited the review instrument and outcome specification before reviewer release.
- Codex, Kimi and Claude Fable surveyed candidate journals.
- AI generated the engine, validators, analysis scripts and review tools to the authors’ specifications, verified by validator tests, byte-identical replay and independent recomputation, and assisted with drafting and language editing.
- Figures 1–2 and the graphical abstract were drawn with plotting scripts written with AI assistance.
- One clinical reviewer used scripts and AI to organise passages and transcribe decisions (Supplement S1.6).

Encoder 1 reviewed and adopted every AI output; the authors decided all content and are responsible for it.

## Acknowledgements

We thank Dr A. Farina and Dr R. Llanas Reus, who served as the independent clinical reviewers, and Dr J.L. Macaya Pascual, who performed the post-hoc second reading. They are not authors.

