## Supplementary Material S1-S3 for "Clinical review before execution of executable guideline encodings: a prospective registered study of three ocular hypertension guidelines"

Contents: S1 Registration, governance, freeze points, instantiation, deviations, branch traces and inter-encoder discrepancies. S2 Statements and limitations required by the registered protocol. S3 Full results tables (S3-1 to S3-12). Supplementary Data S4 (separate PDF file): digest of freeze-register entries 1–285.

All S3 values except Tables S3-11 and S3-12 are transcribed from the committed stage-10 output files (register entry 284) and were reconciled with those files; any discrepancy identified will be corrected. Table S3-11 is a post-hoc tabulation from the sealed exclusion workbook; Table S3-12 reports the post-hoc second reading. Class A denotes retained candidate-proposition units and Class B dependency objects (PG-03 record-depth classes). Results are per guideline; no pooled or cross-guideline figure is given. A blank machine field is shown as NOT COMPUTABLE or NOT APPLICABLE according to its row status.

#### Supplement S1 — Registration, governance, freeze points and deviations

##### S1.1 Registration

- **Registry:** OSF Registries, registration **bhmz2** (https://osf.io/bhmz2), Open-Ended Registration template.
- **Registered:** issued on 1 October 2026 (register entry 8, milestone M-1; verified at entry 9), before v0.2 segmentation and encoding began. The registration was submitted and registered the same day (registry record: date created and date registered, 1 October 2026).
- **What was registered:** package version 3.0, frozen and approved on 30 September 2026.
- **Embargo:** set to 30 June 2030, the date entered in the registry's embargo field; to be lifted at the end of peer review.
- **DOI:** none assigned to the registration at the time of writing.
- **Not a Registered Report:** the study was not submitted in that format.
- **Registration limits recorded at the time (G-1 to G-3):**
    - G-1: the registered summary wording differs from the package wording.
    - G-2: the 58 registered files were uploaded inside one archive.
    - G-3: full documented pre-submission compliance is not certified. One check ran without a filed record, there was no separate pre-submission confirmation, and the sole-administrator setting was confirmed only after submission.

**What was fixed when.** The registration fixed the question, sources, encoding rules, acceptance criteria, outcomes and analysis. Implementation material (tools, schemas, validators, review instrument, engine and analysis scripts) was drafted after v0.2 source contact and locked no later than first use, at the freeze points in S1.2. Post-exposure specifications of the study materials were adopted through the governed amendment route and are dated below. Two later PI decisions had no governed route: the reading of the retained-object frame rule and the encoding-phase closure rule (S1.5, rows 6 and 17).

**Registry updates.** The registration has three approved updates. Each appends text to the registration summary with a stated justification; the registered files are unchanged.

| Date | OSF revision identifier | Content |
|---|---|---|
| 18 Oct 2026 | 6ad4a85c70298170745cc982 | VC-1: PG-11 resolution record, which governs the encoders' joint resolution sessions in reconciliation, v0.1.2 → v0.1.3 (change record P26-10 v1.1; register entries 100–104) |
| 18 Oct 2026 | 6ad4acfe8d86dc5e72ceccb6 | VC-2: stage-5 amendments to PG-11 clauses A–C and to Charter §1 (register entries 136–140) |
| 28 Oct 2026 | 6ae2467d1b4971442bcf4b8e | The amendments listed below |

VC-1 and VC-2 were adopted after source exposure and used during reconciliation (stage 5) before they were entered on OSF; each entry discloses this delay and states that it is not retroactive registration. The 28 October entry records the amendments made after encoding and before any material was released to reviewers:

- PG-16 A3 (term of comparison for omission detection);
- PG-18 A2;
- PG-22 A1 (claim limit for component (c); no change to any denominator, class or bound);
- PG-16 A4 with PG-18 A3 (review instrument).

These amendments were frozen in the register at entries 227–232 and verified at entries 233–235. PG-16 A4 with PG-18 A3 revised the instrument layout (passages grouped by division, prefilled locators, one-click item outcomes, finding fields only for raised findings); none of the amendments changed the exclusion task or the materiality definition.

**Materiality definition and exclusion task, verbatim as frozen in the reviewer manual (M0016) before reviewer release.** Definition: "A loss, addition or distortion of the anchored source's meaning in the representation is **material** if it changes, as represented, what the source states about any of the following: (i) the direction of a recommendation or decision; (ii) its stated strength or certainty; (iii) the population, condition, threshold or scope to which it applies; (iv) the criteria or conditions for applying it. For a documented exclusion or an enumerated division, loss includes decision content of these kinds that the source states and the representation omits. Any other loss, addition or distortion is not material under this definition." Question on every finding: "Does this finding identify a loss or distortion of the source's meaning in the representation that you judge material under the frozen definition?" Exclusion task: "Each row is material the study excluded, with its location only. Read it in the PDF and judge whether excluding it loses decision content."

**Retained-object frame rule (PG-16, OV-1), verbatim as registered.** "For each body h, the frame is the set of in-scope retained objects of all output-intended lineages (including any ADMITTED_ALTERNATIVE) at the review-version freeze point (DP v0.2.1 §4.L). Operationally it is identical to R-A's C-1 assignment register for body h, generated mechanically from the frozen review versions and hash-frozen at that point. N_h = size of that register. Objects added or changed before the review-version freeze, including via a disposed PARTITION_FAILURE, are therefore inside the frame."

**Segmentation rule (PG-03 §10.1), verbatim as registered.** "A child unit is created for each separately assertable guidance-bearing proposition within a parent." A guidance-bearing proposition "asserts, defines, recommends, permits, prohibits, conditions, qualifies, excepts, or assigns decision authority in relation to clinical care or to the interpretation/applicability of clinical guidance"; it is separately assertable when "the source makes a distinct claim that could hold or not hold independently of the parent's other claims". The review instrument and the outcome-computation specification were locked (manifest M0016) before reviewer release (entry 247).

The protocol describes its integrity controls as prespecified internal freezes, with external signed time evidence (RFC 3161) at named decisive steps. It does not claim that every artefact was externally time-stamped at creation. A verified time-stamp token is evidence, conditional on the time-stamping authority and its trust chain, that a manifest digest existed no later than the token time. It does not prove the true time of artefact creation, the completeness of unrecorded attempts, or that an artefact was never replaced before the token.

#### S1.2 Freeze points and register entries

All study artefacts were committed by SHA-256 to an append-only freeze register (PG-26). The register held 286 entries at the time of writing; entries 1–285 cover the study from the initial lock to the analysis, and later entries record publication administration. Supplementary Data S4 gives, for entries 1–285, the entry number, entry date (UTC), entry type, governing provisions, manifest identifier, subject identifiers with their full SHA-256 values, the previous-entry hash and the SHA-256 of each register line, so that every prespecified count and record can be traced to its commitment; the post-hoc Tables S3-11 and S3-12 carry their own provenance (sealed-workbook hash; sampling, returned and analysis files with their hashes). `entry_line_sha256` is the SHA-256 of the raw register line (UTF-8 JSON, without its trailing newline); each entry's `prev_entry_sha256` equals the preceding entry's `entry_line_sha256`, so the predecessor links can be checked from S4; recomputing the line hashes themselves requires the raw register lines, which will be released with the register. Entry dates are given to the day; full timestamps are in the register to be released on OSF. Entries after 285 record publication administration and are not listed. A hash proves identity and integrity, not correctness, and custody is a procedural control, not an independence mechanism.

| Freeze point | Content | Register entries |
|---|---|---|
| Initial lock M0001 | Registered package and tools | 1–3 |
| FP-A (M0002; replacements M0007, M0011) | Pre-encoding freeze and its governed replacements | 5–7; 100–104; 138 |
| Milestone M-1 | Registration issued and checked | 8–9 |
| Stage 4 | Coverage enumeration; duplicate-encoding seed; frame snapshot | 12–20 |
| Stage 5 | Sealed initial encodings; decisive step DS-R0; reconciliation R1–R6; canonical record R6′ | 52–82; 151–157 |
| Stage 6 locks | LT-2 lock M0012; M0013; decisive step REMEDY | 161; 164–170 |
| Rule families (registered before inspection) | NICE; EGS; AAO | 173–184; 185–204; 205–224 |
| Amendments to omission detection and review instrument (M0014–M0016) | See S1.1 | 227–235 |
| FP-B (M0017) | Frozen review versions RV-NICE-V1, RV-AAO-V1 | 236–240 |
| Review | Reviewer matching; release; sealed initial-phase judgements; zero-entry supplementary phase | 245–258 |
| Disposition | Disposition records; partition correction; audit | 259–266 |
| Acceptance | PASS determinations (procedural) | 267–268 |
| FP-C1 (M0018) | Stage-8 frozen representations | 269–271 |
| FP-C2 (M0019 LT-2 tools; M0020 case-space specification) | Engine, validator, analysis plan and scripts locked; case space | 272–277 |
| FP-C3 (M0021) | Case list and execution inputs | 278–280 |
| FP-D (M0022) | Sealed execution records | 281–283 |
| Analysis | Stage-10 outputs committed when produced; independent recomputation | 284; 285 |

**External time evidence.** Two RFC 3161 tokens (Sigstore) were obtained at decisive steps. The values below are as recorded in the register; the register-entry date in S4 is distinct from the token generation time. A token dates its manifest from the token time only.

| Decisive step | Manifest SHA-256 | Token generation time (UTC) | Token response SHA-256 | Token entry | Verification entry, recorded result, check-record SHA-256 |
|---|---|---|---|---|---|
| DS-R0 | dd1b8bc2fb2c76a335f595d020787d6299626e4c8c2f4e26363101169e3c33f6 | 2026-10-03T14:14:12Z | 44d2bfd4bc81f644197330b18450549a3758f9219b6cda24302175bc5b88a96b | 60 | 61; PASS; 03af678ea77d55d5d7cec478af1e06c93b4aed639bccc9db68171b5f5d1226d8 |
| REMEDY | 934455640a981591d675a4d8e8787223f919b46d14495e16dc56e203416a839c | 2026-10-18T09:44:42Z | f5902624479fd62f99378a9460c2d32db25ee97df13cec73bbea41b3d75e0fec | 166 | 167; PASS; 50bcf9584282e01071e57655205e284cf87111aa97489da129144e975a315c66 |

**Limits of the register.** The initial-lock manifest evidences its contents only from its own commitment, not from any earlier commitment. Routine commitments between decisive steps were internal; an unrecorded or coordinated replacement before the next token cannot be ruled out. Replaced lock manifests (M0003→M0008, M0006→M0009, and the M0011 consolidation) are recorded as replacements with their predecessors, and superseded records are kept rather than deleted. S4 lists commitments; it is not by itself a complete account of every change, invalidation or stop. Those histories are in the underlying records (replacement manifests, gatekeeper reports, PI acts and the state log), which are not reproduced in this supplement and will be released with the study records.

#### S1.3 Roles

| Role | Person |
|---|---|
| Principal investigator; Encoder 1 (principal semantic encoder; author of the executable encoding) | Andrea Ragosta |
| Encoder 2 (independent initial semantic record on the duplicate sample; transcription verification of the executable versions) | Francesco Bruni |
| Joint reconciliation and disposition authority | Andrea Ragosta and Francesco Bruni, jointly |
| Reviewer A (retained objects) | Adrian Farina |
| Reviewer B (exclusions, rationale check, omission detection, structural check, overlap) | Raimundo Llanas Reus |

**Prior exposure.** Both encoders know the earlier v0.1 study: Encoder 1 authored its encoding and execution chain, and Encoder 2 completed its clinical review. Reviewer A had previously reviewed material from the historical v0.1 study; this exposure is disclosed, and no claim of naïveté to the historical project is made. Per the PI's determination, the reviewers were naïve to v0.2 material and v0.2 semantic design.

**Reuse of earlier infrastructure.** RU-0 (PG-28) prohibited screening or reusing v0/v0.1 components and repairing, rerunning, regenerating or benchmarking v0.1 results. No reuse is recorded in the study records; final adherence was not independently verified against the complete implementation, change and defect records. Non-reuse of code does not remove knowledge of v0.1 (Supplement S2.5).

AI use is described in the AI usage declaration of the manuscript.

#### S1.4 Instantiation by guideline (context)

| Guideline | Eligible units | Executable branches (frozen) | Non-instantiated units, as recorded at phase closure |
|---|---|---|---|
| EGS | 748 | 0 | 83 preliminary transformations in a non-directional follow-up family, outside the executable format; 657 preliminary non-terminal *no admissible operationalisation*; 8 suspended. No transformation in the four directional families. The linked candidate recommendations were diagnostic or follow-up statements, not treat-or-monitor decisions |

| Guideline | Eligible units | Executable branches (frozen) | Non-instantiated units, as recorded at phase closure |
| --- | --- | --- | --- |
| AAO | 322 | 2 (AAO-CD-0009-C06 and AAO-CD-0036-C02; S1.7) | Source-bounded choice spaces without selection (3); descriptive transformations (68); one co-equal transformation without record (1); stops (3); 244 preliminary non-terminal *no admissible operationalisation*; one suspended (1) |
| NICE | 198 | 1 (NICE-CD-0032-C01, recommendation 1.4.4; S1.7) | 3 decided not instantiable at the gate; 11 suspended pending governance; 183 preliminary non-terminal *no admissible operationalisation* |

Within each guideline the categories do not overlap and sum to the eligible units. All three branches are *source-constrained* with a choice rule declared at study level before inspection. No unit reached a terminal state of non-instantiability, so these counts describe what this pipeline instantiated, not what cannot be made executable.

**Meaning of the recorded states.** *Preliminary non-terminal "no admissible operationalisation"* is Encoder 1's first-pass disposition that no admissible executable form existed under the registered rule families; it was not taken through the procedure that would make it terminal. *Suspended* (or *pending governance*) units depended on a governance decision that was not taken before closure. *Decided not instantiable at the gate* records a dated gate decision by the PI as Encoder 1, with the record left incomplete rather than terminal. The other labels (preliminary transformations, choice spaces without selection, descriptive transformations, stops) are the recorded transformation states, shown as recorded.

**Phase closure.** Each guideline's encoding phase was closed by PI decision (NICE 22 October, EGS 26 October and AAO 28 October 2026) under the stop rule that the PI adopted on 22 October 2026 from a project reset report of 21 October, which reviewed workload and governance and limited further governance work (no artefact without a governing clause; one internal check; at most one external audit). The rule's status is given in S1.5, row 17. At the gate, what was not demonstrated remained non-instantiated or pending; no terminal state was assigned, nothing was promoted and no family was added retroactively. Retained but non-instantiated units are reported here as instantiation yield, descriptively; they are not counted in the primary outcome.

**EGS units carrying a recommendation-strength label at the gate.** The 16 EGS units that reached the gate with a linked strength label were test lists or diagnostic statements; none states when ocular hypertension is treated or observed. No retained EGS unit was transformed in the four directional families. The study did not separately enumerate every EGS passage that states when ocular hypertension is treated or observed; excluded passages whose recorded rationale names OHT and treatment are listed after the next table.

| Units | Source question (division) | Text (L2, abridged) | Label | Recorded outcome and reason |
|---|---|---|---|---|
| EGS-CD-0029-C01 to C07 | Q.V.1 Recommended tests at first assessment (EGS-CD-0029) | Visual acuity and refraction; slit lamp; gonioscopy; tonometry; CCT; visual field testing; optic nerve head, RNFL and macula assessment | strong (CCT weak) | Not instantiated (preliminary non-terminal): list item without an autonomous decision proposition |
| EGS-CD-0029-C08 | Q.V.1 (EGS-CD-0029) | "OCT … can be useful, but the diagnosis of glaucoma cannot be made on the basis of OCT alone" | weak | Not instantiated: a diagnostic limitation, not a negative form on the treatment branch |
| EGS-CD-0035-C01 to C05 | Q.V.7 Recommended tests for monitoring (EGS-CD-0035) | Visual acuity; visual field testing; optic disc and RNFL examination; tonometry; OCT imaging | strong (OCT weak) | Not instantiated (preliminary non-terminal): list item without an autonomous decision proposition |
| EGS-CD-0035-C06 | Q.V.7 (EGS-CD-0035) | "Repeated gonioscopy in some circumstances" | weak | Stop: literal non-directional examination or follow-up transcription |
| EGS-CD-0030 | Tonometry (EGS-CD-0030, sheet 26) | "Consensus could not be reached about which alternative tonometers … can be used" | weak | Not instantiated (preliminary non-terminal) |
| EGS-CD-0032-C01 | Q.V.4 Central corneal thickness | "CCT may be useful for baseline risk profiling" | weak | Not instantiated (preliminary non-terminal) |

**Excluded EGS passages whose recorded rationale names OHT and treatment.** Selected mechanically: the limb-(i) exclusion rationale recorded at extraction mentions OHT or ocular hypertension together with a treatment term (treatment, treat, laser, trabeculoplasty, SLT, MIGS, surgical procedure), after removing the standard phrase "initial treat-vs-observe decision". The list is not a clinical judgement of which passages state when ocular hypertension is treated or observed; that judgement was not made in this study. All were kept at disposition. The rule searches the recorded rationale text only: an excluded passage whose rationale does not name OHT and a treatment term would not appear, so the list is a lower bound on such passages, not an enumeration of them.

| Excluded unit | Location (PDF sheet; section, abridged) | Recorded exclusion rationale, limb (i) | Reviewer B outcome |
|---|---|---|---|
| EGS-CD-0019-C05 | PDF sheet 21; …mendations > I.3.1 New questions for this edition > Q2. What is the recommended first-line treatment for POAG? | Although OHT is named, the claim concerns laser as the modality of initial treatment (modality selection), outside the domain. | FINDING-RAISED |
| EGS-CD-0094-C01 | PDF sheet 62; …s comparing treatment in open angle glaucoma > I.7.3.8 Laser in Glaucoma and Ocular Hypertension Trial (LiGHT) | Comparison of first-line treatment modalities/intensity in patients already selected for treatment; modality choice after the treatment decision is outside the domain. (SLT vs drops as first-line treatment in newly diagnosed OHT/OAG). | NO-FINDING-RAISED |
| EGS-CD-0094-C02 | PDF sheet 62; …s comparing treatment in open angle glaucoma > I.7.3.8 Laser in Glaucoma and Ocular Hypertension Trial (LiGHT) | Comparison of first-line treatment modalities/intensity in patients already selected for treatment; modality choice after the treatment decision is outside the domain. (SLT vs drops as first-line treatment in newly diagnosed OHT/OAG). | NO-FINDING-RAISED |
| EGS-CD-0094-C03 | PDF sheet 62; …s comparing treatment in open angle glaucoma > I.7.3.8 Laser in Glaucoma and Ocular Hypertension Trial (LiGHT) | Comparison of first-line treatment modalities/intensity in patients already selected for treatment; modality choice after the treatment decision is outside the domain. (SLT vs drops as first-line treatment in newly diagnosed OHT/OAG). | FINDING-RAISED |
| EGS-CD-0094-C04 | PDF sheet 62; …s comparing treatment in open angle glaucoma > I.7.3.8 Laser in Glaucoma and Ocular Hypertension Trial (LiGHT) | Comparison of first-line treatment modalities/intensity in patients already selected for treatment; modality choice after the treatment decision is outside the domain. (SLT vs drops as first-line treatment in newly diagnosed OHT/OAG). | NO-FINDING-RAISED |
| EGS-CD-0094-C05 | PDF sheet 62; …s comparing treatment in open angle glaucoma > I.7.3.8 Laser in Glaucoma and Ocular Hypertension Trial (LiGHT) | Comparison of first-line treatment modalities/intensity in patients already selected for treatment; modality choice after the treatment decision is outside the domain. (SLT vs drops as first-line treatment in newly diagnosed OHT/OAG). | NO-FINDING-RAISED |
| EGS-CD-0094-C06 | PDF sheet 62; …s comparing treatment in open angle glaucoma > I.7.3.8 Laser in Glaucoma and Ocular Hypertension Trial (LiGHT) | Comparison of first-line treatment modalities/intensity in patients already selected for treatment; modality choice after the treatment decision is outside the domain. (SLT vs drops as first-line treatment in newly diagnosed OHT/OAG). | NO-FINDING-RAISED |

| Excluded unit | Location (PDF sheet; section, abridged) | Recorded exclusion rationale, limb (i) | Reviewer B outcome |
|---|---|---|---|
| EGS-CD-0094-C07 | PDF sheet 62; …s comparing treatment in open angle glaucoma > I.7.3.8 Laser in Glaucoma and Ocular Hypertension Trial (LiGHT) | Comparison of first-line treatment modalities/intensity in patients already selected for treatment; modality choice after the treatment decision is outside the domain. (SLT vs drops as first-line treatment in newly diagnosed OHT/OAG). | NO-FINDING-RAISED |
| EGS-CD-0094-C08 | PDF sheet 62; …s comparing treatment in open angle glaucoma > I.7.3.8 Laser in Glaucoma and Ocular Hypertension Trial (LiGHT) | Primarily a first-line modality statement (SLT), but phrased as routinely offering treatment to all newly diagnosed OHT patients; frozen wording does not settle whether this bears on the initiate-vs-observe decision or only on modality after treatment is selected. | FINDING-RAISED |
| EGS-CD-0240-C02 | PDF sheet 188; …Part II > II.3 Treatment > II.3.12 Laser surgery > II.3.12.2 Laser trabeculoplasty | Laser trabeculoplasty as primary/first-line treatment (incl. high-risk OHT/OHT): choice of modality once treatment is selected (lasers as treatment selection), outside the domain. | NO-FINDING-RAISED |
| EGS-CD-0240-C04 | PDF sheet 188; …Part II > II.3 Treatment > II.3.12 Laser surgery > II.3.12.2 Laser trabeculoplasty | Laser trabeculoplasty as primary/first-line treatment (incl. high-risk OHT/OHT): choice of modality once treatment is selected (lasers as treatment selection), outside the domain. | NO-FINDING-RAISED |
| EGS-CD-0242-C26 | PDF sheet 193; …Part II > II.3 Treatment > II.3.13 Cyclo-ablative procedures | Laser trabeculoplasty as primary/first-line treatment (incl. high-risk OHT/OHT): choice of modality once treatment is selected (lasers as treatment selection), outside the domain. | FINDING-RAISED |
| EGS-CD-0248-C05 | PDF sheet 196; …t > II.3.14 Incisional surgery > II.3.14.2 Techniques > II.3.14.2.4 Additional/alternative surgical techniques | Mentions OHT but concerns selection/indication of a surgical procedure (MIGS); surgical-treatment selection is outside the domain. | NO-FINDING-RAISED |
| EGS-CD-0248-C27 | PDF sheet 198; …t > II.3.14 Incisional surgery > II.3.14.2 Techniques > II.3.14.2.4 Additional/alternative surgical techniques | Mentions OHT but concerns selection/indication of a surgical procedure (MIGS); surgical-treatment selection is outside the domain. | FINDING-RAISED |
| EGS-CD-0248-C29 | PDF sheet 198; …t > II.3.14 Incisional surgery > II.3.14.2 Techniques > II.3.14.2.4 Additional/alternative surgical techniques | Mentions OHT but concerns selection/indication of a surgical procedure (MIGS); surgical-treatment selection is outside the domain. | FINDING-RAISED |

#### S1.5 Deviations and incidents

Every recorded deviation is listed below with its date, the affected record (register entry and, where one exists, the SHA-256 of the owning record), what was done differently, the actual reason and whether a governed route authorised it. No deviation is described as prespecified. Dates are those of the events; the corresponding register entries may be dated later, at commitment.

| # | Date | Affected record (register entry; SHA-256) | What happened | Reason as recorded | Authorisation / status |
|---|---|---|---|---|---|
| 1 | 30 Sep–1 Oct 2026 | Registration (entries 8–9; limits G-1–G-3) | Summary wording difference; single-archive upload; pre-submission compliance not fully documented | Registry interface; process gaps | No governed route; disclosed limitation |
| 2 | 18 Sep 2026 (source lock) | NICE source file (PG-02 source-lock record, registered package RP-02a §4.3; locked file 1806de5bbeceb5a0b2febad86ef3c900d97350dc0bea62d59e06d00b4d691c3c) | Automated download failed; file obtained outside the acquisition manifest; route attested by the PI | Server refusal (HTTP 403) | No governed route; documentation gap; file identity verified by hash |
| 3 | 3–4 Oct 2026 | Encoder-1 reconciliation positions (entry 75; deviation record 6715a548ff41c3eae59b9ab80c27c0796d5f3d71b54ff49c0585bfa3be6504d1) | 34 positions derive from consultative proposals by Codex, outside its role | Consultative use during reconciliation | Recorded deviation; Codex not independent for those 34 positions |
| 4 | 5 Oct 2026 | Stage-1 session S1-SESSION-01 (entries 96–98; returned form a01e2bba0c9fe6fe5a4043a5516f958c3aabbedb4e249d8704c42046458508f2) | Session started before verification of its preparation; discordant human declarations | Sequencing error | Accepted administratively by the PI, not certified |

| # | Date | Affected record (register entry; SHA-256) | What happened | Reason as recorded | Authorisation / status |
|---|---|---|---|---|---|
| 5 | 27 Oct 2026 | AAO Encoder-1 workbook as returned (6b48f5ca10817d1da58d0b255b218f7cb765aac96d3a4f2b7eb8549a10c4072a); the AI tool was subsequently identified as Codex | Compiled with AI assistance outside the recorded role structure; provenance corrected by the PI | Workbook formatting and compilation | PI correction; disclosed in the AI usage declaration |
| 6 | 28 Oct 2026 | Retained-object review frame (release, entry 247) | The PI first chose the reading 'all retained objects of AAO and NICE' (539), then, on the same day and before release, the reading 'objects encoded in the frozen executable representations' (3: AAO two, NICE one) | PI's choice to shorten the review (instruction recorded verbatim, 28 Oct 2026) | PI reading of the registered rule; the independent pre-release AI audit (Kimi, role A-11; M0016 freeze act, entry 231) found the adopted reading compatible with the frozen records. No governed route. Deviation affecting component (a) of the primary outcome: its denominator rests on this reading |
| 7 | 22–28 Oct 2026 | Reviewer recruitment and contact (contact and query log, entry 267; 276a488c830adcb50a59e81e2be26d46c3414daf6c7b220d14387fb119a0fc15) | Verbal availability check; invitation sent by messaging app; log compiled retrospectively from verbatim records and confirmed by the PI | Practicality | Disclosed limitation (main text); gatekeeper observation answered |
| 8 | 29 Oct 2026 | Reviewer A queries (same log, entry 267) | Five queries: one procedural, answered by re-sending the reviewer manual; four on study interpretation, not answered on the merits under the contact rule (reply: the judgement belongs in the workbook). Replies sent to both reviewers | Reviewer requests | Within the reviewer manual and contact rule; disclosed limitation (main text) |
| 9 | 29 Oct 2026 | Supplementary-phase (R-II) records (corrected records, entries 255–256; eligibility determination 682cdf35f32329de4719ea3e297cee6b013a2cbec7d5adbbdcd848c8076bf3d1) | The notice and zero-entry records preceded the eligibility determination and did not name the versions; the procedural check failed; the sequence was corrected and re-recorded; the earlier records are kept, not relied upon | Sequencing error | Corrected through the governed route |
| 10 | 29–30 Oct 2026 | NICE source partition PF-NICE-0001 (entry 260; final act cc60fac462601212bd992e7179eca613beecadce182a7235364fcd7ff77ed320) | Structural check found PDF sheet 34 outside the enumeration; partition corrected | Structural-check finding | Governed correction (partition-failure procedure; DSP-CHANGE) |
| 11 | 30 Oct 2026 | NICE representation, overlap finding DSP-RB-OV-001 (entry 262; owner act 18f424977a52ca939d9c1a44e64195e906ebcfb0dd0074590f3b5807314c7e3a) | Material omission of 360° SLT; no successor version, because the frozen two-value output vocabulary cannot carry treatment modality | Design limitation exposed by review | Joint owner act through the governed route; reported in main text |
| 12 | 30 Oct 2026 | Committed R1 comparison table read during analysis preparation (a7d2a75297a5585eab608136994fc4bb0aa94e166850cb08f0dff59e7e4ddc4c); no artefact of the printed output was kept; no register entry | While designing a script, the preparer printed the per-component status vocabulary of the committed reconciliation table (no count or rate); the AC1 closed-scale choice was made afterwards | Preparer error | Disclosed; procedural check agreed |
| 13 | 30 Oct 2026 | Register adapters F280 (before fix b42806c70f6337687945bcc70721e17275ce7591222ce048fcaa36deed2f9ba3; fixed 7053843d6c20e1473478a1d90beb996d8fd0b18ea2ddc57228c911f0906025ea) and F281_282 (before fix e53bee39298e278c6e1c9659a3b7a561320f3599bac7032a7b9ab2afbf49ce3f; fixed db608d15b5d376b3fb249ff58288b3668cba722d1bacc9b4cda743281e70cf41); executed adapters with the same gap: F272_275 (e8cd1ddc58a31e053d1cdec3e67a6bfe1f1f8b6f83c5c1e92c400f366aeac5b7) and F278_279 (f74a1a8c96db11c9c6b0486eac473c15280e0fa2426aeb491aabcefbea255216) | Guard gaps in link and subject checks, found by the procedural check and fixed before use; the two executed adapters had the same gap; entries 271, 277 and 280 were verified to have correct subjects and links | Preparer error | Fixed before use; no effect on entries |

| # | Date | Affected record (register entry; SHA-256) | What happened | Reason as recorded | Authorisation / status |
|---|---|---|---|---|---|
| 14 | 30 Oct 2026 | Stage-10 inputs (analysis run sheet; outputs entry 284) | 20 of 56 inputs (stage-10 tools, input maps, verifier, unit-list workbooks, representation records) pinned by hash in the run sheet rather than in a lock manifest | Implementation | PI authorisation of the run sheet; disclosed |
| 15 | 30 Oct 2026 | Stage-10 replay (entries 284–285) | Replay interrupted by the execution environment (background processes ended); resumed under PI authorisation; full replay byte-identical | Technical | PI authorisation; disclosed |
| 16 | 31 Oct 2026 | Reviewer B clarifying addendum (filed separately, not in the register; 6a5e6f5dfd963c1aaad57597d2b1834e57d2975b39b0daca7835621c6098a691) | Clarification obtained after the analysis; the sealed record is unchanged | Request for factual clarification for reporting | Separate dated record (S1.6) |
| 17 | 22 Oct 2026 | Encoding-phase closure rule (project record; not in the freeze register) | The PI adopted the stop rule of a project reset report; each guideline's encoding phase was then closed (NICE 22, EGS 26, AAO 28 October) with the remaining units non-instantiated and non-terminal | Workload and project reset | PI implementation decision outside the registered protocol; not a registered amendment; no governed route; not prespecified (S1.4) |

#### S1.6 Reviewer B clarifying addendum (31 October 2026)

The addendum was received by email on 31 October 2026 and filed as a separate dated record. The specific tools and prompts Reviewer B used were not recorded. The sealed X-1 workbook is unchanged (SHA-256 1e641ffd6ab3bec361bd64c7c87364906dd8d46a57dc8219f30982220f13fc41). It clarifies the meaning of the recorded answers after the analysis; it is not an item-level adjudication, and the reported findings remain the original sealed judgements.

##### Verbatim (Spanish):

1. Utilicé una fórmula estándar para los hallazgos: «Excluding this passage omits decision-relevant source content concerning [sección]: “[pasaje fuente]”». Para organizar los pasajes, aplicar esa fórmula de manera uniforme y transcribir mis decisiones a la hoja utilicé apoyo informático, incluidos scripts y asistencia de IA. La valoración de cada ítem y la responsabilidad sobre las decisiones registradas fueron mías. Las comillas reproducen el texto extraído del PDF, incluidos eventuales artefactos o truncamientos de extracción; no pretenden corregir ni completar el documento fuente.

2. En X-1 apliqué la definición de materialidad del manual §4 a la pérdida de contenido decisorio que la fuente declara en el pasaje excluido. Por tanto, MATERIAL no implica por sí solo que la exclusión fuera indebida respecto del dominio declarado del estudio, ni constituye una valoración final de la adecuación de su justificación. Esa cuestión se examinó separadamente en RA-1. No deseo añadir aclaraciones específicas sobre ítems concretos ni modificar ninguna respuesta de la hoja sellada.

##### Translation (authors):

1. I used a standard formula for the findings: "Excluding this passage omits decision-relevant source content concerning [section]: '[source passage]'". To organise the passages, apply that formula uniformly and transcribe my decisions to the sheet, I used computing support, including scripts and AI assistance. The assessment of each item and responsibility for the recorded decisions were mine. The quotation marks reproduce the text extracted from the PDF, including any extraction artefacts or truncations; they do not aim to correct or complete the source document.

2. In X-1 I applied the materiality definition of the manual (§4) to the loss of decision content that the source states in the excluded passage. Therefore, MATERIAL does not by itself imply that the exclusion was improper with respect to the study's declared domain, nor is it a final assessment of the adequacy of its rationale. That question was examined separately in RA-1. I do not wish to add item-specific clarifications or to modify any answer in the sealed sheet.

The authors' disposition reasons for exclusion findings used one standard sentence, which cites the recorded exclusion rationale (examples in Table S3-9).

#### S1.7 Traces of the admitted executable branches and of one non-instantiated recommendation

Each trace is transcribed from the frozen records: the canonical semantic record (R6′, SHA-256 1f9b67998212fecebbcbb5aefd24ea4d2baa335029749ee8849f8bb1860aa72b) and the frozen representations RV-NICE-V1 and RV-AAO-V1 (FP-C1, entries 269–271). Both representations record their authoring provenance as human-authored by Encoder 1. Missing-data policy for every comparison: a required variable that is not established yields *unresolved*, with no coercion (PG-07 M18). Judgement-bearing conditions are supplied by the case as literal atomic values; the study does not compute them from risk factors or calculators.

| Step | NICE-CD-0032-C01 (NICE NG81 recommendation 1.4.4) | AAO-CD-0009-C06 | AAO-CD-0036-C02 |
|---|---|---|---|
| Source text (segmented unit, L2) | "Offer 360° selective laser trabeculoplasty (SLT) to people with newly diagnosed OHT with IOP of 24 mmHg or more (excluding cases associated with pigment dispersion syndrome) if they are at risk of visual impairment within their lifetime (see the recommendation on taking account of risk factors in the section on diagnosis)." | "Treatment should be selectively directed toward patients with ocular hypertension at high risk of developing POAG." | "Lower IOP through treatment if an eye is likely to progress to POAG" |
| Semantic fields used (L3, as stated) | modality "Offer"; intervention "360° SLT"; conditions "newly diagnosed", "if they are at risk of visual impairment within their lifetime"; quantitative parameter "24 mmHg or more"; exception "excluding cases associated with pigment dispersion syndrome" | modality "should"; intervention "Treatment"; qualifier "selectively"; population "patients with ocular hypertension at high risk of developing POAG" | modality "Lower"; intervention "IOP", "treatment"; condition "if an eye is likely to progress to POAG"; qualifier "likely" |

| Step | NICE-CD-0032-C01 (NICE NG81 recommendation 1.4.4) | AAO-CD-0009-C06 | AAO-CD-0036-C02 |
|---|---|---|---|
| Admissibility class and choice rule | Source-constrained, study-level predeclared; "at risk of visual impairment within their lifetime" supplied by the case as a literal atomic condition (RR-OHT-01 (1), as recorded in the case-variable convention of RV-NICE-V1) | Source-constrained, study-level predeclared; admitted under RR-AAO-01 (4-bis), without a recommendation-strength label; "at high risk of developing POAG" supplied by the case as a literal atomic condition (RR-AAO-01 (1), as recorded in the case-variable convention of RV-AAO-V1) | Source-constrained, study-level predeclared; admitted under RR-AAO-01 (4-bis), without a recommendation-strength label; "likely to progress to POAG" supplied by the case for the eye under assessment as a literal atomic condition (RR-AAO-01 (1), as recorded in RV-AAO-V1) |
| Case variables | newly diagnosed OHT (binary); IOP (mmHg, numeric); associated pigment dispersion syndrome (binary); at risk of visual impairment within lifetime (binary, supplied) | ocular hypertension (binary, declared by the case); high risk of developing POAG (binary, supplied) | eye likely to progress to POAG (binary, supplied) |
| Executable condition | ALL OF: newly diagnosed OHT = true; IOP ≥ 24 mmHg; associated pigment dispersion syndrome = false; at risk of visual impairment within lifetime = true | ALL OF: ocular hypertension = true; high risk of developing POAG = true | eye likely to progress to POAG = true |
| Output when the condition holds | Resolved: initiate IOP-lowering treatment (direction only; the modality, 360° SLT, is not carried by the output vocabulary; see the overlap finding) | Resolved: initiate IOP-lowering treatment | Resolved: initiate IOP-lowering treatment |
| When the condition does not hold, or a variable is not established (branch level) | Unresolved (no branch states when to observe) | This branch does not resolve; the version-level output combines both AAO branches under the frozen evaluation semantics (ESD-OFTYGR-S6-01) | As for AAO-CD-0009-C06 |
| Case-space result (parents) | 8/96 resolved to initiate treatment; 88/96 unresolved (Table 3) | Representation RV-AAO-V1 (both branches): 72/96 resolved to initiate treatment; 24/96 unresolved (Table 3) | (as for RV-AAO-V1) |

**Case space.** The shared synthetic case set was the Cartesian product of the seven case variables, one set executed against both representations; each representation consults only its own variables (AAO three, NICE four) and ignores the others. IOP took 23, 24 and 25 mmHg around the NICE cut point of 24 mmHg; every other variable was binary. Ocular hypertension (AAO) was fixed at true, because only that side is in the study population (Table S3-10). Adults without established glaucomatous damage and without previous IOP-lowering treatment were project constants, not variables. This gives 96 parent cases (2 × 1 × 2 × 2 × 3 × 2 × 2); each parent has seven variants in which one variable is not established (672).

**Key to the choice rules and evaluation semantics.** *Literal risk predicates* (RR-OHT-01 and RR-AAO-01, points 1–2): a qualitative risk qualifier such as "at risk of visual impairment within their lifetime" or "at high risk of developing POAG" is carried as one atomic condition with its literal text, without numeric thresholds absent from the source, and its value is supplied by the case; a combination of factors that the source does not specify is marked undetermined rather than resolved. *Direction and default* (RR-OHT-01 points 3–4; RR-AAO-01 point 4-bis): a strong directive (NICE "offer"; an unhedged AAO "should" that directly governs starting treatment) makes the recommended action the default, with observation as a subordinate alternative; hedged wording ("consider", "may") gives co-equal alternatives and no default. Translating strength into this default is the study's own criterion, not the guideline developers'. Negative forms are excluded from these rules (point 5). *Output* *mapping*: when a branch's condition is true, the output is *resolved: initiate IOP-lowering treatment*; the output carries direction only. *Evaluation semantics* (ESD-OFTYGR-S6-01): all branches of a version are evaluated exhaustively under three-valued (Kleene) logic; a variable not established makes a condition unknown and is never coerced to true or false; a case resolves to *initiate* when at least one branch's condition is true and is otherwise *unresolved*, because no admitted branch states when to observe.

**Non-instantiation trace: NICE 1.4.3 (NICE-CD-0031-C01).** Transcribed from the recorded gate decision; no justification has been added.

| **Step** | **NICE-CD-0031-C01 (NICE NG81 recommendation 1.4.3, first sentence)** |
|---|---|
| Source text (segmented unit, L2) | "Do not offer treatment to people with OHT who are not at risk of visual impairment within their lifetime." |
| Extraction disposition | Retained, within the declared scope (it states when not to start treatment in people with OHT) |
| Encoder 1 classification before the gate | Source-derived, admissible primary, under the NICE prohibition rule RR-OHT-03 (prohibition represented as a constraint, treatment = no, under the stated condition) |
| Gate decision | Not instantiated; decision sheet N-1, PI as Encoder 1, 22 October 2026, under the rule "what is not demonstrated remains non-instantiated or pending" |
| Recorded reasons | (1) Under RR-OHT-03 the prohibition exists only as an exception on a directional treatment branch, and no admissible treatment branch existed to attach it to. (2) The rule that carries the lifetime-risk qualifier literally (RR-OHT-01 (1)–(2), used for recommendation 1.4.4) excludes negative forms (point 5), and no registered rule defined how the qualifier of a negative form is carried; defining it would have required an author choice (author-defined, not executable) |
| Record state | Incomplete; not terminal; not pending governance; classification and workbook unchanged |
| Companion unit | NICE-CD-0031-C02 ("Advise people to continue regular visits…"), not instantiated by the same decision: follow-up advice gives no direction |
| Consequence | No NICE branch states when to observe; the NICE representation cannot resolve to *observe without treatment* (Table 3) |

#### S1.8 Inter-encoder discrepancies and their resolution

Counts are transcribed from the committed discrepancy register (SHA-256 c5dbbe62add332b458e93ce6fb3c710669c43447d1c0ac53466b7d39322a056b, entry 71), the reconciliation outcome record (f20b1bcfea6e9982b392a1dd728ab922f6e014440feb045787edb6e41fd5bbf0, entry 79) and the residual register (18f0cfc1a38e196343ef90cea91d90abb9bfa13e7ecafde110caaddfba34ae6a, entry 81). A discrepancy is one difference between the two encoders' sealed initial records of a duplicated unit or of a dependency object linked to it, detected by the locked comparison tool.

| | **EGS** | **AAO** | **NICE** |
|---|---|---|---|
| Units and dependency objects with at least one discrepancy | 139 | 87 | 90 |
| Discrepancies, total | 1,441 | 437 | 556 |
| Structured-field state (L3 slot present or absent) | 780 | 172 | 271 |
| Dependency relation (Class B link) | 450 | 90 | 120 |
| Structured-field value (L3) | 182 | 136 | 142 |
| Authority dimensions (PG-06 outcome) | 29 | 29 | 22 |
| Segmented text (L2), exact | 0 | 5 | 0 |
| Defect flag | 0 | 5 | 1 |
| Resolved by joint agreement on Encoder 2's value | 1,232 | 269 | 405 |

| | EGS | AAO | NICE |
|---|---|---|---|
| Resolved by joint agreement on Encoder 1's value | 199 | 165 | 147 |
| Residual, unresolved (non-use rule) | 10 | 3 | 4 |

The 17 residual disagreements concern structured-field state (12), structured-field value (3) and authority dimensions (2). Each guideline is reported separately; no pooled figure is given.

#### Supplement S2 — Statements and limitations required by the registered protocol

The registered protocol requires the following statements and limitations to be reported with the study. Quoted text is reproduced from the frozen protocol records; other items are given in substance. They complement the limitations paragraph of the main text.

##### S2.1 What a fidelity statement can mean

• A fidelity determination is conditional on this corpus, this pipeline, this standard and this procedure. It is not a property of the EGS, AAO or NICE guidelines and does not transfer.

• No unconditional "faithful", "fidelity confirmed" or equivalent is made. No object-, item- or division-level fidelity claim is made from an absence of findings.

• Primary conclusions are limited to the components actually reviewed. They do not state or suggest that the source content of the corpus was shown to be faithfully transferred into the executable version.

• No value of the primary outcome defines study success or failure. There is no co-primary outcome and no corpus-level primary figure.

• No secondary or exploratory outcome qualifies, replaces, conditions or is reported as the primary result or as a fidelity determination. Exploratory outcomes support no confirmatory, comparative or causal claim.

##### S2.2 Review components and their limits

• **Materiality.** "A materiality answer is the reviewer's own judgment. It establishes nothing about whether the finding is correct and is not a verified defect. A source-side observation may carry any answer, including MATERIAL; counts of answers therefore cannot be presented as counts of verified encoding departures." A source-side MATERIAL answer counts in the lower bound of P-1.

• **Severity and categories.** There is no reviewer-rated severity; the materiality answer is a categorical judgement against a frozen definition, not a severity rating. A category is the reviewer's own label for a finding and establishes nothing about whether the finding is correct. Category counts are descriptive, not fidelity rates.

• **Omission detection (component c).** "An O-2 result concerns whether decision content that the source states in an enumerated division has a counterpart in the division record set … It says nothing about whether content retained in that record set is present in, or was justifiably left out of, any executable representation version." Component (c) covers enumerated divisions only.

• **Structural check.** "SEC is a single human structural pass. It makes detection of partition misfit a tested procedure; it does not guarantee detection." NO-OBSERVATION-RECORDED does not establish partition fit.

• **Materiality task.** The materiality task adds workload for each initial-phase finding and requires clinical judgement, which may yield CANNOT-JUDGE. Supplementary-phase findings carry no materiality answer, so the two phases have different record structures. (The supplementary phase closed with zero entries.)

• **Review counts.** Outside the overlap, each item received one prespecified review judgement; a post-hoc second reading covered a sample of exclusions (Table S3-12). Within the overlap it received two. Within the prespecified review, reviewer-dependence is observable only on the retained-object overlap, and the overlap is descriptive only.

• **Scope of clinical review.** Clinical review establishes nothing about the correctness of the software implementation.

• **Coverage ledger.** "The coverage ledger establishes that every structural division of the locked corpus was reviewed and accounted for in the coverage/disposition workflow; it establishes nothing about whether the correct units were drawn, whether meaning was preserved, or whether the encoding is faithful."

• **Disposition ledger.** "The ledger establishes that every disposition-bearing object produced by the extraction workflow received an explicit, attributable and traceable ledger state and, by ledger closure, a terminal disposition; it establishes nothing about whether those dispositions were correct, whether the retained material preserves the meaning of the source, or whether the encoding is faithful."

• **Structural and authority invariants.** "The PG-05 invariants establish only that every terminally retained object has a structurally complete, provenance-intact representation containing no prohibited content. They establish nothing about whether any representation says what its source says, and nothing about fidelity."

Likewise, "The PG-06 invariants establish only that every retained PG-05 object was assessed on both authority dimensions and that every positive assessment is supported by explicitly cited licensing source evidence. They establish nothing about whether the encoder interpreted those licensing expressions correctly, and nothing about fidelity."

• **Original exclusion decision.** "The original PG-04 disposition is a prior governance act. It is NOT a second independent PG-16 judgment and must not be described as one."

##### S2.3 No back-translation

"No back-translation was performed. The executable layer was read clinically only by source-first independent reviewers … and design-side by the second encoder (E-10). RC-1 to RC-6 are mechanical checks and not clinical readings. No source-unprimed reading of the executable was performed. Therefore no check tested whether the executable, read without the source, states what the source states. An ambiguous or under-specified executable element may have been read in the light of the source span just read."

The residual risk is a charitable reading of ambiguous condition logic, precedence, missing-data handling or output mapping. The size of what is forgone is unknown. The rationale for omitting back-translation is feasibility and the composition of the study team; it is not a finding that back-translation would add nothing.

##### S2.4 Disposition and acceptance

• The disposers are the encoders whose work was reviewed; Encoder 2 also verified it. "Their reasons, evidence and disagreements are visible and auditable; they are not independent."

• "DSP-NO-CHANGE is the disposers' judgment, not a refutation of the finding." A PASSed version can include content on which a finding was not adopted by the encoders. The finding and its reasons remain on record.

• "Under RD-1 nothing reconciles the two reviewers. Divergence is shown, not explained."

• Completing stage-7 disposition is necessary, not sufficient, for freeze. The procedural audit checks procedure, not clinical correctness.

• "PASS states that the frozen acceptance conditions were met for the named unit under the frozen procedure. It is not a fidelity determination beyond the PG-16 §7 limits, and it says nothing about unreviewed material or software correctness." PASS is procedural and has no numeric threshold.

• **Overlap commitments (signed by both authors, 30 October 2026).**

1. The material omission of SLT, the divergence between the two reviewers and the disposition reasons are reported.
2. The overlap finding is descriptive and outside the primary outcome; the absence of primary findings on NICE does not support a claim of no observed loss.
3. PASS is a governance result, never evidence of semantic fidelity.
4. The two-value output vocabulary is reported as a design limitation that independent review exposed.

##### S2.5 Encoding, agreement and interpretation

• **What duplicate encoding measures.** The design measures "semantic / interpretive reproducibility within the prospectively duplicated sample". It does not measure "extraction reproducibility; PG-03 unitisation reproducibility; PG-04 disposition reproducibility; whole-corpus semantic reproducibility; source fidelity; clinical validity; outcome validity." Inter-encoder agreement is not source fidelity, and duplicate semantic encoding is not independent clinical fidelity review.

• **Non-duplicated units.** "Units flagged NOT_DUPLICATED were semantically encoded by one encoder only. They were not independently double-encoded and did not receive inter-encoder reconciliation. Reported inter-encoder agreement describes the prospectively duplicated sample; for the pre-specified unconditional estimands it is accompanied by design-based sampling uncertainty per guideline body. It does not establish corpus-wide semantic reproducibility and establishes nothing about source fidelity."

• **Shared exposure.** "Both encoders have historical v0.1 exposure. They may share a misreading, so agreement may overstate reproducibility relative to unexposed encoders."

- **Single-pass extraction.** Extraction, unitisation and exclusion decisions were made once. Under-segmentation or false exclusion upstream is single-pass and is not corrected by duplication.
- **Precision and descriptive estimands.** The ±d precision applies only to the four unconditional estimands. Class-B agreement is descriptive, within duplicated dependency objects only. κ and AC1 are reported together, with no benchmark; they can disagree sharply in skewed tables.
- **Prior knowledge.** The principal encoder knows the historical v0.1 study; prior knowledge cannot be erased, no naïveté is claimed, and v0.1 material was prohibited as prompts. "Before source encoding" means before v0.2 extraction, not before any exposure.
- **Contamination risks, disclosed in both directions:**
  - over-identifying source-specified content, and the opposite risk of over-identifying authority allocation or delegation;
  - threshold text may look more determinate than it is;
  - mentions of clinicians or patients may attract over-allocation of authority;
  - "consider" or "may" may attract erroneous authority readings.

Symmetric controls cannot eliminate these risks. Structural completeness can coexist with systematic semantic misclassification.

- **Competing readings.** Historical knowledge could encourage competing readings that would not otherwise have been proposed and direct where they were sought, while reaction against the earlier study could favour forced uniqueness or overlook another reading. Remembered thresholds could influence which source-supported scope reading appeared natural without entering the record as numbers. The controls cannot distinguish a source-supported reading discovered through remembered logic from one discovered without that influence.

- **Non-identification and case variables.** Possible bias toward inferred absence, and the opposite pressure to overuse NOT_ESTABLISHED. Numeric variables may look easier, and broad non-identification claims may be overclaimed. "TARGET DRIFT is a material risk"; target definitions were frozen before assessment. ESTABLISHED does not by itself certify evidence sufficiency.
- **Interpretation closure.** "A READING NEVER IDENTIFIED CANNOT BE RECORDED." Procedural closure does not establish exhaustive natural-language interpretation; the study claims only that all candidate readings identified under the frozen procedure were recorded and dispositioned.
- **Non-inheritance.** The v0.1 taxonomy was not inherited; the three-state primary execution taxonomy was prospectively re-derived.
- **Non-reuse.** RU-0 prohibited screening or reusing v0/v0.1 components; no reuse is recorded in the study records, and final adherence was not independently verified against the complete implementation, change and defect records (S1.3). Non-reuse of code does not remove v0.1 knowledge.

##### S2.6 Case space and execution

- "The case space is synthetic. Output counts over it describe the grid, not patients." No frequency over it is a clinical prevalence.
- The case space is shaped by what was encoded. It cannot expose an omitted variable or cut point.
- One-at-a-time missing-value variants do not exercise joint missingness. Population-defining conditions are not exercised outside scope, and excluded boundary sides are not tested (Table S3-10).
- Execution outputs are not clinical recommendations and do not carry the primary fidelity claim.
- Determinacy is not fidelity and not clinical correctness.

##### S2.7 Implementation timing and integrity

- Implementation material was drafted after v0.2 source contact and locked no later than first use. "No statement may say that all implementation material was frozen before source contact; only the P26-04 set was."

- "A freeze fixes what is used; it does not make locked material correct." Custody and timestamps are not independence mechanisms.
- The registration fixes when, not how, the design was arrived at, and does not make it naïve.
- No cross-guideline divergence result is reported. The study says nothing about whether the three bodies' encodings behave alike on the same cases, and no body is ranked by fidelity.

##### S2.8 Conflict of roles

The PI holds final decision authority and is also an author, the principal encoder and the author of the executable encoding. The protocol's controls limit self-certification but do not remove this conflict.

### Supplement S3 — Full results tables

#### Table S3-1. Primary outcome with bounds, classes and flags (PRIMARY_BY_BODY.csv, CLASS_X_J.csv, SUPPLEMENTAL_DE.csv, S_INDICATOR.csv)

Counts of units with reviewer-judged answers, not verified encoding departures; findings may concern the source rather than the encoding (P22-27). NO-FINDING-RAISED is never 'fidelity confirmed' (P18-17, P22-23). Per body only; no cross-body total (P22-21, P24-10).

| body | component | status | denominator | P1_lower | P1_upper | P2_lower | P2_upper | QUERY_ONLY | J_units | R_units | K_units |
|---|---|---|---|---|---|---|---|---|---|---|---|
| EGS | (a) retained objects | not computable | 0 | NOT COMPUTABLE | NOT COMPUTABLE | NOT COMPUTABLE | NOT COMPUTABLE | NOT COMPUTABLE | NOT COMPUTABLE | NOT COMPUTABLE | NOT COMPUTABLE |
| EGS | (b) documented exclusions | computed | 2109 | 601 | 601 | 601 | 601 | 0 | 0 | 0 | 0 |
| EGS | (c) enumerated divisions | computed | 262 | 1 | 1 | 1 | 1 | 0 | 0 | 0 | 0 |
| EGS | (d) SEC pass | computed | 1 | 0 | 0 | 0 | 0 | 0 | 0 | 0 | 0 |
| EGS | (e) version-level items | not applicable | 0 | NOT APPLICABLE | NOT APPLICABLE | NOT APPLICABLE | NOT APPLICABLE | NOT APPLICABLE | NOT APPLICABLE | NOT APPLICABLE | NOT APPLICABLE |
| AAO | (a) retained objects | computed | 2 | 0 | 0 | 0 | 0 | 0 | 0 | 0 | 0 |
| AAO | (b) documented exclusions | computed | 234 | 216 | 216 | 216 | 216 | 0 | 0 | 0 | 0 |
| AAO | (c) enumerated divisions | computed | 58 | 0 | 0 | 0 | 0 | 0 | 0 | 0 | 0 |
| AAO | (d) SEC pass | computed | 1 | 0 | 0 | 0 | 0 | 0 | 0 | 0 | 0 |
| AAO | (e) version-level items | not applicable | 0 | NOT APPLICABLE | NOT APPLICABLE | NOT APPLICABLE | NOT APPLICABLE | NOT APPLICABLE | NOT APPLICABLE | NOT APPLICABLE | NOT APPLICABLE |
| NICE | (a) retained objects | computed | 1 | 0 | 0 | 0 | 0 | 0 | 0 | 0 | 0 |
| NICE | (b) documented exclusions | computed | 179 | 151 | 151 | 151 | 151 | 0 | 0 | 0 | 0 |
| NICE | (c) enumerated divisions | computed | 124 | 0 | 0 | 0 | 0 | 0 | 0 | 0 | 0 |
| NICE | (d) SEC pass | computed | 1 | 0 | 0 | 1 | 1 | 0 | 0 | 0 | 0 |
| NICE | (e) version-level items | not applicable | 0 | NOT APPLICABLE | NOT APPLICABLE | NOT APPLICABLE | NOT APPLICABLE | NOT APPLICABLE | NOT APPLICABLE | NOT APPLICABLE | NOT APPLICABLE |

Component (c) compares source-stated decision content with each enumerated division's records, not with an executable version; it covers enumerated divisions only. The structural indicator S (below) is reported with it and lies outside every bound.

Classes by component and J flag:

| body | component | p1_class | J | units |
|---|---|---|---|---|
| AAO | (a) retained objects | NO-FINDING | 0 | 2 |
| AAO | (b) documented exclusions | MATERIAL | 0 | 216 |
| AAO | (b) documented exclusions | NO-FINDING | 0 | 18 |
| AAO | (c) enumerated divisions | NO-FINDING | 0 | 58 |
| AAO | (d) SEC pass | NO-FINDING | 0 | 1 |
| EGS | (b) documented exclusions | MATERIAL | 0 | 601 |

| body | component | p1_class | J | units |
|---|---|---|---|---|
| EGS | (b) documented exclusions | NO-FINDING | 0 | 1508 |
| EGS | (c) enumerated divisions | MATERIAL | 0 | 1 |
| EGS | (c) enumerated divisions | NO-FINDING | 0 | 261 |
| EGS | (d) SEC pass | NO-FINDING | 0 | 1 |
| NICE | (a) retained objects | NO-FINDING | 0 | 1 |
| NICE | (b) documented exclusions | MATERIAL | 0 | 151 |
| NICE | (b) documented exclusions | NO-FINDING | 0 | 28 |
| NICE | (c) enumerated divisions | NO-FINDING | 0 | 124 |
| NICE | (d) SEC pass | NOT-MATERIAL-ONLY | 0 | 1 |

Supplemental counts for components (d) and (e):

| body | component | status | findings | stance_ASSERTED | stance_QUERIED | stance_MIXED | stance_NOT-MARKED | stance_blank | answer_MATERIAL | answer_NOT-MATERIAL | answer_CANNOT-JUDGE | answer_blank |
|---|---|---|---|---|---|---|---|---|---|---|---|---|
| EGS | (d) SEC pass | computed | 0 | 0 | 0 | 0 | 0 | 0 | 0 | 0 | 0 | 0 |
| EGS | (e) version-level items | not applicable | NOT APPLICABLE | NOT APPLICABLE | NOT APPLICABLE | NOT APPLICABLE | NOT APPLICABLE | NOT APPLICABLE | NOT APPLICABLE | NOT APPLICABLE | NOT APPLICABLE | NOT APPLICABLE |
| AAO | (d) SEC pass | computed | 0 | 0 | 0 | 0 | 0 | 0 | 0 | 0 | 0 | 0 |
| AAO | (e) version-level items | not applicable | NOT APPLICABLE | NOT APPLICABLE | NOT APPLICABLE | NOT APPLICABLE | NOT APPLICABLE | NOT APPLICABLE | NOT APPLICABLE | NOT APPLICABLE | NOT APPLICABLE | NOT APPLICABLE |
| NICE | (d) SEC pass | computed | 1 | 1 | 0 | 0 | 0 | 0 | 0 | 1 | 0 | 0 |
| NICE | (e) version-level items | not applicable | NOT APPLICABLE | NOT APPLICABLE | NOT APPLICABLE | NOT APPLICABLE | NOT APPLICABLE | NOT APPLICABLE | NOT APPLICABLE | NOT APPLICABLE | NOT APPLICABLE | NOT APPLICABLE |

Structural indicator S, outside every bound:

| body | S | unenumerated_observations | handling_status_summary |
|---|---|---|---|
| EGS | NO-OBSERVATION-RECORDED | 0 | none |
| AAO | NO-OBSERVATION-RECORDED | 0 | none |
| NICE | OBSERVATION-RECORDED | 1 | DSP-RB-OBS-001: DSP-CHANGE (route: PG-03 §19 (routed; P19-01, P19-03); transcribed: 22_DA2_TRASCRIZIONE_P19-08_PAGINA34_FIRMATO.md): 1 |

#### Table S3-2. Secondary and exploratory review outcomes C-01 to C-07 (C01_C07_REVIEW.csv)

Counts of units with reviewer-judged answers, not verified encoding departures; findings may concern the source rather than the encoding (P22-27). NO-FINDING-RAISED is never 'fidelity confirmed' (P18-17, P22-23). Per body only; no cross-body total (P22-21, P24-10). Exploratory outcomes support no confirmatory, comparative or causal claim (P23-21).

| outcome | label | body | scope | category_or_item | value | note |
|---|---|---|---|---|---|---|
| C-01 | SECONDARY | EGS | (a) retained objects | (no finding) | 0 | count; no denominator (P23-06) |
| C-01 | SECONDARY | AAO | (a) retained objects | (no finding) | 0 | count; no denominator (P23-06) |

| outcome | label | body | scope | category_or_item | value | note |
|---|---|---|---|---|---|---|
| C-01 | SECONDARY | NICE | (a) retained objects | (no finding) | 0 | count; no denominator (P23-06) |
| C-01 | SECONDARY | EGS | (b) documented exclusions | RF-EXCLUSION-QUESTIONED | 601 | count; no denominator (P23-06) |
| C-01 | SECONDARY | AAO | (b) documented exclusions | RF-EXCLUSION-QUESTIONED | 216 | count; no denominator (P23-06) |
| C-01 | SECONDARY | NICE | (b) documented exclusions | RF-EXCLUSION-QUESTIONED | 151 | count; no denominator (P23-06) |
| C-01 | SECONDARY | EGS | (c) enumerated divisions | RF-POSSIBLE-OMISSION | 1 | count; no denominator (P23-06) |
| C-01 | SECONDARY | AAO | (c) enumerated divisions | (no finding) | 0 | count; no denominator (P23-06) |
| C-01 | SECONDARY | NICE | (c) enumerated divisions | (no finding) | 0 | count; no denominator (P23-06) |
| C-01 | SECONDARY | EGS | (d) SEC pass | (no finding) | 0 | count; no denominator (P23-06) |
| C-01 | SECONDARY | AAO | (d) SEC pass | (no finding) | 0 | count; no denominator (P23-06) |
| C-01 | SECONDARY | NICE | (d) SEC pass | RF-NOT-CLASSIFIED | 1 | count; no denominator (P23-06) |
| C-02 | SECONDARY | EGS | R-I findings on TV-1 | DSP-CHANGE | 0/602 (0.0%) | denominator: all R-I findings on TV-1 (P23-07) |
| C-02 | SECONDARY | EGS | R-I findings on TV-1 | DSP-NO-CHANGE | 602/602 (100.0%) | denominator: all R-I findings on TV-1 (P23-07) |
| C-02 | SECONDARY | EGS | R-I findings on TV-1 | DSP-UNRESOLVED | 0/602 (0.0%) | denominator: all R-I findings on TV-1 (P23-07) |
| C-02 | SECONDARY | EGS | R-I findings on TV-1 | OPEN | 0/602 (0.0%) | denominator: all R-I findings on TV-1 (P23-07) |
| C-02 | SECONDARY | EGS | R-I findings on TV-1 | DSP-ROUTED | 0/602 (0.0%) | denominator: all R-I findings on TV-1 (P23-07) |
| C-02 | SECONDARY | EGS | R-I findings on TV-1 | RECORD-MISSING | 0/602 (0.0%) | denominator: all R-I findings on TV-1 (P23-07) |
| C-02 | SECONDARY | EGS | R-I findings on TV-1 | of which terminal via referral (P19-08) | 0 | supplemental |
| C-02 | SECONDARY | AAO | R-I findings on TV-1 | DSP-CHANGE | 0/216 (0.0%) | denominator: all R-I findings on TV-1 (P23-07) |
| C-02 | SECONDARY | AAO | R-I findings on TV-1 | DSP-NO-CHANGE | 216/216 (100.0%) | denominator: all R-I findings on TV-1 (P23-07) |
| C-02 | SECONDARY | AAO | R-I findings on TV-1 | DSP-UNRESOLVED | 0/216 (0.0%) | denominator: all R-I findings on TV-1 (P23-07) |
| C-02 | SECONDARY | AAO | R-I findings on TV-1 | OPEN | 0/216 (0.0%) | denominator: all R-I findings on TV-1 (P23-07) |
| C-02 | SECONDARY | AAO | R-I findings on TV-1 | DSP-ROUTED | 0/216 (0.0%) | denominator: all R-I findings on TV-1 (P23-07) |
| C-02 | SECONDARY | AAO | R-I findings on TV-1 | RECORD-MISSING | 0/216 (0.0%) | denominator: all R-I findings on TV-1 (P23-07) |
| C-02 | SECONDARY | AAO | R-I findings on TV-1 | of which terminal via referral (P19-08) | 0 | supplemental |
| C-02 | SECONDARY | NICE | R-I findings on TV-1 | DSP-CHANGE | 1/153 (0.7%) | denominator: all R-I findings on TV-1 (P23-07) |
| C-02 | SECONDARY | NICE | R-I findings on TV-1 | DSP-NO-CHANGE | 152/153 (99.3%) | denominator: all R-I findings on TV-1 (P23-07) |
| C-02 | SECONDARY | NICE | R-I findings on TV-1 | DSP-UNRESOLVED | 0/153 (0.0%) | denominator: all R-I findings on TV-1 (P23-07) |
| C-02 | SECONDARY | NICE | R-I findings on TV-1 | OPEN | 0/153 (0.0%) | denominator: all R-I findings on TV-1 (P23-07) |
| C-02 | SECONDARY | NICE | R-I findings on TV-1 | DSP-ROUTED | 0/153 (0.0%) | denominator: all R-I findings on TV-1 (P23-07) |
| C-02 | SECONDARY | NICE | R-I findings on TV-1 | RECORD-MISSING | 0/153 (0.0%) | denominator: all R-I findings on TV-1 (P23-07) |
| C-02 | SECONDARY | NICE | R-I findings on TV-1 | of which terminal via referral (P19-08) | 2 | supplemental |
| C-02 | SECONDARY | NICE | observation (reported separately) | DSP-RB-OBS-001 | DSP-CHANGE | not a finding; PG-03 §19 route |
| C-02 | SECONDARY | EGS | flags (reported separately) | count | 0 | |
| C-02 | SECONDARY | EGS | overlap divergences (reported separately) | count | 0 | |
| C-02 | SECONDARY | AAO | flags (reported separately) | count | 0 | |
| C-02 | SECONDARY | AAO | overlap divergences (reported separately) | count | 0 | |
| C-02 | SECONDARY | NICE | flags (reported separately) | count | 0 | |
| C-02 | SECONDARY | NICE | overlap divergences (reported separately) | count | 1 | |
| C-03 | SECONDARY | EGS | output-intended lineage versions | (none) | NOT APPLICABLE | no output-intended lineage |
| C-03 | SECONDARY | AAO | RL-AAO-PRIMARY (CANONICAL_PRIMARY) | RV-AAO-V1 | PASS | freeze entry: YES cfb188efe4962ccd2497d8ea1cc7de13368156f2e1e6749e6858c96b9ab63c90; PASS is procedural (P20-02) |
| C-03 | SECONDARY | NICE | RL-NICE-PRIMARY (CANONICAL_PRIMARY) | RV-NICE-V1 | PASS | freeze entry: YES a44c40974d8f6833c0fe19004af17a112ee7a973c36870d498bb692eceb165ec; PASS is procedural (P20-02) |
| C-04 | SECONDARY | EGS | final PASSed version | (none) | NOT APPLICABLE | no PASSed version |

| outcome | label | body | scope | category_or_item | value | note |
|---|---|---|---|---|---|---|
| C-04 | SECONDARY | AAO | final PASSed version | RV-AAO-V1 | = P-1/P-2 tables (outcome_compute.py) | version_seq 1: the final PASSed version is the first frozen review version; no successor version |
| C-04 | SECONDARY | NICE | final PASSed version | RV-NICE-V1 | = P-1/P-2 tables (outcome_compute.py) | version_seq 1: the final PASSed version is the first frozen review version; no successor version |
| C-05 | EXPLORATORY | EGS | admitted alternative lineages | 0 | NOT APPLICABLE | exploratory |
| C-05 | EXPLORATORY | AAO | admitted alternative lineages | 0 | NOT APPLICABLE | exploratory |
| C-05 | EXPLORATORY | NICE | admitted alternative lineages | 0 | NOT APPLICABLE | exploratory |
| C-06 | EXPLORATORY | EGS | R-II findings (count; no denominator) | all stances / categories | 0 | zero-entry final R-II seals (RII-ZERO-ENTRY-FINAL-SEAL-RA-V2, RII-ZERO-ENTRY-FINAL-SEAL-RB-V2) |
| C-06 | EXPLORATORY | AAO | R-II findings (count; no denominator) | all stances / categories | 0 | zero-entry final R-II seals (RII-ZERO-ENTRY-FINAL-SEAL-RA-V2, RII-ZERO-ENTRY-FINAL-SEAL-RB-V2) |
| C-06 | EXPLORATORY | NICE | R-II findings (count; no denominator) | all stances / categories | 0 | zero-entry final R-II seals (RII-ZERO-ENTRY-FINAL-SEAL-RA-V2, RII-ZERO-ENTRY-FINAL-SEAL-RB-V2) |
| C-07 | EXPLORATORY | EGS | documented exclusions with >=1 RA-1 finding | units | 0/2109 (0.0%) | denominator: documented exclusions in (b) |
| C-07 | EXPLORATORY | EGS | class (PG-22 precedence) | ASSERTED | 0/2109 (0.0%) | |
| C-07 | EXPLORATORY | EGS | class (PG-22 precedence) | STANCE-UNRESOLVED | 0/2109 (0.0%) | |
| C-07 | EXPLORATORY | EGS | class (PG-22 precedence) | QUERY-ONLY | 0/2109 (0.0%) | |
| C-07 | EXPLORATORY | EGS | J flag | units | 0 | |
| C-07 | EXPLORATORY | EGS | R flag | units | 0 | |
| C-07 | EXPLORATORY | AAO | documented exclusions with >=1 RA-1 finding | units | 0/234 (0.0%) | denominator: documented exclusions in (b) |
| C-07 | EXPLORATORY | AAO | class (PG-22 precedence) | ASSERTED | 0/234 (0.0%) | |
| C-07 | EXPLORATORY | AAO | class (PG-22 precedence) | STANCE-UNRESOLVED | 0/234 (0.0%) | |
| C-07 | EXPLORATORY | AAO | class (PG-22 precedence) | QUERY-ONLY | 0/234 (0.0%) | |
| C-07 | EXPLORATORY | AAO | J flag | units | 0 | |
| C-07 | EXPLORATORY | AAO | R flag | units | 0 | |

| outcome | label | body | scope | category_or_item | value | note |
|---|---|---|---|---|---|---|
| C-07 | EXPLORATORY | NICE | documented exclusions with >=1 RA-1 finding | units | 0/179 (0.0%) | denominator: documented exclusions in (b) |
| C-07 | EXPLORATORY | NICE | class (PG-22 precedence) | ASSERTED | 0/179 (0.0%) | |
| C-07 | EXPLORATORY | NICE | class (PG-22 precedence) | STANCE-UNRESOLVED | 0/179 (0.0%) | |
| C-07 | EXPLORATORY | NICE | class (PG-22 precedence) | QUERY-ONLY | 0/179 (0.0%) | |
| C-07 | EXPLORATORY | NICE | J flag | units | 0 | |
| C-07 | EXPLORATORY | NICE | R flag | units | 0 | |

#### Table S3-3. Execution determinacy C-08 and Case-B stratum C-09 (C08_C09_EXECUTION.csv)

Synthetic case grid (PG-21): counts describe the designed grid, carry no prevalence; determinacy is not fidelity and not clinical correctness (PG-08 §27; PG-23 P23-13); per body, no cross-body contrast.

| outcome | label | body | representation_version_id | pairing_ref | stratum | case_kind | state_or_reason | value |
|---|---|---|---|---|---|---|---|---|
| C-08 | SECONDARY | AAO | RV-AAO-V1 | PAIR-RV-AAO-V1-TSC-PG02-S8 | PRIMARY_FIDELITY_DENOMINATOR | PARENT | RESOLVED__INITIATE_IOP_LOWERING_TREATMENT | 72/96 (75.0%) |
| C-08 | SECONDARY | AAO | RV-AAO-V1 | PAIR-RV-AAO-V1-TSC-PG02-S8 | PRIMARY_FIDELITY_DENOMINATOR | PARENT | RESOLVED__OBSERVE_WITHOUT_TREATMENT | 0/96 (0.0%) |
| C-08 | SECONDARY | AAO | RV-AAO-V1 | PAIR-RV-AAO-V1-TSC-PG02-S8 | PRIMARY_FIDELITY_DENOMINATOR | PARENT | UNRESOLVED | 24/96 (25.0%) |
| C-08 | SECONDARY | AAO | RV-AAO-V1 | PAIR-RV-AAO-V1-TSC-PG02-S8 | PRIMARY_FIDELITY_DENOMINATOR | VARIANT | RESOLVED__INITIATE_IOP_LOWERING_TREATMENT | 432/672 (64.3%) |
| C-08 | SECONDARY | AAO | RV-AAO-V1 | PAIR-RV-AAO-V1-TSC-PG02-S8 | PRIMARY_FIDELITY_DENOMINATOR | VARIANT | RESOLVED__OBSERVE_WITHOUT_TREATMENT | 0/672 (0.0%) |
| C-08 | SECONDARY | AAO | RV-AAO-V1 | PAIR-RV-AAO-V1-TSC-PG02-S8 | PRIMARY_FIDELITY_DENOMINATOR | VARIANT | UNRESOLVED | 240/672 (35.7%) |
| C-09 | EXPLORATORY | AAO | RV-AAO-V1 | PAIR-RV-AAO-V1-TSC-PG02-S8 | SOURCE_NONIDENTIFICATION_APPLICATION_STRATUM | PARENT | RESOLVED__INITIATE_IOP_LOWERING_TREATMENT | NOT APPLICABLE |
| C-09 | EXPLORATORY | AAO | RV-AAO-V1 | PAIR-RV-AAO-V1-TSC-PG02-S8 | SOURCE_NONIDENTIFICATION_APPLICATION_STRATUM | PARENT | RESOLVED__OBSERVE_WITHOUT_TREATMENT | NOT APPLICABLE |
| C-09 | EXPLORATORY | AAO | RV-AAO-V1 | PAIR-RV-AAO-V1-TSC-PG02-S8 | SOURCE_NONIDENTIFICATION_APPLICATION_STRATUM | PARENT | UNRESOLVED | NOT APPLICABLE |
| C-09 | EXPLORATORY | AAO | RV-AAO-V1 | PAIR-RV-AAO-V1-TSC-PG02-S8 | SOURCE_NONIDENTIFICATION_APPLICATION_STRATUM | VARIANT | RESOLVED__INITIATE_IOP_LOWERING_TREATMENT | NOT APPLICABLE |
| C-09 | EXPLORATORY | AAO | RV-AAO-V1 | PAIR-RV-AAO-V1-TSC-PG02-S8 | SOURCE_NONIDENTIFICATION_APPLICATION_STRATUM | VARIANT | RESOLVED__OBSERVE_WITHOUT_TREATMENT | NOT APPLICABLE |
| C-09 | EXPLORATORY | AAO | RV-AAO-V1 | PAIR-RV-AAO-V1-TSC-PG02-S8 | SOURCE_NONIDENTIFICATION_APPLICATION_STRATUM | VARIANT | UNRESOLVED | NOT APPLICABLE |
| C-08 | SECONDARY | NICE | RV-NICE-V1 | PAIR-RV-NICE-V1-TSC-PG02-S8 | PRIMARY_FIDELITY_DENOMINATOR | PARENT | RESOLVED__INITIATE_IOP_LOWERING_TREATMENT | 8/96 (8.3%) |
| C-08 | SECONDARY | NICE | RV-NICE-V1 | PAIR-RV-NICE-V1-TSC-PG02-S8 | PRIMARY_FIDELITY_DENOMINATOR | PARENT | RESOLVED__OBSERVE_WITHOUT_TREATMENT | 0/96 (0.0%) |
| C-08 | SECONDARY | NICE | RV-NICE-V1 | PAIR-RV-NICE-V1-TSC-PG02-S8 | PRIMARY_FIDELITY_DENOMINATOR | PARENT | UNRESOLVED | 88/96 (91.7%) |

| outcome | label | body | representation_version_id | pairing_ref | stratum | case_kind | state_or_reason | value |
|---|---|---|---|---|---|---|---|---|
| C-08 | SECONDARY | NICE | RV-NICE-V1 | PAIR-RV-NICE-V1-TSC-PG02-S8 | PRIMARY_FIDELITY_DENOMINATOR | VARIANT | RESOLVED__INITIATE_IOP_LOWERING_TREATMENT | 24/672 (3.6%) |
| C-08 | SECONDARY | NICE | RV-NICE-V1 | PAIR-RV-NICE-V1-TSC-PG02-S8 | PRIMARY_FIDELITY_DENOMINATOR | VARIANT | RESOLVED__OBSERVE_WITHOUT_TREATMENT | 0/672 (0.0%) |
| C-08 | SECONDARY | NICE | RV-NICE-V1 | PAIR-RV-NICE-V1-TSC-PG02-S8 | PRIMARY_FIDELITY_DENOMINATOR | VARIANT | UNRESOLVED | 648/672 (96.4%) |
| C-09 | EXPLORATORY | NICE | RV-NICE-V1 | PAIR-RV-NICE-V1-TSC-PG02-S8 | SOURCE_NONIDENTIFICATION_APPLICATION_STRATUM | PARENT | RESOLVED__INITIATE_IOP_LOWERING_TREATMENT | NOT APPLICABLE |
| C-09 | EXPLORATORY | NICE | RV-NICE-V1 | PAIR-RV-NICE-V1-TSC-PG02-S8 | SOURCE_NONIDENTIFICATION_APPLICATION_STRATUM | PARENT | RESOLVED__OBSERVE_WITHOUT_TREATMENT | NOT APPLICABLE |
| C-09 | EXPLORATORY | NICE | RV-NICE-V1 | PAIR-RV-NICE-V1-TSC-PG02-S8 | SOURCE_NONIDENTIFICATION_APPLICATION_STRATUM | PARENT | UNRESOLVED | NOT APPLICABLE |
| C-09 | EXPLORATORY | NICE | RV-NICE-V1 | PAIR-RV-NICE-V1-TSC-PG02-S8 | SOURCE_NONIDENTIFICATION_APPLICATION_STRATUM | VARIANT | RESOLVED__INITIATE_IOP_LOWERING_TREATMENT | NOT APPLICABLE |
| C-09 | EXPLORATORY | NICE | RV-NICE-V1 | PAIR-RV-NICE-V1-TSC-PG02-S8 | SOURCE_NONIDENTIFICATION_APPLICATION_STRATUM | VARIANT | RESOLVED__OBSERVE_WITHOUT_TREATMENT | NOT APPLICABLE |
| C-09 | EXPLORATORY | NICE | RV-NICE-V1 | PAIR-RV-NICE-V1-TSC-PG02-S8 | SOURCE_NONIDENTIFICATION_APPLICATION_STRATUM | VARIANT | UNRESOLVED | NOT APPLICABLE |
| PES-VALIDATION | SECONDARY (C-08 companion) | AAO | RV-AAO-V1 | PAIR-RV-AAO-V1-TSC-PG02-S8 | | PARENT | VALIDATION FAILURE | 0/96 (0.0%) |
| PES-VALIDATION | SECONDARY (C-08 companion) | AAO | RV-AAO-V1 | PAIR-RV-AAO-V1-TSC-PG02-S8 | | VARIANT | VALIDATION FAILURE | 0/672 (0.0%) |
| PES-VALIDATION | SECONDARY (C-08 companion) | NICE | RV-NICE-V1 | PAIR-RV-NICE-V1-TSC-PG02-S8 | | PARENT | VALIDATION FAILURE | 0/96 (0.0%) |
| PES-VALIDATION | SECONDARY (C-08 companion) | NICE | RV-NICE-V1 | PAIR-RV-NICE-V1-TSC-PG02-S8 | | VARIANT | VALIDATION FAILURE | 0/672 (0.0%) |

#### Table S3-4. Inter-encoder agreement C-10 — semantic slot states, by guideline (C10_REPRODUCIBILITY.csv)

INTER-ENCODER AGREEMENT ≠ SOURCE FIDELITY. These descriptors measure the reproducibility of two independent initial semantic records produced on a shared canonical unitisation by two encoders who share historical v0.1 exposure, within the prospectively duplicated sample. They measure neither extraction nor disposition reproducibility, and establish nothing about whether any record is faithful to the source. Per body; no pooled reproducibility score; no acceptance threshold (PG-15 sec. 11.2; PG-20).

**EGS** — duplicated sample 83 of frame 581 units (n_h / N_h); intervals are exact design-based hypergeometric 95% intervals (HI-1).

| Slot | Raw agreement | HI-1 95% interval | Contingency V/V, V/N, N/V, N/N | Category-specific agreement (VALUE) | Category-specific agreement (NO_EXPLICIT_EXPRESSION_IDENTIFIED_IN_UNIT) | Cohen κ | Gwet AC1 |
|---|---|---|---|---|---|---|---|
| subject_population_as_stated | 79/83 (95.2%) | [88.6%, 98.5%] | 10 / 4 / 0 / 69 | 20/24 (83.3%) | 138/142 (97.2%) | 0.81 | 0.94 |
| context_as_stated | 75/83 (90.4%) | [82.6%, 95.4%] | 9 / 7 / 1 / 66 | 18/26 (69.2%) | 132/140 (94.3%) | 0.64 | 0.87 |
| condition_expression_as_stated | 73/83 (88.0%) | [79.7%, 93.6%] | 9 / 10 / 0 / 64 | 18/28 (64.3%) | 128/138 (92.8%) | 0.58 | 0.83 |
| proposition_content_as_stated | 81/83 (97.6%) | [92.1%, 99.5%] | 81 / 0 / 2 / 0 | 162/164 (98.8%) | 0/2 (0.0%) | 0.00 | 0.98 |
| object_intervention_as_stated | 48/83 (57.8%) | [47.3%, 67.8%] | 35 / 32 / 3 / 13 | 70/105 (66.7%) | 26/61 (42.6%) | 0.20 | 0.21 |

| Slot | Raw agreement | HI-1 95% interval | Contingency V/V, V/N, N/V, N/N | Category-specific agreement (VALUE) | Category-specific agreement (NO_EXPLICIT_EXPRESSION_IDENTIFIED_IN_UNIT) | Cohen κ | Gwet AC1 |
|---|---|---|---|---|---|---|---|
| alternative_as_stated | 76/83 (91.6%) | [84.0%, 96.2%] | 2 / 0 / 7 / 74 | 4/11 (36.4%) | 148/155 (95.5%) | 0.34 | 0.90 |
| modality_as_stated | 81/83 (97.6%) | [92.1%, 99.5%] | 26 / 2 / 0 / 55 | 52/54 (96.3%) | 110/112 (98.2%) | 0.95 | 0.96 |
| negation_expression_as_stated | 81/83 (97.6%) | [92.1%, 99.5%] | 11 / 1 / 1 / 70 | 22/24 (91.7%) | 140/142 (98.6%) | 0.90 | 0.97 |
| qualifier_as_stated | 66/83 (79.5%) | [70.1%, 86.9%] | 24 / 9 / 8 / 42 | 48/65 (73.8%) | 84/101 (83.2%) | 0.57 | 0.61 |
| exception_expression_as_stated | 82/83 (98.8%) | [94.0%, 99.8%] | 0 / 0 / 1 / 82 | 0/1 (0.0%) | 164/165 (99.4%) | 0.00 | 0.99 |
| quantitative_parameter_as_stated | 82/83 (98.8%) | [94.0%, 99.8%] | 17 / 1 / 0 / 65 | 34/35 (97.1%) | 130/131 (99.2%) | 0.96 | 0.98 |
| connective_as_stated | 79/83 (95.2%) | [88.6%, 98.5%] | 39 / 3 / 1 / 40 | 78/82 (95.1%) | 80/84 (95.2%) | 0.90 | 0.90 |
| cross_reference_as_stated | 83/83 (100.0%) | [96.0%, 100.0%] | 5 / 0 / 0 / 78 | 10/10 (100.0%) | 156/156 (100.0%) | 1.00 | 1.00 |
| actor_expression_as_stated | 81/83 (97.6%) | [92.1%, 99.5%] | 8 / 0 / 2 / 73 | 16/18 (88.9%) | 146/148 (98.6%) | 0.88 | 0.97 |

**AAO** — duplicated sample 74 of frame 302 units (n_h / N_h); intervals are exact design-based hypergeometric 95% intervals (HI-1).

| Slot | Raw agreement | HI-1 95% interval | Contingency V/V, V/N, N/V, N/N | Category-specific agreement (VALUE) | Category-specific agreement (NO_EXPLICIT_EXPRESSION_IDENTIFIED_IN_UNIT) | Cohen κ | Gwet AC1 |
|---|---|---|---|---|---|---|---|
| subject_population_as_stated | 61/74 (82.4%) | [73.2%, 89.4%] | 11 / 13 / 0 / 50 | 22/35 (62.9%) | 100/113 (88.5%) | 0.53 | 0.73 |
| context_as_stated | 66/74 (89.2%) | [81.1%, 94.4%] | 3 / 7 / 1 / 63 | 6/14 (42.9%) | 126/134 (94.0%) | 0.38 | 0.87 |
| condition_expression_as_stated | 62/74 (83.8%) | [74.8%, 90.4%] | 5 / 11 / 1 / 57 | 10/22 (45.5%) | 114/126 (90.5%) | 0.38 | 0.78 |
| proposition_content_as_stated | 70/74 (94.6%) | [87.7%, 98.0%] | 70 / 0 / 4 / 0 | 140/144 (97.2%) | 0/4 (0.0%) | 0.00 | 0.94 |
| object_intervention_as_stated | 65/74 (87.8%) | [79.5%, 93.4%] | 31 / 2 / 7 / 34 | 62/71 (87.3%) | 68/77 (88.3%) | 0.76 | 0.76 |
| alternative_as_stated | 72/74 (97.3%) | [91.7%, 99.3%] | 1 / 0 / 2 / 71 | 2/4 (50.0%) | 142/144 (98.6%) | 0.49 | 0.97 |
| modality_as_stated | 67/74 (90.5%) | [82.8%, 95.4%] | 17 / 7 / 0 / 50 | 34/41 (82.9%) | 100/107 (93.5%) | 0.77 | 0.84 |
| negation_expression_as_stated | 73/74 (98.6%) | [93.7%, 99.7%] | 11 / 0 / 1 / 62 | 22/23 (95.7%) | 124/125 (99.2%) | 0.95 | 0.98 |
| qualifier_as_stated | 58/74 (78.4%) | [68.9%, 86.1%] | 15 / 3 / 13 / 43 | 30/46 (65.2%) | 86/102 (84.3%) | 0.51 | 0.62 |
| exception_expression_as_stated | 73/74 (98.6%) | [93.7%, 99.7%] | 0 / 1 / 0 / 73 | 0/1 (0.0%) | 146/147 (99.3%) | 0.00 | 0.99 |
| quantitative_parameter_as_stated | 69/74 (93.2%) | [86.1%, 97.0%] | 6 / 4 / 1 / 63 | 12/17 (70.6%) | 126/131 (96.2%) | 0.67 | 0.92 |
| connective_as_stated | 63/74 (85.1%) | [76.5%, 91.4%] | 34 / 4 / 7 / 29 | 68/79 (86.1%) | 58/69 (84.1%) | 0.70 | 0.70 |
| cross_reference_as_stated | 71/74 (95.9%) | [89.7%, 98.7%] | 1 / 3 / 0 / 70 | 2/5 (40.0%) | 140/143 (97.9%) | 0.39 | 0.96 |
| actor_expression_as_stated | 72/74 (97.3%) | [91.7%, 99.3%] | 9 / 0 / 2 / 63 | 18/20 (90.0%) | 126/128 (98.4%) | 0.88 | 0.96 |

**NICE** — duplicated sample 60 of frame 156 units (n_h / N_h); intervals are exact design-based hypergeometric 95% intervals (HI-1).

| Slot | Raw agreement | HI-1 95% interval | Contingency V/V, V/N, N/V, N/N | Category-specific agreement (VALUE) | Category-specific agreement (NO_EXPLICIT_EXPRESSION_IDENTIFIED_IN_UNIT) | Cohen κ | Gwet AC1 |
|---|---|---|---|---|---|---|---|
| subject_population_as_stated | 51/60 (85.0%) | [76.3%, 91.0%] | 2 / 8 / 1 / 49 | 4/13 (30.8%) | 98/107 (91.6%) | 0.25 | 0.81 |
| context_as_stated | 51/60 (85.0%) | [76.3%, 91.0%] | 1 / 7 / 2 / 50 | 2/11 (18.2%) | 100/109 (91.7%) | 0.12 | 0.82 |
| condition_expression_as_stated | 55/60 (91.7%) | [84.0%, 96.2%] | 18 / 5 / 0 / 37 | 36/41 (87.8%) | 74/79 (93.7%) | 0.82 | 0.85 |
| proposition_content_as_stated | 51/60 (85.0%) | [76.3%, 91.0%] | 51 / 0 / 9 / 0 | 102/111 (91.9%) | 0/9 (0.0%) | 0.00 | 0.83 |
| object_intervention_as_stated | 48/60 (80.0%) | [70.5%, 87.2%] | 42 / 6 / 6 / 6 | 84/96 (87.5%) | 12/24 (50.0%) | 0.38 | 0.71 |
| alternative_as_stated | 59/60 (98.3%) | [92.9%, 99.4%] | 1 / 1 / 0 / 58 | 2/3 (66.7%) | 116/117 (99.1%) | 0.66 | 0.98 |

| Slot | Raw agreement | HI-1 95% interval | Contingency V/V, V/N, N/V, N/N | Category-specific agreement (VALUE) | Category-specific agreement (NO_EXPLICIT_EXPRESSION_IDENTIFIED_IN_UNIT) | Cohen κ | Gwet AC1 |
|---|---|---|---|---|---|---|---|
| modality_as_stated | 60/60 (100.0%) | [95.5%, 100.0%] | 25 / 0 / 0 / 35 | 50/50 (100.0%) | 70/70 (100.0%) | 1.00 | 1.00 |
| negation_expression_as_stated | 57/60 (95.0%) | [88.5%, 98.1%] | 7 / 0 / 3 / 50 | 14/17 (82.4%) | 100/103 (97.1%) | 0.80 | 0.93 |
| qualifier_as_stated | 46/60 (76.7%) | [66.7%, 84.6%] | 3 / 10 / 4 / 43 | 6/20 (30.0%) | 86/100 (86.0%) | 0.17 | 0.68 |
| exception_expression_as_stated | 59/60 (98.3%) | [92.9%, 99.4%] | 2 / 1 / 0 / 57 | 4/5 (80.0%) | 114/115 (99.1%) | 0.79 | 0.98 |
| quantitative_parameter_as_stated | 60/60 (100.0%) | [95.5%, 100.0%] | 8 / 0 / 0 / 52 | 16/16 (100.0%) | 104/104 (100.0%) | 1.00 | 1.00 |
| connective_as_stated | 58/60 (96.7%) | [90.4%, 98.7%] | 36 / 2 / 0 / 22 | 72/74 (97.3%) | 44/46 (95.7%) | 0.93 | 0.94 |
| cross_reference_as_stated | 59/60 (98.3%) | [92.9%, 99.4%] | 4 / 1 / 0 / 55 | 8/9 (88.9%) | 110/111 (99.1%) | 0.88 | 0.98 |
| actor_expression_as_stated | 58/60 (96.7%) | [90.4%, 98.7%] | 9 / 1 / 1 / 49 | 18/20 (90.0%) | 98/100 (98.0%) | 0.88 | 0.95 |

V = VALUE; N = NO_EXPLICIT_EXPRESSION_IDENTIFIED_IN_UNIT. No benchmark is applied to any coefficient. A value shown as NOT COMPUTABLE is transcribed as such from the output.

#### Table S3-5. Inter-encoder agreement C-10 — all other estimands (C10_REPRODUCIBILITY.csv)

Each block (estimand, qualifier, guideline) is kept together; the contingency counts of each two-category estimand are listed beside its κ and AC1.

**L2_EXACT_TEXT_MATCH**

| Guideline | Measure | Value | Note |
|---|---|---|---|
| EGS | raw agreement | 83/83 (100.0%) | denominator: duplicated Class-A units |
| EGS | HI-1 95% interval | [96.0%, 100.0%] | design-based, finite frame N_h = 581 (P25-09) |
| AAO | raw agreement | 70/74 (94.6%) | denominator: duplicated Class-A units |
| AAO | HI-1 95% interval | [87.7%, 98.0%] | design-based, finite frame N_h = 302 (P25-09) |
| NICE | raw agreement | 60/60 (100.0%) | denominator: duplicated Class-A units |
| NICE | HI-1 95% interval | [95.5%, 100.0%] | design-based, finite frame N_h = 156 (P25-09) |

**PG06_OUTCOME_AGREEMENT** — SOURCE_SPECIFIED_DECISION_CONTENT_EVIDENCE

| Guideline | Measure | Value | Note |
|---|---|---|---|
| EGS | raw agreement | 65/83 (78.3%) | denominator: duplicated Class-A units |
| EGS | HI-1 95% interval | [68.7%, 86.1%] | design-based, finite frame N_h = 581 (P25-09) |
| EGS | contingency E1=EXPLICIT_EVIDENCE_IDENTIFIED / E2=EXPLICIT_EVIDENCE_IDENTIFIED | 30 | |
| EGS | contingency E1=EXPLICIT_EVIDENCE_IDENTIFIED / E2=NO_QUALIFYING_LICENSING_EXPRESSION_IDENTIFIED | 1 | |
| EGS | contingency E1=NO_QUALIFYING_LICENSING_EXPRESSION_IDENTIFIED / E2=EXPLICIT_EVIDENCE_IDENTIFIED | 17 | |
| EGS | contingency E1=NO_QUALIFYING_LICENSING_EXPRESSION_IDENTIFIED / E2=NO_QUALIFYING_LICENSING_EXPRESSION_IDENTIFIED | 35 | |
| EGS | category-specific agreement EXPLICIT_EVIDENCE_IDENTIFIED | 60/78 (76.9%) | 2 x both-EXPLICIT_EVIDENCE_IDENTIFIED / (E1-EXPLICIT_EVIDENCE_IDENTIFIED + E2-EXPLICIT_EVIDENCE_IDENTIFIED) (PG-15 sec. 11.2) |
| EGS | category-specific agreement NO_QUALIFYING_LICENSING_EXPRESSION_IDENTIFIED | 70/88 (79.5%) | 2 x both-NO_QUALIFYING_LICENSING_EXPRESSION_IDENTIFIED / (E1-NO_QUALIFYING_LICENSING_EXPRESSION_IDENTIFIED + E2-NO_QUALIFYING_LICENSING_EXPRESSION_IDENTIFIED) (PG-15 sec. 11.2) |
| EGS | Cohen kappa | 0.58 | closed scale; no interval; no benchmark (P25-10) |
| EGS | Gwet AC1 | 0.57 | closed scale (q = 2); no interval (P25-10) |

| Guideline | Measure | Value | Note |
|---|---|---|---|
| AAO | raw agreement | 50/74 (67.6%) | denominator: duplicated Class-A units |
| AAO | HI-1 95% interval | [57.3%, 76.5%] | design-based, finite frame N_h = 302 (P25-09) |
| AAO | contingency E1=EXPLICIT_EVIDENCE_IDENTIFIED / E2=EXPLICIT_EVIDENCE_IDENTIFIED | 28 | |
| AAO | contingency E1=EXPLICIT_EVIDENCE_IDENTIFIED / E2=NO_QUALIFYING_LICENSING_EXPRESSION_IDENTIFIED | 2 | |
| AAO | contingency E1=NO_QUALIFYING_LICENSING_EXPRESSION_IDENTIFIED / E2=EXPLICIT_EVIDENCE_IDENTIFIED | 22 | |
| AAO | contingency E1=NO_QUALIFYING_LICENSING_EXPRESSION_IDENTIFIED / E2=NO_QUALIFYING_LICENSING_EXPRESSION_IDENTIFIED | 22 | |
| AAO | category-specific agreement EXPLICIT_EVIDENCE_IDENTIFIED | 56/80 (70.0%) | 2 x both-EXPLICIT_EVIDENCE_IDENTIFIED / (E1-EXPLICIT_EVIDENCE_IDENTIFIED + E2-EXPLICIT_EVIDENCE_IDENTIFIED) (PG-15 sec. 11.2) |
| AAO | category-specific agreement NO_QUALIFYING_LICENSING_EXPRESSION_IDENTIFIED | 44/68 (64.7%) | 2 x both-NO_QUALIFYING_LICENSING_EXPRESSION_IDENTIFIED / (E1-NO_QUALIFYING_LICENSING_EXPRESSION_IDENTIFIED + E2-NO_QUALIFYING_LICENSING_EXPRESSION_IDENTIFIED) (PG-15 sec. 11.2) |
| AAO | Cohen kappa | 0.39 | closed scale; no interval; no benchmark (P25-10) |
| AAO | Gwet AC1 | 0.36 | closed scale (q = 2); no interval (P25-10) |
| NICE | raw agreement | 44/60 (73.3%) | denominator: duplicated Class-A units |
| NICE | HI-1 95% interval | [63.5%, 81.4%] | design-based, finite frame N_h = 156 (P25-09) |
| NICE | contingency E1=EXPLICIT_EVIDENCE_IDENTIFIED / E2=EXPLICIT_EVIDENCE_IDENTIFIED | 38 | |
| NICE | contingency E1=EXPLICIT_EVIDENCE_IDENTIFIED / E2=NO_QUALIFYING_LICENSING_EXPRESSION_IDENTIFIED | 0 | |
| NICE | contingency E1=NO_QUALIFYING_LICENSING_EXPRESSION_IDENTIFIED / E2=EXPLICIT_EVIDENCE_IDENTIFIED | 16 | |
| NICE | contingency E1=NO_QUALIFYING_LICENSING_EXPRESSION_IDENTIFIED / E2=NO_QUALIFYING_LICENSING_EXPRESSION_IDENTIFIED | 6 | |
| NICE | category-specific agreement EXPLICIT_EVIDENCE_IDENTIFIED | 76/92 (82.6%) | 2 x both-EXPLICIT_EVIDENCE_IDENTIFIED / (E1-EXPLICIT_EVIDENCE_IDENTIFIED + E2-EXPLICIT_EVIDENCE_IDENTIFIED) (PG-15 sec. 11.2) |
| NICE | category-specific agreement NO_QUALIFYING_LICENSING_EXPRESSION_IDENTIFIED | 12/28 (42.9%) | 2 x both-NO_QUALIFYING_LICENSING_EXPRESSION_IDENTIFIED / (E1-NO_QUALIFYING_LICENSING_EXPRESSION_IDENTIFIED + E2-NO_QUALIFYING_LICENSING_EXPRESSION_IDENTIFIED) (PG-15 sec. 11.2) |
| NICE | Cohen kappa | 0.32 | closed scale; no interval; no benchmark (P25-10) |
| NICE | Gwet AC1 | 0.58 | closed scale (q = 2); no interval (P25-10) |

#### **PG06_OUTCOME_AGREEMENT** — EXPLICIT_DECISION_AUTHORITY_ALLOCATION_EVIDENCE

| Guideline | Measure | Value | Note |
|---|---|---|---|
| EGS | raw agreement | 82/83 (98.8%) | denominator: duplicated Class-A units |
| EGS | HI-1 95% interval | [94.0%, 99.8%] | design-based, finite frame N_h = 581 (P25-09) |
| EGS | contingency E1=EXPLICIT_EVIDENCE_IDENTIFIED / E2=EXPLICIT_EVIDENCE_IDENTIFIED | 0 | |
| EGS | contingency E1=EXPLICIT_EVIDENCE_IDENTIFIED / E2=NO_QUALIFYING_LICENSING_EXPRESSION_IDENTIFIED | 0 | |
| EGS | contingency E1=NO_QUALIFYING_LICENSING_EXPRESSION_IDENTIFIED / E2=EXPLICIT_EVIDENCE_IDENTIFIED | 1 | |
| EGS | contingency E1=NO_QUALIFYING_LICENSING_EXPRESSION_IDENTIFIED / E2=NO_QUALIFYING_LICENSING_EXPRESSION_IDENTIFIED | 82 | |
| EGS | category-specific agreement EXPLICIT_EVIDENCE_IDENTIFIED | 0/1 (0.0%) | 2 x both-EXPLICIT_EVIDENCE_IDENTIFIED / (E1-EXPLICIT_EVIDENCE_IDENTIFIED + E2-EXPLICIT_EVIDENCE_IDENTIFIED) (PG-15 sec. 11.2) |
| EGS | category-specific agreement NO_QUALIFYING_LICENSING_EXPRESSION_IDENTIFIED | 164/165 (99.4%) | 2 x both-NO_QUALIFYING_LICENSING_EXPRESSION_IDENTIFIED / (E1-NO_QUALIFYING_LICENSING_EXPRESSION_IDENTIFIED + E2-NO_QUALIFYING_LICENSING_EXPRESSION_IDENTIFIED) (PG-15 sec. 11.2) |
| EGS | Cohen kappa | 0.00 | closed scale; no interval; no benchmark (P25-10) |
| EGS | Gwet AC1 | 0.99 | closed scale (q = 2); no interval (P25-10) |

| Guideline | Measure | Value | Note |
|---|---|---|---|
| AAO | raw agreement | 73/74 (98.6%) | denominator: duplicated Class-A units |
| AAO | HI-1 95% interval | [93.7%, 99.7%] | design-based, finite frame N_h = 302 (P25-09) |
| AAO | contingency E1=EXPLICIT_EVIDENCE_IDENTIFIED / E2=EXPLICIT_EVIDENCE_IDENTIFIED | 1 | |
| AAO | contingency E1=EXPLICIT_EVIDENCE_IDENTIFIED / E2=NO_QUALIFYING_LICENSING_EXPRESSION_IDENTIFIED | 0 | |
| AAO | contingency E1=NO_QUALIFYING_LICENSING_EXPRESSION_IDENTIFIED / E2=EXPLICIT_EVIDENCE_IDENTIFIED | 1 | |
| AAO | contingency E1=NO_QUALIFYING_LICENSING_EXPRESSION_IDENTIFIED / E2=NO_QUALIFYING_LICENSING_EXPRESSION_IDENTIFIED | 72 | |
| AAO | category-specific agreement EXPLICIT_EVIDENCE_IDENTIFIED | 2/3 (66.7%) | 2 x both-EXPLICIT_EVIDENCE_IDENTIFIED / (E1-EXPLICIT_EVIDENCE_IDENTIFIED + E2-EXPLICIT_EVIDENCE_IDENTIFIED) (PG-15 sec. 11.2) |
| AAO | category-specific agreement NO_QUALIFYING_LICENSING_EXPRESSION_IDENTIFIED | 144/145 (99.3%) | 2 x both-NO_QUALIFYING_LICENSING_EXPRESSION_IDENTIFIED / (E1-NO_QUALIFYING_LICENSING_EXPRESSION_IDENTIFIED + E2-NO_QUALIFYING_LICENSING_EXPRESSION_IDENTIFIED) (PG-15 sec. 11.2) |
| AAO | Cohen kappa | 0.66 | closed scale; no interval; no benchmark (P25-10) |
| AAO | Gwet AC1 | 0.99 | closed scale (q = 2); no interval (P25-10) |
| NICE | raw agreement | 59/60 (98.3%) | denominator: duplicated Class-A units |
| NICE | HI-1 95% interval | [92.9%, 99.4%] | design-based, finite frame N_h = 156 (P25-09) |
| NICE | contingency E1=EXPLICIT_EVIDENCE_IDENTIFIED / E2=EXPLICIT_EVIDENCE_IDENTIFIED | 3 | |
| NICE | contingency E1=EXPLICIT_EVIDENCE_IDENTIFIED / E2=NO_QUALIFYING_LICENSING_EXPRESSION_IDENTIFIED | 0 | |
| NICE | contingency E1=NO_QUALIFYING_LICENSING_EXPRESSION_IDENTIFIED / E2=EXPLICIT_EVIDENCE_IDENTIFIED | 1 | |
| NICE | contingency E1=NO_QUALIFYING_LICENSING_EXPRESSION_IDENTIFIED / E2=NO_QUALIFYING_LICENSING_EXPRESSION_IDENTIFIED | 56 | |
| NICE | category-specific agreement EXPLICIT_EVIDENCE_IDENTIFIED | 6/7 (85.7%) | 2 x both-EXPLICIT_EVIDENCE_IDENTIFIED / (E1-EXPLICIT_EVIDENCE_IDENTIFIED + E2-EXPLICIT_EVIDENCE_IDENTIFIED) (PG-15 sec. 11.2) |
| NICE | category-specific agreement NO_QUALIFYING_LICENSING_EXPRESSION_IDENTIFIED | 112/113 (99.1%) | 2 x both-NO_QUALIFYING_LICENSING_EXPRESSION_IDENTIFIED / (E1-NO_QUALIFYING_LICENSING_EXPRESSION_IDENTIFIED + E2-NO_QUALIFYING_LICENSING_EXPRESSION_IDENTIFIED) (PG-15 sec. 11.2) |
| NICE | Cohen kappa | 0.85 | closed scale; no interval; no benchmark (P25-10) |
| NICE | Gwet AC1 | 0.98 | closed scale (q = 2); no interval (P25-10) |

#### STAGE1_RAISED_AGREEMENT

| Guideline | Measure | Value | Note |
|---|---|---|---|
| EGS | raw agreement | 83/83 (100.0%) | denominator: duplicated Class-A units |
| EGS | HI-1 95% interval | [96.0%, 100.0%] | design-based, finite frame N_h = 581 (P25-09) |
| EGS | contingency E1=FALSE / E2=FALSE | 83 | |
| EGS | contingency E1=FALSE / E2=TRUE | 0 | |
| EGS | contingency E1=TRUE / E2=FALSE | 0 | |
| EGS | contingency E1=TRUE / E2=TRUE | 0 | |
| EGS | category-specific agreement FALSE | 166/166 (100.0%) | 2 x both-FALSE / (E1-FALSE + E2-FALSE) (PG-15 sec. 11.2) |
| EGS | category-specific agreement TRUE | NOT COMPUTABLE | 2 x both-TRUE / (E1-TRUE + E2-TRUE) (PG-15 sec. 11.2) |
| EGS | Cohen kappa | NOT COMPUTABLE | closed scale; no interval; no benchmark (P25-10) |
| EGS | Gwet AC1 | 1.00 | closed scale (q = 2); no interval (P25-10) |

| Guideline | Measure | Value | Note |
|---|---|---|---|
| AAO | raw agreement | 74/74 (100.0%) | denominator: duplicated Class-A units |
| AAO | HI-1 95% interval | [96.0%, 100.0%] | design-based, finite frame N_h = 302 (P25-09) |
| AAO | contingency E1=FALSE / E2=FALSE | 74 | |
| AAO | contingency E1=FALSE / E2=TRUE | 0 | |
| AAO | contingency E1=TRUE / E2=FALSE | 0 | |
| AAO | contingency E1=TRUE / E2=TRUE | 0 | |
| AAO | category-specific agreement FALSE | 148/148 (100.0%) | 2 x both-FALSE / (E1-FALSE + E2-FALSE) (PG-15 sec. 11.2) |
| AAO | category-specific agreement TRUE | NOT COMPUTABLE | 2 x both-TRUE / (E1-TRUE + E2-TRUE) (PG-15 sec. 11.2) |
| AAO | Cohen kappa | NOT COMPUTABLE | closed scale; no interval; no benchmark (P25-10) |
| AAO | Gwet AC1 | 1.00 | closed scale (q = 2); no interval (P25-10) |
| NICE | raw agreement | 60/60 (100.0%) | denominator: duplicated Class-A units |
| NICE | HI-1 95% interval | [95.5%, 100.0%] | design-based, finite frame N_h = 156 (P25-09) |
| NICE | contingency E1=FALSE / E2=FALSE | 60 | |
| NICE | contingency E1=FALSE / E2=TRUE | 0 | |
| NICE | contingency E1=TRUE / E2=FALSE | 0 | |
| NICE | contingency E1=TRUE / E2=TRUE | 0 | |
| NICE | category-specific agreement FALSE | 120/120 (100.0%) | 2 x both-FALSE / (E1-FALSE + E2-FALSE) (PG-15 sec. 11.2) |
| NICE | category-specific agreement TRUE | NOT COMPUTABLE | 2 x both-TRUE / (E1-TRUE + E2-TRUE) (PG-15 sec. 11.2) |
| NICE | Cohen kappa | NOT COMPUTABLE | closed scale; no interval; no benchmark (P25-10) |
| NICE | Gwet AC1 | 1.00 | closed scale (q = 2); no interval (P25-10) |

##### **L3_VALUE_EXACT_TEXT_MATCH** — subject_population_as_stated

| Guideline | Measure | Value | Note |
|---|---|---|---|
| EGS | exact match | 10/10 (100.0%) | denominator: VALUE/VALUE pairs (conditional) |
| AAO | exact match | 8/11 (72.7%) | denominator: VALUE/VALUE pairs (conditional) |
| NICE | exact match | 2/2 (100.0%) | denominator: VALUE/VALUE pairs (conditional) |

##### **EVIDENCE_SPAN_CONCORDANCE** — L3_VALUE subject_population_as_stated

| Guideline | Measure | Value | Note |
|---|---|---|---|
| EGS | IDENTICAL | 10/10 (100.0%) | denominator: VALUE/VALUE pairs (descriptive) |
| EGS | OVERLAPPING | 0/10 (0.0%) | denominator: VALUE/VALUE pairs (descriptive) |
| EGS | DISJOINT | 0/10 (0.0%) | denominator: VALUE/VALUE pairs (descriptive) |
| EGS | ONE_SIDED | 0/10 (0.0%) | denominator: VALUE/VALUE pairs (descriptive) |
| EGS | NOT_APPLICABLE | 0/10 (0.0%) | denominator: VALUE/VALUE pairs (descriptive) |
| AAO | IDENTICAL | 8/11 (72.7%) | denominator: VALUE/VALUE pairs (descriptive) |
| AAO | OVERLAPPING | 3/11 (27.3%) | denominator: VALUE/VALUE pairs (descriptive) |
| AAO | DISJOINT | 0/11 (0.0%) | denominator: VALUE/VALUE pairs (descriptive) |
| AAO | ONE_SIDED | 0/11 (0.0%) | denominator: VALUE/VALUE pairs (descriptive) |
| AAO | NOT_APPLICABLE | 0/11 (0.0%) | denominator: VALUE/VALUE pairs (descriptive) |
| NICE | IDENTICAL | 2/2 (100.0%) | denominator: VALUE/VALUE pairs (descriptive) |
| NICE | OVERLAPPING | 0/2 (0.0%) | denominator: VALUE/VALUE pairs (descriptive) |
| NICE | DISJOINT | 0/2 (0.0%) | denominator: VALUE/VALUE pairs (descriptive) |
| NICE | ONE_SIDED | 0/2 (0.0%) | denominator: VALUE/VALUE pairs (descriptive) |
| NICE | NOT_APPLICABLE | 0/2 (0.0%) | denominator: VALUE/VALUE pairs (descriptive) |

**L3_VALUE_EXACT_TEXT_MATCH** — context_as_stated

| Guideline | Measure | Value | Note |
|---|---|---|---|
| EGS | exact match | 7/9 (77.8%) | denominator: VALUE/VALUE pairs (conditional) |
| AAO | exact match | 3/3 (100.0%) | denominator: VALUE/VALUE pairs (conditional) |
| NICE | exact match | 1/1 (100.0%) | denominator: VALUE/VALUE pairs (conditional) |

**EVIDENCE_SPAN_CONCORDANCE** — L3_VALUE context_as_stated

| Guideline | Measure | Value | Note |
|---|---|---|---|
| EGS | IDENTICAL | 7/9 (77.8%) | denominator: VALUE/VALUE pairs (descriptive) |
| EGS | OVERLAPPING | 1/9 (11.1%) | denominator: VALUE/VALUE pairs (descriptive) |
| EGS | DISJOINT | 1/9 (11.1%) | denominator: VALUE/VALUE pairs (descriptive) |
| EGS | ONE_SIDED | 0/9 (0.0%) | denominator: VALUE/VALUE pairs (descriptive) |
| EGS | NOT_APPLICABLE | 0/9 (0.0%) | denominator: VALUE/VALUE pairs (descriptive) |
| AAO | IDENTICAL | 3/3 (100.0%) | denominator: VALUE/VALUE pairs (descriptive) |
| AAO | OVERLAPPING | 0/3 (0.0%) | denominator: VALUE/VALUE pairs (descriptive) |
| AAO | DISJOINT | 0/3 (0.0%) | denominator: VALUE/VALUE pairs (descriptive) |
| AAO | ONE_SIDED | 0/3 (0.0%) | denominator: VALUE/VALUE pairs (descriptive) |
| AAO | NOT_APPLICABLE | 0/3 (0.0%) | denominator: VALUE/VALUE pairs (descriptive) |
| NICE | IDENTICAL | 1/1 (100.0%) | denominator: VALUE/VALUE pairs (descriptive) |
| NICE | OVERLAPPING | 0/1 (0.0%) | denominator: VALUE/VALUE pairs (descriptive) |
| NICE | DISJOINT | 0/1 (0.0%) | denominator: VALUE/VALUE pairs (descriptive) |
| NICE | ONE_SIDED | 0/1 (0.0%) | denominator: VALUE/VALUE pairs (descriptive) |
| NICE | NOT_APPLICABLE | 0/1 (0.0%) | denominator: VALUE/VALUE pairs (descriptive) |

**L3_VALUE_EXACT_TEXT_MATCH** — condition_expression_as_stated

| Guideline | Measure | Value | Note |
|---|---|---|---|
| EGS | exact match | 4/9 (44.4%) | denominator: VALUE/VALUE pairs (conditional) |
| AAO | exact match | 5/5 (100.0%) | denominator: VALUE/VALUE pairs (conditional) |
| NICE | exact match | 11/18 (61.1%) | denominator: VALUE/VALUE pairs (conditional) |

**EVIDENCE_SPAN_CONCORDANCE** — L3_VALUE condition_expression_as_stated

| Guideline | Measure | Value | Note |
|---|---|---|---|
| EGS | IDENTICAL | 4/9 (44.4%) | denominator: VALUE/VALUE pairs (descriptive) |
| EGS | OVERLAPPING | 5/9 (55.6%) | denominator: VALUE/VALUE pairs (descriptive) |
| EGS | DISJOINT | 0/9 (0.0%) | denominator: VALUE/VALUE pairs (descriptive) |
| EGS | ONE_SIDED | 0/9 (0.0%) | denominator: VALUE/VALUE pairs (descriptive) |
| EGS | NOT_APPLICABLE | 0/9 (0.0%) | denominator: VALUE/VALUE pairs (descriptive) |
| AAO | IDENTICAL | 5/5 (100.0%) | denominator: VALUE/VALUE pairs (descriptive) |
| AAO | OVERLAPPING | 0/5 (0.0%) | denominator: VALUE/VALUE pairs (descriptive) |
| AAO | DISJOINT | 0/5 (0.0%) | denominator: VALUE/VALUE pairs (descriptive) |
| AAO | ONE_SIDED | 0/5 (0.0%) | denominator: VALUE/VALUE pairs (descriptive) |
| AAO | NOT_APPLICABLE | 0/5 (0.0%) | denominator: VALUE/VALUE pairs (descriptive) |
| NICE | IDENTICAL | 11/18 (61.1%) | denominator: VALUE/VALUE pairs (descriptive) |
| NICE | OVERLAPPING | 7/18 (38.9%) | denominator: VALUE/VALUE pairs (descriptive) |
| NICE | DISJOINT | 0/18 (0.0%) | denominator: VALUE/VALUE pairs (descriptive) |
| NICE | ONE_SIDED | 0/18 (0.0%) | denominator: VALUE/VALUE pairs (descriptive) |
| NICE | NOT_APPLICABLE | 0/18 (0.0%) | denominator: VALUE/VALUE pairs (descriptive) |

**L3_VALUE_EXACT_TEXT_MATCH** — proposition_content_as_stated

| Guideline | Measure | Value | Note |
|---|---|---|---|
| EGS | exact match | 13/81 (16.0%) | denominator: VALUE/VALUE pairs (conditional) |
| AAO | exact match | 29/70 (41.4%) | denominator: VALUE/VALUE pairs (conditional) |
| NICE | exact match | 11/51 (21.6%) | denominator: VALUE/VALUE pairs (conditional) |

**EVIDENCE_SPAN_CONCORDANCE** — L3_VALUE proposition_content_as_stated

| Guideline | Measure | Value | Note |
|---|---|---|---|
| EGS | IDENTICAL | 13/81 (16.0%) | denominator: VALUE/VALUE pairs (descriptive) |
| EGS | OVERLAPPING | 68/81 (84.0%) | denominator: VALUE/VALUE pairs (descriptive) |
| EGS | DISJOINT | 0/81 (0.0%) | denominator: VALUE/VALUE pairs (descriptive) |
| EGS | ONE_SIDED | 0/81 (0.0%) | denominator: VALUE/VALUE pairs (descriptive) |
| EGS | NOT_APPLICABLE | 0/81 (0.0%) | denominator: VALUE/VALUE pairs (descriptive) |
| AAO | IDENTICAL | 29/70 (41.4%) | denominator: VALUE/VALUE pairs (descriptive) |
| AAO | OVERLAPPING | 41/70 (58.6%) | denominator: VALUE/VALUE pairs (descriptive) |
| AAO | DISJOINT | 0/70 (0.0%) | denominator: VALUE/VALUE pairs (descriptive) |
| AAO | ONE_SIDED | 0/70 (0.0%) | denominator: VALUE/VALUE pairs (descriptive) |
| AAO | NOT_APPLICABLE | 0/70 (0.0%) | denominator: VALUE/VALUE pairs (descriptive) |
| NICE | IDENTICAL | 11/51 (21.6%) | denominator: VALUE/VALUE pairs (descriptive) |
| NICE | OVERLAPPING | 40/51 (78.4%) | denominator: VALUE/VALUE pairs (descriptive) |
| NICE | DISJOINT | 0/51 (0.0%) | denominator: VALUE/VALUE pairs (descriptive) |
| NICE | ONE_SIDED | 0/51 (0.0%) | denominator: VALUE/VALUE pairs (descriptive) |
| NICE | NOT_APPLICABLE | 0/51 (0.0%) | denominator: VALUE/VALUE pairs (descriptive) |

**L3_VALUE_EXACT_TEXT_MATCH** — object_intervention_as_stated

| Guideline | Measure | Value | Note |
|---|---|---|---|
| EGS | exact match | 15/35 (42.9%) | denominator: VALUE/VALUE pairs (conditional) |
| AAO | exact match | 19/31 (61.3%) | denominator: VALUE/VALUE pairs (conditional) |
| NICE | exact match | 29/42 (69.0%) | denominator: VALUE/VALUE pairs (conditional) |

**EVIDENCE_SPAN_CONCORDANCE** — L3_VALUE object_intervention_as_stated

| Guideline | Measure | Value | Note |
|---|---|---|---|
| EGS | IDENTICAL | 15/35 (42.9%) | denominator: VALUE/VALUE pairs (descriptive) |
| EGS | OVERLAPPING | 19/35 (54.3%) | denominator: VALUE/VALUE pairs (descriptive) |
| EGS | DISJOINT | 1/35 (2.9%) | denominator: VALUE/VALUE pairs (descriptive) |
| EGS | ONE_SIDED | 0/35 (0.0%) | denominator: VALUE/VALUE pairs (descriptive) |
| EGS | NOT_APPLICABLE | 0/35 (0.0%) | denominator: VALUE/VALUE pairs (descriptive) |
| AAO | IDENTICAL | 19/31 (61.3%) | denominator: VALUE/VALUE pairs (descriptive) |
| AAO | OVERLAPPING | 11/31 (35.5%) | denominator: VALUE/VALUE pairs (descriptive) |
| AAO | DISJOINT | 1/31 (3.2%) | denominator: VALUE/VALUE pairs (descriptive) |
| AAO | ONE_SIDED | 0/31 (0.0%) | denominator: VALUE/VALUE pairs (descriptive) |
| AAO | NOT_APPLICABLE | 0/31 (0.0%) | denominator: VALUE/VALUE pairs (descriptive) |

| Guideline | Measure | Value | Note |
|---|---|---|---|
| NICE | IDENTICAL | 29/42 (69.0%) | denominator: VALUE/VALUE pairs (descriptive) |
| NICE | OVERLAPPING | 9/42 (21.4%) | denominator: VALUE/VALUE pairs (descriptive) |
| NICE | DISJOINT | 4/42 (9.5%) | denominator: VALUE/VALUE pairs (descriptive) |
| NICE | ONE_SIDED | 0/42 (0.0%) | denominator: VALUE/VALUE pairs (descriptive) |
| NICE | NOT_APPLICABLE | 0/42 (0.0%) | denominator: VALUE/VALUE pairs (descriptive) |

##### **L3_VALUE_EXACT_TEXT_MATCH** — alternative_as_stated

| Guideline | Measure | Value | Note |
|---|---|---|---|
| EGS | exact match | 0/2 (0.0%) | denominator: VALUE/VALUE pairs (conditional) |
| AAO | exact match | 0/1 (0.0%) | denominator: VALUE/VALUE pairs (conditional) |
| NICE | exact match | 0/1 (0.0%) | denominator: VALUE/VALUE pairs (conditional) |

##### **EVIDENCE_SPAN_CONCORDANCE** — L3_VALUE alternative_as_stated

| Guideline | Measure | Value | Note |
|---|---|---|---|
| EGS | IDENTICAL | 0/2 (0.0%) | denominator: VALUE/VALUE pairs (descriptive) |
| EGS | OVERLAPPING | 2/2 (100.0%) | denominator: VALUE/VALUE pairs (descriptive) |
| EGS | DISJOINT | 0/2 (0.0%) | denominator: VALUE/VALUE pairs (descriptive) |
| EGS | ONE_SIDED | 0/2 (0.0%) | denominator: VALUE/VALUE pairs (descriptive) |
| EGS | NOT_APPLICABLE | 0/2 (0.0%) | denominator: VALUE/VALUE pairs (descriptive) |
| AAO | IDENTICAL | 0/1 (0.0%) | denominator: VALUE/VALUE pairs (descriptive) |
| AAO | OVERLAPPING | 1/1 (100.0%) | denominator: VALUE/VALUE pairs (descriptive) |
| AAO | DISJOINT | 0/1 (0.0%) | denominator: VALUE/VALUE pairs (descriptive) |
| AAO | ONE_SIDED | 0/1 (0.0%) | denominator: VALUE/VALUE pairs (descriptive) |
| AAO | NOT_APPLICABLE | 0/1 (0.0%) | denominator: VALUE/VALUE pairs (descriptive) |
| NICE | IDENTICAL | 0/1 (0.0%) | denominator: VALUE/VALUE pairs (descriptive) |
| NICE | OVERLAPPING | 1/1 (100.0%) | denominator: VALUE/VALUE pairs (descriptive) |
| NICE | DISJOINT | 0/1 (0.0%) | denominator: VALUE/VALUE pairs (descriptive) |
| NICE | ONE_SIDED | 0/1 (0.0%) | denominator: VALUE/VALUE pairs (descriptive) |
| NICE | NOT_APPLICABLE | 0/1 (0.0%) | denominator: VALUE/VALUE pairs (descriptive) |

##### **L3_VALUE_EXACT_TEXT_MATCH** — modality_as_stated

| Guideline | Measure | Value | Note |
|---|---|---|---|
| EGS | exact match | 4/26 (15.4%) | denominator: VALUE/VALUE pairs (conditional) |
| AAO | exact match | 1/17 (5.9%) | denominator: VALUE/VALUE pairs (conditional) |
| NICE | exact match | 12/25 (48.0%) | denominator: VALUE/VALUE pairs (conditional) |

##### **EVIDENCE_SPAN_CONCORDANCE** — L3_VALUE modality_as_stated

| Guideline | Measure | Value | Note |
|---|---|---|---|
| EGS | IDENTICAL | 4/26 (15.4%) | denominator: VALUE/VALUE pairs (descriptive) |
| EGS | OVERLAPPING | 21/26 (80.8%) | denominator: VALUE/VALUE pairs (descriptive) |
| EGS | DISJOINT | 1/26 (3.8%) | denominator: VALUE/VALUE pairs (descriptive) |
| EGS | ONE_SIDED | 0/26 (0.0%) | denominator: VALUE/VALUE pairs (descriptive) |
| EGS | NOT_APPLICABLE | 0/26 (0.0%) | denominator: VALUE/VALUE pairs (descriptive) |

| Guideline | Measure | Value | Note |
|---|---|---|---|
| AAO | IDENTICAL | 1/17 (5.9%) | denominator: VALUE/VALUE pairs (descriptive) |
| AAO | OVERLAPPING | 16/17 (94.1%) | denominator: VALUE/VALUE pairs (descriptive) |
| AAO | DISJOINT | 0/17 (0.0%) | denominator: VALUE/VALUE pairs (descriptive) |
| AAO | ONE_SIDED | 0/17 (0.0%) | denominator: VALUE/VALUE pairs (descriptive) |
| AAO | NOT_APPLICABLE | 0/17 (0.0%) | denominator: VALUE/VALUE pairs (descriptive) |
| NICE | IDENTICAL | 12/25 (48.0%) | denominator: VALUE/VALUE pairs (descriptive) |
| NICE | OVERLAPPING | 13/25 (52.0%) | denominator: VALUE/VALUE pairs (descriptive) |
| NICE | DISJOINT | 0/25 (0.0%) | denominator: VALUE/VALUE pairs (descriptive) |
| NICE | ONE_SIDED | 0/25 (0.0%) | denominator: VALUE/VALUE pairs (descriptive) |
| NICE | NOT_APPLICABLE | 0/25 (0.0%) | denominator: VALUE/VALUE pairs (descriptive) |

##### **L3_VALUE_EXACT_TEXT_MATCH** — negation_expression_as_stated

| Guideline | Measure | Value | Note |
|---|---|---|---|
| EGS | exact match | 3/11 (27.3%) | denominator: VALUE/VALUE pairs (conditional) |
| AAO | exact match | 1/11 (9.1%) | denominator: VALUE/VALUE pairs (conditional) |
| NICE | exact match | 2/7 (28.6%) | denominator: VALUE/VALUE pairs (conditional) |

##### **EVIDENCE_SPAN_CONCORDANCE** — L3_VALUE negation_expression_as_stated

| Guideline | Measure | Value | Note |
|---|---|---|---|
| EGS | IDENTICAL | 3/11 (27.3%) | denominator: VALUE/VALUE pairs (descriptive) |
| EGS | OVERLAPPING | 8/11 (72.7%) | denominator: VALUE/VALUE pairs (descriptive) |
| EGS | DISJOINT | 0/11 (0.0%) | denominator: VALUE/VALUE pairs (descriptive) |
| EGS | ONE_SIDED | 0/11 (0.0%) | denominator: VALUE/VALUE pairs (descriptive) |
| EGS | NOT_APPLICABLE | 0/11 (0.0%) | denominator: VALUE/VALUE pairs (descriptive) |
| AAO | IDENTICAL | 1/11 (9.1%) | denominator: VALUE/VALUE pairs (descriptive) |
| AAO | OVERLAPPING | 10/11 (90.9%) | denominator: VALUE/VALUE pairs (descriptive) |
| AAO | DISJOINT | 0/11 (0.0%) | denominator: VALUE/VALUE pairs (descriptive) |
| AAO | ONE_SIDED | 0/11 (0.0%) | denominator: VALUE/VALUE pairs (descriptive) |
| AAO | NOT_APPLICABLE | 0/11 (0.0%) | denominator: VALUE/VALUE pairs (descriptive) |
| NICE | IDENTICAL | 2/7 (28.6%) | denominator: VALUE/VALUE pairs (descriptive) |
| NICE | OVERLAPPING | 4/7 (57.1%) | denominator: VALUE/VALUE pairs (descriptive) |
| NICE | DISJOINT | 1/7 (14.3%) | denominator: VALUE/VALUE pairs (descriptive) |
| NICE | ONE_SIDED | 0/7 (0.0%) | denominator: VALUE/VALUE pairs (descriptive) |
| NICE | NOT_APPLICABLE | 0/7 (0.0%) | denominator: VALUE/VALUE pairs (descriptive) |

##### **L3_VALUE_EXACT_TEXT_MATCH** — qualifier_as_stated

| Guideline | Measure | Value | Note |
|---|---|---|---|
| EGS | exact match | 14/24 (58.3%) | denominator: VALUE/VALUE pairs (conditional) |
| AAO | exact match | 3/15 (20.0%) | denominator: VALUE/VALUE pairs (conditional) |
| NICE | exact match | 0/3 (0.0%) | denominator: VALUE/VALUE pairs (conditional) |

**EVIDENCE_SPAN_CONCORDANCE** — L3_VALUE qualifier_as_stated

| Guideline | Measure | Value | Note |
|---|---|---|---|
| EGS | IDENTICAL | 14/24 (58.3%) | denominator: VALUE/VALUE pairs (descriptive) |
| EGS | OVERLAPPING | 9/24 (37.5%) | denominator: VALUE/VALUE pairs (descriptive) |
| EGS | DISJOINT | 1/24 (4.2%) | denominator: VALUE/VALUE pairs (descriptive) |
| EGS | ONE_SIDED | 0/24 (0.0%) | denominator: VALUE/VALUE pairs (descriptive) |
| EGS | NOT_APPLICABLE | 0/24 (0.0%) | denominator: VALUE/VALUE pairs (descriptive) |
| AAO | IDENTICAL | 3/15 (20.0%) | denominator: VALUE/VALUE pairs (descriptive) |
| AAO | OVERLAPPING | 12/15 (80.0%) | denominator: VALUE/VALUE pairs (descriptive) |
| AAO | DISJOINT | 0/15 (0.0%) | denominator: VALUE/VALUE pairs (descriptive) |
| AAO | ONE_SIDED | 0/15 (0.0%) | denominator: VALUE/VALUE pairs (descriptive) |
| AAO | NOT_APPLICABLE | 0/15 (0.0%) | denominator: VALUE/VALUE pairs (descriptive) |
| NICE | IDENTICAL | 0/3 (0.0%) | denominator: VALUE/VALUE pairs (descriptive) |
| NICE | OVERLAPPING | 3/3 (100.0%) | denominator: VALUE/VALUE pairs (descriptive) |
| NICE | DISJOINT | 0/3 (0.0%) | denominator: VALUE/VALUE pairs (descriptive) |
| NICE | ONE_SIDED | 0/3 (0.0%) | denominator: VALUE/VALUE pairs (descriptive) |
| NICE | NOT_APPLICABLE | 0/3 (0.0%) | denominator: VALUE/VALUE pairs (descriptive) |

**L3_VALUE_EXACT_TEXT_MATCH** — exception_expression_as_stated

| Guideline | Measure | Value | Note |
|---|---|---|---|
| EGS | exact match | NOT COMPUTABLE | denominator: VALUE/VALUE pairs (conditional) |
| AAO | exact match | NOT COMPUTABLE | denominator: VALUE/VALUE pairs (conditional) |
| NICE | exact match | 2/2 (100.0%) | denominator: VALUE/VALUE pairs (conditional) |

**EVIDENCE_SPAN_CONCORDANCE** — L3_VALUE exception_expression_as_stated

| Guideline | Measure | Value | Note |
|---|---|---|---|
| EGS | IDENTICAL | NOT COMPUTABLE | denominator: VALUE/VALUE pairs (descriptive) |
| EGS | OVERLAPPING | NOT COMPUTABLE | denominator: VALUE/VALUE pairs (descriptive) |
| EGS | DISJOINT | NOT COMPUTABLE | denominator: VALUE/VALUE pairs (descriptive) |
| EGS | ONE_SIDED | NOT COMPUTABLE | denominator: VALUE/VALUE pairs (descriptive) |
| EGS | NOT_APPLICABLE | NOT COMPUTABLE | denominator: VALUE/VALUE pairs (descriptive) |
| AAO | IDENTICAL | NOT COMPUTABLE | denominator: VALUE/VALUE pairs (descriptive) |
| AAO | OVERLAPPING | NOT COMPUTABLE | denominator: VALUE/VALUE pairs (descriptive) |
| AAO | DISJOINT | NOT COMPUTABLE | denominator: VALUE/VALUE pairs (descriptive) |
| AAO | ONE_SIDED | NOT COMPUTABLE | denominator: VALUE/VALUE pairs (descriptive) |
| AAO | NOT_APPLICABLE | NOT COMPUTABLE | denominator: VALUE/VALUE pairs (descriptive) |
| NICE | IDENTICAL | 2/2 (100.0%) | denominator: VALUE/VALUE pairs (descriptive) |
| NICE | OVERLAPPING | 0/2 (0.0%) | denominator: VALUE/VALUE pairs (descriptive) |
| NICE | DISJOINT | 0/2 (0.0%) | denominator: VALUE/VALUE pairs (descriptive) |
| NICE | ONE_SIDED | 0/2 (0.0%) | denominator: VALUE/VALUE pairs (descriptive) |
| NICE | NOT_APPLICABLE | 0/2 (0.0%) | denominator: VALUE/VALUE pairs (descriptive) |

**L3_VALUE_EXACT_TEXT_MATCH** — quantitative_parameter_as_stated

| Guideline | Measure | Value | Note |
|---|---|---|---|
| EGS | exact match | 10/17 (58.8%) | denominator: VALUE/VALUE pairs (conditional) |
| AAO | exact match | 3/6 (50.0%) | denominator: VALUE/VALUE pairs (conditional) |
| NICE | exact match | 6/8 (75.0%) | denominator: VALUE/VALUE pairs (conditional) |

**EVIDENCE_SPAN_CONCORDANCE** — L3_VALUE quantitative_parameter_as_stated

| Guideline | Measure | Value | Note |
|---|---|---|---|
| EGS | IDENTICAL | 10/17 (58.8%) | denominator: VALUE/VALUE pairs (descriptive) |
| EGS | OVERLAPPING | 7/17 (41.2%) | denominator: VALUE/VALUE pairs (descriptive) |
| EGS | DISJOINT | 0/17 (0.0%) | denominator: VALUE/VALUE pairs (descriptive) |
| EGS | ONE_SIDED | 0/17 (0.0%) | denominator: VALUE/VALUE pairs (descriptive) |
| EGS | NOT_APPLICABLE | 0/17 (0.0%) | denominator: VALUE/VALUE pairs (descriptive) |
| AAO | IDENTICAL | 3/6 (50.0%) | denominator: VALUE/VALUE pairs (descriptive) |
| AAO | OVERLAPPING | 3/6 (50.0%) | denominator: VALUE/VALUE pairs (descriptive) |
| AAO | DISJOINT | 0/6 (0.0%) | denominator: VALUE/VALUE pairs (descriptive) |
| AAO | ONE_SIDED | 0/6 (0.0%) | denominator: VALUE/VALUE pairs (descriptive) |
| AAO | NOT_APPLICABLE | 0/6 (0.0%) | denominator: VALUE/VALUE pairs (descriptive) |
| NICE | IDENTICAL | 6/8 (75.0%) | denominator: VALUE/VALUE pairs (descriptive) |
| NICE | OVERLAPPING | 2/8 (25.0%) | denominator: VALUE/VALUE pairs (descriptive) |
| NICE | DISJOINT | 0/8 (0.0%) | denominator: VALUE/VALUE pairs (descriptive) |
| NICE | ONE_SIDED | 0/8 (0.0%) | denominator: VALUE/VALUE pairs (descriptive) |
| NICE | NOT_APPLICABLE | 0/8 (0.0%) | denominator: VALUE/VALUE pairs (descriptive) |

**L3_VALUE_EXACT_TEXT_MATCH** — connective_as_stated

| Guideline | Measure | Value | Note |
|---|---|---|---|
| EGS | exact match | 22/39 (56.4%) | denominator: VALUE/VALUE pairs (conditional) |
| AAO | exact match | 25/34 (73.5%) | denominator: VALUE/VALUE pairs (conditional) |
| NICE | exact match | 23/36 (63.9%) | denominator: VALUE/VALUE pairs (conditional) |

**EVIDENCE_SPAN_CONCORDANCE** — L3_VALUE connective_as_stated

| Guideline | Measure | Value | Note |
|---|---|---|---|
| EGS | IDENTICAL | 22/39 (56.4%) | denominator: VALUE/VALUE pairs (descriptive) |
| EGS | OVERLAPPING | 15/39 (38.5%) | denominator: VALUE/VALUE pairs (descriptive) |
| EGS | DISJOINT | 2/39 (5.1%) | denominator: VALUE/VALUE pairs (descriptive) |
| EGS | ONE_SIDED | 0/39 (0.0%) | denominator: VALUE/VALUE pairs (descriptive) |
| EGS | NOT_APPLICABLE | 0/39 (0.0%) | denominator: VALUE/VALUE pairs (descriptive) |
| AAO | IDENTICAL | 22/34 (64.7%) | denominator: VALUE/VALUE pairs (descriptive) |
| AAO | OVERLAPPING | 7/34 (20.6%) | denominator: VALUE/VALUE pairs (descriptive) |
| AAO | DISJOINT | 5/34 (14.7%) | denominator: VALUE/VALUE pairs (descriptive) |
| AAO | ONE_SIDED | 0/34 (0.0%) | denominator: VALUE/VALUE pairs (descriptive) |
| AAO | NOT_APPLICABLE | 0/34 (0.0%) | denominator: VALUE/VALUE pairs (descriptive) |
| NICE | IDENTICAL | 21/36 (58.3%) | denominator: VALUE/VALUE pairs (descriptive) |
| NICE | OVERLAPPING | 12/36 (33.3%) | denominator: VALUE/VALUE pairs (descriptive) |
| NICE | DISJOINT | 3/36 (8.3%) | denominator: VALUE/VALUE pairs (descriptive) |
| NICE | ONE_SIDED | 0/36 (0.0%) | denominator: VALUE/VALUE pairs (descriptive) |
| NICE | NOT_APPLICABLE | 0/36 (0.0%) | denominator: VALUE/VALUE pairs (descriptive) |

**L3_VALUE_EXACT_TEXT_MATCH** — cross_reference_as_stated

| Guideline | Measure | Value | Note |
|---|---|---|---|
| EGS | exact match | 3/5 (60.0%) | denominator: VALUE/VALUE pairs (conditional) |

| Guideline | Measure | Value | Note |
|---|---|---|---|
| AAO | exact match | 0/1 (0.0%) | denominator: VALUE/VALUE pairs (conditional) |
| NICE | exact match | 3/4 (75.0%) | denominator: VALUE/VALUE pairs (conditional) |

##### **EVIDENCE_SPAN_CONCORDANCE** — L3_VALUE cross_reference_as_stated

| Guideline | Measure | Value | Note |
|---|---|---|---|
| EGS | IDENTICAL | 3/5 (60.0%) | denominator: VALUE/VALUE pairs (descriptive) |
| EGS | OVERLAPPING | 2/5 (40.0%) | denominator: VALUE/VALUE pairs (descriptive) |
| EGS | DISJOINT | 0/5 (0.0%) | denominator: VALUE/VALUE pairs (descriptive) |
| EGS | ONE_SIDED | 0/5 (0.0%) | denominator: VALUE/VALUE pairs (descriptive) |
| EGS | NOT_APPLICABLE | 0/5 (0.0%) | denominator: VALUE/VALUE pairs (descriptive) |
| AAO | IDENTICAL | 0/1 (0.0%) | denominator: VALUE/VALUE pairs (descriptive) |
| AAO | OVERLAPPING | 1/1 (100.0%) | denominator: VALUE/VALUE pairs (descriptive) |
| AAO | DISJOINT | 0/1 (0.0%) | denominator: VALUE/VALUE pairs (descriptive) |
| AAO | ONE_SIDED | 0/1 (0.0%) | denominator: VALUE/VALUE pairs (descriptive) |
| AAO | NOT_APPLICABLE | 0/1 (0.0%) | denominator: VALUE/VALUE pairs (descriptive) |
| NICE | IDENTICAL | 3/4 (75.0%) | denominator: VALUE/VALUE pairs (descriptive) |
| NICE | OVERLAPPING | 1/4 (25.0%) | denominator: VALUE/VALUE pairs (descriptive) |
| NICE | DISJOINT | 0/4 (0.0%) | denominator: VALUE/VALUE pairs (descriptive) |
| NICE | ONE_SIDED | 0/4 (0.0%) | denominator: VALUE/VALUE pairs (descriptive) |
| NICE | NOT_APPLICABLE | 0/4 (0.0%) | denominator: VALUE/VALUE pairs (descriptive) |

##### **L3_VALUE_EXACT_TEXT_MATCH** — actor_expression_as_stated

| Guideline | Measure | Value | Note |
|---|---|---|---|
| EGS | exact match | 8/8 (100.0%) | denominator: VALUE/VALUE pairs (conditional) |
| AAO | exact match | 8/9 (88.9%) | denominator: VALUE/VALUE pairs (conditional) |
| NICE | exact match | 7/9 (77.8%) | denominator: VALUE/VALUE pairs (conditional) |

##### **EVIDENCE_SPAN_CONCORDANCE** — L3_VALUE actor_expression_as_stated

| Guideline | Measure | Value | Note |
|---|---|---|---|
| EGS | IDENTICAL | 8/8 (100.0%) | denominator: VALUE/VALUE pairs (descriptive) |
| EGS | OVERLAPPING | 0/8 (0.0%) | denominator: VALUE/VALUE pairs (descriptive) |
| EGS | DISJOINT | 0/8 (0.0%) | denominator: VALUE/VALUE pairs (descriptive) |
| EGS | ONE_SIDED | 0/8 (0.0%) | denominator: VALUE/VALUE pairs (descriptive) |
| EGS | NOT_APPLICABLE | 0/8 (0.0%) | denominator: VALUE/VALUE pairs (descriptive) |
| AAO | IDENTICAL | 8/9 (88.9%) | denominator: VALUE/VALUE pairs (descriptive) |
| AAO | OVERLAPPING | 1/9 (11.1%) | denominator: VALUE/VALUE pairs (descriptive) |
| AAO | DISJOINT | 0/9 (0.0%) | denominator: VALUE/VALUE pairs (descriptive) |
| AAO | ONE_SIDED | 0/9 (0.0%) | denominator: VALUE/VALUE pairs (descriptive) |
| AAO | NOT_APPLICABLE | 0/9 (0.0%) | denominator: VALUE/VALUE pairs (descriptive) |
| NICE | IDENTICAL | 7/9 (77.8%) | denominator: VALUE/VALUE pairs (descriptive) |
| NICE | OVERLAPPING | 2/9 (22.2%) | denominator: VALUE/VALUE pairs (descriptive) |
| NICE | DISJOINT | 0/9 (0.0%) | denominator: VALUE/VALUE pairs (descriptive) |
| NICE | ONE_SIDED | 0/9 (0.0%) | denominator: VALUE/VALUE pairs (descriptive) |
| NICE | NOT_APPLICABLE | 0/9 (0.0%) | denominator: VALUE/VALUE pairs (descriptive) |

#### **EVIDENCE_SPAN_CONCORDANCE** — PG06_OUTCOME SOURCE_SPECIFIED_DECISION_CONTENT_EVIDENCE

| Guideline | Measure | Value | Note |
|---|---|---|---|
| EGS | IDENTICAL | 10/83 (12.0%) | denominator: duplicated Class-A units (descriptive) |
| EGS | OVERLAPPING | 19/83 (22.9%) | denominator: duplicated Class-A units (descriptive) |
| EGS | DISJOINT | 1/83 (1.2%) | denominator: duplicated Class-A units (descriptive) |
| EGS | ONE_SIDED | 18/83 (21.7%) | denominator: duplicated Class-A units (descriptive) |
| EGS | NOT_APPLICABLE | 35/83 (42.2%) | denominator: duplicated Class-A units (descriptive) |
| AAO | IDENTICAL | 11/74 (14.9%) | denominator: duplicated Class-A units (descriptive) |
| AAO | OVERLAPPING | 17/74 (23.0%) | denominator: duplicated Class-A units (descriptive) |
| AAO | DISJOINT | 0/74 (0.0%) | denominator: duplicated Class-A units (descriptive) |
| AAO | ONE_SIDED | 24/74 (32.4%) | denominator: duplicated Class-A units (descriptive) |
| AAO | NOT_APPLICABLE | 22/74 (29.7%) | denominator: duplicated Class-A units (descriptive) |
| NICE | IDENTICAL | 6/60 (10.0%) | denominator: duplicated Class-A units (descriptive) |
| NICE | OVERLAPPING | 32/60 (53.3%) | denominator: duplicated Class-A units (descriptive) |
| NICE | DISJOINT | 0/60 (0.0%) | denominator: duplicated Class-A units (descriptive) |
| NICE | ONE_SIDED | 16/60 (26.7%) | denominator: duplicated Class-A units (descriptive) |
| NICE | NOT_APPLICABLE | 6/60 (10.0%) | denominator: duplicated Class-A units (descriptive) |

#### **EVIDENCE_SPAN_CONCORDANCE** — PG06_OUTCOME EXPLICIT_DECISION_AUTHORITY_ALLOCATION_EVIDENCE

| Guideline | Measure | Value | Note |
|---|---|---|---|
| EGS | IDENTICAL | 0/83 (0.0%) | denominator: duplicated Class-A units (descriptive) |
| EGS | OVERLAPPING | 0/83 (0.0%) | denominator: duplicated Class-A units (descriptive) |
| EGS | DISJOINT | 0/83 (0.0%) | denominator: duplicated Class-A units (descriptive) |
| EGS | ONE_SIDED | 1/83 (1.2%) | denominator: duplicated Class-A units (descriptive) |
| EGS | NOT_APPLICABLE | 82/83 (98.8%) | denominator: duplicated Class-A units (descriptive) |
| AAO | IDENTICAL | 0/74 (0.0%) | denominator: duplicated Class-A units (descriptive) |
| AAO | OVERLAPPING | 1/74 (1.4%) | denominator: duplicated Class-A units (descriptive) |
| AAO | DISJOINT | 0/74 (0.0%) | denominator: duplicated Class-A units (descriptive) |
| AAO | ONE_SIDED | 1/74 (1.4%) | denominator: duplicated Class-A units (descriptive) |
| AAO | NOT_APPLICABLE | 72/74 (97.3%) | denominator: duplicated Class-A units (descriptive) |
| NICE | IDENTICAL | 1/60 (1.7%) | denominator: duplicated Class-A units (descriptive) |
| NICE | OVERLAPPING | 2/60 (3.3%) | denominator: duplicated Class-A units (descriptive) |
| NICE | DISJOINT | 0/60 (0.0%) | denominator: duplicated Class-A units (descriptive) |
| NICE | ONE_SIDED | 1/60 (1.7%) | denominator: duplicated Class-A units (descriptive) |
| NICE | NOT_APPLICABLE | 56/60 (93.3%) | denominator: duplicated Class-A units (descriptive) |

#### **CLASSB_RELATION**

| Guideline | Measure | Value | Note |
|---|---|---|---|
| EGS | exact equality | 34/484 (7.0%) | denominator: dependency links of duplicated Class-B objects (descriptive) |
| AAO | exact equality | 9/99 (9.1%) | denominator: dependency links of duplicated Class-B objects (descriptive) |
| NICE | exact equality | 23/143 (16.1%) | denominator: dependency links of duplicated Class-B objects (descriptive) |

##### DEFECT_PRESENCE

| Guideline | Measure | Value | Note |
|---|---|---|---|
| EGS | presence agreement | 83/83 (100.0%) | denominator: duplicated Class-A units (descriptive) |
| EGS | contingency E1=FALSE / E2=FALSE | 83 | |
| EGS | contingency E1=FALSE / E2=TRUE | 0 | |
| EGS | contingency E1=TRUE / E2=FALSE | 0 | |
| EGS | contingency E1=TRUE / E2=TRUE | 0 | |
| AAO | presence agreement | 70/74 (94.6%) | denominator: duplicated Class-A units (descriptive) |
| AAO | contingency E1=FALSE / E2=FALSE | 70 | |
| AAO | contingency E1=FALSE / E2=TRUE | 0 | |
| AAO | contingency E1=TRUE / E2=FALSE | 4 | |
| AAO | contingency E1=TRUE / E2=TRUE | 0 | |
| NICE | presence agreement | 59/60 (98.3%) | denominator: duplicated Class-A units (descriptive) |
| NICE | contingency E1=FALSE / E2=FALSE | 59 | |
| NICE | contingency E1=FALSE / E2=TRUE | 0 | |
| NICE | contingency E1=TRUE / E2=FALSE | 1 | |
| NICE | contingency E1=TRUE / E2=TRUE | 0 | |

##### PG11_PHASE2_DESCRIPTORS

| Guideline | Measure | Value | Note |
|---|---|---|---|
| EGS | NOT_APPLICABLE | 140/140 (100.0%) | denominator: duplicated units; paired comparison fails closed (OP-4) |
| EGS | NOT_PAIRED | 0/140 (0.0%) | denominator: duplicated units; paired comparison fails closed (OP-4) |
| AAO | NOT_APPLICABLE | 90/90 (100.0%) | denominator: duplicated units; paired comparison fails closed (OP-4) |
| AAO | NOT_PAIRED | 0/90 (0.0%) | denominator: duplicated units; paired comparison fails closed (OP-4) |
| NICE | NOT_APPLICABLE | 92/92 (100.0%) | denominator: duplicated units; paired comparison fails closed (OP-4) |
| NICE | NOT_PAIRED | 0/92 (0.0%) | denominator: duplicated units; paired comparison fails closed (OP-4) |

##### **PHASE2_REVISION_EVENTS** — E1

| Guideline | Measure | Value | Note |
|---|---|---|---|
| EGS | STAGE1 EVENT_RECORD_REVISED | 0 | count; does not alter any Phase-I numerator (PG-15 sec. 11.2) |
| EGS | READING_CANDIDATE PROMOTION | 0 | count; does not alter any Phase-I numerator (PG-15 sec. 11.2) |
| EGS | READING_CANDIDATE DEMOTION | 0 | count; does not alter any Phase-I numerator (PG-15 sec. 11.2) |
| EGS | READING_CANDIDATE GOVERNANCE_USE_CHANGE | 0 | count; does not alter any Phase-I numerator (PG-15 sec. 11.2) |
| EGS | F1_F5 SUPERSEDING ASSESSMENT | 0 | count; does not alter any Phase-I numerator (PG-15 sec. 11.2) |
| AAO | STAGE1 EVENT_RECORD_REVISED | 0 | count; does not alter any Phase-I numerator (PG-15 sec. 11.2) |
| AAO | READING_CANDIDATE PROMOTION | 0 | count; does not alter any Phase-I numerator (PG-15 sec. 11.2) |
| AAO | READING_CANDIDATE DEMOTION | 0 | count; does not alter any Phase-I numerator (PG-15 sec. 11.2) |
| AAO | READING_CANDIDATE GOVERNANCE_USE_CHANGE | 0 | count; does not alter any Phase-I numerator (PG-15 sec. 11.2) |
| AAO | F1_F5 SUPERSEDING ASSESSMENT | 0 | count; does not alter any Phase-I numerator (PG-15 sec. 11.2) |

| Guideline | Measure | Value | Note |
|---|---|---|---|
| NICE | STAGE1 EVENT_RECORD_REVISED | 0 | count; does not alter any Phase-I numerator (PG-15 sec. 11.2) |
| NICE | READING_CANDIDATE PROMOTION | 0 | count; does not alter any Phase-I numerator (PG-15 sec. 11.2) |
| NICE | READING_CANDIDATE DEMOTION | 0 | count; does not alter any Phase-I numerator (PG-15 sec. 11.2) |
| NICE | READING_CANDIDATE GOVERNANCE_USE_CHANGE | 0 | count; does not alter any Phase-I numerator (PG-15 sec. 11.2) |
| NICE | F1_F5 SUPERSEDING ASSESSMENT | 0 | count; does not alter any Phase-I numerator (PG-15 sec. 11.2) |

**PHASE2_REVISION_EVENTS** — E2

| Guideline | Measure | Value | Note |
|---|---|---|---|
| EGS | STAGE1 EVENT_RECORD_REVISED | 0 | count; does not alter any Phase-I numerator (PG-15 sec. 11.2) |
| EGS | READING_CANDIDATE PROMOTION | 0 | count; does not alter any Phase-I numerator (PG-15 sec. 11.2) |
| EGS | READING_CANDIDATE DEMOTION | 0 | count; does not alter any Phase-I numerator (PG-15 sec. 11.2) |
| EGS | READING_CANDIDATE GOVERNANCE_USE_CHANGE | 0 | count; does not alter any Phase-I numerator (PG-15 sec. 11.2) |
| EGS | F1_F5 SUPERSEDING ASSESSMENT | 0 | count; does not alter any Phase-I numerator (PG-15 sec. 11.2) |
| AAO | STAGE1 EVENT_RECORD_REVISED | 0 | count; does not alter any Phase-I numerator (PG-15 sec. 11.2) |
| AAO | READING_CANDIDATE PROMOTION | 0 | count; does not alter any Phase-I numerator (PG-15 sec. 11.2) |
| AAO | READING_CANDIDATE DEMOTION | 0 | count; does not alter any Phase-I numerator (PG-15 sec. 11.2) |
| AAO | READING_CANDIDATE GOVERNANCE_USE_CHANGE | 0 | count; does not alter any Phase-I numerator (PG-15 sec. 11.2) |
| AAO | F1_F5 SUPERSEDING ASSESSMENT | 0 | count; does not alter any Phase-I numerator (PG-15 sec. 11.2) |
| NICE | STAGE1 EVENT_RECORD_REVISED | 0 | count; does not alter any Phase-I numerator (PG-15 sec. 11.2) |
| NICE | READING_CANDIDATE PROMOTION | 0 | count; does not alter any Phase-I numerator (PG-15 sec. 11.2) |
| NICE | READING_CANDIDATE DEMOTION | 0 | count; does not alter any Phase-I numerator (PG-15 sec. 11.2) |
| NICE | READING_CANDIDATE GOVERNANCE_USE_CHANGE | 0 | count; does not alter any Phase-I numerator (PG-15 sec. 11.2) |
| NICE | F1_F5 SUPERSEDING ASSESSMENT | 0 | count; does not alter any Phase-I numerator (PG-15 sec. 11.2) |

#### Table S3-6. Overlap judgements C-11, descriptive context (C11_OVERLAP.csv)

Key to the 12 items answered for each retained object (instrument wording abridged): SPAN, does the anchored source span correspond to the object? L23, does the semantic record (segmented text, L2, and structured fields, L3) state what the source says, no more and no less? PG06, are the two recorded authority dimensions (source-specified decision content; explicit decision-authority allocation) supported by the source? PG10-Q1 to Q8, admissibility checks: Q1 transformation class assigned correctly; Q2 source-derived status justified by a valid predeclared rule; Q3 semantic losslessness supported; Q4 choice space genuinely bounded by the source; Q5 study-level selection provenance visible; Q6 an author-defined assumption presented as source-supported; Q7 an inadmissible invention entering a representation; Q8 pending dependencies improperly bypassed. EXEC, does the executable rendering state what the source says, no more and no less?

C-11 R-B overlap judgments and answer differences - NOT AN OUTCOME, descriptive context (P23-17).

Both reviewers' fields as recorded; an answer difference is not an overlap divergence (PG18-A1-11); no agreement statistic.

| body | object_id | item_id | item | RA item_outcome | RA category | RA stance | RA materiality_ answer | RB item_outcome | RB category | RB stance | RB materiality_ answer | materiality answers |
|---|---|---|---|---|---|---|---|---|---|---|---|---|
| NICE | NICE-CD-0032-C01 | RA-NICE-CD-0032-C01-SPAN | SPAN | NO-FINDING-RAISED | | | | NO-FINDING-RAISED | | | | BOTH NO FINDING |
| NICE | NICE-CD-0032-C01 | RA-NICE-CD-0032-C01-L23 | L23 | NO-FINDING-RAISED | | | | NO-FINDING-RAISED | | | | BOTH NO FINDING |

| body | object_id | item_id | item | RA item_outcome | RA category | RA stance | RA materiality_answer | RB item_outcome | RB category | RB stance | RB materiality_answer | materiality answers |
|---|---|---|---|---|---|---|---|---|---|---|---|---|
| NICE | NICE-CD-0032-C01 | RA-NICE-CD-0032-C01-PG06 | PG06 | NO-FINDING-RAISED | | | | NO-FINDING-RAISED | | | | BOTH NO FINDING |
| NICE | NICE-CD-0032-C01 | RA-NICE-CD-0032-C01-PG10-Q1 | PG10-Q1 | NO-FINDING-RAISED | | | | NO-FINDING-RAISED | | | | BOTH NO FINDING |
| NICE | NICE-CD-0032-C01 | RA-NICE-CD-0032-C01-PG10-Q2 | PG10-Q2 | NO-FINDING-RAISED | | | | NO-FINDING-RAISED | | | | BOTH NO FINDING |
| NICE | NICE-CD-0032-C01 | RA-NICE-CD-0032-C01-PG10-Q3 | PG10-Q3 | NO-FINDING-RAISED | | | | NO-FINDING-RAISED | | | | BOTH NO FINDING |
| NICE | NICE-CD-0032-C01 | RA-NICE-CD-0032-C01-PG10-Q4 | PG10-Q4 | NO-FINDING-RAISED | | | | NO-FINDING-RAISED | | | | BOTH NO FINDING |
| NICE | NICE-CD-0032-C01 | RA-NICE-CD-0032-C01-PG10-Q5 | PG10-Q5 | NO-FINDING-RAISED | | | | NO-FINDING-RAISED | | | | BOTH NO FINDING |
| NICE | NICE-CD-0032-C01 | RA-NICE-CD-0032-C01-PG10-Q6 | PG10-Q6 | NO-FINDING-RAISED | | | | NO-FINDING-RAISED | | | | BOTH NO FINDING |
| NICE | NICE-CD-0032-C01 | RA-NICE-CD-0032-C01-PG10-Q7 | PG10-Q7 | NO-FINDING-RAISED | | | | NO-FINDING-RAISED | | | | BOTH NO FINDING |
| NICE | NICE-CD-0032-C01 | RA-NICE-CD-0032-C01-PG10-Q8 | PG10-Q8 | NO-FINDING-RAISED | | | | NO-FINDING-RAISED | | | | BOTH NO FINDING |
| NICE | NICE-CD-0032-C01 | RA-NICE-CD-0032-C01-EXEC | EXEC | NO-FINDING-RAISED | | | | FINDING-RAISED | RF-MISSING | ASSERTED | MATERIAL | FINDING ON ONE SIDE ONLY |
| AAO | AAO-CD-0009-C06 | RA-AAO-CD-0009-C06-SPAN | SPAN | NO-FINDING-RAISED | | | | NO-FINDING-RAISED | | | | BOTH NO FINDING |
| AAO | AAO-CD-0009-C06 | RA-AAO-CD-0009-C06-L23 | L23 | NO-FINDING-RAISED | | | | NO-FINDING-RAISED | | | | BOTH NO FINDING |
| AAO | AAO-CD-0009-C06 | RA-AAO-CD-0009-C06-PG06 | PG06 | NO-FINDING-RAISED | | | | NO-FINDING-RAISED | | | | BOTH NO FINDING |
| AAO | AAO-CD-0009-C06 | RA-AAO-CD-0009-C06-PG10-Q1 | PG10-Q1 | NO-FINDING-RAISED | | | | NO-FINDING-RAISED | | | | BOTH NO FINDING |
| AAO | AAO-CD-0009-C06 | RA-AAO-CD-0009-C06-PG10-Q2 | PG10-Q2 | NO-FINDING-RAISED | | | | NO-FINDING-RAISED | | | | BOTH NO FINDING |
| AAO | AAO-CD-0009-C06 | RA-AAO-CD-0009-C06-PG10-Q3 | PG10-Q3 | NO-FINDING-RAISED | | | | NO-FINDING-RAISED | | | | BOTH NO FINDING |
| AAO | AAO-CD-0009-C06 | RA-AAO-CD-0009-C06-PG10-Q4 | PG10-Q4 | NO-FINDING-RAISED | | | | NO-FINDING-RAISED | | | | BOTH NO FINDING |
| AAO | AAO-CD-0009-C06 | RA-AAO-CD-0009-C06-PG10-Q5 | PG10-Q5 | NO-FINDING-RAISED | | | | NO-FINDING-RAISED | | | | BOTH NO FINDING |
| AAO | AAO-CD-0009-C06 | RA-AAO-CD-0009-C06-PG10-Q6 | PG10-Q6 | NO-FINDING-RAISED | | | | NO-FINDING-RAISED | | | | BOTH NO FINDING |
| AAO | AAO-CD-0009-C06 | RA-AAO-CD-0009-C06-PG10-Q7 | PG10-Q7 | NO-FINDING-RAISED | | | | NO-FINDING-RAISED | | | | BOTH NO FINDING |
| AAO | AAO-CD-0009-C06 | RA-AAO-CD-0009-C06-PG10-Q8 | PG10-Q8 | NO-FINDING-RAISED | | | | NO-FINDING-RAISED | | | | BOTH NO FINDING |
| AAO | AAO-CD-0009-C06 | RA-AAO-CD-0009-C06-EXEC | EXEC | NO-FINDING-RAISED | | | | NO-FINDING-RAISED | | | | BOTH NO FINDING |
| AAO | AAO-CD-0036-C02 | RA-AAO-CD-0036-C02-SPAN | SPAN | NO-FINDING-RAISED | | | | NO-FINDING-RAISED | | | | BOTH NO FINDING |
| AAO | AAO-CD-0036-C02 | RA-AAO-CD-0036-C02-L23 | L23 | NO-FINDING-RAISED | | | | NO-FINDING-RAISED | | | | BOTH NO FINDING |
| AAO | AAO-CD-0036-C02 | RA-AAO-CD-0036-C02-PG06 | PG06 | NO-FINDING-RAISED | | | | NO-FINDING-RAISED | | | | BOTH NO FINDING |
| AAO | AAO-CD-0036-C02 | RA-AAO-CD-0036-C02-PG10-Q1 | PG10-Q1 | NO-FINDING-RAISED | | | | NO-FINDING-RAISED | | | | BOTH NO FINDING |
| AAO | AAO-CD-0036-C02 | RA-AAO-CD-0036-C02-PG10-Q2 | PG10-Q2 | NO-FINDING-RAISED | | | | NO-FINDING-RAISED | | | | BOTH NO FINDING |
| AAO | AAO-CD-0036-C02 | RA-AAO-CD-0036-C02-PG10-Q3 | PG10-Q3 | NO-FINDING-RAISED | | | | NO-FINDING-RAISED | | | | BOTH NO FINDING |

| body | object_id | item_id | item | RA item_outcome | RA category | RA stance | RA materiality_ answer | RB item_outcome | RB category | RB stance | RB materiality_ answer | materiality answers |
|---|---|---|---|---|---|---|---|---|---|---|---|---|
| AAO | AAO-CD-0036-C02 | RA-AAO-CD-0036-C02-PG10-Q4 | PG10-Q4 | NO-FINDING-RAISED | | | | NO-FINDING-RAISED | | | | BOTH NO FINDING |
| AAO | AAO-CD-0036-C02 | RA-AAO-CD-0036-C02-PG10-Q5 | PG10-Q5 | NO-FINDING-RAISED | | | | NO-FINDING-RAISED | | | | BOTH NO FINDING |
| AAO | AAO-CD-0036-C02 | RA-AAO-CD-0036-C02-PG10-Q6 | PG10-Q6 | NO-FINDING-RAISED | | | | NO-FINDING-RAISED | | | | BOTH NO FINDING |
| AAO | AAO-CD-0036-C02 | RA-AAO-CD-0036-C02-PG10-Q7 | PG10-Q7 | NO-FINDING-RAISED | | | | NO-FINDING-RAISED | | | | BOTH NO FINDING |
| AAO | AAO-CD-0036-C02 | RA-AAO-CD-0036-C02-PG10-Q8 | PG10-Q8 | NO-FINDING-RAISED | | | | NO-FINDING-RAISED | | | | BOTH NO FINDING |
| AAO | AAO-CD-0036-C02 | RA-AAO-CD-0036-C02-EXEC | EXEC | NO-FINDING-RAISED | | | | NO-FINDING-RAISED | | | | BOTH NO FINDING |

Finding text, locators and importance, for every item with a finding (verbatim):

**RA-NICE-CD-0032-C01-EXEC — Reviewer B** (RF-MISSING; ASSERTED; MATERIAL)

Finding: The executable output reduces the recommendation to generic initiation of IOP-lowering treatment and does not carry forward the anchored intervention, 360° selective laser trabeculoplasty (SLT).

Locators: SOURCE_NICE.pdf, PDF sheet 9, recommendation 1.4.4; RA_OBJECTS_VIEW.html, executable rendering RV-NICE-V1, output mapping E-0032C01-OM.

Importance (optional, own words): The source-specified treatment is absent from the executable output.

#### Table S3-7. Structural-check detail and out-of-frame observation C-12 (C12_SEC_OBSERVATIONS.csv)

C-12 SEC observation detail; out-of-frame observations - NOT AN OUTCOME, descriptive context (P23-17). Fields as recorded.

| record | body | item_id | item_outcome | category | stance | materiality_answer |
|---|---|---|---|---|---|---|
| SEC_ITEM | EGS | SEC-EGS | NO-FINDING-RAISED | | | |
| SEC_ITEM | AAO | SEC-AAO | NO-FINDING-RAISED | | | |
| SEC_ITEM | NICE | SEC-NICE | FINDING-RAISED | RF-NOT-CLASSIFIED | ASSERTED | NOT-MATERIAL |
| OUT_OF_FRAME_OBSERVATION | NICE | obs 1 | | | | |
| PG03_S19_PARTITION_FAILURE_ACT | | PF-NICE-0001_PG03_S19_FINAL_ACT.md | sha256 cc60fac462601212bd992e7179eca613beecadce182a7235364fcd7ff77ed320 | | | |

**SEC-NICE (SEC_ITEM)**

Finding (own words, verbatim): The enumeration leaves PDF sheet 34 uncovered. That sheet contains the continuation text “procedure.” and the link “Return to recommendations 1.6.6 and 1.6.7”.

Locators: SOURCE_NICE.pdf, PDF sheet 34; RB_1_SEC_STRUCTURE_CHECK.xlsx, ENUMERATION (no NICE division spans sheet 34).

Importance (optional, own words): —

**obs 1 (OUT_OF_FRAME_OBSERVATION)**

Finding (own words, verbatim): PDF sheet 34 is not covered by any enumerated division; it contains the continuation “procedure.” and the link “Return to recommendations 1.6.6 and 1.6.7”.

Locators: SOURCE_NICE.pdf, PDF sheet 34.

Importance (optional, own words): —

**Component (c) finding, EGS-CD-0151 (sealed O-2 record DSP-RB-O2-001; disposition record, 29 October 2026)**

Division: EGS Part II > II.1 Patient Examination > II.1.4 Perimetry > II.1.4.2 Interpreting test results > II.1.4.2.6 Detecting and quantifying glaucomatous visual field deterioration (PDF sheets 107–109)

Finding (verbatim): The division view omits the source's explicit OHT-specific visual-field follow-up statement: patients with OHT do not need frequent VF testing; once every one or two years is often sufficient.

Category / stance / materiality: RF-POSSIBLE-OMISSION / ASSERTED / MATERIAL

Locators (verbatim): SOURCE_EGS.pdf, PDF sheet 109, passage beginning 'Most often, the frequency of testing can then be reduced' and continuing through 'once every one or two years is often sufficient.'; VIEWS_EGS.html#EGS-CD-0151.

Disposition and reason (translated from Italian): DSP-NO-CHANGE. The content reported as omitted is enumerated and represented in the division: record EGS-CD-0151-C35 keeps the OHT population, the negation of frequent testing and the one-to-two-year interval (L2 and L3 proposition content). No change is needed; the finding stays on record.

#### Table S3-8. PG-12 §17 reporting groups C-13 (C13_PG12_GROUPS.csv); groups are non-additive

Key: groups 1–4 classify transfers of a source statement beyond what it states (admitted, prohibited, barred, pending); none occurred. Groups 5 and 6 list non-identified axes: for each listed axis (care setting, terminology system, measurement convention) no qualifying source expression was identified within the assessment universe; these are procedural disclosures, not findings. Case A and Case B are the two attribution routes of the protocol; there was no Case B entry. Group 7 lists governance-incomplete pairings (none). Group counts are never summed.

C-13 PG-12 §17 seven reporting groups - NOT AN OUTCOME, reporting obligation (P23-17).

PG-12 §17.2: the seven groups report along two dimensions (Case A / Case B: attribution; governance-incomplete: pairing-level blocker); a NOT_GRANTED Case-B pairing appears in both; the group counts MUST NEVER be summed.

| group | pairing_ref | item | status_or_count | detail | flag |
|---|---|---|---|---|---|
| 1 ADMITTED TRANSFERS | | | EMPTY BY CONSTRUCTION | EMPTY BY CONSTRUCTION for genuine transfers: the resolved architecture family (Family A) permits no execution ceiling above NOT_EXECUTION_ELIGIBLE (PG-12 §17.3). Not 'none found'. | |
| 2 PROHIBITED TRANSFERS | | | 0 record(s) | intrinsic prohibition_ground; a statement about what the source says | |
| 3 BARRED TRANSFERS | | | 0 record(s), individually listed | ARCHITECTURE_FAMILY_CEILING-only subset identifiable (PG-12 §17.4); not executed, not a stratum, not pooled | |
| 4 PENDING TRANSFERS | | | EMPTY | EMPTY for genuine transfers under Family A (TRANSFER_PENDING unreachable, PG-12 §13.2); pre-determination dependencies are reported under group 7. | |
| 5 NON-IDENTIFIED AXES - CASE A | | | 5 axis entr(ies) | mandatory procedural disclosure, PG-07 §22 canonical form (PG-12 §18.2 row 6) | |
| 5 NON-IDENTIFIED AXES - CASE A | PAIR-RV-AAO-V1-TSC-PG02-S8 | CARE_SETTING | SNA-AAO-RV-AAO-V1-CARE_SETTING | No qualifying expression for target [TGT-AX-CARE-SETTING @ version 1] - a constraint on [CARE_SETTING] - was identified within [DOCUMENT AAO] over assessment universe [AU-AAO-RV-AAO-V1] under assessment pass [AP-S8-G2-01]. | |
| 5 NON-IDENTIFIED AXES - CASE A | PAIR-RV-AAO-V1-TSC-PG02-S8 | TERMINOLOGY_SYSTEM | SNA-AAO-RV-AAO-V1-TERMINOLOGY_SYSTEM | No qualifying expression for target [TGT-AX-TERMINOLOGY @ version 1] - a constraint on [TERMINOLOGY_SYSTEM] - was identified within [DOCUMENT AAO] over assessment universe [AU-AAO-RV-AAO-V1] under assessment pass [AP-S8-G2-01]. | |
| 5 NON-IDENTIFIED AXES - CASE A | PAIR-RV-AAO-V1-TSC-PG02-S8 | MEASUREMENT_CONVENTION | SNA-AAO-RV-AAO-V1-MEASUREMENT_CONVENTION | No qualifying expression for target [TGT-AX-MEASUREMENT @ version 1] - a constraint on [MEASUREMENT_CONVENTION] - was identified within [DOCUMENT AAO] over assessment universe [AU-AAO-RV-AAO-V1] under assessment pass [AP-S8-G2-01]. | |
| 5 NON-IDENTIFIED AXES - CASE A | PAIR-RV-NICE-V1-TSC-PG02-S8 | CARE_SETTING | SNA-NICE-0032C01-CARE_SETTING | No qualifying expression for target [TGT-AX-CARE-SETTING @ version 1] - a constraint on [CARE_SETTING] - was identified within [UNIT NICE-CD-0032-C01] over assessment universe [AU-NICE-0032C01] under assessment pass [AP-S8-G2-01]. | |
| 5 NON-IDENTIFIED AXES - CASE A | PAIR-RV-NICE-V1-TSC-PG02-S8 | TERMINOLOGY_SYSTEM | SNA-NICE-0032C01-TERMINOLOGY_SYSTEM | No qualifying expression for target [TGT-AX-TERMINOLOGY @ version 1] - a constraint on [TERMINOLOGY_SYSTEM] - was identified within [UNIT NICE-CD-0032-C01] over assessment universe [AU-NICE-0032C01] under assessment pass [AP-S8-G2-01]. | |
| 6 NON-IDENTIFIED AXES - CASE B | | | 0 axis entr(ies) | all Case-B pairings, granted and not granted | |
| 7 GOVERNANCE-INCOMPLETE PAIRINGS | | | 0 pairing(s) | pairing-level blockers, as derived (PG-12 §15.2) | |

#### Table S3-9. Examples of documented exclusions on which Reviewer B answered material (selected by Rule C)

Selection rule, fixed and recorded before any example was extracted (31 October 2026; a presentation decision after the analysis): per guideline, the three exclusions in class MATERIAL with the lowest SHA-256("OFTYGR-EXAMPLES-C" ‖ unit_id); no substitution. The examples carry no denominator and are not representative. Reviewer finding text is reproduced verbatim from the sealed X-1 workbook (SHA-256 1e641ffd…13fc41), including PDF-extraction artefacts. Per the reviewer's clarifying addendum (S1.6), the finding text used a standard formula, and *material* refers to the decision content the source states in the passage, not to whether the exclusion was proper for the study's domain. Disposition reasons are verbatim (Italian), with the authors' English translation.

#### Example 1 — EGS-CD-0055-C07

| Field | Content |
|---|---|
| Source locator | Part I > I.5 What matters to patients > I.5.3 Tips for doctors; PDF sheet 36; top 295.6: begins '4) Explain clearly how and when to administer eye drops.' … ends '4) Explain clearly how and when to administer eye drops.' (to sheet 36 y295.6) |
| Extraction-stage exclusion rationale (PG-04, as recorded) | Information on ongoing medical treatment and drop administration for patients on treatment (procedural detail, H-5) - not the initial decision. / limb (ii): No: removing it would not change the meaning, applicability, scope or interpretation of any named limb-(i)-YES root; proximity alone is not sufficient. |
| Reviewer B finding (verbatim) | Excluding this passage omits decision-relevant source content concerning Part I > I.5 What matters to patients > I.5.3 Tips for doctors: "4) Explain clearly how and when to administer eye drops. r e" |
| Category / stance / materiality | RF-EXCLUSION-QUESTIONED / ASSERTED / MATERIAL |
| Disposition and reason | DSP-NO-CHANGE. Verbatim: «Valutato il contenuto puntuale di questo item, il passo è fuori dal dominio dichiarato oppure non condiziona le unità in scope: Information on ongoing medical treatment and drop administration for patients on treatment (procedural detail, H-5) - not the initial decision. L'esclusione PG-04 resta invariata; il finding resta registrato senza riclassificazioni.» Translation: "Having assessed the specific content of this item, the passage is outside the declared domain or does not condition the in-scope units: Information on ongoing medical treatment and drop administration for patients on treatment (procedural detail, H-5) - not the initial decision. The PG-04 exclusion stands; the finding remains on record without reclassification." |

#### Example 2 — EGS-CD-0199-C07

| Field | Content |
|---|---|
| Source locator | Part II > II.2 Classification and terminology > II.2.4 Angle closure > II.2.4.1 Primary angle closure glaucoma > II.2.4.1.4 Acute angle closure (AAC) > Systemic medication; PDF sheet 147; top 226.9: begins 'Mannitol lowers IOP through its hyperosmotic effects' … ends 'leading to dehydration of the vitreous body.' (to sheet 147 y238.3) |
| Extraction-stage exclusion rationale (PG-04, as recorded) | Acute angle closure: explicitly excluded high-level category (acute angle-closure). / limb (ii): No named limb (i) YES root whose meaning, applicability, scope or interpretation would change if this were removed. |
| Reviewer B finding (verbatim) | Excluding this passage omits decision-relevant source content concerning Part II > II.2 Classification and terminology > II.2.4 Angle closure > II.2.4.1 Primary angle closure glaucoma > II.2.4.1.4 Acute angle closure (AAC) > Systemic medication: "azolamide. Mannitol lowers IOP through its hyperosmotic effects leading to dehydration of the vitreous body. Patients must be evaluated for heart or kidney disease because hyperosmotics clud pht" |
| Category / stance / materiality | RF-EXCLUSION-QUESTIONED / ASSERTED / MATERIAL |
| Disposition and reason | DSP-NO-CHANGE. Verbatim: «Valutato il contenuto puntuale di questo item, il passo è fuori dal dominio dichiarato oppure non condiziona le unità in scope: Acute angle closure: explicitly excluded high-level category (acute angle-closure). L'esclusione PG-04 resta invariata; il finding resta registrato senza riclassificazioni.» Translation: "Having assessed the specific content of this item, the passage is outside the declared domain or does not condition the in-scope units: Acute angle closure: explicitly excluded high-level category (acute angle-closure). The PG-04 exclusion stands; the finding remains on record without reclassification." |

#### Example 3 — EGS-CD-0189-C05

| Field | Content |
|---|---|
| Source locator | Part II > II.2 Classification and terminology > II.2.4 Angle closure; PDF sheet 138; top 182.7: begins 'Anterior segment OCT (AS-OCT) may be used' … ends 'but it cannot replace gonioscopy for diagnosis.' (to sheet 138 y194.1) |
| Extraction-stage exclusion rationale (PG-04, as recorded) | Angle-closure definition, mechanism or diagnosis: specific to the angle-closure context, outside the declared OHT population and decision. / limb (ii): No named limb (i) YES root whose meaning, applicability, scope or interpretation would change if this were removed. |
| Reviewer B finding (verbatim) | Excluding this passage omits decision-relevant source content concerning Part II > II.2 Classification and terminology > II.2.4 Angle closure: "closure. Anterior segment OCT (AS-OCT) may be used as a supplementary test, e.g. for tri- age, but it cannot replace gonioscopy for diagnosis. AS-OCT tends to identify more ITC than pyri 10.11" |
| Category / stance / materiality | RF-EXCLUSION-QUESTIONED / ASSERTED / MATERIAL |
| Disposition and reason | DSP-NO-CHANGE. Verbatim: «Valutato il contenuto puntuale di questo item, il passo è fuori dal dominio dichiarato oppure non condiziona le unità in scope: Angle-closure definition, mechanism or diagnosis: specific to the angle-closure context, outside the declared OHT population and decision. L'esclusione PG-04 resta invariata; il finding resta registrato senza riclassificazioni.» Translation: "Having assessed the specific content of this item, the passage is outside the declared domain or does not condition the in-scope units: Angle-closure definition, mechanism or diagnosis: specific to the angle-closure context, outside the declared OHT population and decision. The PG-04 exclusion stands; the finding remains on record without reclassification." |

#### Example 4 — AAO-CD-0041-C54

| Field | Content |
|---|---|
| Source locator | CARE PROCESS > MANAGEMENT > Choice of Therapy > Medical treatment; PDF sheet 27; top 298.5: begins 'In 2024, a travoprost-eluting implant' … ends 'non-inferiority to topical timolol.' (to sheet 27 y333.0) |
| Extraction-stage exclusion rationale (PG-04, as recorded) | Surgically implanted drug delivery; outside domain. / limb (ii): Not needed to preserve the meaning, applicability, scope or interpretation of any named in-scope root unit. |
| Reviewer B finding (verbatim) | Excluding this passage omits decision-relevant source content concerning CARE PROCESS > MANAGEMENT > Choice of Therapy > Medical treatment: "of corneal endothelial cell loss. In 2024, a travoprost-eluting implant (iDose TR, Glaukos Corporation, Aliso Viejo, CA) was FDA approved for surgical implantation into the trabecular meshwork based on two prospective, randomized, multicenter, double-masked phase 3 trials that showed non-inferiority to topical timolol. At 3 years, a greater" |
| Category / stance / materiality | RF-EXCLUSION-QUESTIONED / ASSERTED / MATERIAL |
| Disposition and reason | DSP-NO-CHANGE. Verbatim: «Valutato il contenuto puntuale di questo item, il passo è fuori dal dominio dichiarato oppure non condiziona le unità in scope: Surgically implanted drug delivery; outside domain. L'esclusione PG-04 resta invariata; il finding resta registrato senza riclassificazioni.» Translation: "Having assessed the specific content of this item, the passage is outside the declared domain or does not condition the in-scope units: Surgically implanted drug delivery; outside domain. The PG-04 exclusion stands; the finding remains on record without reclassification." |

#### Example 5 — AAO-CD-0038-C01

| Field | Content |
|---|---|
| Source locator | CARE PROCESS > MANAGEMENT > Target Intraocular Pressure; PDF sheet 21; top 473.7: begins 'When deciding to treat a glaucoma suspect,' … ends 'over his or her lifetime.' (to sheet 21 y496.9) |
| Extraction-stage exclusion rationale (PG-04, as recorded) | Goal range of treatment underlying target IOP; outside the PG-02 domain (target IOP). / limb (ii): Not needed to preserve the meaning, applicability, scope or interpretation of any named in-scope root unit. |
| Reviewer B finding (verbatim) | Excluding this passage omits decision-relevant source content concerning CARE PROCESS > MANAGEMENT > Target Intraocular Pressure: "When deciding to treat a glaucoma suspect, it is important to remember that the goal of treatment is to maintain the IOP in a range at which visual field loss is unlikely to significantly affect a patient's health-related quality of life over his or her lifetime.214 The estimated upper" |
| Category / stance / materiality | RF-EXCLUSION-QUESTIONED / ASSERTED / MATERIAL |
| Disposition and reason | DSP-NO-CHANGE. Verbatim: «Valutato il contenuto puntuale di questo item, il passo è fuori dal dominio dichiarato oppure non condiziona le unità in scope: Goal range of treatment underlying target IOP; outside the PG-02 domain (target IOP). L'esclusione PG-04 resta invariata; il finding resta registrato senza riclassificazioni.» Translation: "Having assessed the specific content of this item, the passage is outside the declared domain or does not condition the in-scope units: Goal range of treatment underlying target IOP; outside the PG-02 domain (target IOP). The PG-04 exclusion stands; the finding remains on record without reclassification." |

#### Example 6 — AAO-CD-0041-C03

| Field | Content |
|---|---|
| Source locator | CARE PROCESS > MANAGEMENT > Choice of Therapy > Medical treatment; PDF sheet 22; top 332.3: begins 'Therefore, prostaglandin analogues are often considered' … ends 'refusal preclude this.' (to sheet 22 y355.4) |
| Extraction-stage exclusion rationale (PG-04, as recorded) | Choice of first medication once treating; outside domain. / limb (ii): Not needed to preserve the meaning, applicability, scope or interpretation of any named in-scope root unit. |
| Reviewer B finding (verbatim) | Excluding this passage omits decision-relevant source content concerning CARE PROCESS > MANAGEMENT > Choice of Therapy > Medical treatment: "Therefore, prostaglandin analogues are often considered as initial medical therapy unless other considerations such as contraindications, cost, side effects, intolerance, or patient refusal preclude this.220, 221" |
| Category / stance / materiality | RF-EXCLUSION-QUESTIONED / ASSERTED / MATERIAL |

| Field | Content |
|---|---|
| Disposition and reason | DSP-NO-CHANGE. Verbatim: «Valutato il contenuto puntuale di questo item, il passo è fuori dal dominio dichiarato oppure non condiziona le unità in scope: Choice of first medication once treating; outside domain. L'esclusione PG-04 resta invariata; il finding resta registrato senza riclassificazioni.» Translation: "Having assessed the specific content of this item, the passage is outside the declared domain or does not condition the in-scope units: Choice of first medication once treating; outside domain. The PG-04 exclusion stands; the finding remains on record without reclassification." |

#### Example 7 — NICE-CD-0046-C01

| Field | Content |
|---|---|
| Source locator | Glaucoma: diagnosis and management > Recommendations > 1.4 Treatment > Treatment for people with COAG > Treatment for people with advanced COAG > 1.4.14; PDF sheet 12; top 229.1: begins '1.4.14 Offer people with advanced COAG, glaucoma surgery with' … ends 'augmentation (MMC) as indicated.' (to sheet 12 y247.1) |
| Extraction-stage exclusion rationale (PG-04, as recorded) | Specific to treatment of established COAG (excluded category: established glaucoma). / limb (ii): No named in-scope root unit's meaning, applicability, scope or interpretation would change if this were removed. |
| Reviewer B finding (verbatim) | Excluding this passage omits decision-relevant source content concerning Glaucoma: diagnosis and management > Recommendations > 1.4 Treatment > Treatment for people with COAG > Treatment for people with advanced COAG > 1.4.14: "1.4.14 Offer people with advanced COAG, glaucoma surgery with pharmacological augmentation (MMC) as indicated. Give them information on the risks and" |
| Category / stance / materiality | RF-EXCLUSION-QUESTIONED / ASSERTED / MATERIAL |
| Disposition and reason | DSP-NO-CHANGE. Verbatim: «Valutato il contenuto puntuale di questo item, il passo è fuori dal dominio dichiarato oppure non condiziona le unità in scope: Specific to treatment of established COAG (excluded category: established glaucoma). L'esclusione PG-04 resta invariata; il finding resta registrato senza riclassificazioni.» Translation: "Having assessed the specific content of this item, the passage is outside the declared domain or does not condition the in-scope units: Specific to treatment of established COAG (excluded category: established glaucoma). The PG-04 exclusion stands; the finding remains on record without reclassification." |

#### Example 8 — NICE-CD-0093-C07

| Field | Content |
|---|---|
| Source locator | Glaucoma: diagnosis and management > Recommendations > 1.7 Providing information > 1.7.2; PDF sheet 25; top 233.7: begins 'compliance aids (such as dispensers) available from their GP' … ends 'from their GP or community pharmacist' (to sheet 25 y251.7) |
| Extraction-stage exclusion rationale (PG-04, as recorded) | Read with the stem: adherence aids for ongoing treatment. / limb (ii): No named in-scope root unit's meaning, applicability, scope or interpretation would change if this were removed. |
| Reviewer B finding (verbatim) | Excluding this passage omits decision-relevant source content concerning Glaucoma: diagnosis and management > Recommendations > 1.7 Providing information > 1.7.2: "• compliance aids (such as dispensers) available from their GP or community pharmacist" |
| Category / stance / materiality | RF-EXCLUSION-QUESTIONED / ASSERTED / MATERIAL |
| Disposition and reason | DSP-NO-CHANGE. Verbatim: «Valutato il contenuto puntuale di questo item, il passo è fuori dal dominio dichiarato oppure non condiziona le unità in scope: Read with the stem: adherence aids for ongoing treatment. L'esclusione PG-04 resta invariata; il finding resta registrato senza riclassificazioni.» Translation: "Having assessed the specific content of this item, the passage is outside the declared domain or does not condition the in-scope units: Read with the stem: adherence aids for ongoing treatment. The PG-04 exclusion stands; the finding remains on record without reclassification." |

#### Example 9 — NICE-CD-0074-C01

| Field | Content |
|---|---|
| Source locator | Glaucoma: diagnosis and management > Recommendations > 1.5 Reassessment > When to reassess > People with treated OHT (baseline IOP 24 mmHg or more) and a normal optic nerve head and visual field at most recent assessment > 1.5.11; PDF sheet 18; top 89.7: begins 'use clinical judgement to assess control of IOP and' … ends 'risk of conversion to COAG, and' (to sheet 18 y107.7) |
| Extraction-stage exclusion rationale (PG-04, as recorded) | Specific to people with treated OHT (previously treated; outside the initial decision). / limb (ii): No named in-scope root unit's meaning, applicability, scope or interpretation would change if this were removed. |

| Field | Content |
|---|---|
| Reviewer B finding (verbatim) | Excluding this passage omits decision-relevant source content concerning Glaucoma: diagnosis and management > Recommendations > 1.5 Reassessment > When to reassess > People with treated OHT (baseline IOP 24 mmHg or more) and a normal optic nerve head and visual field at most recent assessment > 1.5.11: "• use clinical judgement to assess control of IOP and the risk of conversion to COAG, and" |
| Category / stance / materiality | RF-EXCLUSION-QUESTIONED / ASSERTED / MATERIAL |
| Disposition and reason | DSP-NO-CHANGE. Verbatim: «Valutato il contenuto puntuale di questo item, il passo è fuori dal dominio dichiarato oppure non condiziona le unità in scope: Specific to people with treated OHT (previously treated; outside the initial decision). L'esclusione PG-04 resta invariata; il finding resta registrato senza riclassificazioni.» Translation: "Having assessed the specific content of this item, the passage is outside the declared domain or does not condition the in-scope units: Specific to people with treated OHT (previously treated; outside the initial decision). The PG-04 exclusion stands; the finding remains on record without reclassification." |

#### Table S3-10. Case-space boundary-exclusion log (PG21_EXCLUSION_LOG.csv)

PG-21 boundary-exclusion log in full by reason (P21-07 (5)); excluded sides are listed as not tested (P25-03).

| variable_id | cut_point_ref | excluded_side | reason | coverage |
|---|---|---|---|---|
| CV-AAO-0009C06-OHT | E-0009C06-C1 | false | CASE_LEVEL_POPULATION_ELIGIBILITY (P21-07 (3); P21-10; TX-S8-POP-AAO-0009C06-OHT) | excluded side NOT TESTED |

#### Table S3-11. Exclusion findings by recorded material class (post hoc)

Post-hoc descriptive tabulation of sealed exclusion outcomes by guideline and recorded PG-03 material class (A, candidate proposition; B, meaning-changing dependency; C, coverage-only material with no Class-A candidate or Class-B function). Material class was recorded before review. Entries are finding-raised/total (within-class percentage). No pooled analysis or cross-guideline comparison was performed; the tabulation does not explain the guideline-specific finding rates.

| Recorded PG-03 material class | EGS | AAO | NICE |
|---|---|---|---|
| A | 568/1,779 (31.9%) | 199/200 (99.5%) | 128/139 (92.1%) |
| B | 33/290 (11.4%) | 17/25 (68.0%) | 23/33 (69.7%) |
| C | 0/40 (0.0%) | 0/9 (0.0%) | 0/7 (0.0%) |

Source: sealed exclusion workbook (SHA-256 1e641ffd6ab3bec361bd64c7c87364906dd8d46a57dc8219f30982220f13fc41), fields material_class and item_outcome; the same material_class values appear in the workbook as released to the reviewer.

#### Table S3-12. Post-hoc second reading of a sample of documented exclusions

After analysis, a further glaucoma ophthalmologist not otherwise involved in the study read a random sample of 246 documented exclusions (seed 20261031; strata: guideline × Reviewer B outcome × recorded material class, proportional allocation with largest remainders; all unflagged AAO and NICE exclusions). Materials: a workbook with the same fixed columns as Reviewer B's exclusion workbook, including the recorded material class, but without Reviewer B's answers, the exclusion rationales or the dispositions; the three locked source PDFs; and views of the retained units of each division (unit identifier, location and segmented text, without ledger or rationale). The instructions gave the original exclusion task, the frozen definition and the materiality question verbatim, without Reviewer B's later addendum, and recommended answering D1 and D2 from the source before opening the retained-unit view for D3. Questions: D1, the original exclusion task with its materiality question; D2, "Does this content directly bear on the initial treat-or-observe decision in untreated ocular hypertension?"; D3, "Is this passage needed to preserve the meaning, applicability, scope or interpretation of any retained in-domain unit?" (with the retained unit and a short reason). The reading was post hoc and not registered. PPA and NPA are agreement on raising a finding, conditional on Reviewer B's outcome; their Clopper–Pearson intervals treat each stratum as a sample from a hypothetical population of similar passages and are not finite-frame intervals (the unflagged AAO and NICE strata were read in full). κ and AC1 are not reported because sampling was stratified by Reviewer B's outcome. Each guideline is reported separately; no pooled analysis or cross-guideline comparison was performed. The post-hoc judgements were not adjudicated and did not alter the sealed findings or dispositions.

| Measure | EGS | AAO | NICE |
|---|---|---|---|
| Sampled: Reviewer B finding / no finding | 50 / 50 | 50 / 18 | 50 / 28 |
| Reviewer B finding → reader finding | 31 | 11 | 28 |
| Reviewer B finding → reader no finding | 19 | 39 | 22 |
| Reviewer B no finding → reader finding | 7 | 0 | 0 |
| Reviewer B no finding → reader no finding | 43 | 18 | 28 |
| Positive percent agreement (95% exact CI) | 31/50 (62.0%; 47.2–75.3) | 11/50 (22.0%; 11.5–36.0) | 28/50 (56.0%; 41.3–70.0) |
| Negative percent agreement (95% exact CI) | 43/50 (86.0%; 73.3–94.2) | 18/18 (100.0%; 81.5–100.0) | 28/28 (100.0%; 87.7–100.0) |
| Reviewer B finding → reader material finding | 27/50 | 10/50 | 16/50 |
| Reviewer B no finding → reader material finding | 2/50 | 0/18 | 0/28 |
| Reader findings answered material / not material | 29 / 9 | 10 / 1 | 16 / 12 |
| D2: bears directly on the initial treat-or-observe decision | 0/100 | 0/68 | 1/78 |
| D3: needed to preserve the meaning of a retained unit | 0/100 | 0/68 | 8/78 |
| Unable or cannot judge (any question) | 0 | 0 | 0 |

Strata: frame size, sample size and the second reader's positive answers (D1 finding / D2 yes / D3 yes), by Reviewer B outcome and recorded material class:

| Stratum | EGS | AAO | NICE |
|---|---|---|---|
| Reviewer B finding, class A | N=568; n=47; 31 / 0 / 0 | N=199; n=46; 11 / 0 / 0 | N=128; n=42; 28 / 1 / 8 |
| Reviewer B finding, class B | N=33; n=3; 0 / 0 / 0 | N=17; n=4; 0 / 0 / 0 | N=23; n=8; 0 / 0 / 0 |
| Reviewer B finding, class C | N=0; n=0 | N=0; n=0 | N=0; n=0 |
| Reviewer B no finding, class A | N=1,211; n=40; 7 / 0 / 0 | N=1; n=1; 0 / 0 / 0 | N=11; n=11; 0 / 0 / 0 |
| Reviewer B no finding, class B | N=257; n=9; 0 / 0 / 0 | N=8; n=8; 0 / 0 / 0 | N=10; n=10; 0 / 0 / 0 |
| Reviewer B no finding, class C | N=40; n=1; 0 / 0 / 0 | N=9; n=9; 0 / 0 / 0 | N=7; n=7; 0 / 0 / 0 |

Weighting: for each guideline, estimate = Σ over strata of N_h × (x_h / n_h), divided by the guideline's total N, where N_h is the stratum frame size, n_h the sample size and x_h the number of positive answers. Intervals were not computed: several strata have zero or very few positive answers and small samples, where approximate design-based intervals are unreliable, and no interval method was set in advance.

Design-weighted estimates for all documented exclusions of each guideline (post hoc; weighted as above; intervals not computed):

| **Estimate** | **EGS** | **AAO** | **NICE** |
|---|---|---|---|
| Exclusions on which the second reader would raise a finding | 27.8% | 20.3% | 47.7% |
| D2 = yes | 0.0% | 0.0% | 1.7% |
| D3 = yes | 0.0% | 0.0% | 13.6% |

Items with D3 = yes (not adjudicated). Passages are abridged from Reviewer B's sealed finding text; rationales from the extraction ledger; the second reader's reasons are translated from Spanish. L-180 is also the one D2 = yes passage. For L-230 the second reader raised no D1 finding but named the executed NICE branch (recommendation 1.4.4) as the retained unit. L-180 concerns additional treatment after SLT, whereas D2 asks about the initial decision in untreated OHT; the reader's reason refers to an "initial or adjunctive" decision, so the second reading also shows uncertainty in applying the domain boundary. The answer and the count are reported as given.

**L-177 — X1-NICE-CD-0014-C01** (1.1.5 (case-finding; referral criteria), PDF sheet 6; Reviewer B: FINDING-RAISED; second reader D1: FINDING-RAISED, D2: NO)

Excluded passage: "there is optic nerve head damage on stereoscopic slit lamp biomicroscopy or". Recorded exclusion rationale: Referral criterion defined by optic-nerve findings, not OHT. Retained unit named: NICE-CD-0014-C03: "IOP is 24 mmHg or more using Goldmann-type applanation tonometry". Second reader's reason: Disjunctive referral criterion together with the IOP threshold.

**L-180 — X1-NICE-CD-0035-C04** (1.4.6 (initial treatment for people with OHT), PDF sheet 10; Reviewer B: FINDING-RAISED; second reader D1: FINDING-RAISED, D2: YES)

Excluded passage: "they have had 360° SLT but need additional treatment to reduce their IOP sufficiently to prevent the risk of visual impairment". Recorded exclusion rationale: Additional treatment after SLT (previously treated; escalation). Retained unit named: NICE-CD-0035-C01: "they choose not to have 360° SLT or". Second reader's reason: Defines the condition for adding a prostaglandin analogue after SLT in OHT; comment: bears on the initial or adjunctive decision in OHT.

**L-207 — X1-NICE-CD-0073-C02** (1.5.10 (reassessment), PDF sheet 17; Reviewer B: FINDING-RAISED; second reader D1: FINDING-RAISED, D2: NO)

Excluded passage: "… if appropriate, factors affecting adherence to treatment, including cognitive impairment and any treatment side effects". Recorded exclusion rationale: Adherence and side effects of established treatment (previously treated). Retained unit named: NICE-CD-0073-C01: "1.5.10 At each assessment, ask about general health and,". Second reader's reason: Specifies what to assess in the periodic history.

**L-217 — X1-NICE-CD-0087-C06** (1.6.6 (organisation of care), PDF sheet 22; Reviewer B: FINDING-RAISED; second reader D1: FINDING-RAISED, D2: NO)

Excluded passage: "eligibility for 360° SLT". Recorded exclusion rationale: Eligibility for a specific treatment modality (laser selection after the decision). Retained unit named: NICE-CD-0087-C01: "risk factors for conversion to COAG". Second reader's reason: Competence in selecting patients for SLT.

**L-218 — X1-NICE-CD-0088-C03** (1.6.7 (organisation of care), PDF sheet 23; Reviewer B: FINDING-RAISED; second reader D1: FINDING-RAISED, D2: NO)

Excluded passage: "the suitability of the procedure". Recorded exclusion rationale: Training on SLT suitability (modality selection after the decision). Retained unit named: NICE-CD-0088-C01: "1.6.7 … discuss with the responsible consultant ophthalmologist the decision to offer 360° SLT and how it will be performed". Second reader's reason: Content of the supervised SLT training programme.

**L-219 — X1-NICE-CD-0092-C05** (1.7.1 (providing information), PDF sheet 24; Reviewer B: FINDING-RAISED; second reader D1: FINDING-RAISED, D2: NO)

Excluded passage: "that most people having treatment for COAG will have good quality of life and not go blind". Recorded exclusion rationale: Prognosis statement about people treated for COAG (established glaucoma). Retained unit named: NICE-CD-0092-C03: "about their specific condition (OHT, suspected COAG and COAG), its life-long implications and their prognosis for retention of sight". Second reader's reason: Key prognostic message to give the patient.

**L-230 — X1-NICE-CD-0113-C17** (Rationale and impact, SLT for OHT or COAG, PDF sheet 31; Reviewer B: FINDING-RAISED; second reader D1: NO-FINDING-RAISED, D2: NO)

Excluded passage: "… there was no evidence on the use of 360° SLT in people with pigment dispersion syndrome and … eye drop treatment is more suitable for those people". Recorded exclusion rationale: Modality for pigment dispersion syndrome (secondary-OHT context). Retained unit named: NICE-CD-0032-C01 (the executed NICE branch, recommendation 1.4.4): "Offer 360° SLT … (excluding cases associated with pigment dispersion syndrome) …". Second reader's reason: Clinical justification for excluding pigment dispersion syndrome from the SLT recommendation.

**L-238 — X1-NICE-CD-0119-C04** (Rationale and impact, organisation of care, PDF sheet 33; Reviewer B: FINDING-RAISED; second reader D1: FINDING-RAISED, D2: NO)

Excluded passage: "… with support from a consultant ophthalmologist, healthcare professionals such as specialty doctors … can perform 360° SLT". Recorded exclusion rationale: Who may perform SLT (procedural). Retained unit named: NICE-CD-0088-C01 (recommendation 1.6.7, as above). Second reader's reason: Defines the non-medical professionals who may perform SLT; comment: delimits the professional scope of 1.6.7.

Data handling: forty-eight returned AAO rows (L-121–L-168) lacked the grey guideline field, shifting subsequent fields one column left; AAO was re-inserted by script, after which all nine grey fields in all 246 rows matched the sent workbook, with all returned response values preserved. Both the file as received and the corrected file are kept (hashes below).

Sampling procedure: frames were the documented exclusions of the sealed exclusion workbook (sheet X1_ITEMS; SHA-256 1e641ffd6ab3bec361bd64c7c87364906dd8d46a57dc8219f30982220f13fc41), split by guideline × Reviewer B outcome × recorded material class. Strata were processed in sorted key order; within each, item identifiers were sorted and a fixed number drawn with Python random.Random(20261031).sample (unflagged AAO and NICE strata taken in full); reading identifiers L-001–L-246 follow guideline, then item identifier.

Provenance of the second reading (all files to be released on OSF with the other records):

| File | Role | SHA-256 |
|---|---|---|
| build_macaya.py | Sampling and package script (not sent) | 48297f04220b58e4e6f6d427d5ed891907efe3635f28174b19aec6fa612a46bb |
| SAMPLING_RECORD.json | Seed, frames and allocation (not sent) | 881dc35740af589dcdb2315827dfe2f6f7a980682e220431987587520e1da1eb |
| KEY_LECTURA_TO_X1.csv | Key from reading identifier to exclusion item (not sent) | f9ab5cbbff2844598a1f0b7d9456673acd3e5d544f02707c7e6a2d46b3d9dff7 |
| OFTYGR_SEGUNDA_LECTURA_EXCLUSIONES.xlsx | Masked workbook sent | 4cc09fdb3c7cb281ae112e66d872b2153de452e62d15ced3378702d1b770231e |
| RETENIDAS_EGS.html | Retained-unit view sent, EGS | e7a2a8a176f3f544ddce9b6226b9f9d1b22fb4220b33054bfbacb7e9cfdaaf63 |
| RETENIDAS_AAO.html | Retained-unit view sent, AAO | 5ba5f1a8228399bd90aea2263687c3639863e611912c44183eba4213ead808d6 |
| RETENIDAS_NICE.html | Retained-unit view sent, NICE | c5f4a620643ca479189a7506a75104abeedb0a32e886003dcd3dedced17c0de3 |
| SOURCE_EGS.pdf | Locked source sent, EGS | bb716d8456fcf56e78b77055d9cdb1e4dbc02175b628c0c200170f4fae58d270 |
| SOURCE_AAO.pdf | Locked source sent, AAO | aef601ad5ca7ec9adcc101c09a8c042ccc5ba1e9708be9c3591ce3e94a6e85f1 |
| SOURCE_NICE.pdf | Locked source sent, NICE | 1806de5bbeceb5a0b2febad86ef3c900d97350dc0bea62d59e06d00b4d691c3c |
| MACAYA2_AS_RECEIVED.csv | Returned answers, as received | 0d97ba65fe9e287da2ef3c390f8dda9c7d24fb9aa65ab217b89d130c4212648f |
| MACAYA_REALIGNED.csv | Returned answers, corrected | c81b01d9614f81b283169fe4eb501f76b7b8b8b45992a5655b1d53a8594c7f39 |
| analyse_macaya.py | Correction and analysis script | 7920b019546de70537f33c0081a4fa57377933380dc95199b03f6caf5040ab46 |
| AGREEMENT_SUMMARY.json | Analysis output | 91aba3e8b199cb904685c0a2cbd7faecc35ee2f8c4803ac836379607a80843dd |
