## Supplementary Data S4 - Freeze-register digest for "Clinical review before execution of executable guideline encodings: a prospective registered study of three ocular hypertension guidelines"

### Supplementary Data S4. Freeze-register digest, entries 1–285

Clinical review before execution of executable guideline encodings: a prospective registered study of three ocular hypertension guidelines.

One block per register entry. Field names are those used in Supplement S1.2: entry_seq, entry_date_utc (day only), entry_type, governing_provisions, manifest_id, subject_stable_ids with their subject_sha256 values (listed in pairs), committed_or_manifest_sha256, prev_entry_sha256 and entry_line_sha256. A dash marks an empty field. The content is identical to the digest file committed with the study records (SHA-256 of the source CSV: 867e3fe712d0cbb5ed3cd67590843b32e6e4008a4bccc5b1344fd0984b4f59de).

| **Entry 1** | 2026-09-30 · FREEZE_ACT · manifest M0001 |
|---|---|
| **governing_provisions** | P26-18; PG26-A1-04(a); PG26-A1-06 |
| **subject** | PG26-M0001<br>7eff54880858f0485301456559aa1e9a1f10102158f39001d2a0c325381477d6 |
| **committed_or_manifest_sha256** | 7eff54880858f0485301456559aa1e9a1f10102158f39001d2a0c325381477d6 |
| **prev_entry_sha256** | GENESIS |
| **entry_line_sha256** | cac3539276b341c1a6fe496c72032992c8c51b6b8d9283a087bc465ae80cd7b3 |

| **Entry 2** | 2026-09-30 · COMMITMENT · manifest – |
|---|---|
| **governing_provisions** | P26-02(iii) as amended by CA2-10/CA3-10; PG26-A1-04(c); PG26-A1-06 |
| **subject** | PG26-M0001<br>7eff54880858f0485301456559aa1e9a1f10102158f39001d2a0c325381477d6 |
| **committed_or_manifest_sha256** | 7eff54880858f0485301456559aa1e9a1f10102158f39001d2a0c325381477d6 |
| **prev_entry_sha256** | cac3539276b341c1a6fe496c72032992c8c51b6b8d9283a087bc465ae80cd7b3 |
| **entry_line_sha256** | 4a573cb84d73608dd06e6b331d9f932e3bef6daa87455f11e0651609ac3de2a2 |

| **Entry 3** | 2026-09-30 · VERIFICATION_RECORD · manifest – |
|---|---|
| **governing_provisions** | P26-18; PG26-A1-04(e); PG26-A1-05(i) |
| **subject** | PG26-M0001<br>7eff54880858f0485301456559aa1e9a1f10102158f39001d2a0c325381477d6 |
| **committed_or_manifest_sha256** | – |
| **prev_entry_sha256** | 4a573cb84d73608dd06e6b331d9f932e3bef6daa87455f11e0651609ac3de2a2 |
| **entry_line_sha256** | 3ed8e3498bafa3ee18e01b300cc13d60bbc2594529ed9f0d52dc0bb02da19b34 |

| **Entry 4** | 2026-09-30 · ROLE_RECORD · manifest – |
|---|---|
| **governing_provisions** | P32-14 as amended by PG32-A1-01, A1-03; PG26-A1-06 |
| **subject** | PG32-D1<br>efa001ee06ce6b2b89b29250be4f01ce84ffc62d5297fbda3378c19a2815dd61 |
| **committed_or_manifest_sha256** | – |
| **prev_entry_sha256** | 3ed8e3498bafa3ee18e01b300cc13d60bbc2594529ed9f0d52dc0bb02da19b34 |
| **entry_line_sha256** | d104dbcb81a9efc80caa8f210aa4356ddda17dcb87b97a13ac114880226720d9 |

| **Entry 5** | 2026-09-30 · FREEZE_ACT · manifest M0002 |
|---|---|
| **governing_provisions** | P26-18; P26-04; P26-02(vi) |
| **subject** | PG26-M0002<br>531ec90152882cc386119245cc1111c29fbbecfee1ccebd3523f090cf6c9bbee |
| **committed_or_manifest_sha256** | 531ec90152882cc386119245cc1111c29fbbecfee1ccebd3523f090cf6c9bbee |
| **prev_entry_sha256** | d104dbcb81a9efc80caa8f210aa4356ddda17dcb87b97a13ac114880226720d9 |
| **entry_line_sha256** | 3276331e4a75b083dc8d8ca70eb3a90f95d00495afb5447527887171a9b3071b |

| **Entry 6** | 2026-09-30 · COMMITMENT · manifest – |
|---|---|
| **governing_provisions** | P26-02(iii) as amended by CA2-10/CA3-10; P26-04 |
| **subject** | PG26-M0002<br>531ec90152882cc386119245cc1111c29fbbecfee1ccebd3523f090cf6c9bbee |
| **committed_or_manifest_sha256** | 531ec90152882cc386119245cc1111c29fbbecfee1ccebd3523f090cf6c9bbee |
| **prev_entry_sha256** | 3276331e4a75b083dc8d8ca70eb3a90f95d00495afb5447527887171a9b3071b |
| **entry_line_sha256** | 7cb7c6dfa07fb433cd8f5bd896b82f77ac13d1b3afac7d527522c6fd9dd21e16 |

| **Entry 7** | 2026-09-30 · VERIFICATION_RECORD · manifest – |
|---|---|
| **governing_provisions** | P26-18; P26-04 |
| **subject** | PG26-M0002<br>531ec90152882cc386119245cc1111c29fbbecfee1ccebd3523f090cf6c9bbee |
| **committed_or_manifest_sha256** | – |
| **prev_entry_sha256** | 7cb7c6dfa07fb433cd8f5bd896b82f77ac13d1b3afac7d527522c6fd9dd21e16 |
| **entry_line_sha256** | 54d6e70d7e189166648bc09c4cd166f265607ee81b7a86057d424d11f7a11d9c |

| Entry 8 | 2026-10-01 · MILESTONE_RECORD · manifest – |
|---|---|
| governing_provisions | P29-12; P29-13; P29-16; P29-18; P29-19(b); P26-01; P26-02(vi) |
| subject | M1-RECORD<br>bab6572aaa4e2526e33174733c176ad2f50ff2f9ea1a86fdd09a905acd600413<br>M1-REGISTERED-FILES<br>a0aa25bc8d149906e1e68f87bdd81b4bd193b484cc5fe96ba1fc72f34b06b259<br>OSF-UPLOAD-58-FILES<br>9e645b825b12d9d344483eb4000c6d94296568195f75b3cc1d6db35a67699632<br>D-4<br>419e5ac34d55cef538c3ddd0f4a070596d6e93141726738d977d0cda21ae6428 |
| committed_or_manifest_sha256 | – |
| prev_entry_sha256 | 54d6e70d7e189166648bc09c4cd166f265607ee81b7a86057d424d11f7a11d9c |
| entry_line_sha256 | 2e1ce51b898e6d1dbdfa37fa52ff2802d358a2c5cb811926c9422a6458a2672d |

| Entry 9 | 2026-10-01 · VERIFICATION_RECORD · manifest – |
|---|---|
| governing_provisions | P29-19(b); P29-12; P26-18 |
| subject | M1-RECORD<br>bab6572aaa4e2526e33174733c176ad2f50ff2f9ea1a86fdd09a905acd600413 |
| committed_or_manifest_sha256 | – |
| prev_entry_sha256 | 2e1ce51b898e6d1dbdfa37fa52ff2802d358a2c5cb811926c9422a6458a2672d |
| entry_line_sha256 | 0ee08bede399f5abbe47e7a467a22910daa991b49e63058a1ad8da2f893a8656 |

| Entry 10 | 2026-10-01 · ROLE_RECORD · manifest – |
|---|---|
| governing_provisions | P32-14; P32-13; P32-02 |
| subject | PG32-D1<br>3f16a44b61e09cca02bd13cdcb7a9025f05b7b3b23f05730d1fd09c2adbf8af3 |
| committed_or_manifest_sha256 | – |
| prev_entry_sha256 | 0ee08bede399f5abbe47e7a467a22910daa991b49e63058a1ad8da2f893a8656 |
| entry_line_sha256 | 545345386fbca9c5a2d6eeb086e05e22acbc8aea6f89d1270c5b4b83d72a230c |

| Entry 11 | 2026-10-01 · ROLE_RECORD · manifest – |
|---|---|
| governing_provisions | P32-14; P32-13; P32-02 |
| subject | PG32-D1<br>a40d560e18a15a08f86487e297c1238c36369f1cddab0f41ba4d8b77854b7ab6 |
| committed_or_manifest_sha256 | – |
| prev_entry_sha256 | 545345386fbca9c5a2d6eeb086e05e22acbc8aea6f89d1270c5b4b83d72a230c |
| entry_line_sha256 | 15472debb699fc339931fe404140b34d7565afa4e5e8c3869c99e37fe6551aef |

| Entry 12 | 2026-10-01 · COMMITMENT · manifest – |
|---|---|
| governing_provisions | PG-03 §18 (E-09); PG-03 §19; P26-02(iii) as amended by CA2-10/CA3-10 |
| subject | PG03-S18-ENUMERATION<br>5b65b88e8bc07c3866170e9ad524cd8f555ddf7a226412b8462fec2001170df2<br>PG03-S18-FIXING-ACT<br>d645059dc355437c0ab47da65504bb21f0f19e04137e6bbb67b5dec29fdd4e81 |
| committed_or_manifest_sha256 | 5b65b88e8bc07c3866170e9ad524cd8f555ddf7a226412b8462fec2001170df2 |
| prev_entry_sha256 | 15472debb699fc339931fe404140b34d7565afa4e5e8c3869c99e37fe6551aef |
| entry_line_sha256 | a5a26b54879b2a5eadd5aa488ea12ca0b0d6f6eecf97d1ed10e45f8471f5188c |

| Entry 13 | 2026-10-01 · VERIFICATION_RECORD · manifest – |
|---|---|
| governing_provisions | PG-03 §18 (E-09); P26-18 |
| subject | PG03-S18-ENUMERATION<br>5b65b88e8bc07c3866170e9ad524cd8f555ddf7a226412b8462fec2001170df2 |
| committed_or_manifest_sha256 | – |
| prev_entry_sha256 | a5a26b54879b2a5eadd5aa488ea12ca0b0d6f6eecf97d1ed10e45f8471f5188c |
| entry_line_sha256 | bdc367a05d8df9204020caabf49e0af834976f2cefa36610a3b69a64ff253fc7 |

| Entry 14 | 2026-10-01 · ROLE_RECORD · manifest – |
|---|---|
| governing_provisions | P32-14; P32-13; P32-02; PG-03 §19; PG-04 §23 |
| subject | PG32-D1<br>ae422a04c079ceb0e0ba6ad5dedaa024c5a828fd9a1730ce3fad0f9ea6b0e1fc |
| committed_or_manifest_sha256 | – |
| prev_entry_sha256 | bdc367a05d8df9204020caabf49e0af834976f2cefa36610a3b69a64ff253fc7 |
| entry_line_sha256 | e2ba9e94e318d73ca544774ec8c2cf5e8dd66009160b638f663087efceaee3bb |

| Entry 15 | 2026-10-01 · COMMITMENT · manifest – |
|---|---|
| governing_provisions | PG-15 §7; FPA-6 v0.3; P26-02(iii) as amended by CA2-10/CA3-10; CA2-32 |
| subject | SEED-OB<br>6e7068467108ceab1006986a8561445ad4d747acf6a8ee43e8d7ac74be7af6e4 |
| committed_or_manifest_sha256 | 6e7068467108ceab1006986a8561445ad4d747acf6a8ee43e8d7ac74be7af6e4 |
| prev_entry_sha256 | e2ba9e94e318d73ca544774ec8c2cf5e8dd66009160b638f663087efceaee3bb |
| entry_line_sha256 | 169426dfb0c63d47aa8c2a65de1dc67d3cd4c2f7e00ca8495338882bcb7f6d04 |

| Entry 16 | 2026-10-01 · VERIFICATION_RECORD · manifest – |
|---|---|
| **governing_provisions** | PG-15 §7; FPA-6 v0.3; P26-18; CA2-32 |
| **subject** | SEED-OB<br>`6e7068467108ceab1006986a8561445ad4d747acf6a8ee43e8d7ac74be7af6e4` |
| **committed_or_manifest_sha256** | – |
| **prev_entry_sha256** | `169426dfb0c63d47aa8c2a65de1dc67d3cd4c2f7e00ca8495338882bcb7f6d04` |
| **entry_line_sha256** | `6c5058f8ca8b17fd8819c2a8f50a83340de8c050b4739623a33226181c10e463` |

| Entry 17 | 2026-10-01 · COMMITMENT · manifest – |
|---|---|
| **governing_provisions** | PG-15 §7; FPA-6 v0.3; P26-02(iii) as amended by CA2-10/CA3-10 |
| **subject** | PG15-S7-FRAME-SNAPSHOT<br>`b9942e224e170a691bc3c2265f132f2ceb776c298e5ba45deff8a144f93b23b5`<br>PG15-S7-FRAME-COMPANION-RECORD<br>`415ec25d52026938c7e6c60fcec9389d076d9be5efbb6eaa678fd465f7731e32`<br>PG04-LEDGER-CLOSURE-EXPORT-MANIFEST<br>`f8068304273dcc5fd16e7865b0e33ae350429a99b3a8df77e2775554568131b3`<br>PG04-ADOPTED-LEDGER-AND-CLOSURE-CHECK<br>`dfbb3fdd0b537e9064a224cac7a0b58c7ef36279bb2b97a9b9dd1c0fe4b16c5f` |
| **committed_or_manifest_sha256** | `b9942e224e170a691bc3c2265f132f2ceb776c298e5ba45deff8a144f93b23b5` |
| **prev_entry_sha256** | `6c5058f8ca8b17fd8819c2a8f50a83340de8c050b4739623a33226181c10e463` |
| **entry_line_sha256** | `9b4f651b765be052a5824c67d189fe571da6a39daee5db8b855e99c5fb76118c` |

| Entry 18 | 2026-10-01 · VERIFICATION_RECORD · manifest – |
|---|---|
| **governing_provisions** | PG-15 §7; FPA-6 v0.3; P26-18; CA2-32 |
| **subject** | PG15-S7-FRAME-SNAPSHOT<br>`b9942e224e170a691bc3c2265f132f2ceb776c298e5ba45deff8a144f93b23b5` |
| **committed_or_manifest_sha256** | – |
| **prev_entry_sha256** | `9b4f651b765be052a5824c67d189fe571da6a39daee5db8b855e99c5fb76118c` |
| **entry_line_sha256** | `c5fb02c5d7c41a7369884045c962d51b32660cc01a1af0fbf2cc5657a14148dc` |

| Entry 19 | 2026-10-01 · COMMITMENT · manifest – |
|---|---|
| **governing_provisions** | PG-15 §7; FPA-6 §5.1; P26-03; IP-02 §5 |
| **subject** | SEED-OB-ACCESS-LOG<br>`69237b66baa2309e4afa25b1c09504da933272a35b047c64640c659954110b03`<br>OPTION-B-CLASS-A-SELECTION<br>`d6f2fcaf4bde6a135f430d6942d0ee96a7cc184f28905b6b1b95ea3db9ecc42c`<br>OPTION-B-CLASS-B-DUPLICATION<br>`82abf2a77d1af7178696882dfeb5da152e6fb2a397ac17e7aa522bce478d9343`<br>OPTION-B-ESTABLISHMENT-RECORD<br>`81d9cff053bf6093b1ec9fe97ce6153fabcfbd2104b3bead411658bf0bf7493c`<br>OPTION-B-INDEPENDENT-CHECK<br>`17a2aa1d3c516632bf5c236e775a29640e125ba8135ace1b60659319b414447d` |
| **committed_or_manifest_sha256** | `69237b66baa2309e4afa25b1c09504da933272a35b047c64640c659954110b03` |
| **prev_entry_sha256** | `c5fb02c5d7c41a7369884045c962d51b32660cc01a1af0fbf2cc5657a14148dc` |
| **entry_line_sha256** | `e375dc579bfc1c3d3ec7688f41fd796b56380f8d30e1bc181bc586e7c34e288a` |

| Entry 20 | 2026-10-01 · VERIFICATION_RECORD · manifest – |
|---|---|
| **governing_provisions** | PG-15 §7; FPA-6 §5.1; P26-18; CA2-32 |
| **subject** | SEED-OB-ACCESS-LOG<br>`69237b66baa2309e4afa25b1c09504da933272a35b047c64640c659954110b03` |
| **committed_or_manifest_sha256** | – |
| **prev_entry_sha256** | `e375dc579bfc1c3d3ec7688f41fd796b56380f8d30e1bc181bc586e7c34e288a` |
| **entry_line_sha256** | `8a258e1e4d30f326171874f865936cde0350384f80ea71103d74554c3ab6fd10` |

| Entry 21 | 2026-10-01 · ROLE_RECORD · manifest – |
|---|---|
| **governing_provisions** | P32-14; P32-13; P32-02; PG-14 E-07; PG-15 §7 |
| **subject** | PG32-D1<br>`f6ce4cc11e52c2948d06e53e111cd09dd0156a8ed6645ad1e2d7418689a710dc` |
| **committed_or_manifest_sha256** | – |
| **prev_entry_sha256** | `8a258e1e4d30f326171874f865936cde0350384f80ea71103d74554c3ab6fd10` |
| **entry_line_sha256** | `96edcfc6e514e42a4ea2a418dfbd0e521462af26c44957ecb73cd26a94a60ca2` |

| Entry 22 | 2026-10-01 · FREEZE_ACT · manifest M0003 |
|---|---|
| **governing_provisions** | P26-18; P26-14; P26-02(vi) |
| **subject** | PG26-M0003<br>`f5d31248654d5d4da25efd05221cf86ba64d71489fd090145a94df655a463e1b` |
| **committed_or_manifest_sha256** | `f5d31248654d5d4da25efd05221cf86ba64d71489fd090145a94df655a463e1b` |
| **prev_entry_sha256** | `96edcfc6e514e42a4ea2a418dfbd0e521462af26c44957ecb73cd26a94a60ca2` |
| **entry_line_sha256** | `f49cd69432c8aa3cca00de16120d3a2763606fa05c4a0f6b5f8648825e92e0bd` |

| Entry 23 | 2026-10-01 · COMMITMENT · manifest – |
|---|---|
| **governing_provisions** | P26-02(iii) as amended by CA2-10/CA3-10; P26-14 |
| **subject** | PG26-M0003<br>`f5d31248654d5d4da25efd05221cf86ba64d71489fd090145a94df655a463e1b` |
| **committed_or_manifest_sha256** | `f5d31248654d5d4da25efd05221cf86ba64d71489fd090145a94df655a463e1b` |
| **prev_entry_sha256** | `f49cd69432c8aa3cca00de16120d3a2763606fa05c4a0f6b5f8648825e92e0bd` |
| **entry_line_sha256** | `cd51029602ccc67083523409588172fb9e98d1c55ec27929f601cd55d54b2595` |

| Entry 24 | 2026-10-01 · VERIFICATION_RECORD · manifest – |
|---|---|
| governing_provisions | P26-18; P26-14 |
| subject | PG26-M0003<br>f5d31248654d5d4da25efd05221cf86ba64d71489fd090145a94df655a463e1b |
| committed_or_manifest_sha256 | – |
| prev_entry_sha256 | cd51029602ccc67083523409588172fb9e98d1c55ec27929f601cd55d54b2595 |
| entry_line_sha256 | 80b053da4d6bf38cc17cee10a230d9ad917155ab653e30c2f50defc2337f6075 |

| Entry 25 | 2026-10-01 · VERIFICATION_RECORD · manifest – |
|---|---|
| governing_provisions | P26-18; P32-14; P32-13 |
| subject | PG32-D1<br>f6ce4cc11e52c2948d06e53e111cd09dd0156a8ed6645ad1e2d7418689a710dc |
| committed_or_manifest_sha256 | – |
| prev_entry_sha256 | 80b053da4d6bf38cc17cee10a230d9ad917155ab653e30c2f50defc2337f6075 |
| entry_line_sha256 | b2c19babd6b310774877707af66598116b2f0643731385ceed04d9e3c1cfd534 |

| Entry 26 | 2026-10-01 · FREEZE_ACT · manifest M0004 |
|---|---|
| governing_provisions | P26-18; P26-14; P26-02(vi) |
| subject | PG26-M0004<br>9dd143038a2e137c6143923e2b9f3d4f8cd46dc1d4b7f5347c9dd06df7f86a13 |
| committed_or_manifest_sha256 | 9dd143038a2e137c6143923e2b9f3d4f8cd46dc1d4b7f5347c9dd06df7f86a13 |
| prev_entry_sha256 | b2c19babd6b310774877707af66598116b2f0643731385ceed04d9e3c1cfd534 |
| entry_line_sha256 | 3bd27f2e871d3945f4401bdb2b8c16a5259cf1941a29dd811b596299adb37bc7 |

| Entry 27 | 2026-10-01 · COMMITMENT · manifest – |
|---|---|
| governing_provisions | P26-02(iii) as amended by CA2-10/CA3-10; P26-14 |
| subject | PG26-M0004<br>9dd143038a2e137c6143923e2b9f3d4f8cd46dc1d4b7f5347c9dd06df7f86a13 |
| committed_or_manifest_sha256 | 9dd143038a2e137c6143923e2b9f3d4f8cd46dc1d4b7f5347c9dd06df7f86a13 |
| prev_entry_sha256 | 3bd27f2e871d3945f4401bdb2b8c16a5259cf1941a29dd811b596299adb37bc7 |
| entry_line_sha256 | 72e2cbbf15200720b17934208e3381ebc7416f4aad8505689ee5a49dbbdc1214 |

| Entry 28 | 2026-10-01 · VERIFICATION_RECORD · manifest – |
|---|---|
| governing_provisions | P26-18; P26-14; P26-12 |
| subject | PG26-M0004<br>9dd143038a2e137c6143923e2b9f3d4f8cd46dc1d4b7f5347c9dd06df7f86a13 |
| committed_or_manifest_sha256 | – |
| prev_entry_sha256 | 72e2cbbf15200720b17934208e3381ebc7416f4aad8505689ee5a49dbbdc1214 |
| entry_line_sha256 | 26b24451489185c169e7e5ae92f0d7fc7ceb110c068a6c1a9971dea364f6f928 |

| Entry 29 | 2026-10-02 · COMMITMENT · manifest – |
|---|---|
| governing_provisions | PG-15 sec. 9; PG-15 sec. 10 P-1; PG-15 sec. 10 P-2; PG-15 sec. 10 P-3 as replaced by CA3-15; PG-26 P26-03; PG-26 P26-02(iii) as replaced by CA3-10; CA2-32; IP-03 CF-2 |
| subject | OFTYGR-E1-PHASE1-A05DRAFT-v1<br>fd5ea551496a43be2ba95bca9821c21229fa7447db6e236bdd5e499860f65f1a<br>OFTYGR-E1-PHASE1-A05DRAFT-v1.SEAL_MANIFEST<br>f11b32030e424a98b10de9f4fa66519daae60bab89595036c8ace431583b41cf |
| committed_or_manifest_sha256 | fd5ea551496a43be2ba95bca9821c21229fa7447db6e236bdd5e499860f65f1a |
| prev_entry_sha256 | 26b24451489185c169e7e5ae92f0d7fc7ceb110c068a6c1a9971dea364f6f928 |
| entry_line_sha256 | 2f2c1dcede14c4154e735130aceabfabb78d02ade0db178019d7d0857901452e |

| Entry 30 | 2026-10-02 · VERIFICATION_RECORD · manifest – |
|---|---|
| governing_provisions | PG-26 P26-18 as amended by CA3-11; CA2-32; PG-15 sec. 10 P-3 as replaced by CA3-15; IP-04 |
| subject | OFTYGR-E1-PHASE1-A05DRAFT-v1<br>fd5ea551496a43be2ba95bca9821c21229fa7447db6e236bdd5e499860f65f1a |
| committed_or_manifest_sha256 | – |
| prev_entry_sha256 | 2f2c1dcede14c4154e735130aceabfabb78d02ade0db178019d7d0857901452e |
| entry_line_sha256 | bfb0fc773d0a04ef4cefe4a6b82b95da4736139874da6ed171ff98c44a4532eb |

| Entry 31 | 2026-10-02 · COMMITMENT · manifest – |
|---|---|
| governing_provisions | PG-15 §7; PG-15 §10; Stage-5 prerequisites map v1.1 row P-D; P-D/E1 F-1 focused re-check §4; IP-03 CF-4; P26-03; CA2-32; IP-02 §5 |
| subject | OFTYGR-E2-PHASE1-DELIVERY-IDENTITY-RECORD<br>e25920fe0f75511aeae972b7c7bfe8e23d58b5cad925ff966b4ab1ab44dca43a<br>OFTYGR-E2-PHASE1-DELIVERY-ZIP<br>121720667750cf1f33efac68e291b08f0d6c052511f8f3ff0d1407ae3231cae6<br>OFTYGR-E2-SUBSTRATE-v1.0.SUBSTRATE_MANIFEST<br>c2e59d0ac2281400b974fcf81b0bf71e32837d6ee8e9b5ec5e18981319323268<br>OFTYGR-E2-CONTEXT-SUBSET<br>ec2f695619db6923b15334bafd62092e111aba8a9214c809dd7aa8b857ad7f9a<br>OFTYGR-E2-CONTEXT-SUBSET-BINDING<br>a4432483d34c2328b2b6c3923e6f0d619a18e92785aefbd578b99ed926604182<br>OFTYGR-E2-DELIVERY-CUSTODY-ACCESS-LOG<br>3e0704a7a173550404ea46107b9cc221f043d44505b5836710a085a69c86dea6 |
| committed_or_manifest_sha256 | e25920fe0f75511aeae972b7c7bfe8e23d58b5cad925ff966b4ab1ab44dca43a |
| prev_entry_sha256 | bfb0fc773d0a04ef4cefe4a6b82b95da4736139874da6ed171ff98c44a4532eb |
| entry_line_sha256 | f6bdb2e0ba5a472d0c770824caaacbf1abacb1312e406384f588535181098eca |

| Entry 32 | 2026-10-02 · COMMITMENT · manifest – |
|---|---|
| **governing_provisions** | PG-15 §7; PG-15 §10; Stage-5 prerequisites map v1.1 row P-D; P-D/E1 F-1 focused re-check §4; IP-03 CF-4; P26-03; CA2-32; IP-02 §5 |
| **subject** | OFTYGR-E2-PHASE1-DELIVERY-IDENTITY-RECORD<br>`c1be8cd28c4b04cef389b114159a122d5f73e02e2511cb30d00cf7be2529cbcb` |
| | OFTYGR-E2-PHASE1-DELIVERY-ZIP<br>`eca5076739496056c912816445a2940f69b4c209a089ef1c21dd70af9033987b` |
| | OFTYGR-E2-PHASE1-WORKBOOK<br>`1d8c8bb2ab30b725aed4c98e76c07d4d9e6925617da52919c4052d1a8f73c078` |
| | OFTYGR-E2-WORKBOOK-GENERATOR<br>`0f4e37c26ed7b04616b9d674b952bf3bb429b8070c9cc40313c922e7426470ba` |
| | OFTYGR-E2-WORKBOOK-CONVERTER<br>`e6dae32fab10232367c48a3dbc9f1c013d1b3400ee361f888052a509be19da04` |
| | OFTYGR-E2-REVIEW-RENDERER<br>`7943e6e08c83e5ad1fe3c5f1ed73c85c26fa922a88e744fa18816fc9ab9f4769` |
| | OFTYGR-E2-SUBSTRATE-v1.0.SUBSTRATE_MANIFEST<br>`c2e59d0ac2281400b974fcf81b0bf71e32837d6ee8e9b5ec5e18981319323268` |
| | OFTYGR-E2-CONTEXT-SUBSET<br>`ec2f695619db6923b15334bafd62092e111aba8a9214c809dd7aa8b857ad7f9a` |
| | OFTYGR-E2-DELIVERY-CUSTODY-ACCESS-LOG<br>`3e0704a7a173550404ea46107b9cc221f043d44505b5836710a085a69c86dea6` |
| | OFTYGR-E2-DELIVERY-CUSTODY-ACCESS-LOG<br>`1729ae6e1528c46bf030325fdcda64eb8e6e2aae991e4553c099567379e0d306` |
| **committed_or_manifest_sha256** | `c1be8cd28c4b04cef389b114159a122d5f73e02e2511cb30d00cf7be2529cbcb` |
| **prev_entry_sha256** | `f6bdb2e0ba5a472d0c770824caaacbf1abacb1312e406384f588535181098eca` |
| **entry_line_sha256** | `18ce818fce5415c9d6a67bd4b4f78f2b21aef878cadb273a0cd1a5ebb01165fd` |

| Entry 33 | 2026-10-02 · VERIFICATION_RECORD · manifest – |
|---|---|
| **governing_provisions** | CA2-32; PG-26 P26-18 as amended by CA3-11; PG-15 §7; Stage-5 prerequisites map v1.1 row P-D; IP-04 |
| **subject** | OFTYGR-E2-PHASE1-DELIVERY-IDENTITY-RECORD<br>`c1be8cd28c4b04cef389b114159a122d5f73e02e2511cb30d00cf7be2529cbcb` |
| **committed_or_manifest_sha256** | – |
| **prev_entry_sha256** | `18ce818fce5415c9d6a67bd4b4f78f2b21aef878cadb273a0cd1a5ebb01165fd` |
| **entry_line_sha256** | `b6bb2dcbb1268c50a7037df2267c8235665df00b121d1a4408e12f7309d19da8` |

| Entry 34 | 2026-10-02 · COMMITMENT · manifest – |
|---|---|
| **governing_provisions** | PG-15 sec. 9; PG-15 sec. 10 P-1; PG-15 sec. 10 P-2; PG-15 sec. 10 P-3 as replaced by CA3-15; PG-26 P26-03; PG-26 P26-02(iii) as replaced by CA3-10; CA2-32; IP-03 CF-2 |
| **subject** | OFTYGR-E2-PHASE1-v1<br>`1d81d5ea67df8ce0d6890a003179b5ac45aa39491d36daa9eec48edd119d8b0a` |
| | OFTYGR-E2-PHASE1-v1.SEAL_MANIFEST<br>`42d5415ae0472783218785a72548151c6be6418070e600fb67719c163a99f7d5` |
| **committed_or_manifest_sha256** | `1d81d5ea67df8ce0d6890a003179b5ac45aa39491d36daa9eec48edd119d8b0a` |
| **prev_entry_sha256** | `b6bb2dcbb1268c50a7037df2267c8235665df00b121d1a4408e12f7309d19da8` |
| **entry_line_sha256** | `7ae46c8ee2ff73ef05c765bb9f9846c3f222dde0fe36dc838281436fb062d35d` |

| Entry 35 | 2026-10-02 · VERIFICATION_RECORD · manifest – |
|---|---|
| **governing_provisions** | PG-26 P26-18 as amended by CA3-11; CA2-32; PG-15 sec. 10 P-3 as replaced by CA3-15; IP-04 |
| **subject** | OFTYGR-E2-PHASE1-v1<br>`1d81d5ea67df8ce0d6890a003179b5ac45aa39491d36daa9eec48edd119d8b0a` |
| **committed_or_manifest_sha256** | – |
| **prev_entry_sha256** | `7ae46c8ee2ff73ef05c765bb9f9846c3f222dde0fe36dc838281436fb062d35d` |
| **entry_line_sha256** | `d3110dffb5a4942d670343250e689f140d3ad97480f1cf21b2733bb09e4dfd9f` |

| Entry 36 | 2026-10-02 · ROLE_RECORD · manifest – |
|---|---|
| **governing_provisions** | P32-14; P32-13; P32-02; PG-11 sec. 17.2; PG-15 sec. 14.1; PG-14 E-08 |
| **subject** | PG32-D1<br>`f4c44c92e17482ae8b9aae7ab57a3a4204e365ac2ecfc68b6fc2a1a7f7e9f786` |
| **committed_or_manifest_sha256** | – |
| **prev_entry_sha256** | `d3110dffb5a4942d670343250e689f140d3ad97480f1cf21b2733bb09e4dfd9f` |
| **entry_line_sha256** | `a211d4356835c8c4aa7143787ac2d962672a612034695e629ca7aeba2b7a1187` |

| Entry 37 | 2026-10-02 · VERIFICATION_RECORD · manifest – |
|---|---|
| **governing_provisions** | P26-18; P32-14; P32-13 |
| **subject** | PG32-D1<br>`f4c44c92e17482ae8b9aae7ab57a3a4204e365ac2ecfc68b6fc2a1a7f7e9f786` |
| | A06-INPUT-INVENTORY<br>`e886834c59c69c10801369c9ad34800e57c11850a309d44d906c04b53fcddd83` |
| **committed_or_manifest_sha256** | – |
| **prev_entry_sha256** | `a211d4356835c8c4aa7143787ac2d962672a612034695e629ca7aeba2b7a1187` |
| **entry_line_sha256** | `616820313585989f887ea9c00ef79bf7866800cf7e4e31a3a010cf1af186925c` |

| Entry 38 | 2026-10-02 · ROLE_RECORD · manifest – |
|---|---|
| **governing_provisions** | P32-14; P32-13; P32-02; PG-11 sec. 17.2; PG-15 sec. 14.1; PG-14 E-08 |
| **subject** | PG32-D1<br>`575759b71f3b435b8761dca76cfffcdbfaac49a198c5576094f1a2760e2d29a3` |
| **committed_or_manifest_sha256** | – |
| **prev_entry_sha256** | `616820313585989f887ea9c00ef79bf7866800cf7e4e31a3a010cf1af186925c` |
| **entry_line_sha256** | `67642d93da318100a6362577bfc7cdac93f6de56910d5f5f537e350e9ad1f04c` |

| Entry 39 | 2026-10-02 · COMMITMENT · manifest – |
|---|---|
| **governing_provisions** | PG-15 §14.1 R-6; FPA-6 v0.3; LT-2 v0.4 PG-11 procedure P-1, P-2; P26-02(iii) as amended by CA2-10/CA3-10; CA2-32; IP-03 CF-2, CF-3 |
| **committed_or_manifest_sha256** | `73fa9e905608dc427e399824f1747c7467761a71ab913a9152c85ab4807af4d9` |
| **prev_entry_sha256** | `67642d93da318100a6362577bfc7cdac93f6de56910d5f5f537e350e9ad1f04c` |
| **entry_line_sha256** | `1dd9e12b05e2f60b63708c4987d212dd66d51534cd8208bd3a7e5f5db1e809b0` |

| Entry 40 | 2026-10-02 · COMMITMENT · manifest – |
|---|---|
| **governing_provisions** | PG-15 §14.1 R-6; FPA-6 v0.3; LT-2 v0.4 PG-11 procedure P-1, P-2; P26-02(iii) as amended by CA2-10/CA3-10; CA2-32; IP-03 CF-2, CF-3 |
| **committed_or_manifest_sha256** | `5137b55fa1198dc2b0a04763e4cf13a92d05c9413b176d70a596165459f17962` |
| **prev_entry_sha256** | `1dd9e12b05e2f60b63708c4987d212dd66d51534cd8208bd3a7e5f5db1e809b0` |
| **entry_line_sha256** | `fdc84da4f1a3fcebf95d0d3f2572475c0d7f860f89788111ef6989dd78a95e30` |

| Entry 41 | 2026-10-02 · COMMITMENT · manifest – |
|---|---|
| **governing_provisions** | PG-15 §14.1; PG-11 §17.2; LT-2 v0.4 PG-11 procedure P-3, P-4; P26-03; CA2-32; IP-03 CF-2 |
| **subject** | FREG-0001<br>`ac0bf96aebcc8bc5f14c92e5905de76eb738f93dd2dd0c889ffdc5acad84559e` |
| **committed_or_manifest_sha256** | `ac0bf96aebcc8bc5f14c92e5905de76eb738f93dd2dd0c889ffdc5acad84559e` |
| **prev_entry_sha256** | `fdc84da4f1a3fcebf95d0d3f2572475c0d7f860f89788111ef6989dd78a95e30` |
| **entry_line_sha256** | `bd5bc7dd3f0ca611d8abaf366a9b143340dae93e282a23f6e89bbbd91cb247c1` |

| Entry 42 | 2026-10-02 · COMMITMENT · manifest – |
|---|---|
| **governing_provisions** | PG-15 §14.1; PG-11 §17.2; LT-2 v0.4 PG-11 procedure P-3, P-4; P26-03; CA2-32; IP-03 CF-2 |
| **subject** | FREG-0002<br>`aaae28b2b3df79b18ead11a51ff8e08522412bfa2e76f68fba7e12c58c680f7c` |
| **committed_or_manifest_sha256** | `aaae28b2b3df79b18ead11a51ff8e08522412bfa2e76f68fba7e12c58c680f7c` |
| **prev_entry_sha256** | `bd5bc7dd3f0ca611d8abaf366a9b143340dae93e282a23f6e89bbbd91cb247c1` |
| **entry_line_sha256** | `d4d92c60b6465813e375859e6c34dc852e9fe1c38bc1a9c75cf5a6929bb44609` |

| Entry 43 | 2026-10-02 · VERIFICATION_RECORD · manifest – |
|---|---|
| **governing_provisions** | P26-18; P32-14; P32-13 |
| **subject** | PG32-D1<br>`575759b71f3b435b8761dca76cfffcdbfaac49a198c5576094f1a2760e2d29a3` |
| **committed_or_manifest_sha256** | – |
| **prev_entry_sha256** | `d4d92c60b6465813e375859e6c34dc852e9fe1c38bc1a9c75cf5a6929bb44609` |
| **entry_line_sha256** | `24ae315bf6f9ee5eb1eec69f77de22edc45822efdd1b9dd316fdbad04ca1874b` |

| Entry 44 | 2026-10-02 · VERIFICATION_RECORD · manifest – |
|---|---|
| **governing_provisions** | CA2-32; P26-18; LT-2 v0.4 PG-11 procedure P-5; PG-15 §14.1 R-6 |
| **committed_or_manifest_sha256** | – |
| **prev_entry_sha256** | `24ae315bf6f9ee5eb1eec69f77de22edc45822efdd1b9dd316fdbad04ca1874b` |
| **entry_line_sha256** | `9e75eca186492a47b41c82ff1199becaeda7125af994ed72ce74adf082987caf` |

| Entry 45 | 2026-10-02 · VERIFICATION_RECORD · manifest – |
|---|---|
| **governing_provisions** | CA2-32; P26-18; LT-2 v0.4 PG-11 procedure P-5; PG-15 §14.1 R-6 |
| **committed_or_manifest_sha256** | – |
| **prev_entry_sha256** | `9e75eca186492a47b41c82ff1199becaeda7125af994ed72ce74adf082987caf` |
| **entry_line_sha256** | `f66654f58553d68827705497d4700191d41a8d72dedaa1f230970920dafa5105` |

| Entry 46 | 2026-10-02 · VERIFICATION_RECORD · manifest – |
|---|---|
| **governing_provisions** | CA2-32; P26-18; LT-2 v0.4 PG-11 procedure P-5; PG-15 §14.1 |
| **committed_or_manifest_sha256** | – |
| **prev_entry_sha256** | `f66654f58553d68827705497d4700191d41a8d72dedaa1f230970920dafa5105` |
| **entry_line_sha256** | `61e1939fcf20b68fd61297a27a91bbb869f33eb5a788b92082797a6dec69ce97` |

| Entry 47 | 2026-10-02 · VERIFICATION_RECORD · manifest – |
|---|---|
| **governing_provisions** | CA2-32; P26-18; LT-2 v0.4 PG-11 procedure P-5; PG-15 §14.1 |
| **committed_or_manifest_sha256** | – |
| **prev_entry_sha256** | `61e1939fcf20b68fd61297a27a91bbb869f33eb5a788b92082797a6dec69ce97` |
| **entry_line_sha256** | `5f6d7b2c40a1bfb1cfcdb9f695edad350e5b95a37b6935158240c593a45f7305` |

| Entry 48 | 2026-10-02 · COMMITMENT · manifest – |
|---|---|
| **governing_provisions** | PG-15 §14.3; PG-15 §14.6; LT-2 v0.4 PG-11 procedure P-7 step 8; CA2-32; IP-03 CF-4 |
| **committed_or_manifest_sha256** | `4180abba878be3fd03c42528b9d876a24c0f2df97fbc07218abe3afb9f48d87b` |
| **prev_entry_sha256** | `5f6d7b2c40a1bfb1cfcdb9f695edad350e5b95a37b6935158240c593a45f7305` |
| **entry_line_sha256** | `3cd180baf2ad9390bbaf82ab244269f966a80c86ec95b93cc7517a15685d56ae` |

| Entry 49 | 2026-10-02 · COMMITMENT · manifest – |
| --- | --- |
| **governing_provisions** | PG-15 §14.3; PG-15 §14.6; LT-2 v0.4 PG-11 procedure P-7 step 8; CA2-32; IP-03 CF-4 |
| **committed_or_manifest_sha256** | 5e214bf44604213e39f2e9961092bc3c7d70e4a736b08054406ec081efcd9727 |
| **prev_entry_sha256** | 3cd180baf2ad9390bbaf82ab244269f966a80c86ec95b93cc7517a15685d56ae |
| **entry_line_sha256** | f7629cfcf11749ac46bc84b7efbdca1c5a26c1fdbc5894bb25da1de542080ffd |

| Entry 50 | 2026-10-02 · VERIFICATION_RECORD · manifest – |
| --- | --- |
| **governing_provisions** | CA2-32; P26-18; LT-2 v0.4 PG-11 procedure P-7, P-8; PG-15 §14.3 |
| **committed_or_manifest_sha256** | – |
| **prev_entry_sha256** | f7629cfcf11749ac46bc84b7efbdca1c5a26c1fdbc5894bb25da1de542080ffd |
| **entry_line_sha256** | 992662af44b4f5505b8002868a261a1ad74c0085937fdcaca02157d720541b73 |

| Entry 51 | 2026-10-02 · VERIFICATION_RECORD · manifest – |
| --- | --- |
| **governing_provisions** | CA2-32; P26-18; LT-2 v0.4 PG-11 procedure P-7, P-8; PG-15 §14.3 |
| **committed_or_manifest_sha256** | – |
| **prev_entry_sha256** | 992662af44b4f5505b8002868a261a1ad74c0085937fdcaca02157d720541b73 |
| **entry_line_sha256** | 9acaa05e207089de904cae797cd3c45f0f0ece257a0b082e464cef303f16abfa |

| Entry 52 | 2026-10-03 · COMMITMENT · manifest – |
| --- | --- |
| **governing_provisions** | PG-15 sec. 9; PG-15 sec. 10 P-1; PG-15 sec. 10 P-2; PG-15 sec. 10 P-3 as replaced by CA3-15; PG-26 P26-03; CA2-32; IP-03 CF-2 |
| **subject** | INITIAL_E2<br>525b11d6540f7380b2495daf98d06999671a399ead99c948a90213617ee27804 |
| **committed_or_manifest_sha256** | 525b11d6540f7380b2495daf98d06999671a399ead99c948a90213617ee27804 |
| **prev_entry_sha256** | 9acaa05e207089de904cae797cd3c45f0f0ece257a0b082e464cef303f16abfa |
| **entry_line_sha256** | 1d8a55af08a05193eeeb21047eec38794ddb00721c033dfbda0c768a9e9f1be4 |

| Entry 53 | 2026-10-03 · VERIFICATION_RECORD · manifest – |
| --- | --- |
| **governing_provisions** | PG-26 P26-18 as amended by CA3-11; CA2-32; PG-15 sec. 10 P-3 as replaced by CA3-15; IP-04 |
| **subject** | INITIAL_E2<br>525b11d6540f7380b2495daf98d06999671a399ead99c948a90213617ee27804 |
| **committed_or_manifest_sha256** | – |
| **prev_entry_sha256** | 1d8a55af08a05193eeeb21047eec38794ddb00721c033dfbda0c768a9e9f1be4 |
| **entry_line_sha256** | 9bffa26f73e5b38d0196c43efd7e8ed71bdf6fd749a6108b668d491dec2e24ea |

| Entry 54 | 2026-10-03 · COMMITMENT · manifest – |
| --- | --- |
| **governing_provisions** | PG-15 sec. 9; PG-15 sec. 10 P-1; PG-15 sec. 10 P-2; PG-15 sec. 10 P-3 as replaced by CA3-15; PG-26 P26-03; PG-26 P26-02(iii) as replaced by CA3-10; CA2-32; IP-03 CF-2 |
| **subject** | OFTYGR-E1-PHASE2-FAM-0001-ADOPTED-v1<br>60d314c890cdca492ba27cb904d323f8f0a9c7b829ba64e35c5c32e57364cc47<br>OFTYGR-E1-PHASE2-FAM-0001-ADOPTED-v1.SEAL_MANIFEST<br>6d9ccc9c6ba5d7e92851ce00f7a96fb08232676c3eb9c5a2c82dc4f81c46e015 |
| **committed_or_manifest_sha256** | 60d314c890cdca492ba27cb904d323f8f0a9c7b829ba64e35c5c32e57364cc47 |
| **prev_entry_sha256** | 9bffa26f73e5b38d0196c43efd7e8ed71bdf6fd749a6108b668d491dec2e24ea |
| **entry_line_sha256** | eb9ddaf40a79190499bb526f549ec2a7fc9c7f7f5cb27f240aacf233bd078b20 |

| Entry 55 | 2026-10-03 · VERIFICATION_RECORD · manifest – |
| --- | --- |
| **governing_provisions** | PG-26 P26-18 as amended by CA3-11; CA2-32; PG-15 sec. 10 P-3 as replaced by CA3-15; IP-04 |
| **subject** | OFTYGR-E1-PHASE2-FAM-0001-ADOPTED-v1<br>60d314c890cdca492ba27cb904d323f8f0a9c7b829ba64e35c5c32e57364cc47 |
| **committed_or_manifest_sha256** | – |
| **prev_entry_sha256** | eb9ddaf40a79190499bb526f549ec2a7fc9c7f7f5cb27f240aacf233bd078b20 |
| **entry_line_sha256** | 82a1eef202a5179e895278c4f89aaddb8f69b181fca95e2063159ad5efde93f6 |

| Entry 56 | 2026-10-03 · COMMITMENT · manifest – |
| --- | --- |
| **governing_provisions** | PG-15 sec. 9; PG-15 sec. 10 P-1; PG-15 sec. 10 P-2; PG-15 sec. 10 P-8; PG-15 sec. 10 P-3 as replaced by CA3-15; PG-26 P26-03; CA2-32; IP-03 CF-2 |
| **subject** | INITIAL_E1<br>4332df4a6a87c1d8868daa5c67d5ac7594887d862b22a70e929dcf442f832557 |
| **committed_or_manifest_sha256** | 4332df4a6a87c1d8868daa5c67d5ac7594887d862b22a70e929dcf442f832557 |
| **prev_entry_sha256** | 82a1eef202a5179e895278c4f89aaddb8f69b181fca95e2063159ad5efde93f6 |
| **entry_line_sha256** | c86b4ae530574576f8765749904ee508aa10e1d8034e9ced56299a1aba92d379 |

| Entry 57 | 2026-10-03 · VERIFICATION_RECORD · manifest – |
| --- | --- |
| **governing_provisions** | PG-26 P26-18 as amended by CA3-11; CA2-32; PG-15 sec. 10 P-3 as replaced by CA3-15; IP-04 |
| **subject** | INITIAL_E1<br>4332df4a6a87c1d8868daa5c67d5ac7594887d862b22a70e929dcf442f832557 |
| **committed_or_manifest_sha256** | – |
| **prev_entry_sha256** | c86b4ae530574576f8765749904ee508aa10e1d8034e9ced56299a1aba92d379 |
| **entry_line_sha256** | d5d18dddd8ec739bd101b087168211f13a2793c3d0cbd7c85aabc7f4f110d274 |

| Entry 58 | 2026-10-03 · DECISIVE_STEP_MANIFEST · manifest – |
| --- | --- |
| **governing_provisions** | PG-26 P26-25(3) as replaced by CA3-13; CA3-01; CA3-02; CA3-15; IP-01 sec. 7; IP-02 sec. 6 |
| **subject** | DSR0-01<br>dd1b8bc2fb2c76a335f595d020787d6299626e4c8c2f4e26363101169e3c33f6 |
| **committed_or_manifest_sha256** | – |
| **prev_entry_sha256** | d5d18dddd8ec739bd101b087168211f13a2793c3d0cbd7c85aabc7f4f110d274 |
| **entry_line_sha256** | edca9b6bec5d71cab87fb2cbf079a9eb98cc7948a45c7b4ffe4c0d515911f2cf |

| Entry 59 | 2026-10-03 · VERIFICATION_RECORD · manifest – |
|---|---|
| governing_provisions | PG-26 P26-18 as amended by CA3-11; IP-04 V-10a; CA3-13(4); CA2-32 |
| subject | DSR0-01<br>dd1b8bc2fb2c76a335f595d020787d6299626e4c8c2f4e26363101169e3c33f6 |
| committed_or_manifest_sha256 | – |
| prev_entry_sha256 | edca9b6bec5d71cab87fb2cbf079a9eb98cc7948a45c7b4ffe4c0d515911f2cf |
| entry_line_sha256 | f03fd02ded0272e2489aa4fe0d1c0bb050e83bec29df67563e5dd3c636baa17a |

| Entry 60 | 2026-10-03 · TSA_TOKEN · manifest – |
|---|---|
| governing_provisions | PG-26 P26-25(4), (6); P29-21; CA3-01; CA3-04; IP-06 |
| subject | DSR0-01<br>dd1b8bc2fb2c76a335f595d020787d6299626e4c8c2f4e26363101169e3c33f6 |
| committed_or_manifest_sha256 | – |
| prev_entry_sha256 | f03fd02ded0272e2489aa4fe0d1c0bb050e83bec29df67563e5dd3c636baa17a |
| entry_line_sha256 | f81df9f83b4050a06b2ae9c9d43cdbb7267872b404f2f5de09c459667a4b0f6f |

| Entry 61 | 2026-10-03 · VERIFICATION_RECORD · manifest – |
|---|---|
| governing_provisions | PG-26 P26-18 as amended by CA3-11; IP-04 V-10b; CA3-13(4); CA2-32 |
| subject | DSR0-01<br>dd1b8bc2fb2c76a335f595d020787d6299626e4c8c2f4e26363101169e3c33f6 |
| committed_or_manifest_sha256 | – |
| prev_entry_sha256 | f81df9f83b4050a06b2ae9c9d43cdbb7267872b404f2f5de09c459667a4b0f6f |
| entry_line_sha256 | a38c424edb820b49a2da235facb9a9916e2045e036e6587ab73a69d4e7082a54 |

| Entry 62 | 2026-10-03 · COMMITMENT · manifest – |
|---|---|
| governing_provisions | PG-15 §12.1 R0; R0 exposure act v1.1 (b67416f4); gatekeeper ruling X-1 (ba173453); IP-02 §5; P26-03; CA2-32 |
| subject | OFTYGR-R0-EXPOSURE-RECORD<br>d5ff8768293b9eb3787208823bc1c200c49e7884ae87e77dd5df115b662aa42c<br>OFTYGR-R0-TRANSPORT-INITIAL_E1-FOR-E2<br>97cd7d751c1540083944e89870d54086e56ce93b1cb691b7f3e9a18d2247ec0c<br>OFTYGR-R0-TRANSPORT-INITIAL_E2-FOR-E1<br>57800c74c313ea6b0ec04f33ad87051e3b59fe372a4ea9b053907d3a12c76f9f<br>OFTYGR-DSR0-D8-NOTICE<br>834f414676d5d3791d545b0a5cb446ffad0a0ff9700720c47dbfab2f387208cf<br>OFTYGR-R0-EXPOSURE-ACT<br>b67416f4aad6f4343453078e501ab020dbcf975ab8c63ff265c57d672d4ca3d1<br>OFTYGR-R0-EXPOSURE-ACT-ADOPTION-RECORD<br>186b6c49acccff2c6946757b18878dee623f05260bdd737deefe839e89e8e0ef<br>OFTYGR-R0-E2-RECEIPT-REPLY<br>c7384272b69bc45516465632d5d96ab4b79c13c1f223917b62148113aaa78374<br>OFTYGR-R0-RELEASE-E1-CUSTODY-ACCESS-LOG<br>1a76701851e1172c8f3a9c4e49e257f7acc1488069d7d32985f438ef5e49107e |
| committed_or_manifest_sha256 | d5ff8768293b9eb3787208823bc1c200c49e7884ae87e77dd5df115b662aa42c |
| prev_entry_sha256 | a38c424edb820b49a2da235facb9a9916e2045e036e6587ab73a69d4e7082a54 |
| entry_line_sha256 | f5049e3670c6c52c069a0fc1e910f23e54b973247d47facd487db84dbbf86b18 |

| Entry 63 | 2026-10-03 · VERIFICATION_RECORD · manifest – |
|---|---|
| governing_provisions | CA2-32; PG-26 P26-18 as amended by CA3-11; PG-15 §7; Stage-5 prerequisites map v1.1 row P-D; IP-04; PG-15 §12.1 R0 |
| subject | OFTYGR-R0-EXPOSURE-RECORD<br>d5ff8768293b9eb3787208823bc1c200c49e7884ae87e77dd5df115b662aa42c<br>OFTYGR-R0-TRANSPORT-INITIAL_E1-FOR-E2<br>97cd7d751c1540083944e89870d54086e56ce93b1cb691b7f3e9a18d2247ec0c<br>OFTYGR-R0-TRANSPORT-INITIAL_E2-FOR-E1<br>57800c74c313ea6b0ec04f33ad87051e3b59fe372a4ea9b053907d3a12c76f9f<br>OFTYGR-DSR0-D8-NOTICE<br>834f414676d5d3791d545b0a5cb446ffad0a0ff9700720c47dbfab2f387208cf<br>OFTYGR-R0-EXPOSURE-ACT<br>b67416f4aad6f4343453078e501ab020dbcf975ab8c63ff265c57d672d4ca3d1<br>OFTYGR-R0-EXPOSURE-ACT-ADOPTION-RECORD<br>186b6c49acccff2c6946757b18878dee623f05260bdd737deefe839e89e8e0ef<br>OFTYGR-R0-E2-RECEIPT-REPLY<br>c7384272b69bc45516465632d5d96ab4b79c13c1f223917b62148113aaa78374<br>OFTYGR-R0-RELEASE-E1-CUSTODY-ACCESS-LOG<br>1a76701851e1172c8f3a9c4e49e257f7acc1488069d7d32985f438ef5e49107e |
| committed_or_manifest_sha256 | – |
| prev_entry_sha256 | f5049e3670c6c52c069a0fc1e910f23e54b973247d47facd487db84dbbf86b18 |
| entry_line_sha256 | cd23020cec6abcc28543ffa855efc5b53501593da3a7b2186e7c1106fcba9f91 |

| Entry 64 | 2026-10-03 · COMMITMENT · manifest – |
|---|---|
| **governing_provisions** | PG-15 §14.3; PG-15 §7; FPA-6 §5.1; LT-2 PG-11 procedure P-11; gatekeeper ruling X-2 (32a735ec); gatekeeper ruling A-1/A-2/A-3 (2530c6cd); IP-02 §5; P26-03; CA2-32 |
| **subject** | OFTYGR-P11-S14_3-S7-DISCLOSURE-RECORD<br>b5b129bd5bd27ff252391bd500a704a7bee641d7cdf560bbd122ef3b74192a1b<br>OFTYGR-GATEKEEPER-RULING-A1-A2-A3<br>2530c6cd838b8e86850aaed2bca0a3a49095c1d7a44bc80804af61c425fa6d9b<br>OFTYGR-P11-SEED-RELEASE-ACCESS-LOG<br>7028e1e2558d65c85babdea639501d9a9b29b08a6a2eb142ccda25ba8c92f348<br>OFTYGR-DISCLOSURE-COPIES-ACCESS-LOG<br>f40ded13d66f17f7334f833dda45062da6a07d931b64f8a0d860f5842f1319fe<br>SEED-F-0001.COMMITMENT<br>73fa9e905608dc427e399824f1747c7467761a71ab913a9152c85ab4807af4d9<br>SEED-F-0002.COMMITMENT<br>5137b55fa1198dc2b0a04763e4cf13a92d05c9413b176d70a596165459f17962<br>N08A_AUDIT_SAMPLE_RECORD.FAM-0001<br>4180abba878be3fd03c42528b9d876a24c0f2df97fbc07218abe3afb9f48d87b<br>N08A_AUDIT_SAMPLE_RECORD.FAM-0002<br>5e214bf44604213e39f2e9961092bc3c7d70e4a736b08054406ec081efcd9727<br>OPTION-B-CLASS-A-SELECTION<br>d6f2fcaf4bde6a135f430d6942d0ee96a7cc184f28905b6b1b95ea3db9ecc42c<br>OPTION-B-CLASS-B-DUPLICATION<br>82abf2a77d1af7178696882dfeb5da152e6fb2a397ac17e7aa522bce478d9343 |
| **committed_or_manifest_sha256** | b5b129bd5bd27ff252391bd500a704a7bee641d7cdf560bbd122ef3b74192a1b |
| **prev_entry_sha256** | cd23020cec6abcc28543ffa855efc5b53501593da3a7b2186e7c1106fcba9f91 |
| **entry_line_sha256** | 53af87e50fd23069d8081c6bfd6d0476780eede1d8f7b826fd46224eb8489d5a |

| Entry 65 | 2026-10-03 · VERIFICATION_RECORD · manifest – |
|---|---|
| **governing_provisions** | CA2-32; PG-26 P26-18 as amended by CA3-11; PG-15 §14.3; PG-15 §7; FPA-6 §5.2; LT-2 PG-11 procedure P-11, P-12; IP-04 |
| **subject** | OFTYGR-P11-S14_3-S7-DISCLOSURE-RECORD<br>b5b129bd5bd27ff252391bd500a704a7bee641d7cdf560bbd122ef3b74192a1b<br>OFTYGR-GATEKEEPER-RULING-A1-A2-A3<br>2530c6cd838b8e86850aaed2bca0a3a49095c1d7a44bc80804af61c425fa6d9b<br>OFTYGR-P11-SEED-RELEASE-ACCESS-LOG<br>7028e1e2558d65c85babdea639501d9a9b29b08a6a2eb142ccda25ba8c92f348<br>OFTYGR-DISCLOSURE-COPIES-ACCESS-LOG<br>f40ded13d66f17f7334f833dda45062da6a07d931b64f8a0d860f5842f1319fe<br>SEED-F-0001.COMMITMENT<br>73fa9e905608dc427e399824f1747c7467761a71ab913a9152c85ab4807af4d9<br>SEED-F-0002.COMMITMENT<br>5137b55fa1198dc2b0a04763e4cf13a92d05c9413b176d70a596165459f17962<br>N08A_AUDIT_SAMPLE_RECORD.FAM-0001<br>4180abba878be3fd03c42528b9d876a24c0f2df97fbc07218abe3afb9f48d87b<br>N08A_AUDIT_SAMPLE_RECORD.FAM-0002<br>5e214bf44604213e39f2e9961092bc3c7d70e4a736b08054406ec081efcd9727<br>OPTION-B-CLASS-A-SELECTION<br>d6f2fcaf4bde6a135f430d6942d0ee96a7cc184f28905b6b1b95ea3db9ecc42c<br>OPTION-B-CLASS-B-DUPLICATION<br>82abf2a77d1af7178696882dfeb5da152e6fb2a397ac17e7aa522bce478d9343 |
| **committed_or_manifest_sha256** | – |
| **prev_entry_sha256** | 53af87e50fd23069d8081c6bfd6d0476780eede1d8f7b826fd46224eb8489d5a |
| **entry_line_sha256** | 2d0ad97c44f19c853f931788df470e320f98898e6adf0af97bd8f29a6f303d5e |

| Entry 66 | 2026-10-03 · ROLE_RECORD · manifest – |
|---|---|
| **governing_provisions** | P32-14; P32-13; P32-02; PG-15 sec. 12.1; PG-15 sec. 11.2; P26-14 |
| **subject** | PG32-D1<br>e61abf5d5e38ed28d443921942500b268837670b793d64b6574326c45bf20e57 |
| **committed_or_manifest_sha256** | – |
| **prev_entry_sha256** | 2d0ad97c44f19c853f931788df470e320f98898e6adf0af97bd8f29a6f303d5e |
| **entry_line_sha256** | 6c43df656b0708772d06b656174fa06825b598aeb942f527f8b7496aab7c3b87 |

| Entry 67 | 2026-10-03 · FREEZE_ACT · manifest M0005 |
|---|---|
| **governing_provisions** | P26-18; P26-14; P26-02(vi) |
| **subject** | PG26-M0005<br>6577b69eb31c2ff8524e6db9fd36408740ba3c95d15725a637e1053871aa6c09 |
| **committed_or_manifest_sha256** | 6577b69eb31c2ff8524e6db9fd36408740ba3c95d15725a637e1053871aa6c09 |
| **prev_entry_sha256** | 6c43df656b0708772d06b656174fa06825b598aeb942f527f8b7496aab7c3b87 |
| **entry_line_sha256** | 8aa094836da7eaf1c739a84220ce34359b2d43ac6c571aba69bfe9fa10728061 |

| Entry 68 | 2026-10-03 · COMMITMENT · manifest – |
|---|---|
| **governing_provisions** | P26-02(iii) as amended by CA2-10/CA3-10; P26-14 |
| **subject** | PG26-M0005<br>6577b69eb31c2ff8524e6db9fd36408740ba3c95d15725a637e1053871aa6c09 |
| **committed_or_manifest_sha256** | 6577b69eb31c2ff8524e6db9fd36408740ba3c95d15725a637e1053871aa6c09 |
| **prev_entry_sha256** | 8aa094836da7eaf1c739a84220ce34359b2d43ac6c571aba69bfe9fa10728061 |
| **entry_line_sha256** | fce658208890ac17e811bfa9421a19760515ba7a8259e6a29d5e004932897bff |

| Entry 69 | 2026-10-03 · VERIFICATION_RECORD · manifest – |
|---|---|
| governing_provisions | P26-18; P32-14; P32-13 |
| subject | PG32-D1<br>e61abf5d5e38ed28d443921942500b268837670b793d64b6574326c45bf20e57 |
| committed_or_manifest_sha256 | – |
| prev_entry_sha256 | fce658208890ac17e811bfa9421a19760515ba7a8259e6a29d5e004932897bff |
| entry_line_sha256 | 95f481b70c18777c2d561aff30b82159e858311921a759662789e9a8e7f1cfcb |

| Entry 70 | 2026-10-03 · VERIFICATION_RECORD · manifest – |
|---|---|
| governing_provisions | P26-18; P26-14; P26-12 |
| subject | PG26-M0005<br>6577b69eb31c2ff8524e6db9fd36408740ba3c95d15725a637e1053871aa6c09 |
| committed_or_manifest_sha256 | – |
| prev_entry_sha256 | 95f481b70c18777c2d561aff30b82159e858311921a759662789e9a8e7f1cfcb |
| entry_line_sha256 | cd87165a4aebd4dc1d748cb9174ffd59abd6a0cca421b83127e638f04f1d803d |

| Entry 71 | 2026-10-03 · COMMITMENT · manifest – |
|---|---|
| governing_provisions | PG-15 §11; PG-15 §12.1 R1; Act R v0.2 item 2 (3a20c4da); D-1 v7 A-08 (entry 66, verified 69); M0005 LOCK_LT2 6577b69e (entries 67-68, verified 70); CA2-32 |
| subject | OFTYGR-R1-DISCREPANCY-REGISTER<br>c5dbbe62add332b458e93ce6fb3c710669c43447d1c0ac53466b7d39322a056b<br>OFTYGR-R1-COMPARISON-TABLE<br>a7d2a75297a5585eab608136994fc4bb0aa94e166850cb08f0dff59e7e4ddc4c<br>OFTYGR-R1-COMPARISON-MANIFEST<br>0ea12b3e566c06e766712ccca179114321e13944c1911a876d03dd5238a83c07<br>INITIAL_E1<br>4332df4a6a87c1d8868daa5c67d5ac7594887d862b22a70e929dcf442f832557<br>INITIAL_E2<br>525b11d6540f7380b2495daf98d06999671a399ead99c948a90213617ee27804<br>OPTION-B-CLASS-A-SELECTION<br>d6f2fcaf4bde6a135f430d6942d0ee96a7cc184f28905b6b1b95ea3db9ecc42c<br>OPTION-B-CLASS-B-DUPLICATION<br>82abf2a77d1af7178696882dfeb5da152e6fb2a397ac17e7aa522bce478d9343<br>PG26-M0005<br>6577b69eb31c2ff8524e6db9fd36408740ba3c95d15725a637e1053871aa6c09 |
| committed_or_manifest_sha256 | c5dbbe62add332b458e93ce6fb3c710669c43447d1c0ac53466b7d39322a056b |
| prev_entry_sha256 | cd87165a4aebd4dc1d748cb9174ffd59abd6a0cca421b83127e638f04f1d803d |
| entry_line_sha256 | 2a65f19f897bc3d67bbce50c2e2877ccb337d8a3a2995b54033f048c2bc2720d |

| Entry 72 | 2026-10-03 · VERIFICATION_RECORD · manifest – |
|---|---|
| governing_provisions | PG-26 P26-18 as amended by CA3-11; CA2-32; PG-15 §12.1 R1; Act R v0.2 item 2 |
| subject | OFTYGR-R1-DISCREPANCY-REGISTER<br>c5dbbe62add332b458e93ce6fb3c710669c43447d1c0ac53466b7d39322a056b<br>OFTYGR-R1-COMPARISON-TABLE<br>a7d2a75297a5585eab608136994fc4bb0aa94e166850cb08f0dff59e7e4ddc4c<br>OFTYGR-R1-COMPARISON-MANIFEST<br>0ea12b3e566c06e766712ccca179114321e13944c1911a876d03dd5238a83c07<br>INITIAL_E1<br>4332df4a6a87c1d8868daa5c67d5ac7594887d862b22a70e929dcf442f832557<br>INITIAL_E2<br>525b11d6540f7380b2495daf98d06999671a399ead99c948a90213617ee27804<br>OPTION-B-CLASS-A-SELECTION<br>d6f2fcaf4bde6a135f430d6942d0ee96a7cc184f28905b6b1b95ea3db9ecc42c<br>OPTION-B-CLASS-B-DUPLICATION<br>82abf2a77d1af7178696882dfeb5da152e6fb2a397ac17e7aa522bce478d9343<br>PG26-M0005<br>6577b69eb31c2ff8524e6db9fd36408740ba3c95d15725a637e1053871aa6c09 |
| committed_or_manifest_sha256 | – |
| prev_entry_sha256 | 2a65f19f897bc3d67bbce50c2e2877ccb337d8a3a2995b54033f048c2bc2720d |
| entry_line_sha256 | 173f1c92579beadedc7db0bfb79e7874c50c300d2e0d1c6b4265075a4092496d |

| Entry 73 | 2026-10-03 · COMMITMENT · manifest – |
|---|---|
| **governing_provisions** | PG-15 §12.1 R2; Act R v0.2 items 3 and 6; D-1 v7 A-08; gatekeeper report 4bd0008f |
| **subject** | OFTYGR-R2-INSTRUMENT-AND-E2-DELIVERY-IDENTITY-RECORD<br>ba2aedfda0871cc897bb45a33f6029726757449dacc88a80de2cf8720c700907<br>OFTYGR-R2-POSITION-INSTRUMENT-MANIFEST<br>d3ba9c9986c5f30450908d2f5325f0317a654ade5e1c3ac9df2b9012ccf28d50<br>OFTYGR-R2-E2-DELIVERY-ZIP<br>516ae2309d3cab3e64bdd5351bd07065adf4f3b68bac5fce7452709ee2abe9f6<br>OFTYGR-R2-E2-WORKBOOK<br>4a4fa0f72e18ab32a750b4f8ba9beca4e4e0648b6ea5523a59492ea2576bf21e<br>OFTYGR-R2-GUIDE-IT<br>c443913344c59f354c1f1d9fa94796ab4e47bdb66b48d95667dbc9477fd205ae<br>OFTYGR-R1-DISCREPANCY-REGISTER<br>c5dbbe62add332b458e93ce6fb3c710669c43447d1c0ac53466b7d39322a056b<br>OFTYGR-R2-E2-LEGGIMI<br>ea993119c590a464224004ffa20939b9a93e9729d8d53df09015bcae1be06f0c<br>OFTYGR-R2-E2-ZIP-INTERNAL-MANIFEST<br>e9e53905380a61d65371a45c83e97168728a1c1bb19d2cdb1755cbf719bdb9c9<br>OFTYGR-R2-E2-BUILD-RECORD<br>1198948db062796dd88c3cdae104ecca3f524d22cd5fbdcb76b0be182e15164a<br>OFTYGR-R2-E2-DELIVERY-FOLDER-MANIFEST<br>de365d5f8f18af4362d495ee6739b6541286df63e47bd654d264b386fc3b4d4d<br>OFTYGR-R2-E2-EMAIL-TEXT<br>49d343e7015b3fcdcd29047d41015ed85f92161fdae7b13072485f6c19791070<br>OFTYGR-R2-E2-PI-INSTRUCTIONS<br>9fc684b07f84ab695c36a720b0d433a8b9a45379764e8721dfb31ea1164cc601 |
| **committed_or_manifest_sha256** | ba2aedfda0871cc897bb45a33f6029726757449dacc88a80de2cf8720c700907 |
| **prev_entry_sha256** | 173f1c92579beadedc7db0bfb79e7874c50c300d2e0d1c6b4265075a4092496d |
| **entry_line_sha256** | 5abaec35cd0676da0b2f8fe82611fc1ccef39dfd56d53c4218b90009a613dc43 |

| Entry 74 | 2026-10-03 · VERIFICATION_RECORD · manifest – |
|---|---|
| **governing_provisions** | PG-26 P26-18 as amended by CA3-11; CA2-32; PG-15 §12.1 R2; Act R v0.2 items 3 and 6 |
| **subject** | OFTYGR-R2-INSTRUMENT-AND-E2-DELIVERY-IDENTITY-RECORD<br>ba2aedfda0871cc897bb45a33f6029726757449dacc88a80de2cf8720c700907<br>OFTYGR-R2-POSITION-INSTRUMENT-MANIFEST<br>d3ba9c9986c5f30450908d2f5325f0317a654ade5e1c3ac9df2b9012ccf28d50<br>OFTYGR-R2-E2-DELIVERY-ZIP<br>516ae2309d3cab3e64bdd5351bd07065adf4f3b68bac5fce7452709ee2abe9f6<br>OFTYGR-R2-E2-WORKBOOK<br>4a4fa0f72e18ab32a750b4f8ba9beca4e4e0648b6ea5523a59492ea2576bf21e<br>OFTYGR-R2-GUIDE-IT<br>c443913344c59f354c1f1d9fa94796ab4e47bdb66b48d95667dbc9477fd205ae<br>OFTYGR-R1-DISCREPANCY-REGISTER<br>c5dbbe62add332b458e93ce6fb3c710669c43447d1c0ac53466b7d39322a056b<br>OFTYGR-R2-E2-LEGGIMI<br>ea993119c590a464224004ffa20939b9a93e9729d8d53df09015bcae1be06f0c<br>OFTYGR-R2-E2-ZIP-INTERNAL-MANIFEST<br>e9e53905380a61d65371a45c83e97168728a1c1bb19d2cdb1755cbf719bdb9c9<br>OFTYGR-R2-E2-BUILD-RECORD<br>1198948db062796dd88c3cdae104ecca3f524d22cd5fbdcb76b0be182e15164a<br>OFTYGR-R2-E2-DELIVERY-FOLDER-MANIFEST<br>de365d5f8f18af4362d495ee6739b6541286df63e47bd654d264b386fc3b4d4d<br>OFTYGR-R2-E2-EMAIL-TEXT<br>49d343e7015b3fcdcd29047d41015ed85f92161fdae7b13072485f6c19791070<br>OFTYGR-R2-E2-PI-INSTRUCTIONS<br>9fc684b07f84ab695c36a720b0d433a8b9a45379764e8721dfb31ea1164cc601 |
| **committed_or_manifest_sha256** | – |
| **prev_entry_sha256** | 5abaec35cd0676da0b2f8fe82611fc1ccef39dfd56d53c4218b90009a613dc43 |
| **entry_line_sha256** | 1736c35bed0803c9f3a7a39acc05a09fe9bd7dc4cfa06da53c09b9c7bca1c438 |

| **Entry 75** | 2026-10-04 · COMMITMENT · manifest – |
| --- | --- |
| **governing_provisions** | PG-15 §12.1 R2; Act R v0.2 item 3; D-1 v7 A-07/A-08 (entry 66, verified 69); M0005 LOCK_LT2 6577b69e (entries 67-68, verified 70); R1 register entry 71 (verified 72); CA2-32 |
| **subject** | OFTYGR-R2-E1-POSITION-SEAL-MANIFEST<br>`5a5ea335971c6e5fc7abfbcf36ce93034c2f548e3a1537e75cc609241ee9fa30`<br>OFTYGR-R2-E1-POSITION-SET<br>`b757449e79d17ac0a824b126e9de4ef92dea0b641fca217c6f10313de4991268`<br>OFTYGR-R2-E1-ANSWER-PROVENANCE<br>`fb1aa597ae556899f7d96eeb3000cf716c8199550032047a6ef6b6122ec58431`<br>OFTYGR-R2-E1-IMPORTED-WORKBOOK<br>`a054803d90838cddfd2613834afc372735f4fbc8ceeeff782b5ce90f070eccff`<br>OFTYGR-R2-E1-ADOPTION-DECLARATION<br>`e52368fe3dce7c8986ff165c4790c913b145ea7e1092abd993d4869ce5680885`<br>OFTYGR-R2-E1-CONFIRMED-ANSWERS<br>`a263b082efda17814e4151307d1b938350d4ab68b7e4fd4e69292a6e02dbad92`<br>OFTYGR-R2-E1-DEVIATION-RECORD-34<br>`6715a548ff41c3eae59b9ab80c27c0796d5f3d71b54ff49c0585bfa3be6504d1`<br>OFTYGR-R2-E1-PI-APPROVAL-RECORD<br>`cc2be17d8708930664adebf43595b5939bb624dda23d8cd606f91d6cf62cf61a`<br>OFTYGR-R2-E1-PI-CONFIRMATION-RECORD<br>`268a0260f11b1584cf2b070b884140939546b9afd4728b5d3b58f359be531e2a`<br>OFTYGR-R2-E1-A07-BASE-DRAFT<br>`13286d887f5a9cfc5f25c8d3856f67bc52f9bd0fece6f752ea65c5b65ae919d0`<br>OFTYGR-R1-DISCREPANCY-REGISTER<br>`c5dbbe62add332b458e93ce6fb3c710669c43447d1c0ac53466b7d39322a056b` |
| **committed_or_manifest_sha256** | `5a5ea335971c6e5fc7abfbcf36ce93034c2f548e3a1537e75cc609241ee9fa30` |
| **prev_entry_sha256** | `1736c35bed0803c9f3a7a39acc05a09fe9bd7dc4cfa06da53c09b9c7bca1c438` |
| **entry_line_sha256** | `5bb997bc73459bab941bc9aa4be5295c8f61892b35897b596e8bbd0e9a042cf1` |

| **Entry 76** | 2026-10-04 · VERIFICATION_RECORD · manifest – |
| --- | --- |
| **governing_provisions** | PG-15 §12.1 R2; CA2-32; M0005 §9 / X-1; Codex E1 R2 seal and entry75 check v1.0; execution addendum v1.0 |
| **subject** | OFTYGR-R2-E1-POSITION-SEAL-MANIFEST<br>`5a5ea335971c6e5fc7abfbcf36ce93034c2f548e3a1537e75cc609241ee9fa30`<br>OFTYGR-R2-E1-POSITION-SET<br>`b757449e79d17ac0a824b126e9de4ef92dea0b641fca217c6f10313de4991268`<br>OFTYGR-R2-E1-ANSWER-PROVENANCE<br>`fb1aa597ae556899f7d96eeb3000cf716c8199550032047a6ef6b6122ec58431`<br>OFTYGR-R2-E1-IMPORTED-WORKBOOK<br>`a054803d90838cddfd2613834afc372735f4fbc8ceeeff782b5ce90f070eccff`<br>OFTYGR-R2-E1-ADOPTION-DECLARATION<br>`e52368fe3dce7c8986ff165c4790c913b145ea7e1092abd993d4869ce5680885`<br>OFTYGR-R2-E1-CONFIRMED-ANSWERS<br>`a263b082efda17814e4151307d1b938350d4ab68b7e4fd4e69292a6e02dbad92`<br>OFTYGR-R2-E1-DEVIATION-RECORD-34<br>`6715a548ff41c3eae59b9ab80c27c0796d5f3d71b54ff49c0585bfa3be6504d1`<br>OFTYGR-R2-E1-PI-APPROVAL-RECORD<br>`cc2be17d8708930664adebf43595b5939bb624dda23d8cd606f91d6cf62cf61a`<br>OFTYGR-R2-E1-PI-CONFIRMATION-RECORD<br>`268a0260f11b1584cf2b070b884140939546b9afd4728b5d3b58f359be531e2a`<br>OFTYGR-R2-E1-A07-BASE-DRAFT<br>`13286d887f5a9cfc5f25c8d3856f67bc52f9bd0fece6f752ea65c5b65ae919d0`<br>OFTYGR-R1-DISCREPANCY-REGISTER<br>`c5dbbe62add332b458e93ce6fb3c710669c43447d1c0ac53466b7d39322a056b`<br>OFTYGR-R2-E1-CODEX-CHECK<br>`654dc634cdd05832126060144111873fc51f987bd7b1c554e6b826fa1c6fe9dc`<br>OFTYGR-R2-E1-EXECUTION-OPERATOR-ADDENDUM<br>`3a33205a3986dce76388f7fc204de34f2a2d6b474f06c0150124cdabe18e99a2` |
| **committed_or_manifest_sha256** | – |
| **prev_entry_sha256** | `5bb997bc73459bab941bc9aa4be5295c8f61892b35897b596e8bbd0e9a042cf1` |
| **entry_line_sha256** | `a15828f6acd69b4577f5b43a5249f91ef111108cd7ad699c920b7bde1bee0a57` |

| **Entry 77** | 2026-10-04 · COMMITMENT · manifest – |
| --- | --- |
| **governing_provisions** | PG-15 §12.1 R2; CA2-32; Act R v0.2; M0005 §9 / X-1; D-1 v7 mechanical function A-08; actual operator Codex on PI instruction |
| **subject** | OFTYGR-R2-E2-POSITION-SEAL-MANIFEST<br>`008fce5c17536db44769586ea6aa3d6f96ad5af65806a7dde30394f2df7d5388`<br>OFTYGR-R2-E2-POSITION-SET<br>`c603588abb416bcbab6293da2a44e9eebdaf1d16e1dc6feb82b5b89c5c2849df`<br>OFTYGR-R2-E2-ANSWER-PROVENANCE<br>`fcab1ccac53d2958b020e7cabed9a3bb8d84e75f9bb491aba83e778f61afe99e`<br>OFTYGR-R2-E2-CORRECTED-WORKBOOK<br>`52f0fd8729e349643e53eb1a2cbd06466d5f3c6749d8fc6b1d4f6db9855fc747`<br>OFTYGR-R2-E2-PI-SUPPLIED-CORRECTION-MESSAGE<br>`df09dfdb6584da19a6eab7bbc92a499df1d6e25d8aac26c8b5726f9a91d2e60f`<br>OFTYGR-R2-E2-RETURN-RECEIPT<br>`982b86e4797fafe6fd91deaab1e911d12ab494cc975caf85460fbdb7d50610a8`<br>OFTYGR-R2-E2-CORRECTION-COMPARISON<br>`77261fef29988d267f8e234f34519d23e654a96fbcff0ec1143b31f71974c4ea`<br>OFTYGR-R1-DISCREPANCY-REGISTER<br>`c5dbbe62add332b458e93ce6fb3c710669c43447d1c0ac53466b7d39322a056b` |
| **committed_or_manifest_sha256** | `008fce5c17536db44769586ea6aa3d6f96ad5af65806a7dde30394f2df7d5388` |
| **prev_entry_sha256** | `a15828f6acd69b4577f5b43a5249f91ef111108cd7ad699c920b7bde1bee0a57` |
| **entry_line_sha256** | `bf3836bdd9a40198c49fcaa06f4400a9d9e02dc8d261be5c42e50a8cd7721407` |

| **Entry 78** | 2026-10-04 · VERIFICATION_RECORD · manifest – |
| --- | --- |
| **governing_provisions** | PG-15 §12.1 R2; CA2-32; M0005 §9 / X-1; Codex corrected E2 seal/entry77 report v1.0 |
| **subject** | OFTYGR-R2-E2-POSITION-SEAL-MANIFEST<br>`008fce5c17536db44769586ea6aa3d6f96ad5af65806a7dde30394f2df7d5388`<br>OFTYGR-R2-E2-POSITION-SET<br>`c603588abb416bcbab6293da2a44e9eebdaf1d16e1dc6feb82b5b89c5c2849df`<br>OFTYGR-R2-E2-ANSWER-PROVENANCE<br>`fcab1ccac53d2958b020e7cabed9a3bb8d84e75f9bb491aba83e778f61afe99e`<br>OFTYGR-R2-E2-CORRECTED-WORKBOOK<br>`52f0fd8729e349643e53eb1a2cbd06466d5f3c6749d8fc6b1d4f6db9855fc747`<br>OFTYGR-R2-E2-PI-SUPPLIED-CORRECTION-MESSAGE<br>`df09dfdb6584da19a6eab7bbc92a499df1d6e25d8aac26c8b5726f9a91d2e60f`<br>OFTYGR-R2-E2-RETURN-RECEIPT<br>`982b86e4797fafe6fd91deaab1e911d12ab494cc975caf85460fbdb7d50610a8`<br>OFTYGR-R2-E2-CORRECTION-COMPARISON<br>`77261fef29988d267f8e234f34519d23e654a96fbcff0ec1143b31f71974c4ea`<br>OFTYGR-R1-DISCREPANCY-REGISTER<br>`c5dbbe62add332b458e93ce6fb3c710669c43447d1c0ac53466b7d39322a056b`<br>OFTYGR-R2-E2-CODEX-CHECK<br>`45dc5aaa9bea04821a62a9b92f2bc0e3e2725853ab0a325f93a4438c79c91237` |
| **committed_or_manifest_sha256** | – |
| **prev_entry_sha256** | `bf3836bdd9a40198c49fcaa06f4400a9d9e02dc8d261be5c42e50a8cd7721407` |
| **entry_line_sha256** | `2b4fe9a4c2799435cf8ba6e461a9a2010fd1ce59f84dee75f07f1bdc1e604f97` |

| Entry 79 | 2026-10-04 · COMMITMENT · manifest – |
|---|---|
| **governing_provisions** | PG-15 §12.1 R3-R5; PG-15 §12.2; Act R v0.2 item 4; D-1 v7 A-08 part (c) I-C (entry 66, verified 69); M0005 LOCK_LT2 6577b69e (entries 67-68, verified 70); R1 register entry 71 (verified 72); E1 position seal entry 75 (verified 76); E2 position seal entry 77 (verified 78); CA2-32 |
| **subject** | OFTYGR-R3R5-OUTCOME-RECORD<br>f20b1bcfea6e9982b392a1dd728ab922f6e014440feb045787edb6e41fd5bbf0<br>OFTYGR-R3R5-OUTCOME-TRANSCRIPTION-PROVENANCE<br>111eeda6651ade19a6d6535bd4cae16de15e8316db5b93d360b90ef636f7ab69<br>OFTYGR-R3R5-OUTCOME-TRANSCRIPTION-TOOL<br>bf835e9058f338ea0d5a6d80111edaacff8c78eb835b3d6b9ac375e2262535fb<br>OFTYGR-R3-SESSION-SHEET-CORR3<br>81a6d4fcdb077c73d5e7d740ce44ab1b94b2c7a87e59997ac639419541d8b3bc<br>OFTYGR-R3-SOURCE-ZIP<br>f40d778106575276c6960627aa75e83c6e76a53e9d09167df72d934c69897ea1<br>OFTYGR-R3-TECHNICAL-NOTE<br>34709e4deab1861c01b0b1ed4f36cd256264a850313f3c9cc2d85df82f301913<br>OFTYGR-R3-TIMESTAMP-MAPPING<br>67949d0283e8291b2ce9b41ac625a84c7dd634e40b8fa933d70ae1e0e8f5a85c<br>OFTYGR-R3-PI-CONFIRMATION-AND-CLOSURE-RECORD<br>9c18019c3f9cd2134b007ec84c60ae980a2db4ce3a130bbf616cc7afc40886ed<br>OFTYGR-R1-DISCREPANCY-REGISTER<br>c5dbbe62add332b458e93ce6fb3c710669c43447d1c0ac53466b7d39322a056b<br>OFTYGR-R2-E1-POSITION-SEAL-MANIFEST<br>5a5ea335971c6e5fc7abfbcf36ce93034c2f548e3a1537e75cc609241ee9fa30<br>OFTYGR-R2-E2-POSITION-SEAL-MANIFEST<br>008fce5c17536db44769586ea6aa3d6f96ad5af65806a7dde30394f2df7d5388<br>OFTYGR-R3-EXACT-RECORD-JOINT-RECONFIRMATION-PI-TRANSMITTED<br>3f6e9cce713e3fa1cf6fafa2536ffe6777a44e50c792e79c71552ec8c5c576cf<br>OFTYGR-R3R5-TRANSCRIPTION-TOOL-WINDOWS-FIX<br>d13bdee56b0529eca229fa18fcebe1e7d4ca8d69de2b19fa21e27b87531a5db5 |
| **committed_or_manifest_sha256** | f20b1bcfea6e9982b392a1dd728ab922f6e014440feb045787edb6e41fd5bbf0 |
| **prev_entry_sha256** | 2b4fe9a4c2799435cf8ba6e461a9a2010fd1ce59f84dee75f07f1bdc1e604f97 |
| **entry_line_sha256** | 764e7c2e92c82c7c07a7851c068a3064c205c2ac7205090daed33d96969869fa |

| Entry 80 | 2026-10-04 · VERIFICATION_RECORD · manifest – |
|---|---|
| **governing_provisions** | PG-26 P26-18 as amended by CA3-11; CA2-32; PG-15 §12.1 R3-R5; M0005 §9 / X-1; Act R v0.2 item 4; D-1 v7 I-C; Codex R3 concluding check v1.0 (5850869e) |
| **subject** | OFTYGR-R3R5-OUTCOME-RECORD<br>f20b1bcfea6e9982b392a1dd728ab922f6e014440feb045787edb6e41fd5bbf0<br>OFTYGR-R3R5-OUTCOME-TRANSCRIPTION-PROVENANCE<br>111eeda6651ade19a6d6535bd4cae16de15e8316db5b93d360b90ef636f7ab69<br>OFTYGR-R3R5-OUTCOME-TRANSCRIPTION-TOOL<br>bf835e9058f338ea0d5a6d80111edaacff8c78eb835b3d6b9ac375e2262535fb<br>OFTYGR-R3-SESSION-SHEET-CORR3<br>81a6d4fcdb077c73d5e7d740ce44ab1b94b2c7a87e59997ac639419541d8b3bc<br>OFTYGR-R3-SOURCE-ZIP<br>f40d778106575276c6960627aa75e83c6e76a53e9d09167df72d934c69897ea1<br>OFTYGR-R3-TECHNICAL-NOTE<br>34709e4deab1861c01b0b1ed4f36cd256264a850313f3c9cc2d85df82f301913<br>OFTYGR-R3-TIMESTAMP-MAPPING<br>67949d0283e8291b2ce9b41ac625a84c7dd634e40b8fa933d70ae1e0e8f5a85c<br>OFTYGR-R3-PI-CONFIRMATION-AND-CLOSURE-RECORD<br>9c18019c3f9cd2134b007ec84c60ae980a2db4ce3a130bbf616cc7afc40886ed<br>OFTYGR-R1-DISCREPANCY-REGISTER<br>c5dbbe62add332b458e93ce6fb3c710669c43447d1c0ac53466b7d39322a056b<br>OFTYGR-R2-E1-POSITION-SEAL-MANIFEST<br>5a5ea335971c6e5fc7abfbcf36ce93034c2f548e3a1537e75cc609241ee9fa30<br>OFTYGR-R2-E2-POSITION-SEAL-MANIFEST<br>008fce5c17536db44769586ea6aa3d6f96ad5af65806a7dde30394f2df7d5388<br>OFTYGR-R3-EXACT-RECORD-JOINT-RECONFIRMATION-PI-TRANSMITTED<br>3f6e9cce713e3fa1cf6fafa2536ffe6777a44e50c792e79c71552ec8c5c576cf<br>OFTYGR-R3R5-TRANSCRIPTION-TOOL-WINDOWS-FIX<br>d13bdee56b0529eca229fa18fcebe1e7d4ca8d69de2b19fa21e27b87531a5db5 |
| **committed_or_manifest_sha256** | – |
| **prev_entry_sha256** | 764e7c2e92c82c7c07a7851c068a3064c205c2ac7205090daed33d96969869fa |
| **entry_line_sha256** | cdac8cf32695ed681f36d457ec38cc482e125db1c7d00c64ab8f09f2b2d4908a |

| Entry 81 | 2026-10-04 · COMMITMENT · manifest – |
|---|---|
| **governing_provisions** | PG-15 §12.1 R6; PG-15 §13; Act R v0.2 item 5; D-1 v7 A-08 part (b) (entry 66, verified 69); M0005 LOCK_LT2 6577b69e (entries 67-68, verified 70); R1 register entry 71 (verified 72); E1 position seal entry 75 (verified 76); E2 position seal entry 77 (verified 78); R4/R5 outcome record entry 79 (verification entry 80); CA2-32 |
| **subject** | OFTYGR-R6-CANONICAL-BUILD-MANIFEST<br>`c9fb08cc6ca5e4b5bd0f9e3541187d17311720ac7cb4eae22edbd5d22786d7bb`<br>OFTYGR-R6-CANONICAL-RECORD<br>`8fe5ec6fcee28c7eeb648a4c04602bc1efd11de597fc11b04fb79446771217e6`<br>OFTYGR-R6-RESIDUAL-REGISTER<br>`18f0cfc1a38e196343ef90cea91d90abb9bfa13e7ecafde110caaddfba34ae6a`<br>OFTYGR-R6-PENDING-DEPENDENCIES<br>`2062a22c99f73170270ceee3e1f831078afa744133b062665493a62bfe84ff9f`<br>OFTYGR-R6-BLOCKING-CONDITIONS<br>`f0720a1768fd6d1c5837b032ff063c12612b2369a4f42820e8ac2a45191a22fd`<br>OFTYGR-R6-PROCEDURAL-TRIGGERS<br>`cdba968de58742e14b88b6e0cebf5ccb8e1abf1f5a0e6d1eef16217d454ded88`<br>OFTYGR-R3R5-OUTCOME-RECORD<br>`f20b1bcfea6e9982b392a1dd728ab922f6e014440feb045787edb6e41fd5bbf0`<br>OFTYGR-R3R5-OUTCOME-TRANSCRIPTION-PROVENANCE<br>`111eeda6651ade19a6d6535bd4cae16de15e8316db5b93d360b90ef636f7ab69`<br>OFTYGR-R2-E1-POSITION-SEAL-MANIFEST<br>`5a5ea335971c6e5fc7abfbcf36ce93034c2f548e3a1537e75cc609241ee9fa30`<br>OFTYGR-R2-E2-POSITION-SEAL-MANIFEST<br>`008fce5c17536db44769586ea6aa3d6f96ad5af65806a7dde30394f2df7d5388`<br>OFTYGR-R1-DISCREPANCY-REGISTER<br>`c5dbbe62add332b458e93ce6fb3c710669c43447d1c0ac53466b7d39322a056b` |
| **committed_or_manifest_sha256** | `c9fb08cc6ca5e4b5bd0f9e3541187d17311720ac7cb4eae22edbd5d22786d7bb` |
| **prev_entry_sha256** | `cdac8cf32695ed681f36d457ec38cc482e125db1c7d00c64ab8f09f2b2d4908a` |
| **entry_line_sha256** | `46516925ee0ebf99d3b7bbfa3bc0d9599d47f256cddd0545f449b7b41b0b716b` |

| Entry 82 | 2026-10-04 · VERIFICATION_RECORD · manifest – |
|---|---|
| **governing_provisions** | PG-15 §12.1 R6; PG-15 §13; Act R v0.2 item 5; D-1 v7 A-08 part (b) (entry 66, verified 69); M0005 LOCK_LT2 6577b69e (entries 67-68, verified 70); R1 register entry 71 (verified 72); E1 position seal entry 75 (verified 76); E2 position seal entry 77 (verified 78); R4/R5 outcome record entry 79 (verification entry 80); CA2-32; M0005 §9 / X-1; Codex entries 80-82 guarded execution ruling v1.0 (65a4b290) |
| **subject** | OFTYGR-R6-CANONICAL-BUILD-MANIFEST<br>`c9fb08cc6ca5e4b5bd0f9e3541187d17311720ac7cb4eae22edbd5d22786d7bb`<br>OFTYGR-R6-CANONICAL-RECORD<br>`8fe5ec6fcee28c7eeb648a4c04602bc1efd11de597fc11b04fb79446771217e6`<br>OFTYGR-R6-RESIDUAL-REGISTER<br>`18f0cfc1a38e196343ef90cea91d90abb9bfa13e7ecafde110caaddfba34ae6a`<br>OFTYGR-R6-PENDING-DEPENDENCIES<br>`2062a22c99f73170270ceee3e1f831078afa744133b062665493a62bfe84ff9f`<br>OFTYGR-R6-BLOCKING-CONDITIONS<br>`f0720a1768fd6d1c5837b032ff063c12612b2369a4f42820e8ac2a45191a22fd`<br>OFTYGR-R6-PROCEDURAL-TRIGGERS<br>`cdba968de58742e14b88b6e0cebf5ccb8e1abf1f5a0e6d1eef16217d454ded88`<br>OFTYGR-R3R5-OUTCOME-RECORD<br>`f20b1bcfea6e9982b392a1dd728ab922f6e014440feb045787edb6e41fd5bbf0`<br>OFTYGR-R3R5-OUTCOME-TRANSCRIPTION-PROVENANCE<br>`111eeda6651ade19a6d6535bd4cae16de15e8316db5b93d360b90ef636f7ab69`<br>OFTYGR-R2-E1-POSITION-SEAL-MANIFEST<br>`5a5ea335971c6e5fc7abfbcf36ce93034c2f548e3a1537e75cc609241ee9fa30`<br>OFTYGR-R2-E2-POSITION-SEAL-MANIFEST<br>`008fce5c17536db44769586ea6aa3d6f96ad5af65806a7dde30394f2df7d5388`<br>OFTYGR-R1-DISCREPANCY-REGISTER<br>`c5dbbe62add332b458e93ce6fb3c710669c43447d1c0ac53466b7d39322a056b` |
| **committed_or_manifest_sha256** | – |
| **prev_entry_sha256** | `46516925ee0ebf99d3b7bbfa3bc0d9599d47f256cddd0545f449b7b41b0b716b` |
| **entry_line_sha256** | `b9c49a8dd9777c96a5a838bbc0c9d9eb5b827fd5707919b12c9fb4598ce04ae0` |

| Entry 83 | 2026-10-04 · ROLE_RECORD · manifest – |
|---|---|
| **governing_provisions** | P32-14; P32-13; P32-02; PG-15 sec. 12.1; PG-15 sec. 11.2; P26-14 |
| **subject** | PG32-D1<br>`d824446d7b94fc31b1b4d47af8b3f17bcb0ad7fd5060e7106f78cb9562ecaca1`<br>P13-DRIVER-A08_P13_N08A_Form_To_Append_Attempt<br>`ddad983752e95b442171f44327161823d8275372f1d831a0fb40737451232aa0`<br>GATEKEEPER-REPORT-P13-v1.1-and-D1v8<br>`e1f303a9fecb63f90fe2b4820116fb18fb2a66aad54e140cf4ea65312f32ca41`<br>PG32-D1-v8-APPROVAL-AND-FREEZE-ACT<br>`bfe46ec2887f2fcb4a992e1e973e37a1201d96e407c5de5d223b29d6512d48df` |
| **committed_or_manifest_sha256** | – |
| **prev_entry_sha256** | `b9c49a8dd9777c96a5a838bbc0c9d9eb5b827fd5707919b12c9fb4598ce04ae0` |
| **entry_line_sha256** | `c6d640777ef49b8fecbcce6d6c0df4262397ffa1e1e79dea3e511fd490e1278b` |

| Entry 84 | 2026-10-04 · VERIFICATION_RECORD · manifest – |
|---|---|
| **governing_provisions** | P26-18; P32-14; P32-13 |
| **subject** | PG32-D1<br>d824446d7b94fc31b1b4d47af8b3f17bcb0ad7fd5060e7106f78cb9562ecaca1 |
| **committed_or_manifest_sha256** | – |
| **prev_entry_sha256** | c6d640777ef49b8fecbcce6d6c0df4262397ffa1e1e79dea3e511fd490e1278b |
| **entry_line_sha256** | 21b8570863d939bfa0d5d94835b842a8363212716f79e8654b4aa1934e033fbc |

| Entry 85 | 2026-10-04 · COMMITMENT · manifest – |
|---|---|
| **governing_provisions** | PG-15 §14.6; Act R v0.2 item 7 (3a20c4da); D-1 v8 A-08 part (d) (entry 83, verified 84); R6 canonical build (entry 81, verified 82); N08A records FAM-0001 / FAM-0002 (entries 48-49, verified 50-51); CA2-32 |
| **subject** | OFTYGR-P13-CHAIN-MANIFEST-FAM-0001<br>5271218200ec5fa4f09bcf8a416156c017d07ae53053ad6d4f86ffb4f3fa9568<br>OFTYGR-P13-N08A-RECORD-FAM-0001<br>9e0043981efe6b4fc231a3a6970e558c3f4c1b434ea3d342642d66df900a1aee<br>OFTYGR-P13-N08A-RECORD-FAM-0001<br>3976e84ae112db5b227e4c723f4875d3253c3a13e8ee0560153e24fa2cdb1baf<br>OFTYGR-P13-N08A-RECORD-FAM-0001<br>167051db0c9017d17245573ff19a724022b6209bcae983c9037269afb1ce73b5<br>OFTYGR-P13-N08A-RECORD-FAM-0001<br>a6c04ad197c0097ca8766e3b746b6d69e6fcef610db1b518a06f44e385666af8<br>OFTYGR-P13-N08A-RECORD-FAM-0001<br>9df78dfe8ecf0b2985c813574c1e94296630fae7ce9e4942af27a81d1c5f0595<br>OFTYGR-P13-N08A-RECORD-FAM-0001<br>b5e426e84628f7c6be60474162516bdebc477bf8d479fe9f2c1ba0acdec4d2ac<br>OFTYGR-P13-N08A-RECORD-FAM-0001<br>58097b5e1c7759acc34abb38542e1318234411b0b575b59713a91ff5d59cc488<br>OFTYGR-P13-N08A-RECORD-FAM-0001<br>f7d214495336ec9762a6222ac9a907f414a08364a929d0fdcd4b852873ee9ee8<br>OFTYGR-P13-N08A-RECORD-FAM-0001<br>ec26ba1554425ead67f68e7b784d8eae8f31d762fcf84d12bb95456e2ea0118d<br>OFTYGR-P13-N08A-RECORD-FAM-0001<br>46838ec791c8641c1b1c0d331d6013e91d2ab757092dd7fbbe4b019738a33128<br>OFTYGR-P13-N08A-RECORD-FAM-0001<br>b5e620d920dd9a650816667d5ffdf0a507c2e3f37c7ec1b7a268ba797bb75e5f<br>OFTYGR-P13-N08A-RECORD-FAM-0001<br>1a5e771785a879935afdbda0e4b976d416b79be3cf7eca60ccb1e7bc72f047d2<br>OFTYGR-P13-N08A-RECORD-FAM-0001<br>cca1ed5bf6ca54a24f473bf670923de8b75edb6677928c8c28e30515fcd50ead<br>OFTYGR-P13-N08A-RECORD-FAM-0001<br>c6b990e5afed490a2f87647b427e479586c8b936a488feca8df260d4ed7b4728<br>OFTYGR-P13-N08A-RECORD-FAM-0001<br>ba1944874498152939c3ca4f016801b111e1515797eb50eef795cafeeddab6ad<br>OFTYGR-P13-N08A-RECORD-FAM-0001<br>ebcd3ffbd5e125f2ed3bad8592879011336b90cbdbdab5a09f97cba2f55a84c8<br>OFTYGR-P13-N08A-RECORD-FAM-0001<br>97eb51f0a6b85d9812e4a386918351dbcd2c340d49174a8ae7a4e3831d95b882<br>OFTYGR-P13-N08A-RECORD-FAM-0001<br>92711b70c3d0756c1886e51e49a48c2038c266a0057aebe00d20fcc4786047c3<br>OFTYGR-P13-N08A-RECORD-FAM-0001<br>8e06d2c67bfc333800342d22c7de6dc07cbc2beb65223f46a3aab13a65695429<br>OFTYGR-P13-N08A-RECORD-FAM-0001<br>d6091fe1e7185f69b1be318dbf8e2d7b67aad48fa6ebb5a32b3b43ea400342f8<br>OFTYGR-P13-N08A-RECORD-FAM-0001<br>829a3b73fd30a3095b52d7339e4401921e9b591a0765f0a8bf6ac805ec408c3b<br>OFTYGR-P13-N08A-RECORD-FAM-0001<br>370145c2b92422f78f1b561f5c395376a202b90d92ff28a69ebec63602a69a6f<br>OFTYGR-P13-N08A-RECORD-FAM-0001<br>3e752e98b79cebca139e79696d594a402587e43798b40b60a554868d6d124126<br>OFTYGR-P13-N08A-RECORD-FAM-0001<br>bd7240e91b1d4151658eb1dbb77bec0091717736091dc160d6c1e57670a19d3e<br>OFTYGR-P13-N08A-RECORD-FAM-0001<br>70d7f8c1a9e2e1e6e34b8723d23b31a373753466659f5abe5b97988be969bf6d<br>OFTYGR-P13-N08A-RECORD-FAM-0001<br>030cbf22e039ce3ab287898367d1b4ea245c4506751ef3d8752d2f4ddc1b5935<br>OFTYGR-P13-N08A-RECORD-FAM-0001<br>1f23ced9175f10e645d39e3ff9a237c6ad4ea16d98839b5d1a9d9265ade38926<br>OFTYGR-P13-N08A-RECORD-FAM-0001<br>f5dc34bc9adcb63454ca0d3cfb301e91a9cfe039f4f63fd6051d13e61607cfcf<br>OFTYGR-P13-N08A-RECORD-FAM-0001<br>bf8db711cd3b8d1ce818e361bbf55baefd447a9627282f5913718db08ee1f1f9 |
| **committed_or_manifest_sha256** | 5271218200ec5fa4f09bcf8a416156c017d07ae53053ad6d4f86ffb4f3fa9568 |
| **prev_entry_sha256** | 21b8570863d939bfa0d5d94835b842a8363212716f79e8654b4aa1934e033fbc |
| **entry_line_sha256** | b238c10dd0023569fe91d7891914e46889e6e455d0517cc851a4b931a0102acb |

| Entry 86 | 2026-10-04 · COMMITMENT · manifest – |
|---|---|
| **governing_provisions** | PG-15 §14.6; Act R v0.2 item 7 (3a20c4da); D-1 v8 A-08 part (d) (entry 83, verified 84); R6 canonical build (entry 81, verified 82); N08A records FAM-0001 / FAM-0002 (entries 48-49, verified 50-51); CA2-32 |
| **subject** | OFTYGR-P13-CHAIN-MANIFEST-FAM-0002<br>`ff71f554a61013696867f23fb9d6719bd53068e496369f34123651d008913f5c`<br>OFTYGR-P13-N08A-RECORD-FAM-0002<br>`bedb84aa4067f5a5aa1b205abcd1cabf6d0a3b618c7ce972df25ccb1c026e06d`<br>OFTYGR-P13-N08A-RECORD-FAM-0002<br>`3fa745f634e5075abf53e6eed4225feb663d21aaee53e210beb8ff21dd6b7bad`<br>OFTYGR-P13-N08A-RECORD-FAM-0002<br>`9df076494f5b56672494519d69b83602e2ddfe8cfc7707f3f4edd27db75ddfd3`<br>OFTYGR-P13-N08A-RECORD-FAM-0002<br>`53966aa5836d1f0f8e65732f482fc6e76a89962b15e3dcbd6206deb1b2a2db2a`<br>OFTYGR-P13-N08A-RECORD-FAM-0002<br>`7912ca95a8e4bafcfee47effe702fe8890d22962dbb2da3c77c32340d89f03ca`<br>OFTYGR-P13-N08A-RECORD-FAM-0002<br>`8831dd5c330d1eb25ef943a4efd7e7499f8b38fbbe1e421492ed9d242c46e3a3`<br>OFTYGR-P13-N08A-RECORD-FAM-0002<br>`a90be2d3bca690dfe3c04ab3d59c566c87df874a99bad01274c5f2f7d4991b8f`<br>OFTYGR-P13-N08A-RECORD-FAM-0002<br>`141d7ab1bca265b5ec36d7fb0aea5707c167316a86343bea6f8f9f216e584c40`<br>OFTYGR-P13-N08A-RECORD-FAM-0002<br>`ad288d591612ca9415659c10884d6b87b12886de8305000b27e6d32ee7ffbd6a`<br>OFTYGR-P13-N08A-RECORD-FAM-0002<br>`97f14f1538a4add2017c02373a5ef80a5dc45be1c72aa163e8b31650008b4b29`<br>OFTYGR-P13-N08A-RECORD-FAM-0002<br>`51462d423c8ecfcacec2ab17006d5c1e7804230b6fd7c2e1c68d5e266d271712`<br>OFTYGR-P13-N08A-RECORD-FAM-0002<br>`c0c20fe93e481666099c3760e2c98287126402c7a472253317a10204d9f3d2d2`<br>OFTYGR-P13-N08A-RECORD-FAM-0002<br>`b9a3c68bd5ff1b3f57871bff8a9a404a2ec81c494cd56717f07e556e5adefe1a`<br>OFTYGR-P13-N08A-RECORD-FAM-0002<br>`a9d39b3a7a52bac88030bbd993d0ecda420d602ababf0b3f33bfb071ee0a7fd5`<br>OFTYGR-P13-N08A-RECORD-FAM-0002<br>`5061743a93da667e2d34e4a2e8884518bdff9736cc07585005c70c38859b726b`<br>OFTYGR-P13-N08A-RECORD-FAM-0002<br>`6c4d6febdf2ca17064e7a1cdaf1bb3a149a97f9a08dddc8fbb9fb486dd887f78`<br>OFTYGR-P13-N08A-RECORD-FAM-0002<br>`72a44455f9fefc2a0086556629e20291c440ecca26d336d832c7c6b3542c2667`<br>OFTYGR-P13-N08A-RECORD-FAM-0002<br>`f8c88356346e85300dbc95e2e27b06cbb4c93782b35f0a0130a8d808bf07b603`<br>OFTYGR-P13-N08A-RECORD-FAM-0002<br>`4e16000da250577344b4dce4e3d2fd4644f65c3c90587e73be296b275d70fa57`<br>OFTYGR-P13-N08A-RECORD-FAM-0002<br>`318ff168fb392e22fd968e07e8ec820dcae9cebbfca07e6a336214c5ed1b574a`<br>OFTYGR-P13-N08A-RECORD-FAM-0002<br>`c5172e80cb744fefa5b7e980b672a03d8e54f096aa1564c643a7865ade8886fa`<br>OFTYGR-P13-N08A-RECORD-FAM-0002<br>`776aa6e0917e5f82c1899012538b7b773cca3c39e88481eaa43551ed1ff70555`<br>OFTYGR-P13-N08A-RECORD-FAM-0002<br>`84855f912282645a26554a611d285bab8d15a556659b65c20f0806209a153a30`<br>OFTYGR-P13-N08A-RECORD-FAM-0002<br>`fd6a828d4d70c1b3e4614f6e866317e19276af9567c01df8ce2dc555b5360bb8`<br>OFTYGR-P13-N08A-RECORD-FAM-0002<br>`bb523fe138bace31ff902702d88af3f9f4809896985ded0e62b98b8b8280a23a`<br>OFTYGR-P13-N08A-RECORD-FAM-0002<br>`1580499ce91d66cb725ca8a9debffc490fade18d9c35a82d04274882a7a3d313`<br>OFTYGR-P13-N08A-RECORD-FAM-0002<br>`864a88777c778cab5f7fea835589543bc726db058d6d9cf65f4c12d15f578014` |
| **committed_or_manifest_sha256** | `ff71f554a61013696867f23fb9d6719bd53068e496369f34123651d008913f5c` |
| **prev_entry_sha256** | `b238c10dd0023569fe91d7891914e46889e6e455d0517cc851a4b931a0102acb` |
| **entry_line_sha256** | `6868c21eedda0b88828b969b353f79d9f9c4c450e37699df9b0ed9c2db5dce34` |

| Entry 87 | 2026-10-04 · VERIFICATION_RECORD · manifest – |
| --- | --- |
| **governing_provisions** | P26-18; PG-15 §14.6 |
| **subject** | OFTYGR-P13-CHAIN-MANIFEST-FAM-0001<br>5271218200ec5fa4f09bcf8a416156c017d07ae53053ad6d4f86ffb4f3fa9568<br>OFTYGR-P13-N08A-RECORD-FAM-0001<br>9e0043981efe6b4fc231a3a6970e558c3f4c1b434ea3d342642d66df900a1aee<br>OFTYGR-P13-N08A-RECORD-FAM-0001<br>3976e84ae112db5b227e4c723f4875d3253c3a13e8ee0560153e24fa2cdb1baf<br>OFTYGR-P13-N08A-RECORD-FAM-0001<br>167051db0c9017d17245573ff19a724022b6209bcae983c9037269afb1ce73b5<br>OFTYGR-P13-N08A-RECORD-FAM-0001<br>a6c04ad197c0097ca8766e3b746b6d69e6fcef610db1b518a06f44e385666af8<br>OFTYGR-P13-N08A-RECORD-FAM-0001<br>9df78dfe8ecf0b2985c813574c1e94296630fae7ce9e4942af27a81d1c5f0595<br>OFTYGR-P13-N08A-RECORD-FAM-0001<br>b5e426e84628f7c6be60474162516bdebc477bf8d479fe9f2c1ba0acdec4d2ac<br>OFTYGR-P13-N08A-RECORD-FAM-0001<br>58097b5e1c7759acc34abb38542e1318234411b0b575b59713a91ff5d59cc488<br>OFTYGR-P13-N08A-RECORD-FAM-0001<br>f7d214495336ec9762a6222ac9a907f414a08364a929d0fdcd4b852873ee9ee8<br>OFTYGR-P13-N08A-RECORD-FAM-0001<br>ec26ba1554425ead67f68e7b784d8eae8f31d762fcf84d12bb95456e2ea0118d<br>OFTYGR-P13-N08A-RECORD-FAM-0001<br>46838ec791c8641c1b1c0d331d6013e91d2ab757092dd7fbbe4b019738a33128<br>OFTYGR-P13-N08A-RECORD-FAM-0001<br>b5e620d920dd9a650816667d5ffdf0a507c2e3f37c7ec1b7a268ba797bb75e5f<br>OFTYGR-P13-N08A-RECORD-FAM-0001<br>1a5e771785a879935afdbda0e4b976d416b79be3cf7eca60ccb1e7bc72f047d2<br>OFTYGR-P13-N08A-RECORD-FAM-0001<br>cca1ed5bf6ca54a24f473bf670923de8b75edb6677928c8c28e30515fcd50ead<br>OFTYGR-P13-N08A-RECORD-FAM-0001<br>c6b990e5afed490a2f87647b427e479586c8b936a488feca8df260d4ed7b4728<br>OFTYGR-P13-N08A-RECORD-FAM-0001<br>ba1944874498152939c3ca4f016801b111e1515797eb50eef795cafeeddab6ad<br>OFTYGR-P13-N08A-RECORD-FAM-0001<br>ebcd3ffbd5e125f2ed3bad8592879011336b90cbdbdab5a09f97cba2f55a84c8<br>OFTYGR-P13-N08A-RECORD-FAM-0001<br>97eb51f0a6b85d9812e4a386918351dbcd2c340d49174a8ae7a4e3831d95b882<br>OFTYGR-P13-N08A-RECORD-FAM-0001<br>92711b70c3d0756c1886e51e49a48c2038c266a0057aebe00d20fcc4786047c3<br>OFTYGR-P13-N08A-RECORD-FAM-0001<br>8e06d2c67bfc333800342d22c7de6dc07cbc2beb65223f46a3aab13a65695429<br>OFTYGR-P13-N08A-RECORD-FAM-0001<br>d6091fe1e7185f69b1be318dbf8e2d7b67aad48fa6ebb5a32b3b43ea400342f8<br>OFTYGR-P13-N08A-RECORD-FAM-0001<br>829a3b73fd30a3095b52d7339e4401921e9b591a0765f0a8bf6ac805ec408c3b<br>OFTYGR-P13-N08A-RECORD-FAM-0001<br>370145c2b92422f78f1b561f5c395376a202b90d92ff28a69ebec63602a69a6f<br>OFTYGR-P13-N08A-RECORD-FAM-0001<br>3e752e98b79cebca139e79696d594a402587e43798b40b60a554868d6d124126<br>OFTYGR-P13-N08A-RECORD-FAM-0001<br>bd7240e91b1d4151658eb1dbb77bec0091717736091dc160d6c1e57670a19d3e<br>OFTYGR-P13-N08A-RECORD-FAM-0001<br>70d7f8c1a9e2e1e6e34b8723d23b31a373753466659f5abe5b97988be969bf6d<br>OFTYGR-P13-N08A-RECORD-FAM-0001<br>030cbf22e039ce3ab287898367d1b4ea245c4506751ef3d8752d2f4ddc1b5935<br>OFTYGR-P13-N08A-RECORD-FAM-0001<br>1f23ced9175f10e645d39e3ff9a237c6ad4ea16d98839b5d1a9d9265ade38926<br>OFTYGR-P13-N08A-RECORD-FAM-0001<br>f5dc34bc9adcb63454ca0d3cfb301e91a9cfe039f4f63fd6051d13e61607cfcf<br>OFTYGR-P13-N08A-RECORD-FAM-0001<br>bf8db711cd3b8d1ce818e361bbf55baefd447a9627282f5913718db08ee1f1f9 |
| **committed_or_manifest_sha256** | – |
| **prev_entry_sha256** | 6868c21eedda0b88828b969b353f79d9f9c4c450e37699df9b0ed9c2db5dce34 |
| **entry_line_sha256** | ba16e0cc2bf02015b1e543c3d68630b4977e20a67dedb4a41a8c829e46636dfd |

| **Entry 88** | 2026-10-04 · VERIFICATION_RECORD · manifest – |
|---|---|
| **governing_provisions** | P26-18; PG-15 §14.6 |
| **subject** | OFTYGR-P13-CHAIN-MANIFEST-FAM-0002<br>ff71f554a61013696867f23fb9d6719bd53068e496369f34123651d008913f5c<br>OFTYGR-P13-N08A-RECORD-FAM-0002<br>bedb84aa4067f5a5aa1b205abcd1cabf6d0a3b618c7ce972df25ccb1c026e06d<br>OFTYGR-P13-N08A-RECORD-FAM-0002<br>3fa745f634e5075abf53e6eed4225feb663d21aaee53e210beb8ff21dd6b7bad<br>OFTYGR-P13-N08A-RECORD-FAM-0002<br>9df076494f5b56672494519d69b83602e2ddfe8cfc7707f3f4edd27db75ddfd3<br>OFTYGR-P13-N08A-RECORD-FAM-0002<br>53966aa5836d1f0f8e65732f482fc6e76a89962b15e3dcbd6206deb1b2a2db2a<br>OFTYGR-P13-N08A-RECORD-FAM-0002<br>7912ca95a8e4bafcfee47effe702fe8890d22962dbb2da3c77c32340d89f03ca<br>OFTYGR-P13-N08A-RECORD-FAM-0002<br>8831dd5c330d1eb25ef943a4efd7e7499f8b38fbbe1e421492ed9d242c46e3a3<br>OFTYGR-P13-N08A-RECORD-FAM-0002<br>a90be2d3bca690dfe3c04ab3d59c566c87df874a99bad01274c5f2f7d4991b8f<br>OFTYGR-P13-N08A-RECORD-FAM-0002<br>141d7ab1bca265b5ec36d7fb0aea5707c167316a86343bea6f8f9f216e584c40<br>OFTYGR-P13-N08A-RECORD-FAM-0002<br>ad288d591612ca9415659c10884d6b87b12886de8305000b27e6d32ee7ffbd6a<br>OFTYGR-P13-N08A-RECORD-FAM-0002<br>97f14f1538a4add2017c02373a5ef80a5dc45be1c72aa163e8b31650008b4b29<br>OFTYGR-P13-N08A-RECORD-FAM-0002<br>51462d423c8ecfcacec2ab17006d5c1e7804230b6fd7c2e1c68d5e266d271712<br>OFTYGR-P13-N08A-RECORD-FAM-0002<br>c0c20fe93e481666099c3760e2c98287126402c7a472253317a10204d9f3d2d2<br>OFTYGR-P13-N08A-RECORD-FAM-0002<br>b9a3c68bd5ff1b3f57871bff8a9a404a2ec81c494cd56717f07e556e5adefe1a<br>OFTYGR-P13-N08A-RECORD-FAM-0002<br>a9d39b3a7a52bac88030bbd993d0ecda420d602ababf0b3f33bfb071ee0a7fd5<br>OFTYGR-P13-N08A-RECORD-FAM-0002<br>5061743a93da667e2d34e4a2e8884518bdff9736cc07585005c70c38859b726b<br>OFTYGR-P13-N08A-RECORD-FAM-0002<br>6c4d6febdf2ca17064e7a1cdaf1bb3a149a97f9a08dddc8fbb9fb486dd887f78<br>OFTYGR-P13-N08A-RECORD-FAM-0002<br>72a44455f9fefc2a0086556629e20291c440ecca26d336d832c7c6b3542c2667<br>OFTYGR-P13-N08A-RECORD-FAM-0002<br>f8c88356346e85300dbc95e2e27b06cbb4c93782b35f0a0130a8d808bf07b603<br>OFTYGR-P13-N08A-RECORD-FAM-0002<br>4e16000da250577344b4dce4e3d2fd4644f65c3c90587e73be296b275d70fa57<br>OFTYGR-P13-N08A-RECORD-FAM-0002<br>318ff168fb392e22fd968e07e8ec820dcae9cebbfca07e6a336214c5ed1b574a<br>OFTYGR-P13-N08A-RECORD-FAM-0002<br>c5172e80cb744fefa5b7e980b672a03d8e54f096aa1564c643a7865ade8886fa<br>OFTYGR-P13-N08A-RECORD-FAM-0002<br>776aa6e0917e5f82c1899012538b7b773cca3c39e88481eaa43551ed1ff70555<br>OFTYGR-P13-N08A-RECORD-FAM-0002<br>84855f912282645a26554a611d285bab8d15a556659b65c20f0806209a153a30<br>OFTYGR-P13-N08A-RECORD-FAM-0002<br>fd6a828d4d70c1b3e4614f6e866317e19276af9567c01df8ce2dc555b5360bb8<br>OFTYGR-P13-N08A-RECORD-FAM-0002<br>bb523fe138bace31ff902702d88af3f9f4809896985ded0e62b98b8b8280a23a<br>OFTYGR-P13-N08A-RECORD-FAM-0002<br>1580499ce91d66cb725ca8a9debffc490fade18d9c35a82d04274882a7a3d313<br>OFTYGR-P13-N08A-RECORD-FAM-0002<br>864a88777c778cab5f7fea835589543bc726db058d6d9cf65f4c12d15f578014 |
| **committed_or_manifest_sha256** | – |
| **prev_entry_sha256** | ba16e0cc2bf02015b1e543c3d68630b4977e20a67dedb4a41a8c829e46636dfd |
| **entry_line_sha256** | c585bf22ac3fc094ef299774bd594281f45b69f81d1a96981d7c771323273077 |

| **Entry 89** | 2026-10-04 · FREEZE_ACT · manifest M0006 |
|---|---|
| **governing_provisions** | P26-18; P26-14; P26-02(vi) |
| **subject** | PG26-M0006<br>550273cdba85271ba40072c595b3fd58a0fe8cc7cfcb90d6d6ef1614c1249a75 |
| **committed_or_manifest_sha256** | 550273cdba85271ba40072c595b3fd58a0fe8cc7cfcb90d6d6ef1614c1249a75 |
| **prev_entry_sha256** | c585bf22ac3fc094ef299774bd594281f45b69f81d1a96981d7c771323273077 |
| **entry_line_sha256** | e2d4a3aec1fc93bc73a222b025707be19c0119f5eaed788b44ded3a6e87e3728 |

| **Entry 90** | 2026-10-04 · COMMITMENT · manifest – |
|---|---|
| **governing_provisions** | P26-02(iii) as amended by CA2-10/CA3-10; P26-14 |
| **subject** | PG26-M0006<br>550273cdba85271ba40072c595b3fd58a0fe8cc7cfcb90d6d6ef1614c1249a75 |
| **committed_or_manifest_sha256** | 550273cdba85271ba40072c595b3fd58a0fe8cc7cfcb90d6d6ef1614c1249a75 |
| **prev_entry_sha256** | e2d4a3aec1fc93bc73a222b025707be19c0119f5eaed788b44ded3a6e87e3728 |
| **entry_line_sha256** | f1d708ddde333b93064545e5b01c0292fcc425be8c7a05abd1159b3bb40724d6 |

| Entry 91 | 2026-10-04 · VERIFICATION_RECORD · manifest – |
|---|---|
| **governing_provisions** | P26-18; P26-14; P26-12 |
| **subject** | PG26-M0006<br>`550273cdba85271ba40072c595b3fd58a0fe8cc7cfcb90d6d6ef1614c1249a75` |
| **committed_or_manifest_sha256** | – |
| **prev_entry_sha256** | `f1d708ddde333b93064545e5b01c0292fcc425be8c7a05abd1159b3bb40724d6` |
| **entry_line_sha256** | `0d1f5b87e11f21e0dffabda4f27ac68391bedb3f38200364341af2fe4f1878ed` |

| Entry 92 | 2026-10-04 · ROLE_RECORD · manifest – |
|---|---|
| **governing_provisions** | P32-14; P32-13; P32-02; PG-15 sec. 13.3; PG-11 sec. 11.3; P26-14 |
| **subject** | PG32-D1<br>`f035916d53c2672fbe14df0918e1ea67d81928059428557168a3bc3d55d76cd7`<br>PG26-M0006<br>`550273cdba85271ba40072c595b3fd58a0fe8cc7cfcb90d6d6ef1614c1249a75`<br>GATEKEEPER-REPORT-M0006-v0.3-and-D1v9<br>`f9dfdd0ea5fdde4723c0f4db73e909926fd86deff1d78e39d5f4d33b1bc59c4b`<br>PG32-D1-v9-APPROVAL-AND-FREEZE-ACT<br>`cc0e5cdb753a102444bba553b1f531c227f213fcf2458dc7a79b846b8eaf484a` |
| **committed_or_manifest_sha256** | – |
| **prev_entry_sha256** | `0d1f5b87e11f21e0dffabda4f27ac68391bedb3f38200364341af2fe4f1878ed` |
| **entry_line_sha256** | `657b1047b21f9848ecc631a1de93aa03f5f24462266b67d2b5603f1b61a18281` |

| Entry 93 | 2026-10-04 · VERIFICATION_RECORD · manifest – |
|---|---|
| **governing_provisions** | P26-18; P32-14; P32-13 |
| **subject** | PG32-D1<br>`f035916d53c2672fbe14df0918e1ea67d81928059428557168a3bc3d55d76cd7` |
| **committed_or_manifest_sha256** | – |
| **prev_entry_sha256** | `657b1047b21f9848ecc631a1de93aa03f5f24462266b67d2b5603f1b61a18281` |
| **entry_line_sha256** | `21badace25f4cc4f864be627a005cb1781a7280318314cc5e4867886c70e786e` |

| Entry 94 | 2026-10-04 · COMMITMENT · manifest – |
|---|---|
| **governing_provisions** | Act S1 v0.1.1 item 5 (f06f3268); D-1 v9 A-08 part (e) (entry 92, verified 93); M0006 LOCK_LT2 550273cd (entries 89-90, verified 91); R6 canonical build (entry 81, verified 82); PG-11 sec. 10.6; P26-14 |
| **subject** | OFTYGR-S1-S01-PREFILL-MANIFEST<br>`6d33e092e61b9f3615c64435b6b103babd002c31b556075c6ffbae6a9f921ab4`<br>OFTYGR-S1-S01-FORM-PREFILLED<br>`d20b8cf87a3a8293afe3d8542e6bcc89cd643151e8c0fb1fb2667f340276d428`<br>OFTYGR-S1-S01-SESSION-CONTEXT-MANIFEST<br>`91d29fa9f4ac864ed9e9577301049a1c3aa496dfe804f1ec69489c7ba364530e`<br>OFTYGR-S1-S01-SESSION-INVENTORY-VIEW<br>`ef1a7cada9103142df115ae9c60c2d251ff2f28837097c8b31f561b84ef1bf33`<br>OFTYGR-M0006-FORM-TEMPLATE<br>`6535021f082308f4c34e77bb183ba457ec755e3c26baaaf3901eb74134ce64bc`<br>OFTYGR-S1-S01-DISPLAY-MANIFEST<br>`0b658d2dd0ce8593c327aa28425cbfc96a0785ca4408e2db592ae4e315383f0e`<br>OFTYGR-S1-S01-GUIDE-MD<br>`d2671998096f7cb8cca0adcc4cd21f79eafed342d0f6e5ccb3b1b72f9a086ec1`<br>OFTYGR-S1-S01-GUIDE-PDF<br>`4e81acff9e75b1f5dcd58be266fbc4846d0f773c286975021957a38c14df05da`<br>OFTYGR-S1-S01-ZIP-ENCODER2<br>`fdd2338349a1e9050303fd40fa99aa46211e5b4c53edaf75adfd9fc6cffb1e53` |
| **committed_or_manifest_sha256** | `6d33e092e61b9f3615c64435b6b103babd002c31b556075c6ffbae6a9f921ab4` |
| **prev_entry_sha256** | `21badace25f4cc4f864be627a005cb1781a7280318314cc5e4867886c70e786e` |
| **entry_line_sha256** | `3c4a3db3de97f84adaaa1d8ac9fbc4a34b145abaf103d614504a4c0cc7c61181` |

| Entry 95 | 2026-10-04 · VERIFICATION_RECORD · manifest – |
|---|---|
| **governing_provisions** | P26-18; Act S1 v0.1.1 item 5 (f06f3268) |
| **subject** | OFTYGR-S1-S01-PREFILL-MANIFEST<br>`6d33e092e61b9f3615c64435b6b103babd002c31b556075c6ffbae6a9f921ab4`<br>OFTYGR-S1-S01-FORM-PREFILLED<br>`d20b8cf87a3a8293afe3d8542e6bcc89cd643151e8c0fb1fb2667f340276d428`<br>OFTYGR-S1-S01-SESSION-CONTEXT-MANIFEST<br>`91d29fa9f4ac864ed9e9577301049a1c3aa496dfe804f1ec69489c7ba364530e`<br>OFTYGR-S1-S01-SESSION-INVENTORY-VIEW<br>`ef1a7cada9103142df115ae9c60c2d251ff2f28837097c8b31f561b84ef1bf33`<br>OFTYGR-M0006-FORM-TEMPLATE<br>`6535021f082308f4c34e77bb183ba457ec755e3c26baaaf3901eb74134ce64bc`<br>OFTYGR-S1-S01-DISPLAY-MANIFEST<br>`0b658d2dd0ce8593c327aa28425cbfc96a0785ca4408e2db592ae4e315383f0e`<br>OFTYGR-S1-S01-GUIDE-MD<br>`d2671998096f7cb8cca0adcc4cd21f79eafed342d0f6e5ccb3b1b72f9a086ec1`<br>OFTYGR-S1-S01-GUIDE-PDF<br>`4e81acff9e75b1f5dcd58be266fbc4846d0f773c286975021957a38c14df05da`<br>OFTYGR-S1-S01-ZIP-ENCODER2<br>`fdd2338349a1e9050303fd40fa99aa46211e5b4c53edaf75adfd9fc6cffb1e53` |
| **committed_or_manifest_sha256** | – |
| **prev_entry_sha256** | `3c4a3db3de97f84adaaa1d8ac9fbc4a34b145abaf103d614504a4c0cc7c61181` |
| **entry_line_sha256** | `3e7aaf2622d8dc9b9f2211dc6358b1305606c9665015256e7c53dfbfc6626829` |

| Entry 96 | 2026-10-05 · COMMITMENT · manifest – |
|---|---|
| **governing_provisions** | Act S1 v0.1.1 item 5 (f06f3268); D-1 v9 A-08 part (e) (entry 92, verified 93); M0006 LOCK_LT2 550273cd (entries 89-90, verified 91); R6 canonical build (entry 81, verified 82); PG-11 sec. 10.6; P26-14 |
| **subject** | OFTYGR-S1-S01-FORM-RETURNED-CONFIRMED<br>a01e2bba0c9fe6fe5a4043a5516f958c3aabbedb4e249d8704c42046458508f2<br>OFTYGR-S1-S01-RETURN-EMAIL-RECORD<br>5a8bbc4abc94f53e3963b19e5bf8eba0e037d53b7ff58cdadef7875b722e9446<br>OFTYGR-S1-S01-PREFILL-MANIFEST<br>6d33e092e61b9f3615c64435b6b103babd002c31b556075c6ffbae6a9f921ab4 |
| **committed_or_manifest_sha256** | a01e2bba0c9fe6fe5a4043a5516f958c3aabbedb4e249d8704c42046458508f2 |
| **prev_entry_sha256** | 3e7aaf2622d8dc9b9f2211dc6358b1305606c9665015256e7c53dfbfc6626829 |
| **entry_line_sha256** | 1c4c91bbd5400612769212dddfbeed85e7feebe7d0ec4e2c298b674f96dc6f0b |

| Entry 97 | 2026-10-05 · VERIFICATION_RECORD · manifest – |
|---|---|
| **governing_provisions** | P26-18; Act S1 v0.1.1 item 5 (f06f3268) |
| **subject** | OFTYGR-S1-S01-FORM-RETURNED-CONFIRMED<br>a01e2bba0c9fe6fe5a4043a5516f958c3aabbedb4e249d8704c42046458508f2<br>OFTYGR-S1-S01-RETURN-EMAIL-RECORD<br>5a8bbc4abc94f53e3963b19e5bf8eba0e037d53b7ff58cdadef7875b722e9446<br>OFTYGR-S1-S01-PREFILL-MANIFEST<br>6d33e092e61b9f3615c64435b6b103babd002c31b556075c6ffbae6a9f921ab4 |
| **committed_or_manifest_sha256** | – |
| **prev_entry_sha256** | 1c4c91bbd5400612769212dddfbeed85e7feebe7d0ec4e2c298b674f96dc6f0b |
| **entry_line_sha256** | 2c5b419618ea8beffd0e5f144896ab0711f5cc8506b3c5d408467b9c4625a7bf |

| Entry 98 | 2026-10-05 · COMMITMENT · manifest – |
|---|---|
| **governing_provisions** | Act S1 v0.1.1 item 5 (f06f3268); D-1 v9 A-08 part (e) (entry 92, verified 93); M0006 LOCK_LT2 550273cd (entries 89-90, verified 91); R6 canonical build (entry 81, verified 82); PG-11 sec. 10.6; P26-14 |
| **subject** | OFTYGR-S1-S01-SUCCESSOR-LOG<br>`3f711273b691e72386c8d6f5166d2d6c9eb9d6a697900b6c016bc922825276e6`<br>OFTYGR-S1-S01-TRANSCRIPTION-SUMMARY<br>`9348f7a7c0f6db90e1639f3a4747d13d1908439a21e431b6ea10ab539898004b`<br>OFTYGR-S1-S01-SUCCESSOR-EVENT<br>`14f9e3d5dabde7d93fe0750d144611c33e46936af88d0dbe6a411b82c13e26f4`<br>OFTYGR-S1-S01-SUCCESSOR-EVENT<br>`5d8da91a857d7472c10c37af5d6f71f59dbc31706b7a531841d8938d0bc5ee23`<br>OFTYGR-S1-S01-SUCCESSOR-EVENT<br>`20bafdba03cebf67784d65e18c1d62d481ddd775d214d322c0d9fc8cce179002`<br>OFTYGR-S1-S01-SUCCESSOR-EVENT<br>`41e2adc0ddcc7dadab5c4e97a868b16a36f4493aebe02a84ef3c780473d0e30e`<br>OFTYGR-S1-S01-SUCCESSOR-EVENT<br>`989c09df051696201a8b7880388054a950a3ce161420229804b4c997222f5939`<br>OFTYGR-S1-S01-SUCCESSOR-EVENT<br>`8b47279cd821ef8e04cdd1d2f96c8a944db2e4521bc8ce09d228d010d1dc6b4d`<br>OFTYGR-S1-S01-SUCCESSOR-EVENT<br>`08e1a348cdc2506911da5ae4094dff6e8acfe464e95c78f90f3c5cc8140d687c`<br>OFTYGR-S1-S01-SUCCESSOR-EVENT<br>`570956c0ed9c2de8fb50cf4e1f70c7317e48e926eab709da0298cea5970b7444`<br>OFTYGR-S1-S01-SUCCESSOR-EVENT<br>`ee33adbf0f6950c274e466515ecc13b113b94fd736b13de823a6c8feecb75cd7`<br>OFTYGR-S1-S01-SUCCESSOR-EVENT<br>`fe1d61c01d818d17fa1ba5c2659f3c6e95533b7e2e9ff5f255bc489270b28077`<br>OFTYGR-S1-S01-SUCCESSOR-EVENT<br>`7d6a80ece3159f93468005a54c389a574869f0d47d4529b1efcc2d291590ad72`<br>OFTYGR-S1-S01-SUCCESSOR-EVENT<br>`2ce644febd7c3bf91a1319410c81e27ee525566c35304c5ee0eeb4231a9da78e`<br>OFTYGR-S1-S01-SUCCESSOR-EVENT<br>`2225b3246083c6120bb643472acdffea65696a10061f3d21183bfebf30d55c9c`<br>OFTYGR-S1-S01-SUCCESSOR-EVENT<br>`1bd91a9227d3c8a231246f59294d2a7a46aeac92b81619631654eeee9eeff5b9`<br>OFTYGR-S1-S01-SUCCESSOR-EVENT<br>`d23eee4b8e3b8e6dd46422349f4e61bc4564b7ed9a930b716dcf083a28d57dcd`<br>OFTYGR-S1-S01-SUCCESSOR-EVENT<br>`1326cce8ae668e4b3f42cbd6ecc652b31f4c7991b05fc628bae149b442b70b7e`<br>OFTYGR-S1-S01-SUCCESSOR-EVENT<br>`494a314789daf7451c044b48d6420c6ed23168defa1cfd64f0dce0925e5e6249`<br>OFTYGR-S1-S01-SUCCESSOR-EVENT<br>`440a35ed03769d9b41a1cc2a3ffd91f2eadaca370c947621005b951cc0f1794e`<br>OFTYGR-S1-S01-SUCCESSOR-EVENT<br>`d644f739823a6f53b98e5ef375880a776e022b572bd9c1a0f4f9dfb152053d0c`<br>OFTYGR-S1-S01-SUCCESSOR-EVENT<br>`f7a023c6c3784ce52739f4a0b1914c29babae2ba7996626cecefcf3b9f1ba81a`<br>OFTYGR-S1-S01-SUCCESSOR-EVENT<br>`fbdd7df77063db2e2dd91e970c043f2b237354c66aa587d6c18a28053df94d12`<br>OFTYGR-S1-S01-SUCCESSOR-EVENT<br>`10461577dbf6ccdb16c43646dd1e97b995564c21a74efc769e591bbb08a0ab36`<br>OFTYGR-S1-S01-SUCCESSOR-EVENT<br>`f51336ec756194f896d7ea3cb97ee5c2c3c15e52d98ca2bbf268860a101d964b`<br>OFTYGR-S1-S01-SUCCESSOR-EVENT<br>`99610022c6c7b545d0b135c017d9d25f0806064fb9ecb785053d4d1d4ab6b750` |
| **committed_or_manifest_sha256** | `3f711273b691e72386c8d6f5166d2d6c9eb9d6a697900b6c016bc922825276e6` |
| **prev_entry_sha256** | `2c5b419618ea8beffd0e5f144896ab0711f5cc8506b3c5d408467b9c4625a7bf` |
| **entry_line_sha256** | `437a83c8623a7fe91d999a51cf2e60d0e0ae7cc414e56a23ae6dad9c32c3a890` |

| Entry 99 | 2026-10-05 · VERIFICATION_RECORD · manifest – |
|---|---|
| **governing_provisions** | P26-18; Act S1 v0.1.1 item 5 (f06f3268) |
| **subject** | OFTYGR-S1-S01-SUCCESSOR-LOG<br>3f711273b691e72386c8d6f5166d2d6c9eb9d6a697900b6c016bc922825276e6<br>OFTYGR-S1-S01-TRANSCRIPTION-SUMMARY<br>9348f7a7c0f6db90e1639f3a4747d13d1908439a21e431b6ea10ab539898004b<br>OFTYGR-S1-S01-SUCCESSOR-EVENT<br>14f9e3d5dabde7d93fe0750d144611c33e46936af88d0dbe6a411b82c13e26f4<br>OFTYGR-S1-S01-SUCCESSOR-EVENT<br>5d8da91a857d7472c10c37af5d6f71f59dbc31706b7a531841d8938d0bc5ee23<br>OFTYGR-S1-S01-SUCCESSOR-EVENT<br>20bafdba03cebf67784d65e18c1d62d481ddd775d214d322c0d9fc8cce179002<br>OFTYGR-S1-S01-SUCCESSOR-EVENT<br>41e2adc0ddcc7dadab5c4e97a868b16a36f4493aebe02a84ef3c780473d0e30e<br>OFTYGR-S1-S01-SUCCESSOR-EVENT<br>989c09df051696201a8b7880388054a950a3ce161420229804b4c997222f5939<br>OFTYGR-S1-S01-SUCCESSOR-EVENT<br>8b47279cd821ef8e04cdd1d2f96c8a944db2e4521bc8ce09d228d010d1dc6b4d<br>OFTYGR-S1-S01-SUCCESSOR-EVENT<br>08e1a348cdc2506911da5ae4094dff6e8acfe464e95c78f90f3c5cc8140d687c<br>OFTYGR-S1-S01-SUCCESSOR-EVENT<br>570956c0ed9c2de8fb50cf4e1f70c7317e48e926eab709da0298cea5970b7444<br>OFTYGR-S1-S01-SUCCESSOR-EVENT<br>ee33adbf0f6950c274e466515ecc13b113b94fd736b13de823a6c8feecb75cd7<br>OFTYGR-S1-S01-SUCCESSOR-EVENT<br>fe1d61c01d818d17fa1ba5c2659f3c6e95533b7e2e9ff5f255bc489270b28077<br>OFTYGR-S1-S01-SUCCESSOR-EVENT<br>7d6a80ece3159f93468005a54c389a574869f0d47d4529b1efcc2d291590ad72<br>OFTYGR-S1-S01-SUCCESSOR-EVENT<br>2ce644febd7c3bf91a1319410c81e27ee525566c35304c5ee0eeb4231a9da78e<br>OFTYGR-S1-S01-SUCCESSOR-EVENT<br>2225b3246083c6120bb643472acdffea65696a10061f3d21183bfebf30d55c9c<br>OFTYGR-S1-S01-SUCCESSOR-EVENT<br>1bd91a9227d3c8a231246f59294d2a7a46aeac92b81619631654eeee9eeff5b9<br>OFTYGR-S1-S01-SUCCESSOR-EVENT<br>d23eee4b8e3b8e6dd46422349f4e61bc4564b7ed9a930b716dcf083a28d57dcd<br>OFTYGR-S1-S01-SUCCESSOR-EVENT<br>1326cce8ae668e4b3f42cbd6ecc652b31f4c7991b05fc628bae149b442b70b7e<br>OFTYGR-S1-S01-SUCCESSOR-EVENT<br>494a314789daf7451c044b48d6420c6ed23168defa1cfd64f0dce0925e5e6249<br>OFTYGR-S1-S01-SUCCESSOR-EVENT<br>440a35ed03769d9b41a1cc2a3ffd91f2eadaca370c947621005b951cc0f1794e<br>OFTYGR-S1-S01-SUCCESSOR-EVENT<br>d644f739823a6f53b98e5ef375880a776e022b572bd9c1a0f4f9dfb152053d0c<br>OFTYGR-S1-S01-SUCCESSOR-EVENT<br>f7a023c6c3784ce52739f4a0b1914c29babae2ba7996626cecefcf3b9f1ba81a<br>OFTYGR-S1-S01-SUCCESSOR-EVENT<br>fbdd7df77063db2e2dd91e970c043f2b237354c66aa587d6c18a28053df94d12<br>OFTYGR-S1-S01-SUCCESSOR-EVENT<br>10461577dbf6ccdb16c43646dd1e97b995564c21a74efc769e591bbb08a0ab36<br>OFTYGR-S1-S01-SUCCESSOR-EVENT<br>f51336ec756194f896d7ea3cb97ee5c2c3c15e52d98ca2bbf268860a101d964b<br>OFTYGR-S1-S01-SUCCESSOR-EVENT<br>99610022c6c7b545d0b135c017d9d25f0806064fb9ecb785053d4d1d4ab6b750 |
| **committed_or_manifest_sha256** | – |
| **prev_entry_sha256** | 437a83c8623a7fe91d999a51cf2e60d0e0ae7cc414e56a23ae6dad9c32c3a890 |
| **entry_line_sha256** | 6fb8200e1ce2d4a1f7b9d82dd03aced76fd0a3844b96aca39f0af5d9bb2f08bc |

| Entry 100 | 2026-10-05 · FREEZE_ACT · manifest M0007 |
|---|---|
| **governing_provisions** | P26-18; P26-14; P26-10(i); P26-10(ii); P26-02(vi) |
| **subject** | PG26-M0007<br>543feae4a2b968cef0cad9894c3a331078c15bbec5abd1bdfab654c934ec08d2<br>PG11-RESOLUTION-RECORD<br>9ef6450a40ddb945bda7e7a867b1226ba32d413677e4e90597a1acee386d4b4b |
| **committed_or_manifest_sha256** | 543feae4a2b968cef0cad9894c3a331078c15bbec5abd1bdfab654c934ec08d2 |
| **prev_entry_sha256** | 6fb8200e1ce2d4a1f7b9d82dd03aced76fd0a3844b96aca39f0af5d9bb2f08bc |
| **entry_line_sha256** | 3ef17d676516a0ed775334afb604b7500be3afffa9fc2d3fa5d3b5642a530c35 |

| Entry 101 | 2026-10-05 · COMMITMENT · manifest – |
|---|---|
| **governing_provisions** | P26-10(i); P26-02(iii) as amended by CA2-10/CA3-10; P26-14 |
| **subject** | PG11-RESOLUTION-RECORD<br>`9ef6450a40ddb945bda7e7a867b1226ba32d413677e4e90597a1acee386d4b4b`<br>PG11-v0.1.3-CHANGE-RECORD-P26-10<br>`c1b77a02f9f8f670ae2451caa49458b55b1003031090179657415ebf6ff2f80a`<br>PG11-v0.1.3-CLAUSE-CORRECTION-CC-1<br>`7e5fb75a6fc69b0d40c50a1ea85cac391fe21e60836a9a17a9051a5774fb9ba8` |
| **committed_or_manifest_sha256** | `9ef6450a40ddb945bda7e7a867b1226ba32d413677e4e90597a1acee386d4b4b` |
| **prev_entry_sha256** | `3ef17d676516a0ed775334afb604b7500be3afffa9fc2d3fa5d3b5642a530c35` |
| **entry_line_sha256** | `4723afb18478097bdb9f0491f588c9b42477a72f3e02c9ccefd3df8431b9072d` |

| Entry 102 | 2026-10-05 · VERIFICATION_RECORD · manifest – |
|---|---|
| **governing_provisions** | P26-18; P26-14; P26-10(i) |
| **subject** | PG11-RESOLUTION-RECORD<br>`9ef6450a40ddb945bda7e7a867b1226ba32d413677e4e90597a1acee386d4b4b`<br>PG11-v0.1.3-CHANGE-RECORD-P26-10<br>`c1b77a02f9f8f670ae2451caa49458b55b1003031090179657415ebf6ff2f80a`<br>PG11-v0.1.3-CLAUSE-CORRECTION-CC-1<br>`7e5fb75a6fc69b0d40c50a1ea85cac391fe21e60836a9a17a9051a5774fb9ba8` |
| **committed_or_manifest_sha256** | – |
| **prev_entry_sha256** | `4723afb18478097bdb9f0491f588c9b42477a72f3e02c9ccefd3df8431b9072d` |
| **entry_line_sha256** | `ea4cf867fcad692116ebcfe34248bff6cbbf7b2cc7279960635778e102de937f` |

| Entry 103 | 2026-10-05 · COMMITMENT · manifest – |
|---|---|
| **governing_provisions** | P26-02(iii) as amended by CA2-10/CA3-10; P26-14; P26-10(ii) |
| **subject** | PG26-M0007<br>`543feae4a2b968cef0cad9894c3a331078c15bbec5abd1bdfab654c934ec08d2` |
| **committed_or_manifest_sha256** | `543feae4a2b968cef0cad9894c3a331078c15bbec5abd1bdfab654c934ec08d2` |
| **prev_entry_sha256** | `ea4cf867fcad692116ebcfe34248bff6cbbf7b2cc7279960635778e102de937f` |
| **entry_line_sha256** | `022385a65629c36d37fa974f75769c4d00e582bc23014dd2ac515d07415955bd` |

| Entry 104 | 2026-10-05 · VERIFICATION_RECORD · manifest – |
|---|---|
| **governing_provisions** | P26-18; P26-14; P26-12 |
| **subject** | PG26-M0007<br>`543feae4a2b968cef0cad9894c3a331078c15bbec5abd1bdfab654c934ec08d2` |
| **committed_or_manifest_sha256** | – |
| **prev_entry_sha256** | `022385a65629c36d37fa974f75769c4d00e582bc23014dd2ac515d07415955bd` |
| **entry_line_sha256** | `e9e231bf692d035a5ac3f942b7dda5e48775a0143d2f7b500d6b823d1c76937f` |

| Entry 105 | 2026-10-05 · FREEZE_ACT · manifest M0008 |
|---|---|
| **governing_provisions** | P26-18; P26-14; P26-02(vi) |
| **subject** | PG26-M0008<br>`413bc77045b477dda3d40531b00461615050ab4391cca36fac3b289f67af05cd` |
| **committed_or_manifest_sha256** | `413bc77045b477dda3d40531b00461615050ab4391cca36fac3b289f67af05cd` |
| **prev_entry_sha256** | `e9e231bf692d035a5ac3f942b7dda5e48775a0143d2f7b500d6b823d1c76937f` |
| **entry_line_sha256** | `5a8645f52084782c624e51199d3d8b1d5b5f2f8528039c654cd4f93f1084f785` |

| Entry 106 | 2026-10-05 · COMMITMENT · manifest – |
|---|---|
| **governing_provisions** | P26-02(iii) as amended by CA2-10/CA3-10; P26-14 |
| **subject** | PG26-M0008<br>`413bc77045b477dda3d40531b00461615050ab4391cca36fac3b289f67af05cd` |
| **committed_or_manifest_sha256** | `413bc77045b477dda3d40531b00461615050ab4391cca36fac3b289f67af05cd` |
| **prev_entry_sha256** | `5a8645f52084782c624e51199d3d8b1d5b5f2f8528039c654cd4f93f1084f785` |
| **entry_line_sha256** | `3067d97958ac0e3d1b12e19b6470f84e2933766451af35cbc112f5ff31f72a11` |

| Entry 107 | 2026-10-05 · VERIFICATION_RECORD · manifest – |
|---|---|
| **governing_provisions** | P26-18; P26-14; P26-12 |
| **subject** | PG26-M0008<br>`413bc77045b477dda3d40531b00461615050ab4391cca36fac3b289f67af05cd` |
| **committed_or_manifest_sha256** | – |
| **prev_entry_sha256** | `3067d97958ac0e3d1b12e19b6470f84e2933766451af35cbc112f5ff31f72a11` |
| **entry_line_sha256** | `394a806e3a045b0530651ccf7d42b93b27fbe3b355f71951dc8d5269bd6fca83` |

| Entry 108 | 2026-10-05 · FREEZE_ACT · manifest M0009 |
|---|---|
| **governing_provisions** | P26-18; P26-14; P26-02(vi) |
| **subject** | PG26-M0009<br>`8c90e0b11e987255b5e42ee96796ae0a07e41d84d70d2d2b18c511f9d698abbf` |
| **committed_or_manifest_sha256** | `8c90e0b11e987255b5e42ee96796ae0a07e41d84d70d2d2b18c511f9d698abbf` |
| **prev_entry_sha256** | `394a806e3a045b0530651ccf7d42b93b27fbe3b355f71951dc8d5269bd6fca83` |
| **entry_line_sha256** | `9e81493ac5da3b1da26fa393a9d90e7698b99f72f08b36c999de263f145f5e27` |

| Entry 109 | 2026-10-05 · COMMITMENT · manifest – |
|---|---|
| **governing_provisions** | P26-02(iii) as amended by CA2-10/CA3-10; P26-14 |
| **subject** | PG26-M0009<br>8c90e0b11e987255b5e42ee96796ae0a07e41d84d70d2d2b18c511f9d698abbf |
| **committed_or_manifest_sha256** | 8c90e0b11e987255b5e42ee96796ae0a07e41d84d70d2d2b18c511f9d698abbf |
| **prev_entry_sha256** | 9e81493ac5da3b1da26fa393a9d90e7698b99f72f08b36c999de263f145f5e27 |
| **entry_line_sha256** | d669b9ab4c41a74cfdf2e66ec31a8df9400cc39ba4b8eea313b52885dd65609d |

| Entry 110 | 2026-10-05 · VERIFICATION_RECORD · manifest – |
|---|---|
| **governing_provisions** | P26-18; P26-14; P26-12 |
| **subject** | PG26-M0009<br>8c90e0b11e987255b5e42ee96796ae0a07e41d84d70d2d2b18c511f9d698abbf |
| **committed_or_manifest_sha256** | – |
| **prev_entry_sha256** | d669b9ab4c41a74cfdf2e66ec31a8df9400cc39ba4b8eea313b52885dd65609d |
| **entry_line_sha256** | 6729be4d05fde4fd569f71ee0cdd7751accacdb445b8ae7f11e1641539b713de |

| Entry 111 | 2026-10-05 · ROLE_RECORD · manifest – |
|---|---|
| **governing_provisions** | P32-14; P32-13; P32-02; PG-11 v0.1.3 sec. 10.5.2; PG-11 v0.1.3 sec. 10.7; P26-14 |
| **subject** | PG32-D1<br>b2c8521849d179b8ad257782ef718cd3a1da8e7e325633007e79dc91e4652926<br>PG26-M0009<br>8c90e0b11e987255b5e42ee96796ae0a07e41d84d70d2d2b18c511f9d698abbf<br>GATEKEEPER-REPORT-M0007-r3-M0003P-r3-and-D1v10<br>1dd88633f5712f3ccf517aa53627d1b59dfd9a2f3682a57f1a9fc39fef096ead<br>PG32-D1-v10-APPROVAL-AND-FREEZE-ACT<br>d65bc3953ebefed97d55a969432a7b292dc77819eb5f71753b53cf26c2316776 |
| **committed_or_manifest_sha256** | – |
| **prev_entry_sha256** | 6729be4d05fde4fd569f71ee0cdd7751accacdb445b8ae7f11e1641539b713de |
| **entry_line_sha256** | 3ae82b288e330f788f3cff7a6d6ff6a31921065640c64f50cbf882eff68f1515 |

| Entry 112 | 2026-10-05 · VERIFICATION_RECORD · manifest – |
|---|---|
| **governing_provisions** | P26-18; P32-14; P32-13 |
| **subject** | PG32-D1<br>b2c8521849d179b8ad257782ef718cd3a1da8e7e325633007e79dc91e4652926 |
| **committed_or_manifest_sha256** | – |
| **prev_entry_sha256** | 3ae82b288e330f788f3cff7a6d6ff6a31921065640c64f50cbf882eff68f1515 |
| **entry_line_sha256** | e24d461dd091d2231b90d7978d089e13d5f2be6be83b877be27e5dd08807d391 |

| Entry 113 | 2026-10-05 · COMMITMENT · manifest – |
|---|---|
| **governing_provisions** | Act S1-T v1.1 (b00d4e83); D-1 v10 A-08 part (f) (ROLE_RECORD entry 111, verified 112); PG-11 v0.1.3 (entry 101, verified 102); M0009 LOCK_LT2 8c90e0b1 (entries 108-110); M0008 LOCK_LT2 413bc770 (entries 105-107); R6 canonical build (entry 81, verified 82); PG-11 sec. 10.6; P26-14 |
| **subject** | OFTYGR-S1T-S01-PREPARATION-INPUT-SET<br>dd8e5fe96a88b27c0c6cd18337d54edb453b5d541af15cb48a8dcc52f3775a8b<br>OFTYGR-S1T-S01-INVENTORY-VIEW<br>099f10f4c5ab1685a728c5ee8bdbc1f70fc52664547490585eca58221cd02da6<br>OFTYGR-S1T-S01-FAMILY-FRAME-BINDINGS<br>6705cb0eb8821f7823d8c692623e384a6b3fca709b5865094421fd93098cbc57<br>OFTYGR-S1T-S01-PREDECESSOR-STATES<br>68ab82e09f8a3065b8b9dbb663b6a6211199a64818b99165c7644fffeec66db3<br>OFTYGR-S1T-S01-PX-E1-BASE<br>b7f3a6eed143c300bbd460120d82ef7293a8524698ef91f605071a1aeae3a1f1<br>OFTYGR-S1T-S01-PX-E2-BASE<br>f452a067ed1d678da722deecd90b640a539d321e745e5fac3ce7acf32fc06cea<br>OFTYGR-S1T-S01-DV-STRUCTURAL-INDEX<br>05af46b04969edd6f12ca7584746821d1183da943ef9c2198f81069abd5e2266<br>OFTYGR-S1T-S01-DV-EDGES-ROUND1<br>3dbfcef7c22416c48aa73c45c469275d7f5342f17d84e386a5804cd948328faa<br>OFTYGR-S1T-S01-CC1-BINDING-TABLE<br>a857bce6cbca2e24685ec42c3b8c6071cf73f69c6f23aaf723c1aaa96c14044e<br>OFTYGR-S1T-S01-PREFILL-DECL-MANIFEST<br>6e50d522326646c889d6f3ec9f7344d6f0a452a78951b4e571ff43d775ddac00<br>OFTYGR-S1T-S01-PREFILL-DECL-FORM<br>db527eb71f8f31dc00b35d923f09c435fd6ee5ceb2d83f6ceb27a5861e5a166d<br>OFTYGR-M0009-FORM-TEMPLATE-V013<br>b6d75e94979ea46534eb5fca753cfbe72360f187ff4070260157adb93191dd1d<br>OFTYGR-S1T-S01-DISPLAY-MANIFEST<br>13b64919aaf63ef7d9636715e0cebcff653fef10256128cd6588698b72aea6ab |
| **committed_or_manifest_sha256** | dd8e5fe96a88b27c0c6cd18337d54edb453b5d541af15cb48a8dcc52f3775a8b |
| **prev_entry_sha256** | e24d461dd091d2231b90d7978d089e13d5f2be6be83b877be27e5dd08807d391 |
| **entry_line_sha256** | 6e91571ea14e9daea1ca4dd05b0a24e440db5a5cab60f6137be4246e7a8e5000 |

| Entry 114 | 2026-10-05 · VERIFICATION_RECORD · manifest – |
|---|---|
| **governing_provisions** | P26-18; Act S1-T v1.1 (b00d4e83) |
| **subject** | OFTYGR-S1T-S01-PREPARATION-INPUT-SET<br>dd8e5fe96a88b27c0c6cd18337d54edb453b5d541af15cb48a8dcc52f3775a8b<br>OFTYGR-S1T-S01-INVENTORY-VIEW<br>099f10f4c5ab1685a728c5ee8bdbc1f70fc52664547490585eca58221cd02da6<br>OFTYGR-S1T-S01-FAMILY-FRAME-BINDINGS<br>6705cb0eb8821f7823d8c692623e384a6b3fca709b5865094421fd93098cbc57<br>OFTYGR-S1T-S01-PREDECESSOR-STATES<br>68ab82e09f8a3065b8b9dbb663b6a6211199a64818b99165c7644fffeec66db3<br>OFTYGR-S1T-S01-PX-E1-BASE<br>b7f3a6eed143c300bbd460120d82ef7293a8524698ef91f605071a1aeae3a1f1<br>OFTYGR-S1T-S01-PX-E2-BASE<br>f452a067ed1d678da722deecd90b640a539d321e745e5fac3ce7acf32fc06cea<br>OFTYGR-S1T-S01-DV-STRUCTURAL-INDEX<br>05af46b04969edd6f12ca7584746821d1183da943ef9c2198f81069abd5e2266<br>OFTYGR-S1T-S01-DV-EDGES-ROUND1<br>3dbfcef7c22416c48aa73c45c469275d7f5342f17d84e386a5804cd948328faa<br>OFTYGR-S1T-S01-CC1-BINDING-TABLE<br>a857bce6cbca2e24685ec42c3b8c6071cf73f69c6f23aaf723c1aaa96c14044e<br>OFTYGR-S1T-S01-PREFILL-DECL-MANIFEST<br>6e50d522326646c889d6f3ec9f7344d6f0a452a78951b4e571ff43d775ddac00<br>OFTYGR-S1T-S01-PREFILL-DECL-FORM<br>db527eb71f8f31dc00b35d923f09c435fd6ee5ceb2d83f6ceb27a5861e5a166d<br>OFTYGR-M0009-FORM-TEMPLATE-V013<br>b6d75e94979ea46534eb5fca753cfbe72360f187ff4070260157adb93191dd1d<br>OFTYGR-S1T-S01-DISPLAY-MANIFEST<br>13b64919aaf63ef7d9636715e0cebcff653fef10256128cd6588698b72aea6ab |
| **committed_or_manifest_sha256** | – |
| **prev_entry_sha256** | 6e91571ea14e9daea1ca4dd05b0a24e440db5a5cab60f6137be4246e7a8e5000 |
| **entry_line_sha256** | d717324d707f59c1fb8e15600bd30803a82440e9902d91cd603456fe34cbbffa |

| Entry 115 | 2026-10-05 · COMMITMENT · manifest – |
|---|---|
| **governing_provisions** | PI instruction record 20261005T1357Z (c8434d6e); Codex check bf644a75 L1; D-1 v10 A-08 part (f) (ROLE_RECORD entry 111, verified 112); M0009 LOCK_LT2 8c90e0b1 (entries 108-110); P-13 chain commitments (entries 85-86, verified 87-88); P26-14 |
| **subject** | OFTYGR-P13-LINKED-CLOSURE<br>9c5170b36c0ad7562d84ca75aa1c331baa452432027ae381fa0d153d26948d8c<br>OFTYGR-P13-N08A-PACKAGE-MANIFEST<br>bf1526216b68f3b805e1231818f842979653936b27ab362358077c9c34e12ecf<br>OFTYGR-P13-CHAIN-MANIFEST-FAM-0001<br>5271218200ec5fa4f09bcf8a416156c017d07ae53053ad6d4f86ffb4f3fa9568<br>OFTYGR-P13-CHAIN-MANIFEST-FAM-0002<br>ff71f554a61013696867f23fb9d6719bd53068e496369f34123651d008913f5c |
| **committed_or_manifest_sha256** | 9c5170b36c0ad7562d84ca75aa1c331baa452432027ae381fa0d153d26948d8c |
| **prev_entry_sha256** | d717324d707f59c1fb8e15600bd30803a82440e9902d91cd603456fe34cbbffa |
| **entry_line_sha256** | 91e60b525f365e0bdf9482781530acaae31ccdfbc49eb14964c1ffdac50fdfc4 |

| Entry 116 | 2026-10-05 · VERIFICATION_RECORD · manifest – |
|---|---|
| **governing_provisions** | P26-18; Codex check bf644a75 L1 |
| **subject** | OFTYGR-P13-LINKED-CLOSURE<br>9c5170b36c0ad7562d84ca75aa1c331baa452432027ae381fa0d153d26948d8c<br>OFTYGR-P13-N08A-PACKAGE-MANIFEST<br>bf1526216b68f3b805e1231818f842979653936b27ab362358077c9c34e12ecf<br>OFTYGR-P13-CHAIN-MANIFEST-FAM-0001<br>5271218200ec5fa4f09bcf8a416156c017d07ae53053ad6d4f86ffb4f3fa9568<br>OFTYGR-P13-CHAIN-MANIFEST-FAM-0002<br>ff71f554a61013696867f23fb9d6719bd53068e496369f34123651d008913f5c |
| **committed_or_manifest_sha256** | – |
| **prev_entry_sha256** | 91e60b525f365e0bdf9482781530acaae31ccdfbc49eb14964c1ffdac50fdfc4 |
| **entry_line_sha256** | 182e83be2864e1ea4b778730f3d6a1cb5a1447dc70e42d04e1f2c03b30da6f54 |

| Entry 117 | 2026-10-05 · COMMITMENT · manifest – |
|---|---|
| **governing_provisions** | PI instruction record 20261005T1357Z (c8434d6e); Codex check bf644a75 L1; D-1 v10 A-08 part (f) (ROLE_RECORD entry 111, verified 112); M0009 LOCK_LT2 8c90e0b1 (entries 108-110); S1-T-SESSION-01 preparation (entry 113, verified 114); P-13 LINKED_CLOSURE admission (LC1/LC2); P26-14 |
| **subject** | OFTYGR-S1T-S01-DV-STRUCTURAL-INDEX<br>03f051ff8bcc15db7301c061d9b72c7f9534411dd8ca7bd6ae816af0761a6ca8<br>OFTYGR-S1T-S01-DV-EDGES-ROUND1<br>0ecf722e031acbed133af8a6490af831ae0bdba480b9f1ff88e1c3e7280ee6ec |
| **committed_or_manifest_sha256** | 03f051ff8bcc15db7301c061d9b72c7f9534411dd8ca7bd6ae816af0761a6ca8 |
| **prev_entry_sha256** | 182e83be2864e1ea4b778730f3d6a1cb5a1447dc70e42d04e1f2c03b30da6f54 |
| **entry_line_sha256** | 426226421e49d2b265c2be39905754a3765b3c4ea664fd81fa6fa8098f8713b4 |

| Entry 118 | 2026-10-05 · VERIFICATION_RECORD · manifest – |
|---|---|
| **governing_provisions** | P26-18; Codex check bf644a75 L1 |
| **subject** | OFTYGR-S1T-S01-DV-STRUCTURAL-INDEX<br>`03f051ff8bcc15db7301c061d9b72c7f9534411dd8ca7bd6ae816af0761a6ca8`<br>OFTYGR-S1T-S01-DV-EDGES-ROUND1<br>`0ecf722e031acbed133af8a6490af831ae0bdba480b9f1ff88e1c3e7280ee6ec` |
| **committed_or_manifest_sha256** | – |
| **prev_entry_sha256** | `426226421e49d2b265c2be39905754a3765b3c4ea664fd81fa6fa8098f8713b4` |
| **entry_line_sha256** | `284d27b089e1c8d36851c76dda1a16fbb60048a0098eabc99cfe83b5ff883750` |

| Entry 119 | 2026-10-05 · COMMITMENT · manifest – |
|---|---|
| **governing_provisions** | PI instruction record 20261005T1549Z (3d95f2f9); Codex return corr1 check 3a89730f; Act S1-T v1.1 (b00d4e83); PG-11 v0.1.3 (entry 101, verified 102); D-1 v10 A-08 part (f) (ROLE_RECORD entry 111, verified 112); M0009 LOCK_LT2 8c90e0b1 (entries 108-110); DV index/edges (entry 117, verified 118); P26-14 |
| **subject** | OFTYGR-S1T-S01-V013-SUCCESSOR-LOG<br>`bdbc721489c61167928a710daf31ea252110380a4533053ee0822e753668c77e`<br>OFTYGR-S1T-S01-TRANSCRIPTION-MANIFEST<br>`db9e113a03aa7a03fbb7f4cc8704a7821cf7b97964e789c7fca5f915ded44976`<br>OFTYGR-S1T-S01-RETURN-corr1-WORKBOOK<br>`9ed49bf0b13567eab2f2ae93cdbe7b1fd52e23d0ff2ae0d54eb0943eb4f4f49e`<br>OFTYGR-S1T-S01-PRESTAMP-v0.2-MANIFEST<br>`0d6ae5a9dca2768180bc2ee71d25e64b48568af6962b9cb58d09b61e0e57f6d2`<br>OFTYGR-S1T-S01-PX-E1-AT118<br>`02395badf80bb62c967fe32fad2e3053b45377318b30f660ba4c150717a5bf0b`<br>OFTYGR-S1T-S01-PX-E2-AT118<br>`cf44c2f81b8a70b338c0eeed74b21b363b51ae06c36ec4c37400393d131df9ce` |
| **committed_or_manifest_sha256** | `bdbc721489c61167928a710daf31ea252110380a4533053ee0822e753668c77e` |
| **prev_entry_sha256** | `284d27b089e1c8d36851c76dda1a16fbb60048a0098eabc99cfe83b5ff883750` |
| **entry_line_sha256** | `20a703a3d03729700f93ee76193f90e31deae83e85978310f5a985caeb8cc0d0` |

| Entry 120 | 2026-10-05 · VERIFICATION_RECORD · manifest – |
|---|---|
| **governing_provisions** | P26-18; Codex return corr1 check 3a89730f |
| **subject** | OFTYGR-S1T-S01-V013-SUCCESSOR-LOG<br>`bdbc721489c61167928a710daf31ea252110380a4533053ee0822e753668c77e`<br>OFTYGR-S1T-S01-TRANSCRIPTION-MANIFEST<br>`db9e113a03aa7a03fbb7f4cc8704a7821cf7b97964e789c7fca5f915ded44976`<br>OFTYGR-S1T-S01-RETURN-corr1-WORKBOOK<br>`9ed49bf0b13567eab2f2ae93cdbe7b1fd52e23d0ff2ae0d54eb0943eb4f4f49e`<br>OFTYGR-S1T-S01-PRESTAMP-v0.2-MANIFEST<br>`0d6ae5a9dca2768180bc2ee71d25e64b48568af6962b9cb58d09b61e0e57f6d2`<br>OFTYGR-S1T-S01-PX-E1-AT118<br>`02395badf80bb62c967fe32fad2e3053b45377318b30f660ba4c150717a5bf0b`<br>OFTYGR-S1T-S01-PX-E2-AT118<br>`cf44c2f81b8a70b338c0eeed74b21b363b51ae06c36ec4c37400393d131df9ce` |
| **committed_or_manifest_sha256** | – |
| **prev_entry_sha256** | `20a703a3d03729700f93ee76193f90e31deae83e85978310f5a985caeb8cc0d0` |
| **entry_line_sha256** | `afe4fe9feb4df4968118f73833ac591a1891d12d1922847ac373718b829a9dbd` |

| Entry 121 | 2026-10-05 · COMMITMENT · manifest – |
|---|---|
| **governing_provisions** | PI instruction record 20261005T1549Z (3d95f2f9); Codex return corr1 check 3a89730f; Act S1-T v1.1 (b00d4e83); PG-11 v0.1.3 (entry 101, verified 102); D-1 v10 A-08 part (f) (ROLE_RECORD entry 111, verified 112); M0009 LOCK_LT2 8c90e0b1 (entries 108-110); DV index/edges (entry 117, verified 118); P26-14 |
| **subject** | OFTYGR-S1T-S01-DV-RECORD-DV-S1T-SESSION-01<br>`7b1284ed082b477b77d049a51adc639013e185ed53a75e962c7d0e01279ff8f6`<br>OFTYGR-S1T-S01-V013-SUCCESSOR-LOG<br>`bdbc721489c61167928a710daf31ea252110380a4533053ee0822e753668c77e` |
| **committed_or_manifest_sha256** | `7b1284ed082b477b77d049a51adc639013e185ed53a75e962c7d0e01279ff8f6` |
| **prev_entry_sha256** | `afe4fe9feb4df4968118f73833ac591a1891d12d1922847ac373718b829a9dbd` |
| **entry_line_sha256** | `c786264c3a02a70e0a854035a108c71eb35526aca8b9ada49b5d0d8d7bf0458b` |

| Entry 122 | 2026-10-05 · VERIFICATION_RECORD · manifest – |
|---|---|
| **governing_provisions** | P26-18; Codex return corr1 check 3a89730f |
| **subject** | OFTYGR-S1T-S01-DV-RECORD-DV-S1T-SESSION-01<br>`7b1284ed082b477b77d049a51adc639013e185ed53a75e962c7d0e01279ff8f6`<br>OFTYGR-S1T-S01-V013-SUCCESSOR-LOG<br>`bdbc721489c61167928a710daf31ea252110380a4533053ee0822e753668c77e` |
| **committed_or_manifest_sha256** | – |
| **prev_entry_sha256** | `c786264c3a02a70e0a854035a108c71eb35526aca8b9ada49b5d0d8d7bf0458b` |
| **entry_line_sha256** | `4462bb66e4d115814171e8f28bf0387e4a02ac87594df136a6111c48106474fa` |

| Entry 123 | 2026-10-05 · COMMITMENT · manifest – |
|---|---|
| **governing_provisions** | PI instruction record 20261005T1739Z (30b173f8); Codex follow-up corr1 check e288c9b3; Codex follow-up return check fdfa9488; Act S1-T v1.1 (b00d4e83); PG-11 v0.1.3 (entry 101, verified 102); D-1 v10 A-08 part (f) (ROLE_RECORD entry 111, verified 112); M0009 LOCK_LT2 8c90e0b1 (entries 108-110); transcription log (entry 119, verified 120); DV record (entry 121, verified 122); P26-14 |
| **subject** | OFTYGR-S1T-S01-FOLLOWUP-V013-LOG<br>`9a625a2292bf7804701211ddf8507253542e4b00128408d9c6c85ae5e0cac41e`<br>OFTYGR-S1T-S01-FOLLOWUP-TRANSCRIPTION-MANIFEST<br>`66f6ad7183a4c4d5d721bdc1589664515e0a62c7dbe0c2784b7a0df06a8c936f`<br>OFTYGR-S1T-S01-FOLLOWUP-HRD-ONLY-INPUT<br>`0dfb6bfc9b53b54d340737e7f8d089ca2c85a4ec67a75149644d02a352e0da5c`<br>OFTYGR-S1T-S01-FOLLOWUP-R3-PROVENANCE<br>`57f7a24d45b35e2f7c619ce016ff1ba5d5857c74967c7eb0c4b898111f54496c`<br>OFTYGR-S1T-S01-FOLLOWUP-R2-TRANSCRIPTION-INPUT<br>`1ba2e5c3c1070c45c710de39135e4f25378ef692f98c527d415731540f0d3985`<br>OFTYGR-S1T-S01-FOLLOWUP-RECOVERED-WORKBOOK<br>`d70444d2cf00fc14e2aab85947b12ce8f483a35f1206943f4298664dc5723f61`<br>OFTYGR-S1T-S01-FOLLOWUP-PREFILL-MANIFEST-AT122<br>`7c8574f184d0a0b0fb2929c9c9d76af347790ecca781c3f596796bea36813910`<br>OFTYGR-S1T-S01-PX-E1-AT122<br>`4e344c402c9089fe2ff96884823ab53c8194e6a84b7744bcb2e92f3398e0fd78`<br>OFTYGR-S1T-S01-PX-E2-AT122<br>`5493ed96d9287eab10a0c0f1751103296187daaa8834d2bcf332708dc7169d6c` |
| **committed_or_manifest_sha256** | `9a625a2292bf7804701211ddf8507253542e4b00128408d9c6c85ae5e0cac41e` |
| **prev_entry_sha256** | `4462bb66e4d115814171e8f28bf0387e4a02ac87594df136a6111c48106474fa` |
| **entry_line_sha256** | `80f37d945c15c8dc77945572aa63f409b870d661f13edf012d35eb33a2257e2d` |

| Entry 124 | 2026-10-05 · VERIFICATION_RECORD · manifest – |
|---|---|
| **governing_provisions** | P26-18; Codex follow-up corr1 check e288c9b3 |
| **subject** | OFTYGR-S1T-S01-FOLLOWUP-V013-LOG<br>`9a625a2292bf7804701211ddf8507253542e4b00128408d9c6c85ae5e0cac41e`<br>OFTYGR-S1T-S01-FOLLOWUP-TRANSCRIPTION-MANIFEST<br>`66f6ad7183a4c4d5d721bdc1589664515e0a62c7dbe0c2784b7a0df06a8c936f`<br>OFTYGR-S1T-S01-FOLLOWUP-HRD-ONLY-INPUT<br>`0dfb6bfc9b53b54d340737e7f8d089ca2c85a4ec67a75149644d02a352e0da5c`<br>OFTYGR-S1T-S01-FOLLOWUP-R3-PROVENANCE<br>`57f7a24d45b35e2f7c619ce016ff1ba5d5857c74967c7eb0c4b898111f54496c`<br>OFTYGR-S1T-S01-FOLLOWUP-R2-TRANSCRIPTION-INPUT<br>`1ba2e5c3c1070c45c710de39135e4f25378ef692f98c527d415731540f0d3985`<br>OFTYGR-S1T-S01-FOLLOWUP-RECOVERED-WORKBOOK<br>`d70444d2cf00fc14e2aab85947b12ce8f483a35f1206943f4298664dc5723f61`<br>OFTYGR-S1T-S01-FOLLOWUP-PREFILL-MANIFEST-AT122<br>`7c8574f184d0a0b0fb2929c9c9d76af347790ecca781c3f596796bea36813910`<br>OFTYGR-S1T-S01-PX-E1-AT122<br>`4e344c402c9089fe2ff96884823ab53c8194e6a84b7744bcb2e92f3398e0fd78`<br>OFTYGR-S1T-S01-PX-E2-AT122<br>`5493ed96d9287eab10a0c0f1751103296187daaa8834d2bcf332708dc7169d6c` |
| **committed_or_manifest_sha256** | – |
| **prev_entry_sha256** | `80f37d945c15c8dc77945572aa63f409b870d661f13edf012d35eb33a2257e2d` |
| **entry_line_sha256** | `e0cef05dec7fc7b38fc8d6b332f2bb434eb043ae9990199a9fdac5d8a56c1613` |

| Entry 125 | 2026-10-05 · FREEZE_ACT · manifest M0010 |
|---|---|
| **governing_provisions** | P26-18; P26-14; P26-02(vi) |
| **subject** | PG26-M0010<br>`c01a9d39c41f2deef02698cf8debc533ae2765c774780abeced073ad4ed863d6` |
| **committed_or_manifest_sha256** | `c01a9d39c41f2deef02698cf8debc533ae2765c774780abeced073ad4ed863d6` |
| **prev_entry_sha256** | `e0cef05dec7fc7b38fc8d6b332f2bb434eb043ae9990199a9fdac5d8a56c1613` |
| **entry_line_sha256** | `2035c12cd89a016d2af73d62317340eb038bca0474be465751482c50f37bb92e` |

| Entry 126 | 2026-10-05 · COMMITMENT · manifest – |
|---|---|
| **governing_provisions** | P26-02(iii) as amended by CA2-10/CA3-10; P26-14 |
| **subject** | PG26-M0010<br>`c01a9d39c41f2deef02698cf8debc533ae2765c774780abeced073ad4ed863d6` |
| **committed_or_manifest_sha256** | `c01a9d39c41f2deef02698cf8debc533ae2765c774780abeced073ad4ed863d6` |
| **prev_entry_sha256** | `2035c12cd89a016d2af73d62317340eb038bca0474be465751482c50f37bb92e` |
| **entry_line_sha256** | `b092d30e4a44c28e7f19c1c29ad91b69e96c3d8c82f67b461c9f396ef163eaa4` |

| Entry 127 | 2026-10-05 · VERIFICATION_RECORD · manifest – |
|---|---|
| **governing_provisions** | P26-18; P26-14; P26-12 |
| **subject** | PG26-M0010<br>`c01a9d39c41f2deef02698cf8debc533ae2765c774780abeced073ad4ed863d6` |
| **committed_or_manifest_sha256** | – |
| **prev_entry_sha256** | `b092d30e4a44c28e7f19c1c29ad91b69e96c3d8c82f67b461c9f396ef163eaa4` |
| **entry_line_sha256** | `8cc3ff1cb55b29aa7f099915ba6190d71020acad23de945d1524daf27d188a24` |

| Entry 128 | 2026-10-05 · COMMITMENT · manifest – |
|---|---|
| **governing_provisions** | M0010 LOCK_LT2 (AM-1 screen-3 bridge; freeze act, commitment and OR verification); Codex AM1 bridge check fcb1fd70; AM1 bridge proposal v0.1.1 CODEX 10d24c88; PI instruction record 20261005T1814Z (f7b1ac5a); D-1 v10 A-08 part (f) (ROLE_RECORD entry 111, verified 112); P26-14 |
| **subject** | OFTYGR-AM1-S3-BASIS-BASIS_MANIFEST.json<br>36256778a32d626fd6146ccf507e098b6ffa96983a6c19ae90e8c2f5a12724c8<br>OFTYGR-AM1-S3-BASIS-SCREEN3_UAZ_BASIS_01.json<br>7f26f814130fd8c75525efa14a1dd8458771afd50fef4f755db364ce9d028958<br>OFTYGR-AM1-S3-BASIS-SCREEN3_UAZ_BASIS_02.json<br>ff47b2f0ec06d47d1af891b7b832460e7c4c37dd4ea8a5a8f7e2c7317f7fe9d4<br>OFTYGR-AM1-S3-BASIS-SCREEN3_UAZ_BASIS_03.json<br>4f534a63e4508e52a621d629a3486e3b869c231487cb5fa33cdc89cb67e64234<br>OFTYGR-AM1-S3-BASIS-SCREEN3_UAZ_BASIS_04.json<br>c67a4986a8384f64a24374896c318c36461ddd8b26970e7aa3ffc3360fcd3b7b<br>OFTYGR-AM1-S3-BASIS-SCREEN3_UAZ_BASIS_05.json<br>b8c536c7d88a1f8a2a6a9d87f958f926c4abe3f26630b0cfed620be732053312<br>OFTYGR-AM1-S3-BASIS-SCREEN3_UAZ_BASIS_06.json<br>d052fbb26d4706da07b2ea7e9f42527d61aa7dc5dcc1586b631c4eab80bb1bb8<br>OFTYGR-AM1-S3-BASIS-SCREEN3_UAZ_BASIS_07.json<br>e59eb85f6d10fbaf3e8c476a8153ce8d1cf2be60f56c8e6e6847d72732ab0a6a |
| **committed_or_manifest_sha256** | 36256778a32d626fd6146ccf507e098b6ffa96983a6c19ae90e8c2f5a12724c8 |
| **prev_entry_sha256** | 8cc3ff1cb55b29aa7f099915ba6190d71020acad23de945d1524daf27d188a24 |
| **entry_line_sha256** | b63c854f0531abe62f5a8ad38bb871cee89ba3472d807e95e50ecf3ecfba40f8 |

| Entry 129 | 2026-10-05 · VERIFICATION_RECORD · manifest – |
|---|---|
| **governing_provisions** | P26-18; Codex AM1 bridge check fcb1fd70 |
| **subject** | OFTYGR-AM1-S3-BASIS-BASIS_MANIFEST.json<br>36256778a32d626fd6146ccf507e098b6ffa96983a6c19ae90e8c2f5a12724c8<br>OFTYGR-AM1-S3-BASIS-SCREEN3_UAZ_BASIS_01.json<br>7f26f814130fd8c75525efa14a1dd8458771afd50fef4f755db364ce9d028958<br>OFTYGR-AM1-S3-BASIS-SCREEN3_UAZ_BASIS_02.json<br>ff47b2f0ec06d47d1af891b7b832460e7c4c37dd4ea8a5a8f7e2c7317f7fe9d4<br>OFTYGR-AM1-S3-BASIS-SCREEN3_UAZ_BASIS_03.json<br>4f534a63e4508e52a621d629a3486e3b869c231487cb5fa33cdc89cb67e64234<br>OFTYGR-AM1-S3-BASIS-SCREEN3_UAZ_BASIS_04.json<br>c67a4986a8384f64a24374896c318c36461ddd8b26970e7aa3ffc3360fcd3b7b<br>OFTYGR-AM1-S3-BASIS-SCREEN3_UAZ_BASIS_05.json<br>b8c536c7d88a1f8a2a6a9d87f958f926c4abe3f26630b0cfed620be732053312<br>OFTYGR-AM1-S3-BASIS-SCREEN3_UAZ_BASIS_06.json<br>d052fbb26d4706da07b2ea7e9f42527d61aa7dc5dcc1586b631c4eab80bb1bb8<br>OFTYGR-AM1-S3-BASIS-SCREEN3_UAZ_BASIS_07.json<br>e59eb85f6d10fbaf3e8c476a8153ce8d1cf2be60f56c8e6e6847d72732ab0a6a |
| **committed_or_manifest_sha256** | – |
| **prev_entry_sha256** | b63c854f0531abe62f5a8ad38bb871cee89ba3472d807e95e50ecf3ecfba40f8 |
| **entry_line_sha256** | 65d30d0403b888935e15e5020c189e1aaf0d4e574bc5ade3f8517683e1f08907 |

| Entry 130 | 2026-10-05 · COMMITMENT · manifest – |
|---|---|
| **governing_provisions** | M0010 LOCK_LT2 (AM-1 screen-3 bridge; freeze act, commitment and OR verification); SCREEN3_UAZ_BASIS records (BC1/BC2); DV record (entry 121, verified 122); HRD partial log (entry 123, verified 124); Codex AM1 bridge check fcb1fd70; final Option-1 X-1 report OFTY-GR_v0.2_AM1_S3_Bridge_v0.3_Final_CODEX_Check_v1.0.md; AM1 bridge proposal v0.1.1 CODEX 10d24c88; PI instruction record 20261005T1814Z (f7b1ac5a); D-1 v10 A-08 part (f) (ROLE_RECORD entry 111, verified 112); P26-14 |
| **subject** | S1T-SESSION-01-V013-LOG-117<br>a53bbc8f9706cb4a256e17003cfdb4d4a37dc8ea4b76d2ee9685bf2644a783a2<br>S1T-SESSION-01-SUCCESSOR-TRANSCRIPTION-RESULT<br>3f788c58fecc281e29e1074c32da3e7f3960fd11b42b9985d6ffc26597ba2e87<br>S1T-SESSION-01-FROZEN-TRANSCRIBER-REFUSAL<br>46a3f68a6e1900348e56958017cfe87b7ae66d38fdcab9e66c74319914365a7d<br>S1T-SESSION-01-SUCCESSOR-VALIDATION-RESULT<br>062eb83e497ff0d00a8a5fd42fe965c4669b9ce5be18112cab0c55a303884f42<br>S1T-SESSION-01-FROZEN-VALIDATOR-REFUSAL<br>569b14a59168fca82cb361122e99874f2dac7b5f0b856565cef93f0c15269edb<br>S1T-SESSION-01-CARRIED-FORM<br>fab18764b078dd1aac7c7ab248f3665d40028929d89403e988a076c2040df1fd<br>S1T-SESSION-01-CARRY-PROVENANCE<br>4a195616c1a67501eba8ba14222375b1f41935cfa35d67bee4b57bea07cd8094<br>S1T-SESSION-01-HRD-RETURN-E1-corr1<br>b8cd3f7c7c1795d587ba11d6e47fe4a815b0f07e8aca75449105cdf32635a92b<br>S1T-SESSION-01-HRD-RETURN-E2-corr1<br>d804dd661b55c27572a7732d3f1511375500e23fbae4092e517216c07d0bf0c2<br>S1T-SESSION-01-AM1-FULL-PENDING-SOURCE<br>e272bdc851059874fcbd95c4f7cdf31fcfb44ac20363272342295465ca74368c<br>S1T-SESSION-01-AM1-R3-PROVENANCE<br>57f7a24d45b35e2f7c619ce016ff1ba5d5857c74967c7eb0c4b898111f54496c<br>S1T-SESSION-01-PREFILL-POST-BC2-FORM<br>ff88e98f55e84830b15fb5218bbaef797bfb1be9f19da2b39a3762b9aca5665f<br>S1T-SESSION-01-PREFILL-POST-BC2-MANIFEST<br>c6a9d6cd2b3813f7e1214434a704c4c432196b5408e2ff997d7a31ed17e0bf4a<br>PX-E1-S1T-SESSION-01-AT129<br>1080c4d33c8ee0069cd9f9199a678c242e3ce1af721cf3f3b8d4e22abb79e785<br>PX-E2-S1T-SESSION-01-AT129<br>01c8c827e6707a08aa27e114b11f7e565cc2e1a13a8eaf1cc73ebdc82e04bb2c<br>OFTYGR-AM1-S3-BASIS-MANIFEST<br>36256778a32d626fd6146ccf507e098b6ffa96983a6c19ae90e8c2f5a12724c8<br>CERT-BC1-SCREEN3-UAZ-BASIS-ENTRY-128<br>e89ce95bad6240b4b7ca225cc8b7165cb1b0d57084c228c67c83af6c784fd427 |
| **committed_or_manifest_sha256** | a53bbc8f9706cb4a256e17003cfdb4d4a37dc8ea4b76d2ee9685bf2644a783a2 |
| **prev_entry_sha256** | 65d30d0403b888935e15e5020c189e1aaf0d4e574bc5ade3f8517683e1f08907 |
| **entry_line_sha256** | adce47a1c7da3af4790ff7bf35ac1bbfb282a4feb9464c506b288dc4c6e71498 |

| Entry 131 | 2026-10-05 · VERIFICATION_RECORD · manifest – |
|---|---|
| **governing_provisions** | P26-18; Codex AM1 bridge check fcb1fd70; final Option-1 X-1 report OFTY-GR_v0.2_AM1_S3_Bridge_v0.3_Final_CODEX_Check_v1.0.md |
| **subject** | S1T-SESSION-01-V013-LOG-117<br>`a53bbc8f9706cb4a256e17003cfdb4d4a37dc8ea4b76d2ee9685bf2644a783a2`<br>S1T-SESSION-01-SUCCESSOR-TRANSCRIPTION-RESULT<br>`3f788c58fecc281e29e1074c32da3e7f3960fd11b42b9985d6ffc26597ba2e87`<br>S1T-SESSION-01-FROZEN-TRANSCRIBER-REFUSAL<br>`46a3f68a6e1900348e56958017cfe87b7ae66d38fdcab9e66c74319914365a7d`<br>S1T-SESSION-01-SUCCESSOR-VALIDATION-RESULT<br>`062eb83e497ff0d00a8a5fd42fe965c4669b9ce5be18112cab0c55a303884f42`<br>S1T-SESSION-01-FROZEN-VALIDATOR-REFUSAL<br>`569b14a59168fca82cb361122e99874f2dac7b5f0b856565cef93f0c15269edb`<br>S1T-SESSION-01-CARRIED-FORM<br>`fab18764b078dd1aac7c7ab248f3665d40028929d89403e988a076c2040df1fd`<br>S1T-SESSION-01-CARRY-PROVENANCE<br>`4a195616c1a67501eba8ba14222375b1f41935cfa35d67bee4b57bea07cd8094`<br>S1T-SESSION-01-HRD-RETURN-E1-corr1<br>`b8cd3f7c7c1795d587ba11d6e47fe4a815b0f07e8aca75449105cdf32635a92b`<br>S1T-SESSION-01-HRD-RETURN-E2-corr1<br>`d804dd661b55c27572a7732d3f1511375500e23fbae4092e517216c07d0bf0c2`<br>S1T-SESSION-01-AM1-FULL-PENDING-SOURCE<br>`e272bdc851059874fcbd95c4f7cdf31fcfb44ac20363272342295465ca74368c`<br>S1T-SESSION-01-AM1-R3-PROVENANCE<br>`57f7a24d45b35e2f7c619ce016ff1ba5d5857c74967c7eb0c4b898111f54496c`<br>S1T-SESSION-01-PREFILL-POST-BC2-FORM<br>`ff88e98f55e84830b15fb5218bbaef797bfb1be9f19da2b39a3762b9aca5665f`<br>S1T-SESSION-01-PREFILL-POST-BC2-MANIFEST<br>`c6a9d6cd2b3813f7e1214434a704c4c432196b5408e2ff997d7a31ed17e0bf4a`<br>PX-E1-S1T-SESSION-01-AT129<br>`1080c4d33c8ee0069cd9f9199a678c242e3ce1af721cf3f3b8d4e22abb79e785`<br>PX-E2-S1T-SESSION-01-AT129<br>`01c8c827e6707a08aa27e114b11f7e565cc2e1a13a8eaf1cc73ebdc82e04bb2c`<br>OFTYGR-AM1-S3-BASIS-MANIFEST<br>`36256778a32d626fd6146ccf507e098b6ffa96983a6c19ae90e8c2f5a12724c8`<br>CERT-BC1-SCREEN3-UAZ-BASIS-ENTRY-128<br>`e89ce95bad6240b4b7ca225cc8b7165cb1b0d57084c228c67c83af6c784fd427` |
| **committed_or_manifest_sha256** | – |
| **prev_entry_sha256** | `adce47a1c7da3af4790ff7bf35ac1bbfb282a4feb9464c506b288dc4c6e71498` |
| **entry_line_sha256** | `6d31c090f41817ebca9a397da815cf5a9179fcdd15e797601708d176f7793399` |

| Entry 132 | 2026-10-05 · COMMITMENT · manifest – |
|---|---|
| **governing_provisions** | M0010 LOCK_LT2 (entries 125-127); SCREEN3_UAZ_BASIS records (BC1/BC2, entries 128/129); combined AM-1/HRD-proposal transcription log (AC1/AC2, entries 130/131); Codex HRD assessment package check 89ddd1ca; PI instruction records 20261005T2107Z (0fe12f24) and 20261005T2120Z (7cc64a9d); X-1 report ae89185ccde195b837901d208abdf32a400f770423bb671cd0e20f773843d8d9 (principal) and supplement 2db0c9ecaee71df516a75a2de720b6948bf9ead788d0b0e2e7c7af1cce04934f; D-1 v10 A-08 part (f) (ROLE_RECORD entry 111, verified 112); P26-14 |
| **subject** | S1T-SESSION-01-V013-LOG-HRD-ASSESSMENTS<br>9b0d14f8229f5ee55f1209de4442e4cac943dc5f088c956bc26d609e83bdd548<br>S1T-SESSION-01-HRD-ASSESSMENT-TRANSCRIPTION-RESULT<br>916dce80b98f603d4f4dd3858bbfa4a4d2f57752942b6a541e425b5462321b64<br>S1T-SESSION-01-HRD-ASSESSMENT-FROZEN-REFUSAL<br>19fb64aae70251ce5fbf2bd67f0b9d8aa76a79b2982014632a234190e3387b30<br>S1T-SESSION-01-HRD-ASSESSMENT-VALIDATION-RESULT<br>7299d7fd14f16fef0aa512a90813743dfc20f523f3c60ef5d0ca8c9358186ece<br>S1T-SESSION-01-HRD-ASSESSMENT-MERGED-FORM<br>37b524361cf66e349a93448a9b89d01323b78a1d077c5986c166403731c931be<br>S1T-SESSION-01-HRD-ASSESSMENT-MERGE-PROVENANCE<br>798df7b8705874b242956245e10d81c7ed803a4244cd15563cb10b916da8954f<br>S1T-SESSION-01-HRD-ASSESSMENT-RETURN-E1-corr2<br>d14f788c26d9f1654e06d0c305b2a33458ccc17446492f6d07bd1c5ecb02e3ab<br>S1T-SESSION-01-HRD-ASSESSMENT-RETURN-E2-corr1<br>09916426be9183d17371fe333a8b388188569cc2cfd46a04ab2fbd9339d40a4f<br>S1T-SESSION-01-HRD-ASSESSMENT-RETURN-E1-corr1-PREDECESSOR<br>9fb4235dca7faca94b46e16dd7f10fd0e8aa538ea5acc7fe25d43b0b61b4e72b<br>S1T-SESSION-01-HRD-ASSESSMENT-RETURN-E2-PREDECESSOR<br>ea55b651a060d7c99d10e549fb1260b8f17400a52950edba572a1c2e16f330ba<br>S1T-SESSION-01-PTR011-CORRECTIONS-RECEIPT<br>ef0557ccd175ffce281e2bf2b19350531dddbc4064b2fe404790129080a55015<br>S1T-SESSION-01-E1-corr2-RECEIPT<br>8b87a4b18dc39169e6a4d71a819ff0930e8753d0419cc1b2fc726b97d8ef812a<br>S1T-SESSION-01-PTR011-CORRECTIONS-SUPPLEMENT<br>2db0c9ecaee71df516a75a2de720b6948bf9ead788d0b0e2e7c7af1cce04934f<br>S1T-SESSION-01-PI-RECORD-20261005T2153Z<br>a6827abe55964bb165ce6bdf286c4e8b54dc1710993df7a4e0256bd55455f2c1<br>S1T-SESSION-01-R1-INPUT-VERIFICATION<br>083d77b214fcee4c1034578b43ab8409baeb8ab924b34c23e70a594f623fdb79<br>PX-E1-S1T-SESSION-01-AT131<br>3ea73b7d7eedcfcb32cf2f555ab4eb4e84e03e43727a82f1279f23084b093c10<br>PX-E2-S1T-SESSION-01-AT131<br>e91448b31e2ed28031c2fe3849e2ffe6dadc5e5747c9176668dd22dba872a914<br>OFTYGR-AM1-S3-BASIS-MANIFEST<br>36256778a32d626fd6146ccf507e098b6ffa96983a6c19ae90e8c2f5a12724c8 |
| **committed_or_manifest_sha256** | 9b0d14f8229f5ee55f1209de4442e4cac943dc5f088c956bc26d609e83bdd548 |
| **prev_entry_sha256** | 6d31c090f41817ebca9a397da815cf5a9179fcdd15e797601708d176f7793399 |
| **entry_line_sha256** | d8d2c58e91ebfed5420afbb19e38b1a3f434398aa96bd253e9ab98199861d01a |

| **Entry 133** | 2026-10-05 · VERIFICATION_RECORD · manifest – |
|---|---|
| **governing_provisions** | P26-18; X-1 report ae89185ccde195b837901d208abdf32a400f770423bb671cd0e20f773843d8d9 (principal) and supplement 2db0c9ecaee71df516a75a2de720b6948bf9ead788d0b0e2e7c7af1cce04934f |
| **subject** | S1T-SESSION-01-V013-LOG-HRD-ASSESSMENTS<br>9b0d14f8229f5ee55f1209de4442e4cac943dc5f088c956bc26d609e83bdd548<br>S1T-SESSION-01-HRD-ASSESSMENT-TRANSCRIPTION-RESULT<br>916dce80b98f603d4f4dd3858bbfa4a4d2f57752942b6a541e425b5462321b64<br>S1T-SESSION-01-HRD-ASSESSMENT-FROZEN-REFUSAL<br>19fb64aae70251ce5fbf2bd67f0b9d8aa76a79b2982014632a234190e3387b30<br>S1T-SESSION-01-HRD-ASSESSMENT-VALIDATION-RESULT<br>7299d7fd14f16fef0aa512a90813743dfc20f523f3c60ef5d0ca8c9358186ece<br>S1T-SESSION-01-HRD-ASSESSMENT-MERGED-FORM<br>37b524361cf66e349a93448a9b89d01323b78a1d077c5986c166403731c931be<br>S1T-SESSION-01-HRD-ASSESSMENT-MERGE-PROVENANCE<br>798df7b8705874b242956245e10d81c7ed803a4244cd15563cb10b916da8954f<br>S1T-SESSION-01-HRD-ASSESSMENT-RETURN-E1-corr2<br>d14f788c26d9f1654e06d0c305b2a33458ccc17446492f6d07bd1c5ecb02e3ab<br>S1T-SESSION-01-HRD-ASSESSMENT-RETURN-E2-corr1<br>09916426be9183d17371fe333a8b388188569cc2cfd46a04ab2fbd9339d40a4f<br>S1T-SESSION-01-HRD-ASSESSMENT-RETURN-E1-corr1-PREDECESSOR<br>9fb4235dca7faca94b46e16dd7f10fd0e8aa538ea5acc7fe25d43b0b61b4e72b<br>S1T-SESSION-01-HRD-ASSESSMENT-RETURN-E2-PREDECESSOR<br>ea55b651a060d7c99d10e549fb1260b8f17400a52950edba572a1c2e16f330ba<br>S1T-SESSION-01-PTR011-CORRECTIONS-RECEIPT<br>ef0557ccd175ffce281e2bf2b19350531dddbc4064b2fe404790129080a55015<br>S1T-SESSION-01-E1-corr2-RECEIPT<br>8b87a4b18dc39169e6a4d71a819ff0930e8753d0419cc1b2fc726b97d8ef812a<br>S1T-SESSION-01-PTR011-CORRECTIONS-SUPPLEMENT<br>2db0c9ecaee71df516a75a2de720b6948bf9ead788d0b0e2e7c7af1cce04934f<br>S1T-SESSION-01-PI-RECORD-20261005T2153Z<br>a6827abe55964bb165ce6bdf286c4e8b54dc1710993df7a4e0256bd55455f2c1<br>S1T-SESSION-01-R1-INPUT-VERIFICATION<br>083d77b214fcee4c1034578b43ab8409baeb8ab924b34c23e70a594f623fdb79<br>PX-E1-S1T-SESSION-01-AT131<br>3ea73b7d7eedcfcb32cf2f555ab4eb4e84e03e43727a82f1279f23084b093c10<br>PX-E2-S1T-SESSION-01-AT131<br>e91448b31e2ed28031c2fe3849e2ffe6dadc5e5747c9176668dd22dba872a914<br>OFTYGR-AM1-S3-BASIS-MANIFEST<br>36256778a32d626fd6146ccf507e098b6ffa96983a6c19ae90e8c2f5a12724c8 |
| **committed_or_manifest_sha256** | – |
| **prev_entry_sha256** | d8d2c58e91ebfed5420afbb19e38b1a3f434398aa96bd253e9ab98199861d01a |
| **entry_line_sha256** | ffd1c0316a041589414db3a5d6136f9687bdef80c02600cedd96791fde91c94b |

| Entry 134 | 2026-10-06 · COMMITMENT · manifest – |
|---|---|
| **governing_provisions** | M0010 LOCK_LT2 (entries 125-127); SCREEN3_UAZ_BASIS records (BC1/BC2, entries 128/129); HRD floor-assessment transcription log (HA1/HA2, entries 132/133); Codex HRD joint-comparison return check cd6452ee; Effective bilateral declaration and PI instruction record 20261005T2256Z (42c8b0e4); PI instruction record 20261005T2301Z (f98d6491); PI authorisation record 20261005T2303Z (287998b9); X-1 report 19de71be40eaaa562c99bfffda40e2a350fe40e4dce1c9f0addbed585374bfac (Codex HRD confronto and dossier check v1.0, conditions R-1 to R-7); D-1 v10 A-08 part (f) (ROLE_RECORD entry 111, verified 112); P26-14 |
| **subject** | S1T-SESSION-01-V013-LOG-HRD-CONFRONTO<br>`07ab35499b99c5e4ac93634d34db9e8c46229cb09f64ecf5310012a157cbda2d`<br>S1T-SESSION-01-HRD-CONFRONTO-TRANSCRIPTION-RESULT<br>`ef28eb1a59645c1ab4aec14bf76054dbee75741881bc8fdf3701c3ac69647ec7`<br>S1T-SESSION-01-HRD-CONFRONTO-FROZEN-REFUSAL<br>`19fb64aae70251ce5fbf2bd67f0b9d8aa76a79b2982014632a234190e3387b30`<br>S1T-SESSION-01-HRD-CONFRONTO-VALIDATION-RESULT<br>`fd3c78b2208cbff64cfbea395674a5c015015f871d52205f78a1b29454602397`<br>S1T-SESSION-01-HRD-CONFRONTO-VALIDATION-FROZEN-REFUSAL<br>`569b14a59168fca82cb361122e99874f2dac7b5f0b856565cef93f0c15269edb`<br>S1T-SESSION-01-HRD-CONFRONTO-UNION-FORM<br>`294d105a377948ab72e7dd3f3b4f37633ef47e4ae7fcbdbfdd854ea30b5b4e14`<br>S1T-SESSION-01-HRD-CONFRONTO-UNION-PROVENANCE<br>`2ab884d499db674cba132b4576fc95b2628e35cf1cd3965808190453789c8015`<br>S1T-SESSION-01-HRD-CONFRONTO-REGOLE-DOCUMENTARY-CONTENT<br>`b163f4219fd17070008ead817e8fc58039b97bfb1d0ad3a63f15950514b48680`<br>S1T-SESSION-01-HRD-CONFRONTO-JOINT-DERIVED<br>`d31be85f21b31152a144860d9e981b2b11ec1f51e4f01f763a831bfbe9090490`<br>S1T-SESSION-01-HRD-CONFRONTO-DERIVATION-PROVENANCE<br>`f9a1a187c0726252e16b978268bb0adc14691e2a13f9ba92f2be46dd1ebfd292`<br>S1T-SESSION-01-HRD-CONFRONTO-JOINT-RETURN<br>`3a463063b555bc8ef674f671739dd4a47f6fc12b16e5c4034d6c212090a5cb28`<br>S1T-SESSION-01-HRD-CONFRONTO-SEARCH-RETURN-E1<br>`3df8ea8ee10b2e5d20bd9ba031538aa63bc4ee43c3a06870ed82782d64574c69`<br>S1T-SESSION-01-HRD-CONFRONTO-SEARCH-RETURN-E2<br>`671ad3a4ded6f6bd2f70c69dc741e21fd9deaab65fefbbfad6144f4565005b2c`<br>S1T-SESSION-01-HRD-CONFRONTO-VERBALE<br>`7dc7154654a2cfbf4d419e14d125faab6b39b97448cf5747044076fa0ff3db9e`<br>S1T-SESSION-01-HRD-CONFRONTO-RETURN-ZIP<br>`dae3c7467b5e043c75695de5b38a93b2ea04ebf01a9aaef95d45db92e487ac16`<br>S1T-SESSION-01-HRD-CONFRONTO-LM-DECLARATION-RECORD<br>`42c8b0e456fce9526d799e4e2d84f869e693564fc8658de419614c142b92cb78`<br>S1T-SESSION-01-HRD-CONFRONTO-RETURN-CODEX-CHECK<br>`cd6452ee6088565d3ff1c35299c31f0adbcb8b45d9db12c4c3fcc14249a6fd11`<br>S1T-SESSION-01-HRD-CONFRONTO-RETURN-CODEX-AUDIT<br>`84ba4cc1f508784b8a1f5bc01b7090140ce0b32d7599af9398bd1b9d3865d5da`<br>S1T-SESSION-01-HRD-CONFRONTO-X1-REPORT<br>`19de71be40eaaa562c99bfffda40e2a350fe40e4dce1c9f0addbed585374bfac`<br>S1T-SESSION-01-PI-RECORD-20261005T2301Z<br>`f98d649148d1f5e5c7f07fbb67fc263fe848098f8a67aeaba80997bdf0661c4b`<br>S1T-SESSION-01-PI-RECORD-20261006T0952Z<br>`7158852194efd9f78026e7534373e05075ac19e6d2b02457a24d17ab27013646`<br>S1T-SESSION-01-HRD-CONFRONTO-A3-INDEPENDENT-CHECKS<br>`b1d15a3ea6252020432f004ce6c9425fe542d8f6b1442f0daaf215473bf5da3d`<br>S1T-SESSION-01-HRD-CONFRONTO-LIMITATIONS<br>`dcdc41b9da6847539f13a897afb5769972565a1add74b3c3a391f682e79e3b14`<br>PX-E1-S1T-SESSION-01-AT133<br>`7a0e3f2ea2a83933dbe15ceaae8b948e22cb988a323cf264fe5e149d8363823d`<br>PX-E2-S1T-SESSION-01-AT133<br>`1c0e4aa7d9fde71b82d10e98fd9052a8a7fccf21c8ccd7102aa89706cc1ba300`<br>OFTYGR-AM1-S3-BASIS-MANIFEST<br>`36256778a32d626fd6146ccf507e098b6ffa96983a6c19ae90e8c2f5a12724c8` |
| **committed_or_manifest_sha256** | `07ab35499b99c5e4ac93634d34db9e8c46229cb09f64ecf5310012a157cbda2d` |
| **prev_entry_sha256** | `ffd1c0316a041589414db3a5d6136f9687bdef80c02600cedd96791fde91c94b` |
| **entry_line_sha256** | `70fc7f396ff2de8ff0a61e738d7ebf4094436fddf0e77e187a679225e32b4e64` |

| **Entry 135** | 2026-10-06 · VERIFICATION_RECORD · manifest – |
|---|---|
| **governing_provisions** | P26-18; X-1 report 19de71be40eaaa562c99bfffda40e2a350fe40e4dce1c9f0addbed585374bfac (Codex HRD confronto and dossier check v1.0, conditions R-1 to R-7) |
| **subject** | S1T-SESSION-01-V013-LOG-HRD-CONFRONTO<br>`07ab35499b99c5e4ac93634d34db9e8c46229cb09f64ecf5310012a157cbda2d`<br>S1T-SESSION-01-HRD-CONFRONTO-TRANSCRIPTION-RESULT<br>`ef28eb1a59645c1ab4aec14bf76054dbee75741881bc8fdf3701c3ac69647ec7`<br>S1T-SESSION-01-HRD-CONFRONTO-FROZEN-REFUSAL<br>`19fb64aae70251ce5fbf2bd67f0b9d8aa76a79b2982014632a234190e3387b30`<br>S1T-SESSION-01-HRD-CONFRONTO-VALIDATION-RESULT<br>`fd3c78b2208cbff64cfbea395674a5c015015f871d52205f78a1b29454602397`<br>S1T-SESSION-01-HRD-CONFRONTO-VALIDATION-FROZEN-REFUSAL<br>`569b14a59168fca82cb361122e99874f2dac7b5f0b856565cef93f0c15269edb`<br>S1T-SESSION-01-HRD-CONFRONTO-UNION-FORM<br>`294d105a377948ab72e7dd3f3b4f37633ef47e4ae7fcbdbfdd854ea30b5b4e14`<br>S1T-SESSION-01-HRD-CONFRONTO-UNION-PROVENANCE<br>`2ab884d499db674cba132b4576fc95b2628e35cf1cd3965808190453789c8015`<br>S1T-SESSION-01-HRD-CONFRONTO-REGOLE-DOCUMENTARY-CONTENT<br>`b163f4219fd17070008ead817e8fc58039b97bfb1d0ad3a63f15950514b48680`<br>S1T-SESSION-01-HRD-CONFRONTO-JOINT-DERIVED<br>`d31be85f21b31152a144860d9e981b2b11ec1f51e4f01f763a831bfbe9090490`<br>S1T-SESSION-01-HRD-CONFRONTO-DERIVATION-PROVENANCE<br>`f9a1a187c0726252e16b978268bb0adc14691e2a13f9ba92f2be46dd1ebfd292`<br>S1T-SESSION-01-HRD-CONFRONTO-JOINT-RETURN<br>`3a463063b555bc8ef674f671739dd4a47f6fc12b16e5c4034d6c212090a5cb28`<br>S1T-SESSION-01-HRD-CONFRONTO-SEARCH-RETURN-E1<br>`3df8ea8ee10b2e5d20bd9ba031538aa63bc4ee43c3a06870ed82782d64574c69`<br>S1T-SESSION-01-HRD-CONFRONTO-SEARCH-RETURN-E2<br>`671ad3a4ded6f6bd2f70c69dc741e21fd9deaab65fefbbfad6144f4565005b2c`<br>S1T-SESSION-01-HRD-CONFRONTO-VERBALE<br>`7dc7154654a2cfbf4d419e14d125faab6b39b97448cf5747044076fa0ff3db9e`<br>S1T-SESSION-01-HRD-CONFRONTO-RETURN-ZIP<br>`dae3c7467b5e043c75695de5b38a93b2ea04ebf01a9aaef95d45db92e487ac16`<br>S1T-SESSION-01-HRD-CONFRONTO-LM-DECLARATION-RECORD<br>`42c8b0e456fce9526d799e4e2d84f869e693564fc8658de419614c142b92cb78`<br>S1T-SESSION-01-HRD-CONFRONTO-RETURN-CODEX-CHECK<br>`cd6452ee6088565d3ff1c35299c31f0adbcb8b45d9db12c4c3fcc14249a6fd11`<br>S1T-SESSION-01-HRD-CONFRONTO-RETURN-CODEX-AUDIT<br>`84ba4cc1f508784b8a1f5bc01b7090140ce0b32d7599af9398bd1b9d3865d5da`<br>S1T-SESSION-01-HRD-CONFRONTO-X1-REPORT<br>`19de71be40eaaa562c99bfffda40e2a350fe40e4dce1c9f0addbed585374bfac`<br>S1T-SESSION-01-PI-RECORD-20261005T2301Z<br>`f98d649148d1f5e5c7f07fbb67fc263fe848098f8a67aeaba80997bdf0661c4b`<br>S1T-SESSION-01-PI-RECORD-20261006T0952Z<br>`7158852194efd9f78026e7534373e05075ac19e6d2b02457a24d17ab27013646`<br>S1T-SESSION-01-HRD-CONFRONTO-A3-INDEPENDENT-CHECKS<br>`b1d15a3ea6252020432f004ce6c9425fe542d8f6b1442f0daaf215473bf5da3d`<br>S1T-SESSION-01-HRD-CONFRONTO-LIMITATIONS<br>`dcdc41b9da6847539f13a897afb5769972565a1add74b3c3a391f682e79e3b14`<br>PX-E1-S1T-SESSION-01-AT133<br>`7a0e3f2ea2a83933dbe15ceaae8b948e22cb988a323cf264fe5e149d8363823d`<br>PX-E2-S1T-SESSION-01-AT133<br>`1c0e4aa7d9fde71b82d10e98fd9052a8a7fccf21c8ccd7102aa89706cc1ba300`<br>OFTYGR-AM1-S3-BASIS-MANIFEST<br>`36256778a32d626fd6146ccf507e098b6ffa96983a6c19ae90e8c2f5a12724c8` |
| **committed_or_manifest_sha256** | – |
| **prev_entry_sha256** | `70fc7f396ff2de8ff0a61e738d7ebf4094436fddf0e77e187a679225e32b4e64` |
| **entry_line_sha256** | `b8767bac9fd6f8231c0a260183adc050aa83eea81e52348fdd508fd5a0564364` |

| **Entry 136** | 2026-10-06 · COMMITMENT · manifest – |
|---|---|
| **governing_provisions** | P26-18; AM-S5-LIM; X-1 report 062826a2ab5c1dd166f0701ce4cf297f05a358402294dccd16a254e802e19dad |
| **subject** | S5-PI-ACT<br>`b79983398339bbce9c2b714dd635dbfec3004176823fce097be2f2df0dc5f230`<br>S5-ACT-TEXT<br>`54406cc27d6935a9641ed70c9fa4e93f2da9ebe365f6cb3daf8902c090716905`<br>S5-LIMITATIONS-AMENDMENT<br>`09055ce2fde32ec2fd31cac99bd26b0e574ff65fbf7e7d2ef8dfedf067fc8d25`<br>S5-CHANGE-RECORD<br>`75afd5feac66dc87ab96fb9d9e9cb3ffa89604b65b78cba50372ef88ba8fcb0f`<br>S5-CHANGE-IMPACT-INVENTORY<br>`deadfcfeb52685d4539f6d7453e123df31e4ff4de67fffbd223eab5d9d116ae9`<br>FPA-1.Charter-BASELINE<br>`2dce0ad81c93314f0c409edea2af582f0750137ebd06c9f2a1f6c77d6420ae24`<br>FPA-1.PG11-BASELINE<br>`9ef6450a40ddb945bda7e7a867b1226ba32d413677e4e90597a1acee386d4b4b` |
| **committed_or_manifest_sha256** | `b79983398339bbce9c2b714dd635dbfec3004176823fce097be2f2df0dc5f230` |
| **prev_entry_sha256** | `b8767bac9fd6f8231c0a260183adc050aa83eea81e52348fdd508fd5a0564364` |
| **entry_line_sha256** | `21a5f6b9ebc469834daac0f347a8e93f0fae3b8adb261220ba6b441ee8021641` |

| | |
|---|---|
| **Entry 137** | 2026-10-06 · VERIFICATION_RECORD · manifest – |
| **governing_provisions** | P26-18; AM-S5-LIM; X-1 report 062826a2ab5c1dd166f0701ce4cf297f05a358402294dccd16a254e802e19dad |
| **subject** | S5-PI-ACT<br>b79983398339bbce9c2b714dd635dbfec3004176823fce097be2f2df0dc5f230<br>S5-ACT-TEXT<br>54406cc27d6935a9641ed70c9fa4e93f2da9ebe365f6cb3daf8902c090716905<br>S5-LIMITATIONS-AMENDMENT<br>09055ce2fde32ec2fd31cac99bd26b0e574ff65fbf7e7d2ef8dfedf067fc8d25<br>S5-CHANGE-RECORD<br>75afd5feac66dc87ab96fb9d9e9cb3ffa89604b65b78cba50372ef88ba8fcb0f<br>S5-CHANGE-IMPACT-INVENTORY<br>deadfcfeb52685d4539f6d7453e123df31e4ff4de67fffbd223eab5d9d116ae9<br>FPA-1.Charter-BASELINE<br>2dce0ad81c93314f0c409edea2af582f0750137ebd06c9f2a1f6c77d6420ae24<br>FPA-1.PG11-BASELINE<br>9ef6450a40ddb945bda7e7a867b1226ba32d413677e4e90597a1acee386d4b4b |
| **committed_or_manifest_sha256** | – |
| **prev_entry_sha256** | 21a5f6b9ebc469834daac0f347a8e93f0fae3b8adb261220ba6b441ee8021641 |
| **entry_line_sha256** | 70343835bb0ba7dc30208b4bbb25619df1e38580bfd437fb82d5bb7292521193 |

| | |
|---|---|
| **Entry 138** | 2026-10-06 · FREEZE_ACT · manifest M0011 |
| **governing_provisions** | P26-18; P26-14; P26-02(vi) |
| **subject** | PG26-M0011<br>02ffc74456ad1a3dad834f44fa9ef2dfa6a3347b2a466cae1baee9c069d3814a |
| **committed_or_manifest_sha256** | 02ffc74456ad1a3dad834f44fa9ef2dfa6a3347b2a466cae1baee9c069d3814a |
| **prev_entry_sha256** | 70343835bb0ba7dc30208b4bbb25619df1e38580bfd437fb82d5bb7292521193 |
| **entry_line_sha256** | fc91f8377fab512aa2c238956a7ba9480ad900189a5405a71512409d8d319323 |

| | |
|---|---|
| **Entry 139** | 2026-10-06 · COMMITMENT · manifest – |
| **governing_provisions** | P26-02(iii) as amended by CA2-10/CA3-10; P26-14 |
| **subject** | PG26-M0011<br>02ffc74456ad1a3dad834f44fa9ef2dfa6a3347b2a466cae1baee9c069d3814a |
| **committed_or_manifest_sha256** | 02ffc74456ad1a3dad834f44fa9ef2dfa6a3347b2a466cae1baee9c069d3814a |
| **prev_entry_sha256** | fc91f8377fab512aa2c238956a7ba9480ad900189a5405a71512409d8d319323 |
| **entry_line_sha256** | 724acca2ce3c871234a42627fc1a9631f0eac1d5a6b28c63ae260938f36d283f |

| | |
|---|---|
| **Entry 140** | 2026-10-06 · VERIFICATION_RECORD · manifest – |
| **governing_provisions** | P26-18; P26-14; P26-12 |
| **subject** | PG26-M0011<br>02ffc74456ad1a3dad834f44fa9ef2dfa6a3347b2a466cae1baee9c069d3814a |
| **committed_or_manifest_sha256** | – |
| **prev_entry_sha256** | 724acca2ce3c871234a42627fc1a9631f0eac1d5a6b28c63ae260938f36d283f |
| **entry_line_sha256** | 966581431f3a4d2546e3d616e5d8b04f24df56c71dbf035062f2deb0c78981b1 |

| | |
|---|---|
| **Entry 141** | 2026-10-06 · COMMITMENT · manifest – |
| **governing_provisions** | PG26-M0011 freeze138 commitment139 verification140; PI act b79983398339bbce9c2b714dd635dbfec3004176823fce097be2f2df0dc5f230 / 54406cc27d6935a9641ed70c9fa4e93f2da9ebe365f6cb3daf8902c090716905; Codex 712afa62 / spec 10779562; X-1 report 2279558afd2ca9b3e3293f9338196ed9232192a383155c569f602a8d19049e6f; D-1 v10 A-08 part (f) (ROLE_RECORD entry 111, verified 112); P26-14 |
| **subject** | S1T-SESSION-01-S5-SIDE-LOG<br>1f6109d033e22db1d79e993f974d16e6b9614316cac2c64984d360dff391e9b5<br>S1T-SESSION-01-S5-RESULT<br>fdaf893e4b1eab2fc71203f71324be98e2a2cf56470f75a046e88e02cae0bd1f<br>S1T-SESSION-01-SOURCE-INVENTORY<br>15db611f892ed9f8d5c1c9c2d2682f87da3ba135147669f3707db3bfe66e7d75 |
| **committed_or_manifest_sha256** | 1f6109d033e22db1d79e993f974d16e6b9614316cac2c64984d360dff391e9b5 |
| **prev_entry_sha256** | 966581431f3a4d2546e3d616e5d8b04f24df56c71dbf035062f2deb0c78981b1 |
| **entry_line_sha256** | 137a91b4f359f96aebf07f89a185f2933b7d0ff1c2da3d34c69c55434a6be7cb |

| | |
|---|---|
| **Entry 142** | 2026-10-06 · VERIFICATION_RECORD · manifest – |
| **governing_provisions** | P26-18; X-1 report 105dc391867960bbabf7358ffb27c502f906688ff1276fb77a09643ee69f62ed |
| **subject** | S1T-SESSION-01-S5-SIDE-LOG<br>1f6109d033e22db1d79e993f974d16e6b9614316cac2c64984d360dff391e9b5<br>S1T-SESSION-01-S5-RESULT<br>fdaf893e4b1eab2fc71203f71324be98e2a2cf56470f75a046e88e02cae0bd1f<br>S1T-SESSION-01-SOURCE-INVENTORY<br>15db611f892ed9f8d5c1c9c2d2682f87da3ba135147669f3707db3bfe66e7d75 |
| **committed_or_manifest_sha256** | – |
| **prev_entry_sha256** | 137a91b4f359f96aebf07f89a185f2933b7d0ff1c2da3d34c69c55434a6be7cb |
| **entry_line_sha256** | d9c94f6ac324777bc851d4f6de5dc61b778850117cca1741d43d82f84f12bd97 |

| Entry 143 | 2026-10-06 · COMMITMENT · manifest – |
|---|---|
| **governing_provisions** | PG26-M0011, FREEZE_ACT 138, COMMITMENT 139, VERIFICATION_RECORD 140; PI act b79983398339bbce9c2b714dd635dbfec3004176823fce097be2f2df0dc5f230 / 54406cc27d6935a9641ed70c9fa4e93f2da9ebe365f6cb3daf8902c090716905; committed 136, verified 137; Codex 712afa62 / spec 10779562; X-1 report 2e970475fc54985d95d1a6f1d9b9b56130af05bb8652648d6477d1e10656e00f; D-1 v10 A-08 part (f) (ROLE_RECORD entry 111, verified 112); P26-14 |
| **subject** | S1T-SESSION-01-S5-SIDE-LOG<br>`2904709f1373d6a66a271367a882efc5815f9f1f4741861a53b6e0658140e3e7`<br>S1T-SESSION-01-S5-RECORDS-RESULT<br>`f693fe7a7bd5408d8597bc4e1652491301971fc40a5db32d444ed7e094d702cc`<br>S1T-SESSION-01-S5-DECISION-WORKBOOK<br>`0f33d295d1c46f2cc14812b4e1ece3c368c91370d9b9aaee23bf0fdd404686cf`<br>S1T-SESSION-01-S5-DF3-MANIFEST<br>`f3a0c140c200830a5f6c73250894c07ae1f0bdfe791ed24cf5f51235fc9f720d` |
| **committed_or_manifest_sha256** | `2904709f1373d6a66a271367a882efc5815f9f1f4741861a53b6e0658140e3e7` |
| **prev_entry_sha256** | `d9c94f6ac324777bc851d4f6de5dc61b778850117cca1741d43d82f84f12bd97` |
| **entry_line_sha256** | `75b379a9256ff6cbb7459886c3d318c2a96e68a7aedb6244194d69dc50919e98` |

| Entry 144 | 2026-10-06 · VERIFICATION_RECORD · manifest – |
|---|---|
| **governing_provisions** | P26-18; X-1 report 1b952205b4390624a91c97447943714dac9eb625193e22274f40b9986cd46cb7 |
| **subject** | S1T-SESSION-01-S5-SIDE-LOG<br>`2904709f1373d6a66a271367a882efc5815f9f1f4741861a53b6e0658140e3e7`<br>S1T-SESSION-01-S5-RECORDS-RESULT<br>`f693fe7a7bd5408d8597bc4e1652491301971fc40a5db32d444ed7e094d702cc`<br>S1T-SESSION-01-S5-DECISION-WORKBOOK<br>`0f33d295d1c46f2cc14812b4e1ece3c368c91370d9b9aaee23bf0fdd404686cf`<br>S1T-SESSION-01-S5-DF3-MANIFEST<br>`f3a0c140c200830a5f6c73250894c07ae1f0bdfe791ed24cf5f51235fc9f720d` |
| **committed_or_manifest_sha256** | – |
| **prev_entry_sha256** | `75b379a9256ff6cbb7459886c3d318c2a96e68a7aedb6244194d69dc50919e98` |
| **entry_line_sha256** | `797129f39bc1d47f6d3308cd1e0f807224d15c82d1244e7d37e6ecd761666ca3` |

| Entry 145 | 2026-10-06 · COMMITMENT · manifest – |
|---|---|
| **governing_provisions** | PG26-M0011 FREEZE_ACT138 COMMITMENT139 VERIFICATION140; side-log COMMITMENT143 VERIFICATION144; PI act b79983398339bbce9c2b714dd635dbfec3004176823fce097be2f2df0dc5f230 / 54406cc27d6935a9641ed70c9fa4e93f2da9ebe365f6cb3daf8902c090716905; conditional-chain act 911343c27df783beefdf9a2f7306d4137fad6718e5652110aaa93008cda8be92; Codex 712afa62 / spec 10779562; X-1 report f9969063ca39d2c407a40e88df31f545e0bc5a26653e924d496d12835391bd4d; D-1 v10 A-08 part (f) (ROLE_RECORD entry 111, verified 112); P26-14 |
| **subject** | S1T-SESSION-01-V013-TRANSCRIPTION-LOG<br>`e2beee8265a34b1f671f1ff660fcf2dfd0dc58e5c79850a966509f9c4c642d50`<br>S1T-SESSION-01-S5-SIDE-LOG<br>`242aa079bcc6cefc3ef937010312be13e7a79db3dbaea4bc822dd02487abb0da`<br>S1T-SESSION-01-S5-TRANSCRIPTION-RESULT<br>`904ff999df53d0a1ba9267b21428b63f8d04df3bbd102ab8d4f8edc6d4fc02ac`<br>S1T-SESSION-01-SCOPED-RETURN<br>`b392d7aa97550fe8cc5ef9569e4652c4f4b4d5f8d79323fdc148e83ea5d036eb`<br>S1T-SESSION-01-CARRY-CERTIFICATE<br>`b35f8b565335db03bb96c9a4f77ffaece5b7610c791472fe437eee6d1e007fad`<br>S1T-SESSION-01-TAPPA1-ORIGINAL<br>`0f33d295d1c46f2cc14812b4e1ece3c368c91370d9b9aaee23bf0fdd404686cf`<br>S1T-SESSION-01-MCA-ORIGINAL<br>`fe802b38c18a6a6b7d77fc43a1b553000ffc331d3e9a617398495f49bd05adb1`<br>S1T-SESSION-01-MCA-DECLARATION<br>`61ac21306c1281c916123c5f32194245f56f822dec631e46b477cfe0b2f29d0c`<br>S1T-SESSION-01-PI-CHAIN-ACT<br>`911343c27df783beefdf9a2f7306d4137fad6718e5652110aaa93008cda8be92`<br>S1T-SESSION-01-TRANSFER-POSTCHECK<br>`51f699739cccb0eb2555a45f875f3b648ff256abfa45ad1af4df47bcd21a9b56`<br>S1T-SESSION-01-TRANSCRIPTION-CERTIFICATE<br>`07e16c0edf2d3f425c145080b9b89e4e10f5960190ef3a9fd69690503f779e9f`<br>S1T-SESSION-01-EXECUTION-PROVENANCE<br>`c75142af276c388a055d875d97ccd603efe9c571955af26b9be43e5765427fc0` |
| **committed_or_manifest_sha256** | `e2beee8265a34b1f671f1ff660fcf2dfd0dc58e5c79850a966509f9c4c642d50` |
| **prev_entry_sha256** | `797129f39bc1d47f6d3308cd1e0f807224d15c82d1244e7d37e6ecd761666ca3` |
| **entry_line_sha256** | `714bd5890e0db56e1bfb04b6714cd1712b3b0fc11f5c85745da904647be200e7` |

| Entry 146 | 2026-10-06 · VERIFICATION_RECORD · manifest – |
|---|---|
| **governing_provisions** | P26-18; X-1 report b63a17a9c6d70115bcf160ed9f477c0ecb62e1c171aee777929aa7d911dc1055 |
| **subject** | S1T-SESSION-01-V013-TRANSCRIPTION-LOG<br>e2beee8265a34b1f671f1ff660fcf2dfd0dc58e5c79850a966509f9c4c642d50<br>S1T-SESSION-01-S5-SIDE-LOG<br>242aa079bcc6cefc3ef937010312be13e7a79db3dbaea4bc822dd02487abb0da<br>S1T-SESSION-01-S5-TRANSCRIPTION-RESULT<br>904ff999df53d0a1ba9267b21428b63f8d04df3bbd102ab8d4f8edc6d4fc02ac<br>S1T-SESSION-01-SCOPED-RETURN<br>b392d7aa97550fe8cc5ef9569e4652c4f4b4d5f8d79323fdc148e83ea5d036eb<br>S1T-SESSION-01-CARRY-CERTIFICATE<br>b35f8b565335db03bb96c9a4f77ffaece5b7610c791472fe437eee6d1e007fad<br>S1T-SESSION-01-TAPPA1-ORIGINAL<br>0f33d295d1c46f2cc14812b4e1ece3c368c91370d9b9aaee23bf0fdd404686cf<br>S1T-SESSION-01-MCA-ORIGINAL<br>fe802b38c18a6a6b7d77fc43a1b553000ffc331d3e9a617398495f49bd05adb1<br>S1T-SESSION-01-MCA-DECLARATION<br>61ac21306c1281c916123c5f32194245f56f822dec631e46b477cfe0b2f29d0c<br>S1T-SESSION-01-PI-CHAIN-ACT<br>911343c27df783beefdf9a2f7306d4137fad6718e5652110aaa93008cda8be92<br>S1T-SESSION-01-TRANSFER-POSTCHECK<br>51f699739cccb0eb2555a45f875f3b648ff256abfa45ad1af4df47bcd21a9b56<br>S1T-SESSION-01-TRANSCRIPTION-CERTIFICATE<br>07e16c0edf2d3f425c145080b9b89e4e10f5960190ef3a9fd69690503f779e9f<br>S1T-SESSION-01-EXECUTION-PROVENANCE<br>c75142af276c388a055d875d97ccd603efe9c571955af26b9be43e5765427fc0 |
| **committed_or_manifest_sha256** | – |
| **prev_entry_sha256** | 714bd5890e0db56e1bfb04b6714cd1712b3b0fc11f5c85745da904647be200e7 |
| **entry_line_sha256** | 44466869b3795b02dc29f8ae7823cdfe8e766bf4334e1b5fad904055e182dfe2 |

| Entry 147 | 2026-10-16 · COMMITMENT · manifest – |
|---|---|
| **governing_provisions** | PG-11 v0.1.3 successor clauses as activated (stage-5 transcription log); ATTO_UNICO_EMESSO_PI_v1.0.md; SHA-256 b79983398339bbce9c2b714dd635dbfec3004176823fce097be2f2df0dc5f230; Testo consolidato clausole; A, B, C, D-PROJ, G-M, M-CA, S-SET; SHA-256 54406cc27d6935a9641ed70c9fa4e93f2da9ebe365f6cb3daf8902c090716905; R-3: provenienza, 12 subject e limiti fattuali invariati; CA3 P26-25(1), P26-25(5); Autorizzazione PI AT146 e perimetro interpretativo distinti; SHA-256 ed2b1863aa21954d255594a0005b8dd6d434297476a120c20464056ba54544c0; Post-controllo indipendente Fase A Claude, condizioni Fase B; SHA-256 a4668ac38fa7cdad058fab490bcabbbc9ccba8935ce95ef6bbc96ade0dccee0e |
| **subject** | AT146-V013-LOG<br>9d95168370de42d61b68a65da6eab952012d2cbbc7ce8409ba7d3453eb45fe13<br>AT146-S5-SIDE-LOG<br>242aa079bcc6cefc3ef937010312be13e7a79db3dbaea4bc822dd02487abb0da<br>AT146-AUTHENTIC-RETURN<br>f87a84d49e206110ce5ea09e98a309fa664dd291e0776ca2be494a1141756649<br>AT146-APPROVED-MINUTES<br>eb9222de216500e8a7e87cfea7e640b326400785dd316aad5d2ae04b11de01ac<br>AT146-G6-LINEAGE<br>c3429c2a20dd29b714737cb94e5e808c34dae0ca80dfa98e0ea3413f4bec0a5c<br>AT146-I1-CERTIFICATE<br>17198057e790c4b1a17afd038e9e12ed8062def01cd416c19ed8023ca99fc385<br>AT146-PI-FACTUAL-STATEMENT<br>4353f18a260763e2cbf88ce544336d26199b0359df7d41fcea22d8bb16006eb5<br>AT146-MANAGER-LIMITS<br>f30f0b30c57c020c04e23738a9cb852430f1683af8ad78de906a4b87a464a8dc<br>AT146-TRANSCRIPTION-CERTIFICATE<br>e5c25f65b579976321146686abb4f20897e424fc1381f0c9211fd938a7354ebd<br>AT146-INDEPENDENT-TRANSCRIPTION-POSTCHECK<br>a4668ac38fa7cdad058fab490bcabbbc9ccba8935ce95ef6bbc96ade0dccee0e<br>AT146-SPECIFIC-EXECUTION-AUTHORIZATION<br>ed2b1863aa21954d255594a0005b8dd6d434297476a120c20464056ba54544c0<br>AT146-R3-EXECUTION-CERTIFICATE<br>cd2f3bf15e496d802d916d387567e14facb1c43c327c312db7c98f252d7472ec |
| **committed_or_manifest_sha256** | 9d95168370de42d61b68a65da6eab952012d2cbbc7ce8409ba7d3453eb45fe13 |
| **prev_entry_sha256** | 44466869b3795b02dc29f8ae7823cdfe8e766bf4334e1b5fad904055e182dfe2 |
| **entry_line_sha256** | f5ec16869f74857e5563c432abef15292602442c7894fbda95009c021e5c5eb1 |

| Entry 148 | 2026-10-16 · VERIFICATION_RECORD · manifest – |
|---|---|
| **governing_provisions** | PG-26 P26-23; S5L2 VERIFICATION_RECORD, check_kind commitment, della voce COMMITMENT immediatamente precedente; modello S5L2_AT147_T2_UNFILLED_GATED.py SHA-256 25b63a936bcf50feafaf0197f6ece2c41485d27f8ccbc00928beb56c55bb75b4; Post-controllo indipendente Claude del commit 147: OFTY-GR_v0.2_AT147_COMMIT_POSTCHECK_X1_CLAUDE_v1.0.md; SHA-256 14e2867c47c950ddbafec7fe1977be30c35f8bec76a301319d923f63a7a3c6f0; esito PASS riferito alla verifica meccanica del COMMITMENT 147; COMMITMENT 147; testa f5ec16869f74857e5563c432abef15292602442c7894fbda95009c021e5c5eb1; registro al prefisso 147 SHA-256 5371d14eded4d45b6f934d5fa473186975b2048663a7e7bd9cf4ae441c921ed5; log impegnato SHA-256 9d95168370de42d61b68a65da6eab952012d2cbbc7ce8409ba7d3453eb45fe13; COMMIT147_EXECUTION_CERTIFICATE.json; SHA-256 c4f5982585788502ef338ce11c1fb7ee813aeb125b1f79896b394d15d68d2ab4; Autorizzazione PI per la sequenza AT146; 01_ATTO_PI_E_PERIMETRO.md SHA-256 ed2b1863aa21954d255594a0005b8dd6d434297476a120c20464056ba54544c0; atto testuale PI e perimetro interpretativo del gestore distinti; R-3 e contratto T2 per pre_verify; 12 subject e limiti fattuali del COMMITMENT 147 conservati; nessuna estensione della verifica a disposizioni o contenuto scientifico, chiusure HRD/D2/DV o Stage 6 |
| **subject** | AT146-V013-LOG<br>`9d95168370de42d61b68a65da6eab952012d2cbbc7ce8409ba7d3453eb45fe13`<br>AT146-S5-SIDE-LOG<br>`242aa079bcc6cefc3ef937010312be13e7a79db3dbaea4bc822dd02487abb0da`<br>AT146-AUTHENTIC-RETURN<br>`f87a84d49e206110ce5ea09e98a309fa664dd291e0776ca2be494a1141756649`<br>AT146-APPROVED-MINUTES<br>`eb9222de216500e8a7e87cfea7e640b326400785dd316aad5d2ae04b11de01ac`<br>AT146-G6-LINEAGE<br>`c3429c2a20dd29b714737cb94e5e808c34dae0ca80dfa98e0ea3413f4bec0a5c`<br>AT146-I1-CERTIFICATE<br>`17198057e790c4b1a17afd038e9e12ed8062def01cd416c19ed8023ca99fc385`<br>AT146-PI-FACTUAL-STATEMENT<br>`4353f18a260763e2cbf88ce544336d26199b0359df7d41fcea22d8bb16006eb5`<br>AT146-MANAGER-LIMITS<br>`f30f0b30c57c020c04e23738a9cb852430f1683af8ad78de906a4b87a464a8dc`<br>AT146-TRANSCRIPTION-CERTIFICATE<br>`e5c25f65b579976321146686abb4f20897e424fc1381f0c9211fd938a7354ebd`<br>AT146-INDEPENDENT-TRANSCRIPTION-POSTCHECK<br>`a4668ac38fa7cdad058fab490bcabbbc9ccba8935ce95ef6bbc96ade0dccee0e`<br>AT146-SPECIFIC-EXECUTION-AUTHORIZATION<br>`ed2b1863aa21954d255594a0005b8dd6d434297476a120c20464056ba54544c0`<br>AT146-R3-EXECUTION-CERTIFICATE<br>`cd2f3bf15e496d802d916d387567e14facb1c43c327c312db7c98f252d7472ec` |
| **committed_or_manifest_sha256** | – |
| **prev_entry_sha256** | `f5ec16869f74857e5563c432abef15292602442c7894fbda95009c021e5c5eb1` |
| **entry_line_sha256** | `f3c0b8d5ed1548fc86a03a37834578ec05eb0100d62b8c47eeee493f11a78ff4` |

| Entry 149 | 2026-10-16 · COMMITMENT · manifest – |
|---|---|
| **governing_provisions** | P26-18; AM-S5-LIM; X-1 report 93af9519e9b8de370a1aa285e986cab3e1b67cac83939126614e7dc25e66f9f9 |
| **subject** | S5-LIMITATIONS-DV-CURRENT-STATE<br>`9cd43c9f01680b4d18e06df0c977aaeb269e6cc5197cbeb18a3a1d067541266e`<br>S5-LIMITATIONS-LEDGER<br>`8ba5923a8605c583e9dcb02dc46bb29c1647fa140ac84068c539e206e7349f5e`<br>S5-LIMITATIONS-CORPUS-REVIEW<br>`f7b7fc6ee15aa4624dd6df7ccf8fae16daab427f5ace005c3217063d38f8f837`<br>S5-LIMITATIONS-AUTHENTIC-FORM<br>`a6d7eb0e7bc5ec0132a317575af233b4b7e9729e57d59a01dce8149bbb3d0416`<br>S5-LIMITATIONS-AUTHENTIC-PROVENANCE<br>`8a412a29124dd6822d22a72726602c51511618a6975c81cc9516b3f3a333656f` |
| **committed_or_manifest_sha256** | `8ba5923a8605c583e9dcb02dc46bb29c1647fa140ac84068c539e206e7349f5e` |
| **prev_entry_sha256** | `f3c0b8d5ed1548fc86a03a37834578ec05eb0100d62b8c47eeee493f11a78ff4` |
| **entry_line_sha256** | `b00d14e4e181ef863d20786e7e91797476cd72b3eb3cbcb9543292e33a3fa84b` |

| Entry 150 | 2026-10-16 · VERIFICATION_RECORD · manifest – |
|---|---|
| **governing_provisions** | P26-18; AM-S5-LIM; X-1 report 93af9519e9b8de370a1aa285e986cab3e1b67cac83939126614e7dc25e66f9f9 |
| **subject** | S5-LIMITATIONS-DV-CURRENT-STATE<br>`9cd43c9f01680b4d18e06df0c977aaeb269e6cc5197cbeb18a3a1d067541266e`<br>S5-LIMITATIONS-LEDGER<br>`8ba5923a8605c583e9dcb02dc46bb29c1647fa140ac84068c539e206e7349f5e`<br>S5-LIMITATIONS-CORPUS-REVIEW<br>`f7b7fc6ee15aa4624dd6df7ccf8fae16daab427f5ace005c3217063d38f8f837`<br>S5-LIMITATIONS-AUTHENTIC-FORM<br>`a6d7eb0e7bc5ec0132a317575af233b4b7e9729e57d59a01dce8149bbb3d0416`<br>S5-LIMITATIONS-AUTHENTIC-PROVENANCE<br>`8a412a29124dd6822d22a72726602c51511618a6975c81cc9516b3f3a333656f` |
| **committed_or_manifest_sha256** | – |
| **prev_entry_sha256** | `b00d14e4e181ef863d20786e7e91797476cd72b3eb3cbcb9543292e33a3fa84b` |
| **entry_line_sha256** | `16cb1aa4d7a13cd859f9929686d552c642098ef3b403d0af6d65eb5601c9849e` |

| Entry 151 | 2026-10-16 · COMMITMENT · manifest – |
|---|---|
| **governing_provisions** | Carta section 1 as amended for Stage 5 by AM-S5-LIM section 4; AM-S5-LIM sections 4-6; sha256 09055ce2fde32ec2fd31cac99bd26b0e574ff65fbf7e7d2ef8dfedf067fc8d25; PI S2 adoption and technical mandate sha256 dcafbab8e138913e1d0b2eba434e5506025de39508513857923943f30bbccf7d; PG-26 P26-23; X-1 report b9f9c755b187a8dda16aca6b56bb7a8e632a53bb87bcfedee1105136aabc1e3a |
| **subject** | S5-ACT-S2<br>`8c64e1c3e111f675e82081351ab389f3e0c007cbd4d244f7fab9339e14965185`<br>S5-ACT-S2-PI-DECLARATION<br>`dcafbab8e138913e1d0b2eba434e5506025de39508513857923943f30bbccf7d`<br>S5-LIMITATIONS-CORPUS-REVIEW<br>`f7b7fc6ee15aa4624dd6df7ccf8fae16daab427f5ace005c3217063d38f8f837`<br>S5-LIMITATIONS-LEDGER<br>`8ba5923a8605c583e9dcb02dc46bb29c1647fa140ac84068c539e206e7349f5e` |
| **committed_or_manifest_sha256** | `8c64e1c3e111f675e82081351ab389f3e0c007cbd4d244f7fab9339e14965185` |
| **prev_entry_sha256** | `16cb1aa4d7a13cd859f9929686d552c642098ef3b403d0af6d65eb5601c9849e` |
| **entry_line_sha256** | `39229412f20d4d341713208fe448314169647bcc8b3c2fcd9e85a2a888da7645` |

| Entry 152 | 2026-10-16 · VERIFICATION_RECORD · manifest – |
|---|---|
| **governing_provisions** | Carta section 1 as amended for Stage 5 by AM-S5-LIM section 4; AM-S5-LIM sections 4-6; sha256 09055ce2fde32ec2fd31cac99bd26b0e574ff65fbf7e7d2ef8dfedf067fc8d25; PI S2 adoption and technical mandate sha256 dcafbab8e138913e1d0b2eba434e5506025de39508513857923943f30bbccf7d; PG-26 P26-23; X-1 report b9f9c755b187a8dda16aca6b56bb7a8e632a53bb87bcfedee1105136aabc1e3a |
| **subject** | S5-ACT-S2<br>`8c64e1c3e111f675e82081351ab389f3e0c007cbd4d244f7fab9339e14965185`<br>S5-ACT-S2-PI-DECLARATION<br>`dcafbab8e138913e1d0b2eba434e5506025de39508513857923943f30bbccf7d`<br>S5-LIMITATIONS-CORPUS-REVIEW<br>`f7b7fc6ee15aa4624dd6df7ccf8fae16daab427f5ace005c3217063d38f8f837`<br>S5-LIMITATIONS-LEDGER<br>`8ba5923a8605c583e9dcb02dc46bb29c1647fa140ac84068c539e206e7349f5e` |
| **committed_or_manifest_sha256** | – |
| **prev_entry_sha256** | `39229412f20d4d341713208fe448314169647bcc8b3c2fcd9e85a2a888da7645` |
| **entry_line_sha256** | `83ad7b595b557b16c1aeaf2c9c5ed2a28c3cee2eca38603ec9fa6effb322fe66` |

| Entry 153 | 2026-10-17 · COMMITMENT · manifest – |
|---|---|
| **governing_provisions** | PG-11 v0.1.3 change record section 1 (P26-10(ii)): R6 invalidated for further use, replaced by R6'; sha256 c1b77a02f9f8f670ae2451caa49458b55b1003031090179657415ebf6ff2f80a; PG-15 section 12.1 R6; PI act b79983398339bbce9c2b714dd635dbfec3004176823fce097be2f2df0dc5f230 (Codex allocation: R6 successor build with comparison and final account); M0011 02ffc74456ad1a3dad834f44fa9ef2dfa6a3347b2a466cae1baee9c069d3814a freeze138 commitment139 verification140; s5_projection_records.py r6-build 6b3ed58f9a1d53cd6d3d9a1b1fdc79f16665edf1a72b6be39901a3b8ade9c4a0; PG-26 P26-23; X-1 report 54fa8670771c9f2982bf1330a76eaaec3c32bb2fb0a6310f2bd33d6372dc8ea3 |
| **subject** | OFTYGR-R6P-BUILD-MANIFEST<br>`3d20365ab7a6ef4c5014db8295396453875a7ba8efb5a7f5dcf35d60f97c596d`<br>OFTYGR-R6P-CANONICAL-RECORD<br>`1f9b67998212fecebbcbb5aefd24ea4d2baa335029749ee8849f8bb1860aa72b`<br>OFTYGR-R6P-ROW-DIFF<br>`271a766ebc41a7003cc67eb0a0f6ac42b112f8a46b7bea4478d8b7ae82d2dd10`<br>OFTYGR-R6P-REGRESSION<br>`435141161cef3e5c969e59106eec6d0120660e994a0f15212d03ddca55023903` |
| **committed_or_manifest_sha256** | `3d20365ab7a6ef4c5014db8295396453875a7ba8efb5a7f5dcf35d60f97c596d` |
| **prev_entry_sha256** | `83ad7b595b557b16c1aeaf2c9c5ed2a28c3cee2eca38603ec9fa6effb322fe66` |
| **entry_line_sha256** | `95f896c1c1a73e62212132287a4ae820a32dea96083e31cad2eeab8e572e802f` |

| Entry 154 | 2026-10-17 · VERIFICATION_RECORD · manifest – |
|---|---|
| **governing_provisions** | PG-11 v0.1.3 change record section 1 (P26-10(ii)): R6 invalidated for further use, replaced by R6'; sha256 c1b77a02f9f8f670ae2451caa49458b55b1003031090179657415ebf6ff2f80a; PG-15 section 12.1 R6; PI act b79983398339bbce9c2b714dd635dbfec3004176823fce097be2f2df0dc5f230 (Codex allocation: R6 successor build with comparison and final account); M0011 02ffc74456ad1a3dad834f44fa9ef2dfa6a3347b2a466cae1baee9c069d3814a freeze138 commitment139 verification140; s5_projection_records.py r6-build 6b3ed58f9a1d53cd6d3d9a1b1fdc79f16665edf1a72b6be39901a3b8ade9c4a0; PG-26 P26-23; X-1 report 54fa8670771c9f2982bf1330a76eaaec3c32bb2fb0a6310f2bd33d6372dc8ea3 |
| **subject** | OFTYGR-R6P-BUILD-MANIFEST<br>`3d20365ab7a6ef4c5014db8295396453875a7ba8efb5a7f5dcf35d60f97c596d`<br>OFTYGR-R6P-CANONICAL-RECORD<br>`1f9b67998212fecebbcbb5aefd24ea4d2baa335029749ee8849f8bb1860aa72b`<br>OFTYGR-R6P-ROW-DIFF<br>`271a766ebc41a7003cc67eb0a0f6ac42b112f8a46b7bea4478d8b7ae82d2dd10`<br>OFTYGR-R6P-REGRESSION<br>`435141161cef3e5c969e59106eec6d0120660e994a0f15212d03ddca55023903` |
| **committed_or_manifest_sha256** | – |
| **prev_entry_sha256** | `95f896c1c1a73e62212132287a4ae820a32dea96083e31cad2eeab8e572e802f` |
| **entry_line_sha256** | `0208c9324a154e825681ebd33f3cb2f80ec53c46c5bb2443f0ba2d11722d56e0` |

| Entry 155 | 2026-10-17 · COMMITMENT · manifest – |
|---|---|
| **governing_provisions** | Carta section 1 as amended for Stage 5 by AM-S5-LIM section 4; AM-S5-LIM sections 4-6; sha256 09055ce2fde32ec2fd31cac99bd26b0e574ff65fbf7e7d2ef8dfedf067fc8d25; PI act b79983398339bbce9c2b714dd635dbfec3004176823fce097be2f2df0dc5f230 (S3 remains a separate act); S2 act 8c64e1c3e111f675e82081351ab389f3e0c007cbd4d244f7fab9339e14965185 (entries 151/152); R6' build manifest 3d20365ab7a6ef4c5014db8295396453875a7ba8efb5a7f5dcf35d60f97c596d (entries 153/154); independent gatekeeper conformity check 31082586a68ae70bf9fad95c0c220f5e36c4830f9dc35a33d1062ac412d1b15a; PG-26 P26-23; X-1 report 54fa8670771c9f2982bf1330a76eaaec3c32bb2fb0a6310f2bd33d6372dc8ea3 |
| **subject** | S5-ACT-COMPLETION<br>14e2b440fe7fa0f309e0f0589b65a18f409c7b4226dce1f7f2f9ef3b4af39183<br>S5-ACT-COMPLETION-PI-ADOPTION<br>4e1d6891e209311a670fe103d44000739651dec98ed2617a847f87464a14324b<br>OFTYGR-R6P-BUILD-MANIFEST<br>3d20365ab7a6ef4c5014db8295396453875a7ba8efb5a7f5dcf35d60f97c596d<br>S5-ACT-S2<br>8c64e1c3e111f675e82081351ab389f3e0c007cbd4d244f7fab9339e14965185 |
| **committed_or_manifest_sha256** | 14e2b440fe7fa0f309e0f0589b65a18f409c7b4226dce1f7f2f9ef3b4af39183 |
| **prev_entry_sha256** | 0208c9324a154e825681ebd33f3cb2f80ec53c46c5bb2443f0ba2d11722d56e0 |
| **entry_line_sha256** | 1cab5dd73589e71e32f4861f65db790aa40d6c8e16b185c45ba21b19842e2de4 |

| Entry 156 | 2026-10-17 · VERIFICATION_RECORD · manifest – |
|---|---|
| **governing_provisions** | Carta section 1 as amended for Stage 5 by AM-S5-LIM section 4; AM-S5-LIM sections 4-6; sha256 09055ce2fde32ec2fd31cac99bd26b0e574ff65fbf7e7d2ef8dfedf067fc8d25; PI act b79983398339bbce9c2b714dd635dbfec3004176823fce097be2f2df0dc5f230 (S3 remains a separate act); S2 act 8c64e1c3e111f675e82081351ab389f3e0c007cbd4d244f7fab9339e14965185 (entries 151/152); R6' build manifest 3d20365ab7a6ef4c5014db8295396453875a7ba8efb5a7f5dcf35d60f97c596d (entries 153/154); independent gatekeeper conformity check 31082586a68ae70bf9fad95c0c220f5e36c4830f9dc35a33d1062ac412d1b15a; PG-26 P26-23; X-1 report 54fa8670771c9f2982bf1330a76eaaec3c32bb2fb0a6310f2bd33d6372dc8ea3 |
| **subject** | S5-ACT-COMPLETION<br>14e2b440fe7fa0f309e0f0589b65a18f409c7b4226dce1f7f2f9ef3b4af39183<br>S5-ACT-COMPLETION-PI-ADOPTION<br>4e1d6891e209311a670fe103d44000739651dec98ed2617a847f87464a14324b<br>OFTYGR-R6P-BUILD-MANIFEST<br>3d20365ab7a6ef4c5014db8295396453875a7ba8efb5a7f5dcf35d60f97c596d<br>S5-ACT-S2<br>8c64e1c3e111f675e82081351ab389f3e0c007cbd4d244f7fab9339e14965185 |
| **committed_or_manifest_sha256** | – |
| **prev_entry_sha256** | 1cab5dd73589e71e32f4861f65db790aa40d6c8e16b185c45ba21b19842e2de4 |
| **entry_line_sha256** | 6f768db585689bb25847e74f1dfa335458255703ce969efd7e19bbb83b7de348 |

| Entry 157 | 2026-10-17 · COMMITMENT · manifest – |
|---|---|
| **governing_provisions** | Carta section 1 (specific act opening the next stage); AM-S5-LIM section 4 (S3 authorizes only the admitted corpus after verified closure); sha256 09055ce2fde32ec2fd31cac99bd26b0e574ff65fbf7e7d2ef8dfedf067fc8d25; PI act b79983398339bbce9c2b714dd635dbfec3004176823fce097be2f2df0dc5f230; Stage 5 completion declaration 14e2b440fe7fa0f309e0f0589b65a18f409c7b4226dce1f7f2f9ef3b4af39183 (entries 155/156); Suspensive conditions C-1 to C-6: PG-26 P26-14, P26-18, P26-02(iv), P26-25(4); PG-29 P29-14 as amended by CA3-20 and CA3-21; PG-32 P32-11, P32-14; PG-08 sec. 16; independent gatekeeper conformity check 32a09558d8105542ff2fd9d4a83738b70e3037e0cf5c72fde8083f5d88a26934; PG-26 P26-23; X-1 report 73064b1b065ec910358dd192c99b8f4e5a9ed3d8f8181b0df27ad474f356d2c0 |
| **subject** | S6-ACT-S3<br>5baf5992c1ee21ce5c58907c5fa7d68999f6702b803bec8629088f653dadce1f<br>S6-ACT-S3-PI-ADOPTION<br>a98c56e9855d5c9cdc4b3e1704a9b45dd0a7226a784234c2c90286bbc4166916<br>S5-ACT-COMPLETION<br>14e2b440fe7fa0f309e0f0589b65a18f409c7b4226dce1f7f2f9ef3b4af39183<br>S5-LIMITATIONS-CORPUS-REVIEW<br>f7b7fc6ee15aa4624dd6df7ccf8fae16daab427f5ace005c3217063d38f8f837<br>OFTYGR-R6P-CANONICAL-RECORD<br>1f9b67998212fecebbcbb5aefd24ea4d2baa335029749ee8849f8bb1860aa72b |
| **committed_or_manifest_sha256** | 5baf5992c1ee21ce5c58907c5fa7d68999f6702b803bec8629088f653dadce1f |
| **prev_entry_sha256** | 6f768db585689bb25847e74f1dfa335458255703ce969efd7e19bbb83b7de348 |
| **entry_line_sha256** | 2043ae9e83debf605df797d4008d6f2129a91285116437d4561d05c4fefbedb7 |

| Entry 158 | 2026-10-17 · VERIFICATION_RECORD · manifest – |
|---|---|
| **governing_provisions** | Carta section 1 (specific act opening the next stage); AM-S5-LIM section 4 (S3 authorizes only the admitted corpus after verified closure); sha256 09055ce2fde32ec2fd31cac99bd26b0e574ff65fbf7e7d2ef8dfedf067fc8d25; PI act b79983398339bbce9c2b714dd635dbfec3004176823fce097be2f2df0dc5f230; Stage 5 completion declaration 14e2b440fe7fa0f309e0f0589b65a18f409c7b4226dce1f7f2f9ef3b4af39183 (entries 155/156); Suspensive conditions C-1 to C-6: PG-26 P26-14, P26-18, P26-02(iv), P26-25(4); PG-29 P29-14 as amended by CA3-20 and CA3-21; PG-32 P32-11, P32-14; PG-08 sec. 16; independent gatekeeper conformity check 32a09558d8105542ff2fd9d4a83738b70e3037e0cf5c72fde8083f5d88a26934; PG-26 P26-23; X-1 report 73064b1b065ec910358dd192c99b8f4e5a9ed3d8f8181b0df27ad474f356d2c0 |
| **subject** | S6-ACT-S3<br>5baf5992c1ee21ce5c58907c5fa7d68999f6702b803bec8629088f653dadce1f<br>S6-ACT-S3-PI-ADOPTION<br>a98c56e9855d5c9cdc4b3e1704a9b45dd0a7226a784234c2c90286bbc4166916<br>S5-ACT-COMPLETION<br>14e2b440fe7fa0f309e0f0589b65a18f409c7b4226dce1f7f2f9ef3b4af39183<br>S5-LIMITATIONS-CORPUS-REVIEW<br>f7b7fc6ee15aa4624dd6df7ccf8fae16daab427f5ace005c3217063d38f8f837<br>OFTYGR-R6P-CANONICAL-RECORD<br>1f9b67998212fecebbcbb5aefd24ea4d2baa335029749ee8849f8bb1860aa72b |
| **committed_or_manifest_sha256** | – |
| **prev_entry_sha256** | 2043ae9e83debf605df797d4008d6f2129a91285116437d4561d05c4fefbedb7 |
| **entry_line_sha256** | 372778e4230f77f56e9112fb4672318a74a69238f6521640b8c1035d3ac9907c |

| Entry 159 | 2026-10-17 · ROLE_RECORD · manifest – |
|---|---|
| **governing_provisions** | P32-14; P32-13; P32-02; P32-11; PG-14 E-03; P26-14; P26-18 |
| **subject** | PG32-D1<br>b9a6ee36a3e8e863998ae574198747e669fec03a80dc71ff2f3e34413028c3ae<br>GATEKEEPER-REPORT-D1v11<br>e84b3589e6b7b6e7cb7b56260aba0205bfe799378c160c285b1c2bce0f7cc7fb<br>PG32-D1-v11-APPROVAL-AND-FREEZE-ACT<br>246700d96be2893566d2359cc0b775ef017fa13bab3557d038da7b4b1f31bf00 |
| **committed_or_manifest_sha256** | – |
| **prev_entry_sha256** | 372778e4230f77f56e9112fb4672318a74a69238f6521640b8c1035d3ac9907c |
| **entry_line_sha256** | ae6d3e1c3f4d8f2cdf31984b2f28598b80452b060c92ac8dbade86c5d3976e96 |

| Entry 160 | 2026-10-17 · VERIFICATION_RECORD · manifest – |
|---|---|
| **governing_provisions** | P26-18; P32-14; P32-13 |
| **subject** | PG32-D1<br>b9a6ee36a3e8e863998ae574198747e669fec03a80dc71ff2f3e34413028c3ae |
| **committed_or_manifest_sha256** | – |
| **prev_entry_sha256** | ae6d3e1c3f4d8f2cdf31984b2f28598b80452b060c92ac8dbade86c5d3976e96 |
| **entry_line_sha256** | cae910857fe135809a1f1ea6fbd3769fad614afc2f8768ba91537c16865a7a26 |

| Entry 161 | 2026-10-17 · FREEZE_ACT · manifest M0012 |
|---|---|
| **governing_provisions** | P26-18; P26-14; P26-02(vi); Act S3 v1.3 C-1, C-6 |
| **subject** | PG26-M0012<br>326e741fdf1663e14f2933815930ece864e00d8d4b277558614bb385e901411c |
| **committed_or_manifest_sha256** | 326e741fdf1663e14f2933815930ece864e00d8d4b277558614bb385e901411c |
| **prev_entry_sha256** | cae910857fe135809a1f1ea6fbd3769fad614afc2f8768ba91537c16865a7a26 |
| **entry_line_sha256** | b213c256b4fcde8826aa4799151ed473abe487032930c02d22eb4c9ecc8a7135 |

| Entry 162 | 2026-10-17 · COMMITMENT · manifest – |
|---|---|
| **governing_provisions** | P26-02(iii) as amended by CA2-10/CA3-10; P26-14 |
| **subject** | PG26-M0012<br>326e741fdf1663e14f2933815930ece864e00d8d4b277558614bb385e901411c |
| **committed_or_manifest_sha256** | 326e741fdf1663e14f2933815930ece864e00d8d4b277558614bb385e901411c |
| **prev_entry_sha256** | b213c256b4fcde8826aa4799151ed473abe487032930c02d22eb4c9ecc8a7135 |
| **entry_line_sha256** | 20a91c901ba7864ad03685e4a9c4446cdaf4b77d71e1e3723bbe44aa0e2f695b |

| Entry 163 | 2026-10-17 · VERIFICATION_RECORD · manifest – |
|---|---|
| **governing_provisions** | P26-18; P26-14; P26-12 |
| **subject** | PG26-M0012<br>326e741fdf1663e14f2933815930ece864e00d8d4b277558614bb385e901411c |
| **committed_or_manifest_sha256** | – |
| **prev_entry_sha256** | 20a91c901ba7864ad03685e4a9c4446cdaf4b77d71e1e3723bbe44aa0e2f695b |
| **entry_line_sha256** | bbc9f0eee724eb190442c1a20ca1923145c044dd6ecae6cd4f85ecfd3dee68f8 |

| Entry 164 | 2026-10-18 · DECISIVE_STEP_MANIFEST · manifest – |
|---|---|
| **governing_provisions** | PG-29 P29-14(ii) as replaced by CA3-21; CA3-01; CA3-02; PG-26 P26-25(4); IP-01 sec. 7; IP-02 sec. 6 |
| **subject** | RMD-01<br>934455640a981591d675a4d8e8787223f919b46d14495e16dc56e203416a839c |
| **committed_or_manifest_sha256** | – |
| **prev_entry_sha256** | bbc9f0eee724eb190442c1a20ca1923145c044dd6ecae6cd4f85ecfd3dee68f8 |
| **entry_line_sha256** | bc0dfdf178e2ed07344db5d1166c4a4f071b2b74315cf0c705afeda4961f2e98 |

| **Entry 165** | 2026-10-18 · VERIFICATION_RECORD · manifest – |
|---|---|
| **governing_provisions** | PG-26 P26-18 as amended by CA3-11; IP-04 V-10a; CA3-13(4); CA2-32 |
| **subject** | RMD-01<br>`934455640a981591d675a4d8e8787223f919b46d14495e16dc56e203416a839c` |
| **committed_or_manifest_sha256** | – |
| **prev_entry_sha256** | `bc0dfdf178e2ed07344db5d1166c4a4f071b2b74315cf0c705afeda4961f2e98` |
| **entry_line_sha256** | `5ab894ae6965e4c7fe3cbdeb8e27a176a30df29dad5848eea6c1ea0e113905db` |

| **Entry 166** | 2026-10-18 · TSA_TOKEN · manifest – |
|---|---|
| **governing_provisions** | PG-26 P26-25(4), (6); P29-21; CA3-01; CA3-04; IP-06 |
| **subject** | RMD-01<br>`934455640a981591d675a4d8e8787223f919b46d14495e16dc56e203416a839c` |
| **committed_or_manifest_sha256** | – |
| **prev_entry_sha256** | `5ab894ae6965e4c7fe3cbdeb8e27a176a30df29dad5848eea6c1ea0e113905db` |
| **entry_line_sha256** | `e3b6d89dc131b53e3298724b1fb9bfd4e52ca970bf4707feff82104017b3cc0a` |

| **Entry 167** | 2026-10-18 · VERIFICATION_RECORD · manifest – |
|---|---|
| **governing_provisions** | PG-26 P26-18 as amended by CA3-11; IP-04 V-10b; CA3-13(4); CA2-32 |
| **subject** | RMD-01<br>`934455640a981591d675a4d8e8787223f919b46d14495e16dc56e203416a839c` |
| **committed_or_manifest_sha256** | – |
| **prev_entry_sha256** | `e3b6d89dc131b53e3298724b1fb9bfd4e52ca970bf4707feff82104017b3cc0a` |
| **entry_line_sha256** | `11ba661ed187e989e7935ac31707b5029db789b1634896a19b168ae11e9979e9` |

| **Entry 168** | 2026-10-19 · ROLE_RECORD · manifest – |
|---|---|
| **governing_provisions** | P32-14; P32-13; P32-02; P32-11; PG-14 E-03; P26-14; P26-18 |
| **subject** | PG32-D1<br>`483a7585ef93eb05a326d3bd54bd50d1f2bdbce7bbc96f7749af28d31bc1eae8`<br>GATEKEEPER-REPORT-D1v12<br>`7cd04034a349c002428e726636b90cf933f0ee374c39baa9be0fc11e452ee6fb`<br>PG32-D1-v12-APPROVAL-AND-FREEZE-ACT<br>`eb7b34456638737e2f875130059fd492b7418c6774d666a0a3474e09ef8e2ddb` |
| **committed_or_manifest_sha256** | – |
| **prev_entry_sha256** | `11ba661ed187e989e7935ac31707b5029db789b1634896a19b168ae11e9979e9` |
| **entry_line_sha256** | `acf47caaf86d0e2acc961cd1d8cbb5c96867cb50d350acdf21eab27fdbf79974` |

| **Entry 169** | 2026-10-19 · VERIFICATION_RECORD · manifest – |
|---|---|
| **governing_provisions** | P26-18; P32-14; P32-13 |
| **subject** | PG32-D1<br>`483a7585ef93eb05a326d3bd54bd50d1f2bdbce7bbc96f7749af28d31bc1eae8` |
| **committed_or_manifest_sha256** | – |
| **prev_entry_sha256** | `acf47caaf86d0e2acc961cd1d8cbb5c96867cb50d350acdf21eab27fdbf79974` |
| **entry_line_sha256** | `2127d359d9f9aca7b3d665a97c4ab01a31e2b72c567a3d3a9664d8d90ddf791a` |

| **Entry 170** | 2026-10-19 · FREEZE_ACT · manifest M0013 |
|---|---|
| **governing_provisions** | P26-18; P26-14; P26-02(vi); P26-10(ii) |
| **subject** | PG26-M0013<br>`7c179012f88df1c6636a5165df6b70aa9107e0c5459195e6e58c53eba3d38359` |
| **committed_or_manifest_sha256** | `7c179012f88df1c6636a5165df6b70aa9107e0c5459195e6e58c53eba3d38359` |
| **prev_entry_sha256** | `2127d359d9f9aca7b3d665a97c4ab01a31e2b72c567a3d3a9664d8d90ddf791a` |
| **entry_line_sha256** | `01bd308b1633e6541d7c2a50fa1fe453d90cb2d0e1a7f9f8bccfd53f7cb61120` |

| **Entry 171** | 2026-10-19 · COMMITMENT · manifest – |
|---|---|
| **governing_provisions** | P26-02(iii) as amended by CA2-10/CA3-10; P26-14; P26-10(ii) |
| **subject** | PG26-M0013<br>`7c179012f88df1c6636a5165df6b70aa9107e0c5459195e6e58c53eba3d38359` |
| **committed_or_manifest_sha256** | `7c179012f88df1c6636a5165df6b70aa9107e0c5459195e6e58c53eba3d38359` |
| **prev_entry_sha256** | `01bd308b1633e6541d7c2a50fa1fe453d90cb2d0e1a7f9f8bccfd53f7cb61120` |
| **entry_line_sha256** | `88eea17260fddd57b85f55cc37172b2f18c2e7f0bac2b2dfe636c0f338b46ee7` |

| **Entry 172** | 2026-10-19 · VERIFICATION_RECORD · manifest – |
|---|---|
| **governing_provisions** | P26-18; P26-14; P26-12 |
| **subject** | PG26-M0013<br>`7c179012f88df1c6636a5165df6b70aa9107e0c5459195e6e58c53eba3d38359` |
| **committed_or_manifest_sha256** | – |
| **prev_entry_sha256** | `88eea17260fddd57b85f55cc37172b2f18c2e7f0bac2b2dfe636c0f338b46ee7` |
| **entry_line_sha256** | `292eb0ae6e2fd4653d51d56641bfd3bf985ab3682f7738e3be9f037cd4b35079` |

| **Entry 173** | 2026-10-19 · COMMITMENT · manifest – |
|---|---|
| **governing_provisions** | PG-10 §12 (M-03A); PG-10 sec. 10.1, T6, T7, T31; P26-02(vi); CA2-32 |
| **subject** | RR-OHT-01<br>`10b35b88bff93b3149c821daf2f84e02b82d00952d37f110e97dbb04da867a82` |
| **committed_or_manifest_sha256** | `10b35b88bff93b3149c821daf2f84e02b82d00952d37f110e97dbb04da867a82` |
| **prev_entry_sha256** | `292eb0ae6e2fd4653d51d56641bfd3bf985ab3682f7738e3be9f037cd4b35079` |
| **entry_line_sha256** | `02b07ab6b6cddae95b621da735c7ea2698cb204751eba2e879a726a63a31689e` |

| **Entry 174** | 2026-10-19 · VERIFICATION_RECORD · manifest – |
|---|---|
| **governing_provisions** | CA2-32; P26-18; PG-10 §12 (M-03A) |
| **committed_or_manifest_sha256** | – |
| **prev_entry_sha256** | 02b07ab6b6cddae95b621da735c7ea2698cb204751eba2e879a726a63a31689e |
| **entry_line_sha256** | 8f7822c6fe64c82950a120f4038c6017bef889277bcec05f87831c2c0d5a2f20 |

| **Entry 175** | 2026-10-19 · COMMITMENT · manifest – |
|---|---|
| **governing_provisions** | PG-10 §12 (M-03A); PG-10 sec. 10.1, T6, T7, T31; P26-02(vi); CA2-32 |
| **subject** | FIS-TF-OHT-01<br>34e8821316daadef10f32a0066e7b71d217a498a811fdef387c19e0f1edbbaac |
| **committed_or_manifest_sha256** | 34e8821316daadef10f32a0066e7b71d217a498a811fdef387c19e0f1edbbaac |
| **prev_entry_sha256** | 8f7822c6fe64c82950a120f4038c6017bef889277bcec05f87831c2c0d5a2f20 |
| **entry_line_sha256** | 73e71a995b7e558d4de5b7c066d74e8186f4015231baa37a569f4f73671b7193 |

| **Entry 176** | 2026-10-19 · VERIFICATION_RECORD · manifest – |
|---|---|
| **governing_provisions** | CA2-32; P26-18; PG-10 §12 (M-03A) |
| **committed_or_manifest_sha256** | – |
| **prev_entry_sha256** | 73e71a995b7e558d4de5b7c066d74e8186f4015231baa37a569f4f73671b7193 |
| **entry_line_sha256** | 390b1857534eb8bbeaf28b11916466b138c91532064b93ebfcaded618d185ae8 |

| **Entry 177** | 2026-10-20 · COMMITMENT · manifest – |
|---|---|
| **governing_provisions** | PG-10 §12 (M-03A); PG-10 sec. 10.1, T6, T7, T31; P26-02(vi); CA2-32 |
| **subject** | RR-OHT-02<br>d19e0bec25ac19e22140384e750791105a8d90c8ec547ffc4279278bd80e6aab |
| **committed_or_manifest_sha256** | d19e0bec25ac19e22140384e750791105a8d90c8ec547ffc4279278bd80e6aab |
| **prev_entry_sha256** | 390b1857534eb8bbeaf28b11916466b138c91532064b93ebfcaded618d185ae8 |
| **entry_line_sha256** | e3e461349fa73a15abc3ad2aacb6c3a3bd04b9fcb154723fd343b3215834ed17 |

| **Entry 178** | 2026-10-20 · VERIFICATION_RECORD · manifest – |
|---|---|
| **governing_provisions** | CA2-32; P26-18; PG-10 §12 (M-03A) |
| **committed_or_manifest_sha256** | – |
| **prev_entry_sha256** | e3e461349fa73a15abc3ad2aacb6c3a3bd04b9fcb154723fd343b3215834ed17 |
| **entry_line_sha256** | 6b46c308987eb443d71400c85a0c508e91318346897408e829aaf3afd0e33fa7 |

| **Entry 179** | 2026-10-20 · COMMITMENT · manifest – |
|---|---|
| **governing_provisions** | PG-10 §12 (M-03A); PG-10 sec. 10.1, T6, T7, T31; P26-02(vi); CA2-32 |
| **subject** | RR-OHT-03<br>0d06433623ab4ef20bcb3d2d8eff32d059f9e3d3ad74e9e79bf94e3537da4e0f |
| **committed_or_manifest_sha256** | 0d06433623ab4ef20bcb3d2d8eff32d059f9e3d3ad74e9e79bf94e3537da4e0f |
| **prev_entry_sha256** | 6b46c308987eb443d71400c85a0c508e91318346897408e829aaf3afd0e33fa7 |
| **entry_line_sha256** | e6a626eddf8478e1929e4097ef73978d5c2d2b179d12ff82512396201fa47997 |

| **Entry 180** | 2026-10-20 · VERIFICATION_RECORD · manifest – |
|---|---|
| **governing_provisions** | CA2-32; P26-18; PG-10 §12 (M-03A) |
| **committed_or_manifest_sha256** | – |
| **prev_entry_sha256** | e6a626eddf8478e1929e4097ef73978d5c2d2b179d12ff82512396201fa47997 |
| **entry_line_sha256** | ab159597a20f3cf362d0cee9eaff06edbbcc37e7b2f60137ea3da5ec16fcc3e0 |

| **Entry 181** | 2026-10-20 · COMMITMENT · manifest – |
|---|---|
| **governing_provisions** | PG-10 §12 (M-03A); PG-10 sec. 10.1, T6, T7, T31; P26-02(vi); CA2-32 |
| **subject** | FIS-TF-OHT-02<br>2fde2ae62122c7a36678879792d7063eb57d5f946617523b1dd76db7c09a861a |
| **committed_or_manifest_sha256** | 2fde2ae62122c7a36678879792d7063eb57d5f946617523b1dd76db7c09a861a |
| **prev_entry_sha256** | ab159597a20f3cf362d0cee9eaff06edbbcc37e7b2f60137ea3da5ec16fcc3e0 |
| **entry_line_sha256** | 884f34153fb251b99531c34ccc63dffca965b68b1ce021758828095afbe46b71 |

| **Entry 182** | 2026-10-20 · VERIFICATION_RECORD · manifest – |
|---|---|
| **governing_provisions** | CA2-32; P26-18; PG-10 §12 (M-03A) |
| **committed_or_manifest_sha256** | – |
| **prev_entry_sha256** | 884f34153fb251b99531c34ccc63dffca965b68b1ce021758828095afbe46b71 |
| **entry_line_sha256** | 99376d308a7618465f68ff5b2c86048b10b03c38c7ae3d351ba6d61e6cfb9c88 |

| **Entry 183** | 2026-10-20 · COMMITMENT · manifest – |
|---|---|
| **governing_provisions** | PG-10 §12 (M-03A); PG-10 sec. 10.1, T6, T7, T31; P26-02(vi); CA2-32 |
| **subject** | FIS-TF-OHT-03<br>c993aa5607a62ca73648d4e5eff822badfdfce46388ac1bd0c7eb68b4825acb1 |
| **committed_or_manifest_sha256** | c993aa5607a62ca73648d4e5eff822badfdfce46388ac1bd0c7eb68b4825acb1 |
| **prev_entry_sha256** | 99376d308a7618465f68ff5b2c86048b10b03c38c7ae3d351ba6d61e6cfb9c88 |
| **entry_line_sha256** | f15c6e26f5f5556db92e0f23d7dd5d12740422f56a0807149249ca59d87e1097 |

| **Entry 184** | 2026-10-20 · VERIFICATION_RECORD · manifest – |
|---|---|
| **governing_provisions** | CA2-32; P26-18; PG-10 §12 (M-03A) |
| **committed_or_manifest_sha256** | – |
| **prev_entry_sha256** | f15c6e26f5f5556db92e0f23d7dd5d12740422f56a0807149249ca59d87e1097 |
| **entry_line_sha256** | d419e9057290bfacf6350fa989f5464ddb7d10e2a8c6416675878d102349b1c6 |

| Entry 185 | 2026-10-22 · COMMITMENT · manifest – |
|---|---|
| **governing_provisions** | PG-10 §12 (M-03A); PG-10 sec. 10.1, T6, T7, T31; P26-02(vi); CA2-32 |
| **subject** | RR-EGS-01<br>929f55a5633f86179c8dc751ba8f8ee86948db1bd3a7e19b531e781683cd28e1 |
| **committed_or_manifest_sha256** | 929f55a5633f86179c8dc751ba8f8ee86948db1bd3a7e19b531e781683cd28e1 |
| **prev_entry_sha256** | d419e9057290bfacf6350fa989f5464ddb7d10e2a8c6416675878d102349b1c6 |
| **entry_line_sha256** | 2333cd8b64286c1e074ca5121bca005f522192824df980a92f474099579bc697 |

| Entry 186 | 2026-10-22 · COMMITMENT · manifest – |
|---|---|
| **governing_provisions** | PG-10 §12 (M-03A); PG-10 sec. 10.1, T6, T7, T31; P26-02(vi); CA2-32 |
| **subject** | RR-EGS-02<br>650269abd8e2e64630fe18031ef8868c25f9f6a8a9b92ac8bf582e7068d9e0e7 |
| **committed_or_manifest_sha256** | 650269abd8e2e64630fe18031ef8868c25f9f6a8a9b92ac8bf582e7068d9e0e7 |
| **prev_entry_sha256** | 2333cd8b64286c1e074ca5121bca005f522192824df980a92f474099579bc697 |
| **entry_line_sha256** | 1e0310a7de5932e4de2a6bff41f1f396f95f8ad7190c0ecbae85d9c2d466fb62 |

| Entry 187 | 2026-10-22 · COMMITMENT · manifest – |
|---|---|
| **governing_provisions** | PG-10 §12 (M-03A); PG-10 sec. 10.1, T6, T7, T31; P26-02(vi); CA2-32 |
| **subject** | RR-EGS-03<br>a6a0fdb33d8abd5ad71a76ce8a5beebc9b03119942389f695589e4ae885e8df7 |
| **committed_or_manifest_sha256** | a6a0fdb33d8abd5ad71a76ce8a5beebc9b03119942389f695589e4ae885e8df7 |
| **prev_entry_sha256** | 1e0310a7de5932e4de2a6bff41f1f396f95f8ad7190c0ecbae85d9c2d466fb62 |
| **entry_line_sha256** | febf4c5d23e8d9fa5d8500f78e11269d58661011a90d48857609120db3d47c29 |

| Entry 188 | 2026-10-22 · COMMITMENT · manifest – |
|---|---|
| **governing_provisions** | PG-10 §12 (M-03A); PG-10 sec. 10.1, T6, T7, T31; P26-02(vi); CA2-32 |
| **subject** | RR-EGS-04<br>09d99ea5b81b55ddab88111601579d8fb0b23c7de6c979fccec851a39ea5c9c1 |
| **committed_or_manifest_sha256** | 09d99ea5b81b55ddab88111601579d8fb0b23c7de6c979fccec851a39ea5c9c1 |
| **prev_entry_sha256** | febf4c5d23e8d9fa5d8500f78e11269d58661011a90d48857609120db3d47c29 |
| **entry_line_sha256** | 33a317161514ecc24ea8caa3501f4ab6b5ede31c733cb979ceea39656e4ca8b3 |

| Entry 189 | 2026-10-22 · COMMITMENT · manifest – |
|---|---|
| **governing_provisions** | PG-10 §12 (M-03A); PG-10 sec. 10.1, T6, T7, T31; P26-02(vi); CA2-32 |
| **subject** | RR-EGS-05<br>93a50522cee5307f008e6d5f5c94dfcfd02518ff95551e2720af4246a59ce5a0 |
| **committed_or_manifest_sha256** | 93a50522cee5307f008e6d5f5c94dfcfd02518ff95551e2720af4246a59ce5a0 |
| **prev_entry_sha256** | 33a317161514ecc24ea8caa3501f4ab6b5ede31c733cb979ceea39656e4ca8b3 |
| **entry_line_sha256** | f775424eb6b79a09b378ae8e6c9a64c9fa97e5b59ec9e9c7991cc81aedefc426 |

| Entry 190 | 2026-10-22 · VERIFICATION_RECORD · manifest – |
|---|---|
| **governing_provisions** | CA2-32; P26-18; PG-10 §12 (M-03A) |
| **committed_or_manifest_sha256** | – |
| **prev_entry_sha256** | f775424eb6b79a09b378ae8e6c9a64c9fa97e5b59ec9e9c7991cc81aedefc426 |
| **entry_line_sha256** | eeea0b9dcf6ab36b9e461808e5cbf0e8e0a83096e29d1dc7b38ad3a9565b0023 |

| Entry 191 | 2026-10-22 · VERIFICATION_RECORD · manifest – |
|---|---|
| **governing_provisions** | CA2-32; P26-18; PG-10 §12 (M-03A) |
| **committed_or_manifest_sha256** | – |
| **prev_entry_sha256** | eeea0b9dcf6ab36b9e461808e5cbf0e8e0a83096e29d1dc7b38ad3a9565b0023 |
| **entry_line_sha256** | 429b95e1e90a79efb908ba99d3cd61cbaff2d1b37d59b1a6855b8c828960d011 |

| Entry 192 | 2026-10-22 · VERIFICATION_RECORD · manifest – |
|---|---|
| **governing_provisions** | CA2-32; P26-18; PG-10 §12 (M-03A) |
| **committed_or_manifest_sha256** | – |
| **prev_entry_sha256** | 429b95e1e90a79efb908ba99d3cd61cbaff2d1b37d59b1a6855b8c828960d011 |
| **entry_line_sha256** | 085558a95c49cef5e69fc2fabb6050fe97581174a33a59cc5d20c4ed66d05649 |

| Entry 193 | 2026-10-22 · VERIFICATION_RECORD · manifest – |
|---|---|
| **governing_provisions** | CA2-32; P26-18; PG-10 §12 (M-03A) |
| **committed_or_manifest_sha256** | – |
| **prev_entry_sha256** | 085558a95c49cef5e69fc2fabb6050fe97581174a33a59cc5d20c4ed66d05649 |
| **entry_line_sha256** | 514d678feac3906918faf548f45f5bb149e674c78004ad1f6b9dab4f509e289f |

| Entry 194 | 2026-10-22 · VERIFICATION_RECORD · manifest – |
|---|---|
| **governing_provisions** | CA2-32; P26-18; PG-10 §12 (M-03A) |
| **committed_or_manifest_sha256** | – |
| **prev_entry_sha256** | 514d678feac3906918faf548f45f5bb149e674c78004ad1f6b9dab4f509e289f |
| **entry_line_sha256** | 3ac5f59ceb1eea09f9f6dc15ef99f1042a46abea3ccac26b99e86c5e9a0b805c |

| Entry 195 | 2026-10-22 · COMMITMENT · manifest – |
|---|---|
| **governing_provisions** | PG-10 §12 (M-03A); PG-10 sec. 10.1, T6, T7, T31; P26-02(vi); CA2-32 |
| **subject** | FIS-TF-EGS-01<br>5b4fc12cee20a4cfcf0660616ac9f50a8c5abe50374a4e68067cb60d768b5d2a |
| **committed_or_manifest_sha256** | 5b4fc12cee20a4cfcf0660616ac9f50a8c5abe50374a4e68067cb60d768b5d2a |
| **prev_entry_sha256** | 3ac5f59ceb1eea09f9f6dc15ef99f1042a46abea3ccac26b99e86c5e9a0b805c |
| **entry_line_sha256** | b94d888f089f9c1eb5dec29aac2f08ca45ad41438807fbdc62632b3f6bcdf0a2 |

| Entry 196 | 2026-10-22 · COMMITMENT · manifest – |
|---|---|
| governing_provisions | PG-10 §12 (M-03A); PG-10 sec. 10.1, T6, T7, T31; P26-02(vi); CA2-32 |
| subject | FIS-TF-EGS-02<br>f6be93e823a9a3b7e79c67e15d728d4d450126ee75918007d55b81fd25b88747 |
| committed_or_manifest_sha256 | f6be93e823a9a3b7e79c67e15d728d4d450126ee75918007d55b81fd25b88747 |
| prev_entry_sha256 | b94d888f089f9c1eb5dec29aac2f08ca45ad41438807fbdc62632b3f6bcdf0a2 |
| entry_line_sha256 | 4e2e359e042a6d954ec8200b385497971d7cce1d8d72a56b0ddb3803cea27417 |

| Entry 197 | 2026-10-22 · COMMITMENT · manifest – |
|---|---|
| governing_provisions | PG-10 §12 (M-03A); PG-10 sec. 10.1, T6, T7, T31; P26-02(vi); CA2-32 |
| subject | FIS-TF-EGS-03<br>274ae5fe7a88b001ecd8601b0aac50a962b96a84b821a3734d5da63eb0ca0862 |
| committed_or_manifest_sha256 | 274ae5fe7a88b001ecd8601b0aac50a962b96a84b821a3734d5da63eb0ca0862 |
| prev_entry_sha256 | 4e2e359e042a6d954ec8200b385497971d7cce1d8d72a56b0ddb3803cea27417 |
| entry_line_sha256 | 1fa344832ae98b356c391ed915673b6a5d3f43fb40e01818508151afe6e628a8 |

| Entry 198 | 2026-10-22 · COMMITMENT · manifest – |
|---|---|
| governing_provisions | PG-10 §12 (M-03A); PG-10 sec. 10.1, T6, T7, T31; P26-02(vi); CA2-32 |
| subject | FIS-TF-EGS-04<br>0dd9346cc929949807bd73144940edc744a6f33bc4af732ce0c1548f2ecb154f |
| committed_or_manifest_sha256 | 0dd9346cc929949807bd73144940edc744a6f33bc4af732ce0c1548f2ecb154f |
| prev_entry_sha256 | 1fa344832ae98b356c391ed915673b6a5d3f43fb40e01818508151afe6e628a8 |
| entry_line_sha256 | 40fdadc585bc02a1a3f7dc561bbe48f94fbdd158e4ae2d9a307dd921b7c468d4 |

| Entry 199 | 2026-10-22 · COMMITMENT · manifest – |
|---|---|
| governing_provisions | PG-10 §12 (M-03A); PG-10 sec. 10.1, T6, T7, T31; P26-02(vi); CA2-32 |
| subject | FIS-TF-EGS-05<br>44bb1890bdc2dc24e76cc833e3e7a8c04df4238b0b84de0f00835a79ffad4304 |
| committed_or_manifest_sha256 | 44bb1890bdc2dc24e76cc833e3e7a8c04df4238b0b84de0f00835a79ffad4304 |
| prev_entry_sha256 | 40fdadc585bc02a1a3f7dc561bbe48f94fbdd158e4ae2d9a307dd921b7c468d4 |
| entry_line_sha256 | a9ac508feca3e422d8d205ddc1dfa5974f08c614ee1e2fd593ed26372f773cc3 |

| Entry 200 | 2026-10-22 · VERIFICATION_RECORD · manifest – |
|---|---|
| governing_provisions | CA2-32; P26-18; PG-10 §12 (M-03A) |
| committed_or_manifest_sha256 | – |
| prev_entry_sha256 | a9ac508feca3e422d8d205ddc1dfa5974f08c614ee1e2fd593ed26372f773cc3 |
| entry_line_sha256 | 7cb75b25ead5e6dc5b7f86a535fe1b07342f0b7f96cd1881722889cd3461eddb |

| Entry 201 | 2026-10-22 · VERIFICATION_RECORD · manifest – |
|---|---|
| governing_provisions | CA2-32; P26-18; PG-10 §12 (M-03A) |
| committed_or_manifest_sha256 | – |
| prev_entry_sha256 | 7cb75b25ead5e6dc5b7f86a535fe1b07342f0b7f96cd1881722889cd3461eddb |
| entry_line_sha256 | 21c763e91bb0693b237eac9cbc00b1d0a93701213e90ed8c894d95d0e379f5f3 |

| Entry 202 | 2026-10-22 · VERIFICATION_RECORD · manifest – |
|---|---|
| governing_provisions | CA2-32; P26-18; PG-10 §12 (M-03A) |
| committed_or_manifest_sha256 | – |
| prev_entry_sha256 | 21c763e91bb0693b237eac9cbc00b1d0a93701213e90ed8c894d95d0e379f5f3 |
| entry_line_sha256 | 9dc3b3120924441c665bf3c1a282e767710d47413eb62eac569f434d7a151900 |

| Entry 203 | 2026-10-22 · VERIFICATION_RECORD · manifest – |
|---|---|
| governing_provisions | CA2-32; P26-18; PG-10 §12 (M-03A) |
| committed_or_manifest_sha256 | – |
| prev_entry_sha256 | 9dc3b3120924441c665bf3c1a282e767710d47413eb62eac569f434d7a151900 |
| entry_line_sha256 | 73dc417e636551f3b8ca59d331d8ef2be14f7dd581c4517d29d1beae1778123f |

| Entry 204 | 2026-10-22 · VERIFICATION_RECORD · manifest – |
|---|---|
| governing_provisions | CA2-32; P26-18; PG-10 §12 (M-03A) |
| committed_or_manifest_sha256 | – |
| prev_entry_sha256 | 73dc417e636551f3b8ca59d331d8ef2be14f7dd581c4517d29d1beae1778123f |
| entry_line_sha256 | 7fd6ae30462c266316fa638ce8fc00168ceb94b2612eb515fe426a235b1faeaa |

| Entry 205 | 2026-10-27 · COMMITMENT · manifest – |
|---|---|
| governing_provisions | PG-10 §12 (M-03A); PG-10 sec. 10.1, T6, T7, T31; P26-02(vi); CA2-32 |
| subject | RR-AAO-01<br>d7ab1deaec979e7bf2c1a5bb2a2e2c56cf40f21413972f163b45b27e4f85f985 |
| committed_or_manifest_sha256 | d7ab1deaec979e7bf2c1a5bb2a2e2c56cf40f21413972f163b45b27e4f85f985 |
| prev_entry_sha256 | 7fd6ae30462c266316fa638ce8fc00168ceb94b2612eb515fe426a235b1faeaa |
| entry_line_sha256 | a9fdf87da3bc665e1b88b79e22fb3d839a9de55e2a698384d0d64d5b155632f8 |

| Entry 206 | 2026-10-27 · COMMITMENT · manifest – |
|---|---|
| governing_provisions | PG-10 §12 (M-03A); PG-10 sec. 10.1, T6, T7, T31; P26-02(vi); CA2-32 |
| subject | RR-AAO-02<br>25ee7a5ce7a31a9a9bcc3c365268dedfa3dfca8fb4a6f0664ed69d3c58a0bbdc |
| committed_or_manifest_sha256 | 25ee7a5ce7a31a9a9bcc3c365268dedfa3dfca8fb4a6f0664ed69d3c58a0bbdc |
| prev_entry_sha256 | a9fdf87da3bc665e1b88b79e22fb3d839a9de55e2a698384d0d64d5b155632f8 |
| entry_line_sha256 | f5fffab1a394325152afdb696a0afaeff743a8969b9b2a41d841ade0d9134711 |

| Entry 207 | 2026-10-27 · COMMITMENT · manifest – |
|---|---|
| governing_provisions | PG-10 §12 (M-03A); PG-10 sec. 10.1, T6, T7, T31; P26-02(vi); CA2-32 |
| subject | RR-AAO-03<br>1fa2fef7a033adb179d24212edd0931a5794e0455d4e5294f5b37e88b9aaca56 |
| committed_or_manifest_sha256 | 1fa2fef7a033adb179d24212edd0931a5794e0455d4e5294f5b37e88b9aaca56 |
| prev_entry_sha256 | f5fffab1a394325152afdb696a0afaeff743a8969b9b2a41d841ade0d9134711 |
| entry_line_sha256 | 3966076de52e49db443577390eda8d384badcb040b554311f5ba499cde0c7f72 |

| Entry 208 | 2026-10-27 · COMMITMENT · manifest – |
|---|---|
| governing_provisions | PG-10 §12 (M-03A); PG-10 sec. 10.1, T6, T7, T31; P26-02(vi); CA2-32 |
| subject | RR-AAO-04<br>a7370d6f77b4aca97636e2d583ea98520a187f86909fe3d15b6079125cfe3c61 |
| committed_or_manifest_sha256 | a7370d6f77b4aca97636e2d583ea98520a187f86909fe3d15b6079125cfe3c61 |
| prev_entry_sha256 | 3966076de52e49db443577390eda8d384badcb040b554311f5ba499cde0c7f72 |
| entry_line_sha256 | 7014479147e7debe970b7cca8054d7a928bf9c69241e5bf387b171dc74b63044 |

| Entry 209 | 2026-10-27 · COMMITMENT · manifest – |
|---|---|
| governing_provisions | PG-10 §12 (M-03A); PG-10 sec. 10.1, T6, T7, T31; P26-02(vi); CA2-32 |
| subject | RR-AAO-05<br>2da572fc4ec8ccd0c99ffbf76d2aceb47d0fd2baa84a68f20cb0b8a21630a2a1 |
| committed_or_manifest_sha256 | 2da572fc4ec8ccd0c99ffbf76d2aceb47d0fd2baa84a68f20cb0b8a21630a2a1 |
| prev_entry_sha256 | 7014479147e7debe970b7cca8054d7a928bf9c69241e5bf387b171dc74b63044 |
| entry_line_sha256 | 1c9cd0086a0e602b265f3a7a736ebb458e1b7b814b40f679fa80210afe4fbc08 |

| Entry 210 | 2026-10-27 · VERIFICATION_RECORD · manifest – |
|---|---|
| governing_provisions | CA2-32; P26-18; PG-10 §12 (M-03A) |
| committed_or_manifest_sha256 | – |
| prev_entry_sha256 | 1c9cd0086a0e602b265f3a7a736ebb458e1b7b814b40f679fa80210afe4fbc08 |
| entry_line_sha256 | 3878f310069e04aaaa6f7f84ac02e6c8846b9a499342009dc0c1d1257bba7906 |

| Entry 211 | 2026-10-27 · VERIFICATION_RECORD · manifest – |
|---|---|
| governing_provisions | CA2-32; P26-18; PG-10 §12 (M-03A) |
| committed_or_manifest_sha256 | – |
| prev_entry_sha256 | 3878f310069e04aaaa6f7f84ac02e6c8846b9a499342009dc0c1d1257bba7906 |
| entry_line_sha256 | 0a09196fa2a34ef529f16580641b24ac00aefcaa86302ca092ad511d19b19124 |

| Entry 212 | 2026-10-27 · VERIFICATION_RECORD · manifest – |
|---|---|
| governing_provisions | CA2-32; P26-18; PG-10 §12 (M-03A) |
| committed_or_manifest_sha256 | – |
| prev_entry_sha256 | 0a09196fa2a34ef529f16580641b24ac00aefcaa86302ca092ad511d19b19124 |
| entry_line_sha256 | 16882e73dcaec70b6c46035ae4d4a5eb394db24b775a5af3c859ed11adebc2cf |

| Entry 213 | 2026-10-27 · VERIFICATION_RECORD · manifest – |
|---|---|
| governing_provisions | CA2-32; P26-18; PG-10 §12 (M-03A) |
| committed_or_manifest_sha256 | – |
| prev_entry_sha256 | 16882e73dcaec70b6c46035ae4d4a5eb394db24b775a5af3c859ed11adebc2cf |
| entry_line_sha256 | 70fb74793400828b89da73485938f15fb1d330a20545557c5bc31dbf31a7f8dc |

| Entry 214 | 2026-10-27 · VERIFICATION_RECORD · manifest – |
|---|---|
| governing_provisions | CA2-32; P26-18; PG-10 §12 (M-03A) |
| committed_or_manifest_sha256 | – |
| prev_entry_sha256 | 70fb74793400828b89da73485938f15fb1d330a20545557c5bc31dbf31a7f8dc |
| entry_line_sha256 | e6893518f66534d03e15cb9e98cbfea717359e46a2dba9a6ee704045a6b03b02 |

| Entry 215 | 2026-10-27 · COMMITMENT · manifest – |
|---|---|
| governing_provisions | PG-10 §12 (M-03A); PG-10 sec. 10.1, T6, T7, T31; P26-02(vi); CA2-32 |
| subject | FIS-TF-AAO-01<br>9ecd558d9838e8cd5b3dd2d3154a59c22825f23e9b65c526ca8d4c6fe6552413 |
| committed_or_manifest_sha256 | 9ecd558d9838e8cd5b3dd2d3154a59c22825f23e9b65c526ca8d4c6fe6552413 |
| prev_entry_sha256 | e6893518f66534d03e15cb9e98cbfea717359e46a2dba9a6ee704045a6b03b02 |
| entry_line_sha256 | ffc2330185b2fa0454b760b8b2c78cfce806bbf5cd395762732277bc318db8fb |

| Entry 216 | 2026-10-27 · COMMITMENT · manifest – |
|---|---|
| governing_provisions | PG-10 §12 (M-03A); PG-10 sec. 10.1, T6, T7, T31; P26-02(vi); CA2-32 |
| subject | FIS-TF-AAO-02<br>c22107922f447b7629b913e99ce1e01ee1b6c4a9b4155044f715ddccafe51cc1 |
| committed_or_manifest_sha256 | c22107922f447b7629b913e99ce1e01ee1b6c4a9b4155044f715ddccafe51cc1 |
| prev_entry_sha256 | ffc2330185b2fa0454b760b8b2c78cfce806bbf5cd395762732277bc318db8fb |
| entry_line_sha256 | 7824379f4111b26ff6f61a36fde66dc61a6c30e4388f59b284d424f39ff66b82 |

| Entry 217 | 2026-10-27 · COMMITMENT · manifest – |
|---|---|
| governing_provisions | PG-10 §12 (M-03A); PG-10 sec. 10.1, T6, T7, T31; P26-02(vi); CA2-32 |
| subject | FIS-TF-AAO-03<br>069253e8054c4444d693a03fa1c96211e56cbf3849a2721a7532ffe92f0a8a84 |
| committed_or_manifest_sha256 | 069253e8054c4444d693a03fa1c96211e56cbf3849a2721a7532ffe92f0a8a84 |
| prev_entry_sha256 | 7824379f4111b26ff6f61a36fde66dc61a6c30e4388f59b284d424f39ff66b82 |
| entry_line_sha256 | 3c58f2cd455ace934ea4abc556bb6c5a7775d1d59f2df19fb49adf2e6d7f0cfa |

| Entry 218 | 2026-10-27 · COMMITMENT · manifest – |
|---|---|
| **governing_provisions** | PG-10 §12 (M-03A); PG-10 sec. 10.1, T6, T7, T31; P26-02(vi); CA2-32 |
| **subject** | FIS-TF-AAO-04<br>4d3fa09f75c839c12bfca0f3fc4ecd7416d32fd310ccb7be5151460dd5c15e7d |
| **committed_or_manifest_sha256** | 4d3fa09f75c839c12bfca0f3fc4ecd7416d32fd310ccb7be5151460dd5c15e7d |
| **prev_entry_sha256** | 3c58f2cd455ace934ea4abc556bb6c5a7775d1d59f2df19fb49adf2e6d7f0cfa |
| **entry_line_sha256** | ad4d3b98921113b9f6dffdd5f475af6be03eeda4d944e798d461f4b463474102 |

| Entry 219 | 2026-10-27 · COMMITMENT · manifest – |
|---|---|
| **governing_provisions** | PG-10 §12 (M-03A); PG-10 sec. 10.1, T6, T7, T31; P26-02(vi); CA2-32 |
| **subject** | FIS-TF-AAO-05<br>eaab0e204da163c2399acd061c322adb6a3b295a9d4d096fe86958fbf36b1fbc |
| **committed_or_manifest_sha256** | eaab0e204da163c2399acd061c322adb6a3b295a9d4d096fe86958fbf36b1fbc |
| **prev_entry_sha256** | ad4d3b98921113b9f6dffdd5f475af6be03eeda4d944e798d461f4b463474102 |
| **entry_line_sha256** | 0164a3a7d7122d2656a4f98d45dbb23e74f6a983740d661e96aef272fb742e9a |

| Entry 220 | 2026-10-27 · VERIFICATION_RECORD · manifest – |
|---|---|
| **governing_provisions** | CA2-32; P26-18; PG-10 §12 (M-03A) |
| **committed_or_manifest_sha256** | – |
| **prev_entry_sha256** | 0164a3a7d7122d2656a4f98d45dbb23e74f6a983740d661e96aef272fb742e9a |
| **entry_line_sha256** | 89543b0ea3d6e5d053d6ae6a1c2a4545b2080281f277dda6fc5254d817d5d028 |

| Entry 221 | 2026-10-27 · VERIFICATION_RECORD · manifest – |
|---|---|
| **governing_provisions** | CA2-32; P26-18; PG-10 §12 (M-03A) |
| **committed_or_manifest_sha256** | – |
| **prev_entry_sha256** | 89543b0ea3d6e5d053d6ae6a1c2a4545b2080281f277dda6fc5254d817d5d028 |
| **entry_line_sha256** | e78c459a228df201fdb2a2c00a3bfd3ce8eba1e4bd3a244932a1747d6b1816c6 |

| Entry 222 | 2026-10-27 · VERIFICATION_RECORD · manifest – |
|---|---|
| **governing_provisions** | CA2-32; P26-18; PG-10 §12 (M-03A) |
| **committed_or_manifest_sha256** | – |
| **prev_entry_sha256** | e78c459a228df201fdb2a2c00a3bfd3ce8eba1e4bd3a244932a1747d6b1816c6 |
| **entry_line_sha256** | 04e80faf46108145a0a79d3632edfaebc0e24c84416ade303151163c06ebf3ae |

| Entry 223 | 2026-10-27 · VERIFICATION_RECORD · manifest – |
|---|---|
| **governing_provisions** | CA2-32; P26-18; PG-10 §12 (M-03A) |
| **committed_or_manifest_sha256** | – |
| **prev_entry_sha256** | 04e80faf46108145a0a79d3632edfaebc0e24c84416ade303151163c06ebf3ae |
| **entry_line_sha256** | 1a1e7c30088fc55174be451c1c52a720f3d65af12665028410b715ed7c1fa96a |

| Entry 224 | 2026-10-27 · VERIFICATION_RECORD · manifest – |
|---|---|
| **governing_provisions** | CA2-32; P26-18; PG-10 §12 (M-03A) |
| **committed_or_manifest_sha256** | – |
| **prev_entry_sha256** | 1a1e7c30088fc55174be451c1c52a720f3d65af12665028410b715ed7c1fa96a |
| **entry_line_sha256** | f3d08cdba91e935194d8186751e20076d311e6a4dc6130a9d14c491d51ad8be1 |

| Entry 225 | 2026-10-28 · ROLE_RECORD · manifest – |
|---|---|
| **governing_provisions** | P32-14; P32-13; P32-02; P26-14; P26-17; P26-18; P26-23; P26-24 |
| **subject** | PG32-D1<br>580f55ddfa86d653e1ab4a75e4b3cd4a50c16592e50a495c4dabbbf7e39c1d47<br>GATEKEEPER-REPORT-D1v13<br>4e72f6cfb0be5320be948569a508e467fd25ee1025569fb0c1281fda732c9aca<br>PG32-D1-v13-APPROVAL-AND-FREEZE-ACT<br>82b0c286155647c8ed9538770b42f7da0d18368dd386e1a50ac267f11ea69b4a |
| **committed_or_manifest_sha256** | – |
| **prev_entry_sha256** | f3d08cdba91e935194d8186751e20076d311e6a4dc6130a9d14c491d51ad8be1 |
| **entry_line_sha256** | 1b69a5138dcca153832b19b82a2cc494f4b049281e86cb32301f4f03f786af41 |

| Entry 226 | 2026-10-28 · VERIFICATION_RECORD · manifest – |
|---|---|
| **governing_provisions** | P26-18; P32-14; P32-13 |
| **subject** | PG32-D1<br>580f55ddfa86d653e1ab4a75e4b3cd4a50c16592e50a495c4dabbbf7e39c1d47 |
| **committed_or_manifest_sha256** | – |
| **prev_entry_sha256** | 1b69a5138dcca153832b19b82a2cc494f4b049281e86cb32301f4f03f786af41 |
| **entry_line_sha256** | 64d18274d2fd42748edae0074c9bb08cea08412be312be38f386a0e326ff3f5a |

| Entry 227 | 2026-10-28 · FREEZE_ACT · manifest M0014 |
|---|---|
| **governing_provisions** | P26-18; P26-14; P26-02(vi); P26-10(ii); P26-17 |
| **subject** | PG26-M0014<br>9e857c00e610f13685608ce092e557108f6e8e3e55d8ff971c47021c06470d65 |
| **committed_or_manifest_sha256** | 9e857c00e610f13685608ce092e557108f6e8e3e55d8ff971c47021c06470d65 |
| **prev_entry_sha256** | 64d18274d2fd42748edae0074c9bb08cea08412be312be38f386a0e326ff3f5a |
| **entry_line_sha256** | 6e25cf3790cb3256c5451d4ccf6d273e94dbd9ae95393778fadf6cc01f22c571 |

| Entry 228 | 2026-10-28 · COMMITMENT · manifest – |
|---|---|
| **governing_provisions** | P26-02(iii) as amended by CA2-10/CA3-10; P26-14; P26-10(ii) |
| **subject** | PG26-M0014<br>`9e857c00e610f13685608ce092e557108f6e8e3e55d8ff971c47021c06470d65` |
| **committed_or_manifest_sha256** | `9e857c00e610f13685608ce092e557108f6e8e3e55d8ff971c47021c06470d65` |
| **prev_entry_sha256** | `6e25cf3790cb3256c5451d4ccf6d273e94dbd9ae95393778fadf6cc01f22c571` |
| **entry_line_sha256** | `fcc6d9047af529c10a04283fa0e0e4717d02c0218633bbe5c63d3fd2d7be854a` |

| Entry 229 | 2026-10-28 · FREEZE_ACT · manifest M0015 |
|---|---|
| **governing_provisions** | P26-18; P26-14; P26-02(vi); P26-10(ii) |
| **subject** | PG26-M0015<br>`f5e4a8cfda8635f7c36fed54162b74b09b7d619bd702d2258f20284f1d48c74a` |
| **committed_or_manifest_sha256** | `f5e4a8cfda8635f7c36fed54162b74b09b7d619bd702d2258f20284f1d48c74a` |
| **prev_entry_sha256** | `fcc6d9047af529c10a04283fa0e0e4717d02c0218633bbe5c63d3fd2d7be854a` |
| **entry_line_sha256** | `8c4bc53188cccb6c5cbd62f41a42b5f13950dfb0b8ed47cdccc4587b8d41d0e7` |

| Entry 230 | 2026-10-28 · COMMITMENT · manifest – |
|---|---|
| **governing_provisions** | P26-02(iii) as amended by CA2-10/CA3-10; P26-14; P26-10(ii) |
| **subject** | PG26-M0015<br>`f5e4a8cfda8635f7c36fed54162b74b09b7d619bd702d2258f20284f1d48c74a` |
| **committed_or_manifest_sha256** | `f5e4a8cfda8635f7c36fed54162b74b09b7d619bd702d2258f20284f1d48c74a` |
| **prev_entry_sha256** | `8c4bc53188cccb6c5cbd62f41a42b5f13950dfb0b8ed47cdccc4587b8d41d0e7` |
| **entry_line_sha256** | `29b778310878b256f3281aa2ad3f0f2fb5e7f9f7a7fcb4fd1054139cfb2d2bab` |

| Entry 231 | 2026-10-28 · FREEZE_ACT · manifest M0016 |
|---|---|
| **governing_provisions** | P26-18; P26-14; P26-02(ii); P26-05(6); PG18-D4 |
| **subject** | PG26-M0016<br>`372a44cb5c4bdbdcec30f3b2c6da65e561c0ab2963a16cfb92d13fa3304be17f` |
| **committed_or_manifest_sha256** | `372a44cb5c4bdbdcec30f3b2c6da65e561c0ab2963a16cfb92d13fa3304be17f` |
| **prev_entry_sha256** | `29b778310878b256f3281aa2ad3f0f2fb5e7f9f7a7fcb4fd1054139cfb2d2bab` |
| **entry_line_sha256** | `af41da5d566395c0c3db1cbe805006162a914124f8f00027f913ea202c763e66` |

| Entry 232 | 2026-10-28 · COMMITMENT · manifest – |
|---|---|
| **governing_provisions** | P26-02(iii) as amended by CA2-10/CA3-10; P26-14 |
| **subject** | PG26-M0016<br>`372a44cb5c4bdbdcec30f3b2c6da65e561c0ab2963a16cfb92d13fa3304be17f` |
| **committed_or_manifest_sha256** | `372a44cb5c4bdbdcec30f3b2c6da65e561c0ab2963a16cfb92d13fa3304be17f` |
| **prev_entry_sha256** | `af41da5d566395c0c3db1cbe805006162a914124f8f00027f913ea202c763e66` |
| **entry_line_sha256** | `4cb216d35e110c882dbdf4d65f6d420db74c268e17704637d6744d5088aeec21` |

| Entry 233 | 2026-10-28 · VERIFICATION_RECORD · manifest – |
|---|---|
| **governing_provisions** | P26-18; P26-14; P26-12 |
| **subject** | PG26-M0014<br>`9e857c00e610f13685608ce092e557108f6e8e3e55d8ff971c47021c06470d65` |
| **committed_or_manifest_sha256** | – |
| **prev_entry_sha256** | `4cb216d35e110c882dbdf4d65f6d420db74c268e17704637d6744d5088aeec21` |
| **entry_line_sha256** | `310382da81211092c2f430d08df05a0df10b826bd0f31a563d2ea3c5e99cf5ad` |

| Entry 234 | 2026-10-28 · VERIFICATION_RECORD · manifest – |
|---|---|
| **governing_provisions** | P26-18; P26-14; P26-12 |
| **subject** | PG26-M0015<br>`f5e4a8cfda8635f7c36fed54162b74b09b7d619bd702d2258f20284f1d48c74a` |
| **committed_or_manifest_sha256** | – |
| **prev_entry_sha256** | `310382da81211092c2f430d08df05a0df10b826bd0f31a563d2ea3c5e99cf5ad` |
| **entry_line_sha256** | `6159032bc3f47f9a13ed5f686ddf9c4df9eb50e600f12fc1485e576a375b079c` |

| Entry 235 | 2026-10-28 · VERIFICATION_RECORD · manifest – |
|---|---|
| **governing_provisions** | P26-18; P26-14; P26-12 |
| **subject** | PG26-M0016<br>`372a44cb5c4bdbdcec30f3b2c6da65e561c0ab2963a16cfb92d13fa3304be17f` |
| **committed_or_manifest_sha256** | – |
| **prev_entry_sha256** | `6159032bc3f47f9a13ed5f686ddf9c4df9eb50e600f12fc1485e576a375b079c` |
| **entry_line_sha256** | `df6196679c97ba95d65fd0f9de3a0f9fcacfab99916967ac061643a34aa41580` |

| Entry 236 | 2026-10-28 · COMMITMENT · manifest – |
|---|---|
| **governing_provisions** | PG-16 OV-3; FPA-6 v0.3; P26-05(4); P26-02(iii) as amended by CA2-10/CA3-10; CA2-32 |
| **subject** | SEED-R<br>`abe257c30e23edc21f31b18dc7b5ecbb37b56c9df7119075286ba90ef3a7707d` |
| **committed_or_manifest_sha256** | `abe257c30e23edc21f31b18dc7b5ecbb37b56c9df7119075286ba90ef3a7707d` |
| **prev_entry_sha256** | `df6196679c97ba95d65fd0f9de3a0f9fcacfab99916967ac061643a34aa41580` |
| **entry_line_sha256** | `e74fa757de53d7730dc7ef3a53cc22a644b61e5462e6bddbb4006b02a16ea120` |

| Entry 237 | 2026-10-28 · VERIFICATION_RECORD · manifest – |
|---|---|
| **governing_provisions** | PG-16 OV-3; FPA-6 v0.3 sec. 3.2; P26-18; CA2-32 |
| **subject** | SEED-R<br>abe257c30e23edc21f31b18dc7b5ecbb37b56c9df7119075286ba90ef3a7707d |
| **committed_or_manifest_sha256** | – |
| **prev_entry_sha256** | e74fa757de53d7730dc7ef3a53cc22a644b61e5462e6bddbb4006b02a16ea120 |
| **entry_line_sha256** | 9e7e1248604023f6d0297361f3315b10257255486b8f5b8a617d95fbcadf7c35 |

| Entry 238 | 2026-10-28 · FREEZE_ACT · manifest M0017 |
|---|---|
| **governing_provisions** | P26-18; P26-05; P26-02(vi); PG-16 OV-1; PG-09 sec. 27 |
| **subject** | PG26-M0017<br>ccb43de2880aeed69a4e03e114646eaadc9d13977506c548b75c0b3bbd6c6052 |
| **committed_or_manifest_sha256** | ccb43de2880aeed69a4e03e114646eaadc9d13977506c548b75c0b3bbd6c6052 |
| **prev_entry_sha256** | 9e7e1248604023f6d0297361f3315b10257255486b8f5b8a617d95fbcadf7c35 |
| **entry_line_sha256** | 31ff37eddfc50aa62f72d9ac79960241a63bc2f1f0a6a8115b3e591ae9d8693a |

| Entry 239 | 2026-10-28 · COMMITMENT · manifest – |
|---|---|
| **governing_provisions** | P26-02(iii) as amended by CA2-10/CA3-10; P26-05 |
| **subject** | PG26-M0017<br>ccb43de2880aeed69a4e03e114646eaadc9d13977506c548b75c0b3bbd6c6052 |
| **committed_or_manifest_sha256** | ccb43de2880aeed69a4e03e114646eaadc9d13977506c548b75c0b3bbd6c6052 |
| **prev_entry_sha256** | 31ff37eddfc50aa62f72d9ac79960241a63bc2f1f0a6a8115b3e591ae9d8693a |
| **entry_line_sha256** | 79aee6ccc2574a077f5470fe97cd2ba646de7dbdefcd0c6fb139a50337bfca8b |

| Entry 240 | 2026-10-28 · VERIFICATION_RECORD · manifest – |
|---|---|
| **governing_provisions** | P26-18; P26-05; P26-12 |
| **subject** | PG26-M0017<br>ccb43de2880aeed69a4e03e114646eaadc9d13977506c548b75c0b3bbd6c6052 |
| **committed_or_manifest_sha256** | – |
| **prev_entry_sha256** | 79aee6ccc2574a077f5470fe97cd2ba646de7dbdefcd0c6fb139a50337bfca8b |
| **entry_line_sha256** | 1c80c102c6ebd1be4d5c85b772986471e33de25310eecc3668680e1edab2a616 |

| Entry 241 | 2026-10-28 · COMMITMENT · manifest – |
|---|---|
| **governing_provisions** | PG-16 OV-3; PG-16 OV-1; FPA-6 v0.3 sec. 5; P26-03; CA2-32 |
| **subject** | SEED-R-ACCESS-LOG<br>b5e2b9ce8066b25ab451e4038e853d8271249ccfc370c550a0d077b81527a11a<br>OV3-SELECTION-RECORD<br>3f6e4656eeb49522284e0b1aedb4f9927be3d02dfeeb879fcb07830d465ad05c |
| **committed_or_manifest_sha256** | b5e2b9ce8066b25ab451e4038e853d8271249ccfc370c550a0d077b81527a11a |
| **prev_entry_sha256** | 1c80c102c6ebd1be4d5c85b772986471e33de25310eecc3668680e1edab2a616 |
| **entry_line_sha256** | 6543b2cca64e401836facfb98a34656d6a32f914f06b94b6b5000ba7689ddb68 |

| Entry 242 | 2026-10-28 · COMMITMENT · manifest – |
|---|---|
| **governing_provisions** | PG-30 P30-10; PG-30 P30-12; PG-30 P30-14; PG-30 P30-17; P26-19; P29-22 |
| **subject** | PG30-TARGET-JOURNAL-LIST-LOCK<br>131c3675b10a78106b3659afc90109fe24ebe8d6939c57f9854ffa9ead389c08<br>PG30-LOCK-PI-ACT<br>121b2cf97ed54f5f5f82ea1c95c6b5f331060c39351d59453e350b91cc5bbcc8 |
| **committed_or_manifest_sha256** | 131c3675b10a78106b3659afc90109fe24ebe8d6939c57f9854ffa9ead389c08 |
| **prev_entry_sha256** | 6543b2cca64e401836facfb98a34656d6a32f914f06b94b6b5000ba7689ddb68 |
| **entry_line_sha256** | 5ba5d3adce145167fa7211d0e95447f817da3e8e8dadf2521c382ddb28b0cfb1 |

| Entry 243 | 2026-10-28 · VERIFICATION_RECORD · manifest – |
|---|---|
| **governing_provisions** | PG-16 OV-3; FPA-6 v0.3 sec. 5.2; P26-18; CA2-32 |
| **subject** | SEED-R<br>abe257c30e23edc21f31b18dc7b5ecbb37b56c9df7119075286ba90ef3a7707d<br>OV3-SELECTION-RECORD<br>3f6e4656eeb49522284e0b1aedb4f9927be3d02dfeeb879fcb07830d465ad05c |
| **committed_or_manifest_sha256** | – |
| **prev_entry_sha256** | 5ba5d3adce145167fa7211d0e95447f817da3e8e8dadf2521c382ddb28b0cfb1 |
| **entry_line_sha256** | 63a7401abf1c1d23832ae55ee7e46771ee326879909e4091416995faf7105f35 |

| Entry 244 | 2026-10-28 · VERIFICATION_RECORD · manifest – |
|---|---|
| **governing_provisions** | PG-30 P30-14; PG-30 P30-17; P26-18; CA2-32 |
| **subject** | PG30-TARGET-JOURNAL-LIST-LOCK<br>131c3675b10a78106b3659afc90109fe24ebe8d6939c57f9854ffa9ead389c08 |
| **committed_or_manifest_sha256** | – |
| **prev_entry_sha256** | 63a7401abf1c1d23832ae55ee7e46771ee326879909e4091416995faf7105f35 |
| **entry_line_sha256** | f27778e4c09655ff272d4814c9dd770afee287fcc886983681e28886e071de6b |

| Entry 245 | 2026-10-28 · COMMITMENT · manifest – |
| --- | --- |
| **governing_provisions** | PG-16 P16-17; PG-16 P16-22; PG-16 P16-27; PG-16 §9.4; PG-14 E-17; D-1 v13 I-K |
| **subject** | PG16-REVIEWER-ROLE-RECORD<br>94a7b3149c8718c8f589e2acc465745832a69ba180c6cae746eba0f838d6bd71<br>PG16-F1-RELEASE-PACKAGE-MANIFEST<br>cc1c0b5033f3fc3e006e45276086b403f3a3f3f75ccf10a75a457bc2e6c538ab<br>PG16-MATCHING-PI-ACT<br>777d7501a5a6ce111ed4233ec58120e1b2d9360fee2c15324fd12dda40b33a55 |
| **committed_or_manifest_sha256** | 94a7b3149c8718c8f589e2acc465745832a69ba180c6cae746eba0f838d6bd71 |
| **prev_entry_sha256** | f27778e4c09655ff272d4814c9dd770afee287fcc886983681e28886e071de6b |
| **entry_line_sha256** | c6cc246a7ccf21e807a1b3c6da2e797f9a460d8e58a3bb2ce0d4f37e0882757c |

| Entry 246 | 2026-10-28 · VERIFICATION_RECORD · manifest – |
| --- | --- |
| **governing_provisions** | PG-16 P16-27; PG-16 §9.4; P26-18; CA2-32 |
| **subject** | PG16-REVIEWER-ROLE-RECORD<br>94a7b3149c8718c8f589e2acc465745832a69ba180c6cae746eba0f838d6bd71 |
| **committed_or_manifest_sha256** | – |
| **prev_entry_sha256** | c6cc246a7ccf21e807a1b3c6da2e797f9a460d8e58a3bb2ce0d4f37e0882757c |
| **entry_line_sha256** | 45a1d7f86da7d841d1e0c772b71a11c9f2af37c7ee66d9fd4b834f741d528fb2 |

| Entry 247 | 2026-10-28 · COMMITMENT · manifest – |
| --- | --- |
| **governing_provisions** | PG-16 P16-13; PG-16 P16-22; P26-14; D-1 v13 I-K |
| **subject** | F2-BUILD-INVENTORY<br>484f6aa9c558668caf075db2e1a398f824a809a651a55e9488561c234b43cdb2<br>F2-RELEASE-RA<br>7a72b6bff946e89ad540819d6af65ba7f15f18f1c279198114aa4ff9dc7dfba0<br>F2-RELEASE-RB<br>94601d98944642d1336994af58d132d1516d4130e401e3f2c9146adae66f1fea<br>F2-RELEASE-PI-ACT<br>1308b60e537de6af451db7c2e5183956084280f22ea8ce798876e55fb57af3e1 |
| **committed_or_manifest_sha256** | 484f6aa9c558668caf075db2e1a398f824a809a651a55e9488561c234b43cdb2 |
| **prev_entry_sha256** | 45a1d7f86da7d841d1e0c772b71a11c9f2af37c7ee66d9fd4b834f741d528fb2 |
| **entry_line_sha256** | 52b6320f606196a1cda6fed24fccf33c3629da8e581bb4ee155ec0330bf2d092 |

| Entry 248 | 2026-10-29 · VERIFICATION_RECORD · manifest – |
| --- | --- |
| **governing_provisions** | P26-18; CA2-32; PG-16 P16-13; PG-16 P16-22 |
| **subject** | F2-BUILD-INVENTORY<br>484f6aa9c558668caf075db2e1a398f824a809a651a55e9488561c234b43cdb2<br>F2-RELEASE-RA<br>7a72b6bff946e89ad540819d6af65ba7f15f18f1c279198114aa4ff9dc7dfba0<br>F2-RELEASE-RB<br>94601d98944642d1336994af58d132d1516d4130e401e3f2c9146adae66f1fea<br>F2-RELEASE-PI-ACT<br>1308b60e537de6af451db7c2e5183956084280f22ea8ce798876e55fb57af3e1 |
| **committed_or_manifest_sha256** | – |
| **prev_entry_sha256** | 52b6320f606196a1cda6fed24fccf33c3629da8e581bb4ee155ec0330bf2d092 |
| **entry_line_sha256** | c829811d2dab506c1b626c8aed9b89577f2671f7375faf7054ba8aa969cee5a4 |

| Entry 249 | 2026-10-29 · COMMITMENT · manifest – |
| --- | --- |
| **governing_provisions** | PG-16 P16-15; PG-16 P16-19; P26-14; D-1 v13 I-K |
| **subject** | SEAL-SEC-7a48f507a760f832<br>7a48f507a760f8328327bca552b349f4c38d9b8f89331028dc42f9a7755ab1fe<br>SEAL-O2_SEAL1-d800adecc403c474<br>d800adecc403c4742517e4bea0a0be1cca6dd7fea33ffc9e7dfda04f42ae99bc<br>SEAL-X1-1e641ffd6ab3bec3<br>1e641ffd6ab3bec361bd64c7c87364906dd8d46a57dc8219f30982220f13fc41<br>RV-CUSTODY-LOG<br>9d65260a7bdf7d0fba4d59c21a303c79b021cae672023795ca56fdb10acbf392 |
| **committed_or_manifest_sha256** | 9d65260a7bdf7d0fba4d59c21a303c79b021cae672023795ca56fdb10acbf392 |
| **prev_entry_sha256** | c829811d2dab506c1b626c8aed9b89577f2671f7375faf7054ba8aa969cee5a4 |
| **entry_line_sha256** | 4ccc5ee8eeb0e9275dccdd88d1005f959c8447f134349c3a59386b500d76e8fe |

| Entry 250 | 2026-10-29 · VERIFICATION_RECORD · manifest – |
| --- | --- |
| **governing_provisions** | P26-18; CA2-32; PG-16 P16-15; PG-16 P16-19 |
| **subject** | SEAL-SEC-7a48f507a760f832<br>7a48f507a760f8328327bca552b349f4c38d9b8f89331028dc42f9a7755ab1fe<br>SEAL-O2_SEAL1-d800adecc403c474<br>d800adecc403c4742517e4bea0a0be1cca6dd7fea33ffc9e7dfda04f42ae99bc<br>SEAL-X1-1e641ffd6ab3bec3<br>1e641ffd6ab3bec361bd64c7c87364906dd8d46a57dc8219f30982220f13fc41<br>RV-CUSTODY-LOG<br>9d65260a7bdf7d0fba4d59c21a303c79b021cae672023795ca56fdb10acbf392 |
| **committed_or_manifest_sha256** | – |
| **prev_entry_sha256** | 4ccc5ee8eeb0e9275dccdd88d1005f959c8447f134349c3a59386b500d76e8fe |
| **entry_line_sha256** | 8f96a17a83acd1a8282a366ba8422747d3846d2342727aa89c792e54790ddc08 |

| Entry 251 | 2026-10-29 · COMMITMENT · manifest – |
|---|---|
| **governing_provisions** | PG-16 P16-15; PG-16 P16-19; P26-14; D-1 v13 I-K |
| **subject** | SEAL-O2_SEAL2-fd0b21de9339a5d6<br>fd0b21de9339a5d6a861d295a680311db439dd5cd0b8c71648a71a4b3fac6fce<br>SEAL-RA1-484bfa9466d29569<br>484bfa9466d2956953e32e67433032eaa3b6d7d27390a28940b1be8c5d1027c3<br>SEAL-OVERLAP-007d9eb757458776<br>007d9eb757458776f3ae777d0121c60f778d3ecb40fe93615d01dafc0759dc09<br>RV-CUSTODY-LOG<br>f6a20a8f080455ce8eb8f9c1cbc5725b6b18c27e7cd3b06c0ab449dd51514450 |
| **committed_or_manifest_sha256** | f6a20a8f080455ce8eb8f9c1cbc5725b6b18c27e7cd3b06c0ab449dd51514450 |
| **prev_entry_sha256** | 8f96a17a83acd1a8282a366ba8422747d3846d2342727aa89c792e54790ddc08 |
| **entry_line_sha256** | e35786eb68bc11ab43b7ad5f871f25deeb6616548567f52796ac3ee25ac33fd4 |

| Entry 252 | 2026-10-29 · VERIFICATION_RECORD · manifest – |
|---|---|
| **governing_provisions** | P26-18; CA2-32; PG-16 P16-15; PG-16 P16-19 |
| **subject** | SEAL-O2_SEAL2-fd0b21de9339a5d6<br>fd0b21de9339a5d6a861d295a680311db439dd5cd0b8c71648a71a4b3fac6fce<br>SEAL-RA1-484bfa9466d29569<br>484bfa9466d2956953e32e67433032eaa3b6d7d27390a28940b1be8c5d1027c3<br>SEAL-OVERLAP-007d9eb757458776<br>007d9eb757458776f3ae777d0121c60f778d3ecb40fe93615d01dafc0759dc09<br>RV-CUSTODY-LOG<br>f6a20a8f080455ce8eb8f9c1cbc5725b6b18c27e7cd3b06c0ab449dd51514450 |
| **committed_or_manifest_sha256** | – |
| **prev_entry_sha256** | e35786eb68bc11ab43b7ad5f871f25deeb6616548567f52796ac3ee25ac33fd4 |
| **entry_line_sha256** | fbb4ad7d3152379571390fff79ccdb4a5469ef71e5231d1535c6531749157303 |

| Entry 253 | 2026-10-29 · COMMITMENT · manifest – |
|---|---|
| **governing_provisions** | PG-16 P16-15; PG-16 P16-19; P26-14; D-1 v13 I-K |
| **subject** | SEAL-RA_ITEMS-38cf235379b69749<br>38cf235379b6974954dcc9672e90bed78fae1a3ef774ee24794b59d62ea138f8<br>RV-CUSTODY-LOG<br>ad7aff77520e4da77609c1b5fb15129f12097cfed39ae8a124d866b7cc090b18 |
| **committed_or_manifest_sha256** | ad7aff77520e4da77609c1b5fb15129f12097cfed39ae8a124d866b7cc090b18 |
| **prev_entry_sha256** | fbb4ad7d3152379571390fff79ccdb4a5469ef71e5231d1535c6531749157303 |
| **entry_line_sha256** | 767451300763eda768e1c135aa758578515c28fcbf346fc41bc614dae6830978 |

| Entry 254 | 2026-10-29 · VERIFICATION_RECORD · manifest – |
|---|---|
| **governing_provisions** | P26-18; CA2-32; PG-16 P16-15; PG-16 P16-19 |
| **subject** | SEAL-RA_ITEMS-38cf235379b69749<br>38cf235379b6974954dcc9672e90bed78fae1a3ef774ee24794b59d62ea138f8<br>RV-CUSTODY-LOG<br>ad7aff77520e4da77609c1b5fb15129f12097cfed39ae8a124d866b7cc090b18 |
| **committed_or_manifest_sha256** | – |
| **prev_entry_sha256** | 767451300763eda768e1c135aa758578515c28fcbf346fc41bc614dae6830978 |
| **entry_line_sha256** | 615d389e0612fb1d6fb9744b98fa1b44a8f8f273ca73f6163d051130b7958096 |

| Entry 255 | 2026-10-29 · COMMITMENT · manifest – |
|---|---|
| **governing_provisions** | P26-14; D-1 v13 I-K |
| **subject** | RII-ELIGIBILITY-DETERMINATION-V2<br>682cdf35f32329de4719ea3e297cee6b013a2cbec7d5adbbdcd848c8076bf3d1<br>RII-EMPTY-SET-ATTESTATION-RA-V2<br>816cdcaedf095f62adf4335dab275c4ca21d5d3d93ed8fd2136759138da7ef4d<br>RII-EMPTY-SET-ATTESTATION-RB-V2<br>2f6ee11fc8614bfe3273b7337a73316ddd4b05bb01cd76bb76702bb5208763be |
| **committed_or_manifest_sha256** | 46bec58ada6be5fcfa147da1b704329a35d0f36c55742483872cec9647d46afb |
| **prev_entry_sha256** | 615d389e0612fb1d6fb9744b98fa1b44a8f8f273ca73f6163d051130b7958096 |
| **entry_line_sha256** | 73c8902169bfc42fdd66f072766634c2db60ea9679562caf7951b86f14ec3d97 |

| Entry 256 | 2026-10-29 · COMMITMENT · manifest – |
|---|---|
| **governing_provisions** | P26-14; D-1 v13 I-K |
| **subject** | RII-ZERO-ENTRY-FINAL-SEAL-RA-V2<br>01d8534ab2666f4cb914ede3eb76b8aa37d653de75661dba8331f0c4cb1019fa<br>RII-ZERO-ENTRY-FINAL-SEAL-RB-V2<br>ddce8822f50567c190259d0bd3da15eec83394e78a921550dbb073e7af554285<br>RII-NOTICE-V2-TEXT<br>153df25a2cf8625c1d8b0490031375f4361c3cdecbd2a19da446ec510bb7fe0f |
| **committed_or_manifest_sha256** | e031e3aa5b136336543a262ab8ab06d5aac1fef8e2d6ff819f79dec059888eff |
| **prev_entry_sha256** | 73c8902169bfc42fdd66f072766634c2db60ea9679562caf7951b86f14ec3d97 |
| **entry_line_sha256** | bcbc1729dac19b54d12805c472339d92c633f34458dcadec7751fe1c9ad7ae26 |

| Entry 257 | 2026-10-29 · VERIFICATION_RECORD · manifest – |
|---|---|
| governing_provisions | P26-18; CA2-32; P26-14; D-1 v13 I-K |
| subject | RII-ELIGIBILITY-DETERMINATION-V2<br>682cdf35f32329de4719ea3e297cee6b013a2cbec7d5adbbdcd848c8076bf3d1<br>RII-EMPTY-SET-ATTESTATION-RA-V2<br>816cdcaedf095f62adf4335dab275c4ca21d5d3d93ed8fd2136759138da7ef4d<br>RII-EMPTY-SET-ATTESTATION-RB-V2<br>2f6ee11fc8614bfe3273b7337a73316ddd4b05bb01cd76bb76702bb5208763be |
| committed_or_manifest_sha256 | – |
| prev_entry_sha256 | bcbc1729dac19b54d12805c472339d92c633f34458dcadec7751fe1c9ad7ae26 |
| entry_line_sha256 | 6d5a0805440cfa417d89f359d222d4d277bf78321bb4eb785db4ea3b74783b8a |

| Entry 258 | 2026-10-29 · VERIFICATION_RECORD · manifest – |
|---|---|
| governing_provisions | P26-18; CA2-32; P26-14; D-1 v13 I-K |
| subject | RII-ZERO-ENTRY-FINAL-SEAL-RA-V2<br>01d8534ab2666f4cb914ede3eb76b8aa37d653de75661dba8331f0c4cb1019fa<br>RII-ZERO-ENTRY-FINAL-SEAL-RB-V2<br>ddce8822f50567c190259d0bd3da15eec83394e78a921550dbb073e7af554285<br>RII-NOTICE-V2-TEXT<br>153df25a2cf8625c1d8b0490031375f4361c3cdecbd2a19da446ec510bb7fe0f |
| committed_or_manifest_sha256 | – |
| prev_entry_sha256 | 6d5a0805440cfa417d89f359d222d4d277bf78321bb4eb785db4ea3b74783b8a |
| entry_line_sha256 | e14c0c666c883b217d0356f907237727edae647aee3bba3162bbdd905b7ea396 |

| Entry 259 | 2026-10-29 · COMMITMENT · manifest – |
|---|---|
| governing_provisions | P26-14; D-1 v13 I-K |
| subject | PG19-DISPOSITION-RECORDS-S1<br>91e62c032938a506b01eb80f5508ba4e1c716df5d0732c06905d176bfa0f4aea<br>PG19-DISPOSITION-COMPLETENESS-CHECK-S1<br>485e51c5018a4f9a9d7af6024063d22f5cd4bcc51ef3c9881b5a0304136af9ba |
| committed_or_manifest_sha256 | 3102b1919b2ea4a5f4dd4cdab2a27ef0fced512847037a05fd59e1545a07eaaf |
| prev_entry_sha256 | e14c0c666c883b217d0356f907237727edae647aee3bba3162bbdd905b7ea396 |
| entry_line_sha256 | f84fa32c5b78dea86b3b03841bda0b689f7bfb28686b32db992e4568be8e876b |

| Entry 260 | 2026-10-29 · COMMITMENT · manifest – |
|---|---|
| governing_provisions | P26-14; D-1 v13 I-K |
| subject | PG03-S19-PF-NICE-0001-FINAL-ACT<br>cc60fac462601212bd992e7179eca613beecadce182a7235364fcd7ff77ed320<br>PG03-S19-PF-NICE-0001-ROW<br>20c45999d7e0e71752f59e86fa586057e0a616d1ba6d8f4b9e98009e725b4fac<br>PG03-S19-PF-NICE-0001-AMENDMENT<br>72059111ed57395332099bf85bf856b0a292d57da2067b16f6e59227a5dba2e5<br>PG03-S19-PF-NICE-0001-PI-ADOPTION<br>ffc64a93875dfbeadfad478acc1269024dbfda6bfb4d337d8ea69c41e45dab5e |
| committed_or_manifest_sha256 | e4ae703be66d6d42a8ef4aa7fb884be7e63f748b13746f985e32796e6d5945cf |
| prev_entry_sha256 | f84fa32c5b78dea86b3b03841bda0b689f7bfb28686b32db992e4568be8e876b |
| entry_line_sha256 | f82b99cb69674f6e73db98d9b895b355e1de3a8faaf9b7c24da88ad6908694a8 |

| Entry 261 | 2026-10-29 · COMMITMENT · manifest – |
|---|---|
| governing_provisions | P26-14; D-1 v13 I-K |
| subject | PG19-DA2-S2-TRANSCRIPTION-PAGE34<br>30ae34020c41761c4058bd2c57096e3c3188635ff81acd7e57063a4dfe75715d |
| committed_or_manifest_sha256 | 77d09373e71b6395eb08f37e2adb12fad40abf5f6543e7326fda07cbecb15fa2 |
| prev_entry_sha256 | f82b99cb69674f6e73db98d9b895b355e1de3a8faaf9b7c24da88ad6908694a8 |
| entry_line_sha256 | c2c2cd6944b5ad40560cd768a3534b1ce1697a5acc7010ce8fd53bea95988653 |

| Entry 262 | 2026-10-30 · COMMITMENT · manifest – |
|---|---|
| governing_provisions | P26-14; D-1 v13 I-K |
| subject | E03-NICE-0032C01-NOSUCC-001-AND-DA2-TRANSCRIPTION<br>18f424977a52ca939d9c1a44e64195e906ebcfb0dd0074590f3b5807314c7e3a<br>DA2-DECISION-ROUTE1-SUPERSEDED<br>a8fb6c233f793bfd619ddca961b5ec8969dd9ce4e7713053bab338072258f4dd<br>TRIANGULATION-OV001-PROMPT<br>549491718dea34a5bdeda8f3c3bf8764c93c9e5eeb5e2a5cf779464b1d9cbc87<br>TRIANGULATION-OV001-ANSWER<br>e50c3007eeff151c764928a5643c943a970e92fe562a2b5e13eeb4f3331c72ec |
| committed_or_manifest_sha256 | 6be104779979b465e24884f844dc33f5d513aae1d2dcd2513b1352dc64b7b71c |
| prev_entry_sha256 | c2c2cd6944b5ad40560cd768a3534b1ce1697a5acc7010ce8fd53bea95988653 |
| entry_line_sha256 | 52c3ba6503ddd7ffbfeb9b4103490c34972b74f6130319f485ad2e06d0bcf190 |

| Entry 263 | 2026-10-30 · VERIFICATION_RECORD · manifest – |
|---|---|
| governing_provisions | P26-18; CA2-32; P26-14; D-1 v13 I-K |
| subject | PG19-DISPOSITION-RECORDS-S1<br>91e62c032938a506b01eb80f5508ba4e1c716df5d0732c06905d176bfa0f4aea<br>PG19-DISPOSITION-COMPLETENESS-CHECK-S1<br>485e51c5018a4f9a9d7af6024063d22f5cd4bcc51ef3c9881b5a0304136af9ba |
| committed_or_manifest_sha256 | – |
| prev_entry_sha256 | 52c3ba6503ddd7ffbfeb9b4103490c34972b74f6130319f485ad2e06d0bcf190 |
| entry_line_sha256 | ba2175a5cd459549e63f160475b79739989129be4e24942d166b53f6c9433f6d |

| Entry 264 | 2026-10-30 · VERIFICATION_RECORD · manifest – |
|---|---|
| governing_provisions | P26-18; CA2-32; P26-14; D-1 v13 I-K |
| subject | PG03-S19-PF-NICE-0001-FINAL-ACT<br>cc60fac462601212bd992e7179eca613beecadce182a7235364fcd7ff77ed320<br>PG03-S19-PF-NICE-0001-ROW<br>20c45999d7e0e71752f59e86fa586057e0a616d1ba6d8f4b9e98009e725b4fac<br>PG03-S19-PF-NICE-0001-AMENDMENT<br>72059111ed57395332099bf85bf856b0a292d57da2067b16f6e59227a5dba2e5<br>PG03-S19-PF-NICE-0001-PI-ADOPTION<br>ffc64a93875dfbeadfad478acc1269024dbfda6bfb4d337d8ea69c41e45dab5e |
| committed_or_manifest_sha256 | – |
| prev_entry_sha256 | ba2175a5cd459549e63f160475b79739989129be4e24942d166b53f6c9433f6d |
| entry_line_sha256 | 64876163fd93a169814fd01c82063f8878b6b88666aba4ff2d1db6c53444953f |

| Entry 265 | 2026-10-30 · VERIFICATION_RECORD · manifest – |
|---|---|
| governing_provisions | P26-18; CA2-32; P26-14; D-1 v13 I-K |
| subject | PG19-DA2-S2-TRANSCRIPTION-PAGE34<br>30ae34020c41761c4058bd2c57096e3c3188635ff81acd7e57063a4dfe75715d |
| committed_or_manifest_sha256 | – |
| prev_entry_sha256 | 64876163fd93a169814fd01c82063f8878b6b88666aba4ff2d1db6c53444953f |
| entry_line_sha256 | 8fef4189fca8e91959d208671f5b60b3ee40840f2a6769e568685a92384f1853 |

| Entry 266 | 2026-10-30 · VERIFICATION_RECORD · manifest – |
|---|---|
| governing_provisions | P26-18; CA2-32; P26-14; D-1 v13 I-K |
| subject | E03-NICE-0032C01-NOSUCC-001-AND-DA2-TRANSCRIPTION<br>18f424977a52ca939d9c1a44e64195e906ebcfb0dd0074590f3b5807314c7e3a<br>DA2-DECISION-ROUTE1-SUPERSEDED<br>a8fb6c233f793bfd619ddca961b5ec8969dd9ce4e7713053bab338072258f4dd<br>TRIANGULATION-OV001-PROMPT<br>549491718dea34a5bdeda8f3c3bf8764c93c9e5eeb5e2a5cf779464b1d9cbc87<br>TRIANGULATION-OV001-ANSWER<br>e50c3007eeff151c764928a5643c943a970e92fe562a2b5e13eeb4f3331c72ec |
| committed_or_manifest_sha256 | – |
| prev_entry_sha256 | 8fef4189fca8e91959d208671f5b60b3ee40840f2a6769e568685a92384f1853 |
| entry_line_sha256 | cd592f222d33e8d56f3755efd0f3b4f137a9aedfb3f6f4f703f7a9aea8b595db |

| Entry 267 | 2026-10-30 · COMMITMENT · manifest – |
|---|---|
| governing_provisions | P26-14; D-1 v13 I-K |
| subject | PG20-PASS-RECORDS-RV-NICE-V1-RV-AAO-V1<br>2263417da10a692bc014ba294d93110a90185de1465a87dd55a37a8101c6cd59<br>P16-21-CONTACT-AND-QUERY-LOG<br>276a488c830adcb50a59e81e2be26d46c3414daf6c7b220d14387fb119a0fc15<br>P16-21-LOG-PI-CONFIRMATION<br>d785e7d92215a17c40ab15b569fd214470643bbe91a43f9873e1a5fb8b9e5ad1<br>PG20-A01-PROCEDURAL-CHECK<br>ada5c33c685c3f65f66c36a69c2a363ea235c5b932b35ed51b19ecfc445cfb0d<br>PG20-A01-OBSERVATION-1-RESPONSE<br>58517405a54b1b862dcb64bd58052defa0116bb943bfa0347546129159ee7799<br>PG20-PI-DETERMINATION<br>4775db9ce8e62c40f2ec1847144a1cf7ed333ddd94958580d705d9ecee2992db |
| committed_or_manifest_sha256 | efc32f201344ef9f11019c030c7b81dd14fac57eb06867c038f692b93a35be85 |
| prev_entry_sha256 | cd592f222d33e8d56f3755efd0f3b4f137a9aedfb3f6f4f703f7a9aea8b595db |
| entry_line_sha256 | d77b252ecc8a663144df560f976dba9f0ac8775eef8ba5d696b22d2b9f23664a |

| Entry 268 | 2026-10-30 · VERIFICATION_RECORD · manifest – |
|---|---|
| governing_provisions | P26-18; CA2-32; P26-14; D-1 v13 I-K |
| subject | PG20-PASS-RECORDS-RV-NICE-V1-RV-AAO-V1<br>2263417da10a692bc014ba294d93110a90185de1465a87dd55a37a8101c6cd59<br>P16-21-CONTACT-AND-QUERY-LOG<br>276a488c830adcb50a59e81e2be26d46c3414daf6c7b220d14387fb119a0fc15<br>P16-21-LOG-PI-CONFIRMATION<br>d785e7d92215a17c40ab15b569fd214470643bbe91a43f9873e1a5fb8b9e5ad1<br>PG20-A01-PROCEDURAL-CHECK<br>ada5c33c685c3f65f66c36a69c2a363ea235c5b932b35ed51b19ecfc445cfb0d<br>PG20-A01-OBSERVATION-1-RESPONSE<br>58517405a54b1b862dcb64bd58052defa0116bb943bfa0347546129159ee7799<br>PG20-PI-DETERMINATION<br>4775db9ce8e62c40f2ec1847144a1cf7ed333ddd94958580d705d9ecee2992db |
| committed_or_manifest_sha256 | – |
| prev_entry_sha256 | d77b252ecc8a663144df560f976dba9f0ac8775eef8ba5d696b22d2b9f23664a |
| entry_line_sha256 | 7634cbe70d9f214332eeaaa262a7e3d9fb305c2443752f788fc8b1c55845f1dc |

| Entry 269 | 2026-10-30 · FREEZE_ACT · manifest M0018 |
|---|---|
| governing_provisions | P26-18; P26-07; P26-06; P26-16; P26-02(vi); PG-23 P23-04 |
| subject | PG26-M0018<br>588f8ce03f070a779e4d164d02f75f08617d42b1bebe3f19dd7134cbb0de8d6c |
| committed_or_manifest_sha256 | 588f8ce03f070a779e4d164d02f75f08617d42b1bebe3f19dd7134cbb0de8d6c |
| prev_entry_sha256 | 7634cbe70d9f214332eeaaa262a7e3d9fb305c2443752f788fc8b1c55845f1dc |
| entry_line_sha256 | ca0f8e598e434403ecc23197f39b2f8adf6f1ff612989f62eb7b0d53140c5f55 |

| Entry 270 | 2026-10-30 · COMMITMENT · manifest – |
|---|---|
| governing_provisions | P26-02(iii) as amended by CA2-10/CA3-10; P26-07 |
| subject | PG26-M0018<br>588f8ce03f070a779e4d164d02f75f08617d42b1bebe3f19dd7134cbb0de8d6c |
| committed_or_manifest_sha256 | 588f8ce03f070a779e4d164d02f75f08617d42b1bebe3f19dd7134cbb0de8d6c |
| prev_entry_sha256 | ca0f8e598e434403ecc23197f39b2f8adf6f1ff612989f62eb7b0d53140c5f55 |
| entry_line_sha256 | 97e4fa2f8ce417a0e2d7e3fbc310906d5aba971b333c3680347826fb404e0acd |

| Entry 271 | 2026-10-30 · VERIFICATION_RECORD · manifest – |
|---|---|
| governing_provisions | P26-18; P26-07; P26-12 |
| subject | PG26-M0018<br>588f8ce03f070a779e4d164d02f75f08617d42b1bebe3f19dd7134cbb0de8d6c |
| committed_or_manifest_sha256 | – |
| prev_entry_sha256 | 97e4fa2f8ce417a0e2d7e3fbc310906d5aba971b333c3680347826fb404e0acd |
| entry_line_sha256 | 3190a502fee5470726552b84ff38a21fb16a5b523ee2d2fc41b089d7fb63e37f |

| Entry 272 | 2026-10-30 · FREEZE_ACT · manifest M0019 |
|---|---|
| governing_provisions | P26-18; P26-14; P26-02(vi) |
| subject | PG26-M0019<br>e0133232c8a7c5ac6e2f162b956f5c02021f2fa565f1235da198cb1d72599456 |
| committed_or_manifest_sha256 | e0133232c8a7c5ac6e2f162b956f5c02021f2fa565f1235da198cb1d72599456 |
| prev_entry_sha256 | 3190a502fee5470726552b84ff38a21fb16a5b523ee2d2fc41b089d7fb63e37f |
| entry_line_sha256 | bfa1a9f3f16a4e87979b76cb2ee545185be2324c0d67266f3f2638f00870a8df |

| Entry 273 | 2026-10-30 · COMMITMENT · manifest – |
|---|---|
| governing_provisions | P26-02(iii) as amended by CA2-10/CA3-10; P26-14 |
| subject | PG26-M0019<br>e0133232c8a7c5ac6e2f162b956f5c02021f2fa565f1235da198cb1d72599456 |
| committed_or_manifest_sha256 | e0133232c8a7c5ac6e2f162b956f5c02021f2fa565f1235da198cb1d72599456 |
| prev_entry_sha256 | bfa1a9f3f16a4e87979b76cb2ee545185be2324c0d67266f3f2638f00870a8df |
| entry_line_sha256 | 1ecc31ec72fd4ea12f46dd4967ca01923a926e61f44f57569af9a5964a6eadf1 |

| Entry 274 | 2026-10-30 · FREEZE_ACT · manifest M0020 |
|---|---|
| governing_provisions | P26-18; P26-07; P26-02(vi); PG-21 P21-01; PG-21 P21-11 |
| subject | PG26-M0020<br>087ec2a2b9953f005216047f2fab6d1924424b9445337f7e4edb0d9104a3a933 |
| committed_or_manifest_sha256 | 087ec2a2b9953f005216047f2fab6d1924424b9445337f7e4edb0d9104a3a933 |
| prev_entry_sha256 | 1ecc31ec72fd4ea12f46dd4967ca01923a926e61f44f57569af9a5964a6eadf1 |
| entry_line_sha256 | 1de676ad32b0c022b985eaab05ae77b97a6f4505d743df630dc80f36da80429a |

| Entry 275 | 2026-10-30 · COMMITMENT · manifest – |
|---|---|
| governing_provisions | P26-02(iii) as amended by CA2-10/CA3-10; P26-07 |
| subject | PG26-M0020<br>087ec2a2b9953f005216047f2fab6d1924424b9445337f7e4edb0d9104a3a933 |
| committed_or_manifest_sha256 | 087ec2a2b9953f005216047f2fab6d1924424b9445337f7e4edb0d9104a3a933 |
| prev_entry_sha256 | 1de676ad32b0c022b985eaab05ae77b97a6f4505d743df630dc80f36da80429a |
| entry_line_sha256 | 6e3f62806c9453ad246697b507e8219c38d2946ab18960f8d281a6c10265bc41 |

| Entry 276 | 2026-10-30 · VERIFICATION_RECORD · manifest – |
|---|---|
| **governing_provisions** | P26-18; P26-14; P26-12 |
| **subject** | PG26-M0019<br>e0133232c8a7c5ac6e2f162b956f5c02021f2fa565f1235da198cb1d72599456 |
| **committed_or_manifest_sha256** | – |
| **prev_entry_sha256** | 6e3f62806c9453ad246697b507e8219c38d2946ab18960f8d281a6c10265bc41 |
| **entry_line_sha256** | 7b5c23ea04dbb88b0cffb057107c258f449c60a2b7c78fe85deae1222c2ea845 |

| Entry 277 | 2026-10-30 · VERIFICATION_RECORD · manifest – |
|---|---|
| **governing_provisions** | P26-18; P26-07; P26-12 |
| **subject** | PG26-M0020<br>087ec2a2b9953f005216047f2fab6d1924424b9445337f7e4edb0d9104a3a933 |
| **committed_or_manifest_sha256** | – |
| **prev_entry_sha256** | 7b5c23ea04dbb88b0cffb057107c258f449c60a2b7c78fe85deae1222c2ea845 |
| **entry_line_sha256** | ff23344af05bdb71a86c7b7d1bc0c406298a034c2edc00fb546ccf416af7ac23 |

| Entry 278 | 2026-10-30 · FREEZE_ACT · manifest M0021 |
|---|---|
| **governing_provisions** | P26-18; P26-07; P26-02(vi); PG-21 P21-11 |
| **subject** | PG26-M0021<br>c5527efadfe5b2606631c4cd24dee669bfd7c2ce1a54bfa1e6ce28bbf9a24b33 |
| **committed_or_manifest_sha256** | c5527efadfe5b2606631c4cd24dee669bfd7c2ce1a54bfa1e6ce28bbf9a24b33 |
| **prev_entry_sha256** | ff23344af05bdb71a86c7b7d1bc0c406298a034c2edc00fb546ccf416af7ac23 |
| **entry_line_sha256** | c2bfc4b1e4f2f525597dacf38d6857c7758d986a99ef4ca25caf76b22dd58998 |

| Entry 279 | 2026-10-30 · COMMITMENT · manifest – |
|---|---|
| **governing_provisions** | P26-02(iii) as amended by CA2-10/CA3-10; P26-07 |
| **subject** | PG26-M0021<br>c5527efadfe5b2606631c4cd24dee669bfd7c2ce1a54bfa1e6ce28bbf9a24b33 |
| **committed_or_manifest_sha256** | c5527efadfe5b2606631c4cd24dee669bfd7c2ce1a54bfa1e6ce28bbf9a24b33 |
| **prev_entry_sha256** | c2bfc4b1e4f2f525597dacf38d6857c7758d986a99ef4ca25caf76b22dd58998 |
| **entry_line_sha256** | 528b616be217ca9ff13021e895c43036edcd0de7b0343852b5347b174fcb6c41 |

| Entry 280 | 2026-10-30 · VERIFICATION_RECORD · manifest – |
|---|---|
| **governing_provisions** | P26-18; P26-07; P26-12 |
| **subject** | PG26-M0021<br>c5527efadfe5b2606631c4cd24dee669bfd7c2ce1a54bfa1e6ce28bbf9a24b33 |
| **committed_or_manifest_sha256** | – |
| **prev_entry_sha256** | 528b616be217ca9ff13021e895c43036edcd0de7b0343852b5347b174fcb6c41 |
| **entry_line_sha256** | f09a8d424d845be5e1e8692affc07e634ae2c18ed7709ca897b70d3f2b0e4507 |

| Entry 281 | 2026-10-30 · FREEZE_ACT · manifest M0022 |
|---|---|
| **governing_provisions** | P26-18; P26-09; P26-02(vi); PG-25 P25-05 |
| **subject** | PG26-M0022<br>8da8b49a93999c09d224bebe0f72972ded38acab0e46e61ec3466fe3ea3581f6 |
| **committed_or_manifest_sha256** | 8da8b49a93999c09d224bebe0f72972ded38acab0e46e61ec3466fe3ea3581f6 |
| **prev_entry_sha256** | f09a8d424d845be5e1e8692affc07e634ae2c18ed7709ca897b70d3f2b0e4507 |
| **entry_line_sha256** | 6f25534db822a02abc67acd48ddf8b4b32f236d7664e19155e32251a89f19bd7 |

| Entry 282 | 2026-10-30 · COMMITMENT · manifest – |
|---|---|
| **governing_provisions** | P26-02(iii) as amended by CA2-10/CA3-10; P26-09 |
| **subject** | PG26-M0022<br>8da8b49a93999c09d224bebe0f72972ded38acab0e46e61ec3466fe3ea3581f6 |
| **committed_or_manifest_sha256** | 8da8b49a93999c09d224bebe0f72972ded38acab0e46e61ec3466fe3ea3581f6 |
| **prev_entry_sha256** | 6f25534db822a02abc67acd48ddf8b4b32f236d7664e19155e32251a89f19bd7 |
| **entry_line_sha256** | 0b15a377c948e15f722f62859968b3a852f7d45b2ff6cee78b9226fbb79fc993 |

| Entry 283 | 2026-10-30 · VERIFICATION_RECORD · manifest – |
|---|---|
| **governing_provisions** | P26-18; P26-09; P26-12 |
| **subject** | PG26-M0022<br>8da8b49a93999c09d224bebe0f72972ded38acab0e46e61ec3466fe3ea3581f6 |
| **committed_or_manifest_sha256** | – |
| **prev_entry_sha256** | 0b15a377c948e15f722f62859968b3a852f7d45b2ff6cee78b9226fbb79fc993 |
| **entry_line_sha256** | 1be4c4fc0ca09dc93be6f6ec66611fd3a93ed7cde36a38eeac8a78c2112fa82c |

| Entry 284 | 2026-10-30 · COMMITMENT · manifest – |
|---|---|
| **governing_provisions** | P26-14; D-1 v13 I-K |
| **subject** | S10-OUT-001<br>22512f9a7edd6faecda5aa5ba22bc7b1319dc42d5cbfec0f8e78463a2cd722ca<br>S10-OUT-002<br>145afad0f810851fc364b774da9005c9b1fbb43d0c7edab65224d39ddeab0f5b<br>S10-OUT-003<br>aca32c4688ff26bbeed121fa8388a1f8fbd1fa9edb0af4ad12f09e6b8dabc559<br>S10-OUT-004<br>44cc71bec4056b7582b559b08ae966ba6461f6e00bf3ed2d4c7c4ee5e8ac3379<br>S10-OUT-005<br>974767aefd60ebe0bd07d36eaff5cef0177d749519ab895d15cfe3f2e4ac6aa0<br>S10-OUT-006<br>c620b87b471df4770b9729a8914844ace64b3b5fe61938c8b316bb3b8846f52a<br>S10-OUT-007<br>c1323fbeef9bf07a237431e7b0da4054132d471d9cf6b5fcb738c2d86c646e96<br>S10-OUT-008<br>f375f2f7db8f7b295558b8e76c0f58cdea8eb9287c970cb39bfdf864b8ae4ad7<br>S10-OUT-009<br>b8eb8e6dee1c4277cb3883cfd3e5a15c476cc990b2f52783284069e7b4147177<br>S10-OUT-010<br>323733ad783e88831a69cb8dbb5e09be5df5319ac2b4f40ca1cf408325b83ee7<br>S10-OUT-011<br>cb40dbb29db12d14997399949be05d40e716cc2744cb7956e9d52dc728e8fcdf<br>S10-OUT-012<br>a213e2471ff2da74ea3ff99e8c0e5168cb584733cbf643ff6b13b6cbe8afadf8<br>S10-OUT-013<br>b2fc03709205c26200928336382ff70cab3b0e21ce100aabbc104415cf691e96<br>S10-OUT-014<br>5e4f403206d2ee1944978b073a64f005488505ed5e79a7b3631baef20c79140f<br>S10-OUT-015<br>701f18f472fbb853dff8903ff443692cd09805f68d82f1804ccaa32a2f3e1b5a<br>S10-OUT-016<br>bdeb5669e4e2df21a2b8caca88ed80ef930e8e099413c901147eac21f23e39fa |
| **committed_or_manifest_sha256** | b34379168dbe6cf2b44de90c538c8d23ec179e3bdcc3609e9410880d499cd510 |
| **prev_entry_sha256** | 1be4c4fc0ca09dc93be6f6ec66611fd3a93ed7cde36a38eeac8a78c2112fa82c |
| **entry_line_sha256** | 82810b2e817229e8ddbb24993446c67bc52b686c799739c5189d97735bf3140c |

| Entry 285 | 2026-10-30 · COMMITMENT · manifest – |
|---|---|
| **governing_provisions** | P26-14; D-1 v13 I-K |
| **subject** | S10-CODEX-VERBATIM<br>0d9df846affff598b324a172e815b39a005a92463cb7d02d1ab7540ca0ea58de<br>S10-CODEX-REQUEST<br>f1f8b8c98c21c55393ae90acb3600ba18933444b0809e29daaa3c2c5bcd819be<br>S10-CODEX-PACKAGE<br>a2a8d715668306162072acc2cfe8e1a73bbef1565f002d94e1014ac25690c423 |
| **committed_or_manifest_sha256** | 3a557c71061eff5576f7875a3e1441ff6f4305cdcb3ce1c88f1a8ead6d8ddc89 |
| **prev_entry_sha256** | 82810b2e817229e8ddbb24993446c67bc52b686c799739c5189d97735bf3140c |
| **entry_line_sha256** | 4e0493825aa1f5c5b8e496bfe6424ae79fc5537378cc993a5bca4f072f5c8268 |
